# Model Misspecification Misleads Inference of the Spatial Dynamics of Disease Outbreaks

**DOI:** 10.1101/2022.08.24.22278802

**Authors:** Jiansi Gao, Michael R. May, Bruce Rannala, Brian R. Moore

## Abstract

Epidemiology has been transformed by the advent of Bayesian phylodynamic models that allow researchers to infer the geographic history of pathogen dispersal over a set of discrete geographic areas (1, 2). These models provide powerful tools for understanding the spatial dynamics of disease outbreaks, but contain many parameters that are inferred from minimal geographic information (*i*.*e*., the single area in which each pathogen was sampled). Consequently, inferences under these models are inherently sensitive to our prior assumptions about the model parameters. Here, we demonstrate that the default priors used in empirical phylodynamic studies make strong and biologically unrealistic assumptions about the underlying geographic process. We provide empirical evidence that these unrealistic priors strongly (and adversely) impact commonly reported aspects of epidemiological studies, including: (1) the relative rates of dispersal between areas; (2) the importance of dispersal routes for the spread of pathogens among areas; (3) the number of dispersal events between areas, and; (4) the ancestral area in which a given outbreak originated. We offer strategies to avoid these problems, and develop tools to help researchers specify more biologically reasonable prior models that will realize the full potential of phylodynamic methods to elucidate pathogen biology and, ultimately, inform surveillance and monitoring policies to mitigate the impacts of disease outbreaks.

**Significance Statement:** Bayesian phylodynamic models have revolutionized epidemiology by enabling researchers to infer key aspects of the geographic history of disease outbreaks. These models contain many parameters that must be estimated from minimal information (the area from which each pathogen was sampled), rendering inferences under this approach inherently sensitive to the choice of priors on the model parameters. Here, we demonstrate that: (1) the priors assumed in *≈*93% of surveyed phylodynamic studies make strong and biologically unrealistic assumptions, and; (2) these priors distort the conclusions of epidemiological studies. We offer strategies and tools to specify more reasonable priors that will enhance our ability to understand pathogen biology and, thereby, to mitigate disease.

---

**P**hylogenies are now central to epidemiological studies; this *phylodynamic* approach is used to infer various aspects of pathogen biology, including patterns of variation in demographic and geographic history. The approach developed by Lemey *et al*. (1, 2)—implemented in the `BEAST` software package (3, 4)—is now the standard approach used to elucidate the geographic history of disease outbreaks and has featured prominently in studies of the COVID-19 pandemic (5–11). These discrete-geographic models allow us to infer key aspects of disease outbreaks, including: (1) the area in which an epidemic originated; (2) the dispersal routes by which the pathogen spread among geographic areas (where a dispersal route is a direct path between a pair of geographic areas), and; (3) the number of dispersal events between areas.

Under this approach, geographic history involves dispersal among a set of discrete areas (*e*.*g*., cities, states, countries) over the branches of the pathogen phylogeny. Geographic history is modeled as a probabilistic process with parameters that specify the average rate of pathogen dispersal among all geographic areas, and the relative rates of pathogen dispersal between pairs of geographic areas. Inference under these discrete-geographic models is performed within a Bayesian statistical framework. Bayesian inference requires that we specify a *prior probability* distribution for each parameter of the geographic model (reflecting our beliefs about the corresponding parameter values *before* evaluating the data at hand); the priors are updated by the information in our data (the geographic area from which each pathogen was sampled) to provide a *posterior probability* distribution for each of the model parameters (reflecting our beliefs about the parameter values *after* evaluating our study data).

These geographic models contain many parameters that must be inferred from minimal information; the data are limited to a single observation for each sampled pathogen (*i*.*e*., the area in which each pathogen was sampled). Accordingly, geographic inference under this approach is inherently sensitive to the assumed priors. Here, we demonstrate that the priors on the average dispersal rate and the number of dispersal routes implemented as defaults in `BEAST` (and used in most empirical studies; Figure 1) make strong and biologically unrealistic assumptions about the underlying geographic process. We present empirical evidence demonstrating that these priors are strongly disfavored by real data, and that these priors strongly (and adversely) distort central conclusions of epidemiological studies. Finally, we offer strategies—and develop tools—to help researchers specify more biologically reasonable priors that will enhance the potential of phylodynamic methods to elucidate pathogen biology.

**Fig. 1.**
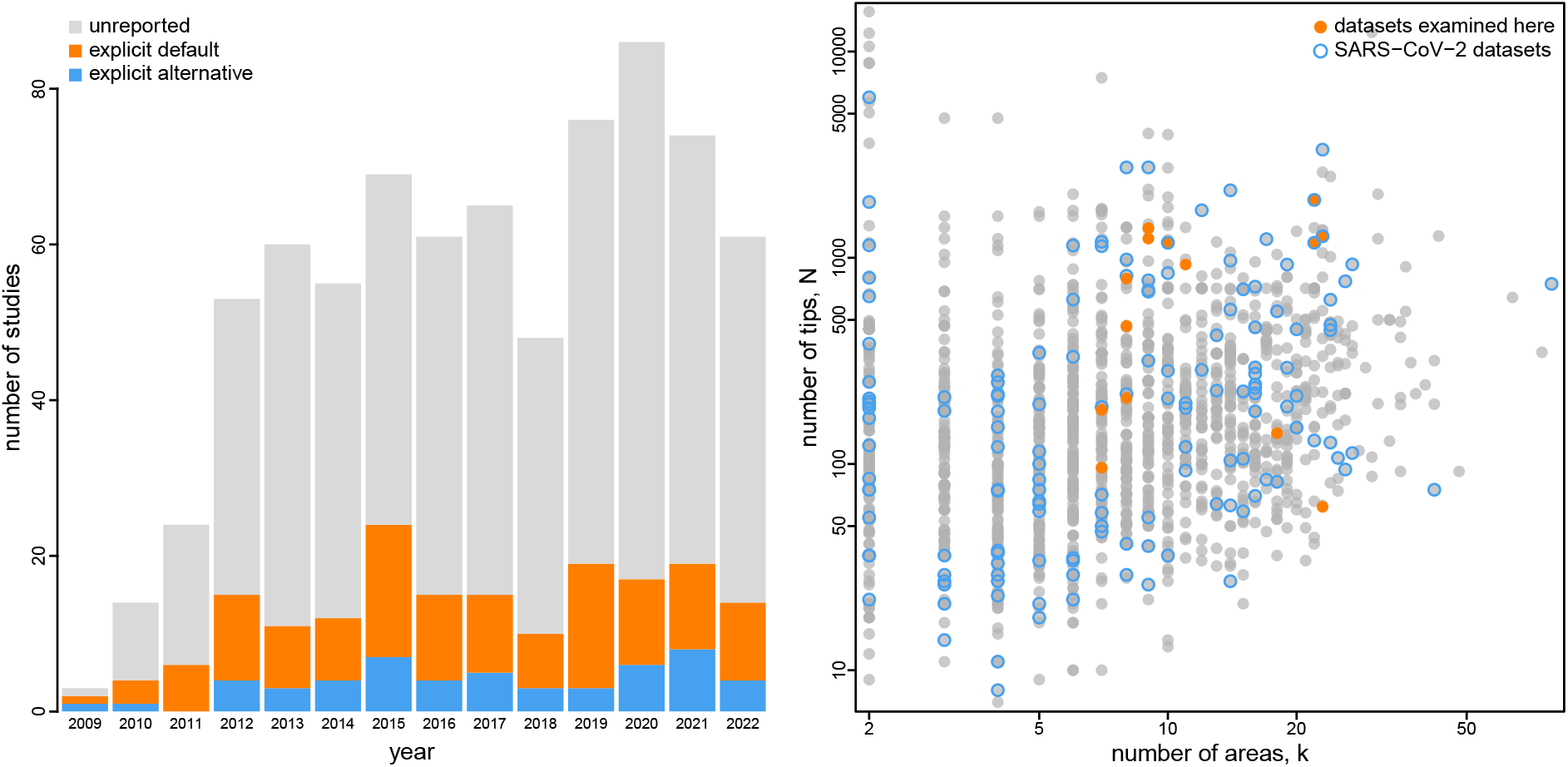
Empirical phylodynamic studies of pathogen geographic history. The bar plot at left depicts the choice of priors on the average dispersal rate and/or number of dispersal routes in the 749 published empirical studies (obtained from Google Scholar on August 11, 2022) that inferred the geographic history of pathogens using the approach of Lemey *et al*. (1). The vast majority of these studies explicitly (orange) or implicitly (gray) specified default priors on these parameters; only 7.1% of published studies used non-default priors on the average dispersal rate and/or number of dispersal routes (blue). The right panel depicts the size of published empirical datasets in terms of the number of geographic areas (*x*-axis) and the number of tips (*y*-axis). Orange dots indicate empirical datasets included in our study; open blue dots indicate SARS-CoV-2 datasets.

## Theoretical Concerns and Proposed Solutions

Each tip of the study phylogeny corresponds to a sampled pathogen that occurs in one of *k* discrete geographic areas. We simplify our presentation by assuming that the study phylogeny with divergence times is known. (In practice, the geographic history and study phylogeny are usually inferred simultaneously; see Supplemental Material.) We first describe the discrete-geographic model proposed by Lemey *et al*. (1); we then discuss theoretical concerns related to the priors on the parameters of that model and suggest alternative priors to address these concerns.

### The Model

Discrete-geographic models describe the history of pathogen dispersal over the tree, Ψ, as a continuous-time Markov chain (CTMC). For a geographic history with *k* discrete areas, this stochastic process is fully specified by a *k* × *k* instantaneous-rate matrix, *Q*, where an element of the matrix, *qij*, is the instantaneous rate of change between states *i* and *j* (*i*.*e*., the instantaneous rate of dispersal from area *i* to area *j*). In principle, we may wish to treat each element of this matrix as a free parameter to be estimated from the data. In practice, *k* is typically large, such that the geographic model includes many parameters, while the data are limited to a single geographic observation (the location where each pathogen was sampled). This raises concerns about our ability to estimate each parameter in the matrix, which motivated Lemey *et al*. (1) to develop a Bayesian approach to simplify the geographic model. This is accomplished by specifying each element, *qij*, of the instantaneous-rate matrix, *Q*, as:

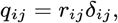

where *r*_*ij*_ is the relative rate of dispersal between areas *i* and *j*, and *δ*_*ij*_ is an indicator variable that takes one of two states (0 or 1). When *δ*_*ij*_ = 1, the instantaneous dispersal rate for the corresponding element, *q*_*ij*_, is simply *q*_*ij*_ = *r*_*ij*_. Conversely, when *δ*_*ij*_ = 0, the instantaneous dispersal rate for the corresponding element, *q*_*ij*_, is zero, effectively removing that parameter from the geographic model. For a given *Q* matrix there is a vector of *δ*_*ij*_ and a vector of *r*_*ij*_. Each unique vector of *δ*_*ij*_ —*i*.*e*., ***δ***, a string of zeros and ones for each of the possible pairwise dispersal routes between the *k* geographic areas—corresponds to a unique geographic model (Figure 2). By convention, the *Q* matrix is rescaled such that the expected number of dispersal events in one time unit is equal to the parameter *μ* (12).

**Fig. 2.**
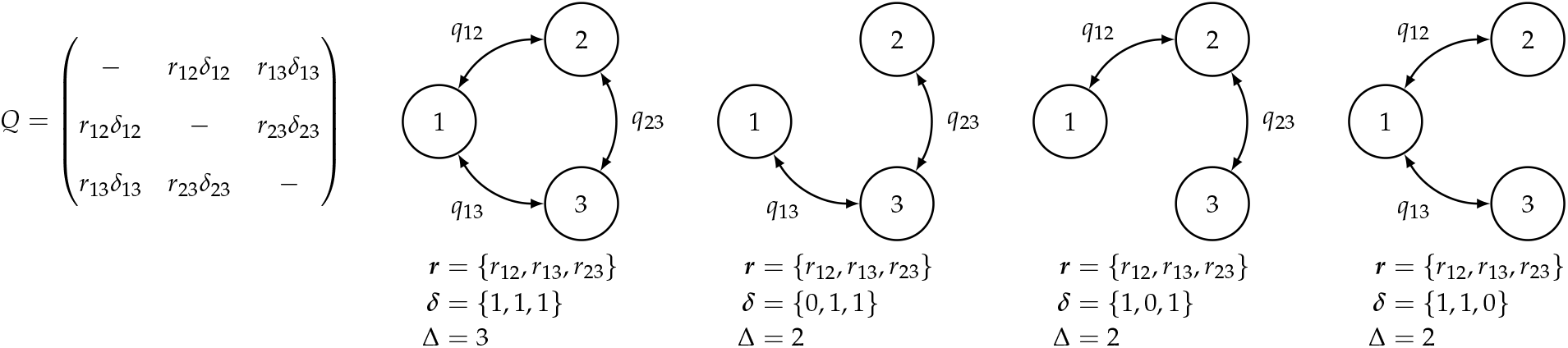
Discrete-geographic models for *k* = 3 areas. The approach of Lemey *et al*. (1) models the evolution of geographic range using a continuous-time Markov chain (CTMC). The CTMC is completely described by the instantaneous-rate matrix, *Q*, where each element *q*_*ij*_ specifies the instantaneous rate of dispersal between areas *i* and *j*. Each element, *q*_*ij*_, is a function of the relative-rate parameter, *r*_*ij*_, and a dispersal-route indicator, *δ*_*ij*_ (left panel). The dispersal-route indicator, *δ*_*ij*_, is 1 when the corresponding dispersal route exists, and 0 when it does not exist. Alternative geographic models are specified by different configurations of dispersal routes. In the first model, all possible dispersal routes exist; the remaining models have two viable dispersal routes, corresponding to different vectors of dispersal-route indicators, ***δ*** (right panels). The total number of dispersal routes for a given geographic model is Δ. Note that there may be multiple distinct geographic models with an equal number of dispersal routes, Δ (*e*.*g*., the three distinct models depicted here for which Δ = 2). The models depicted are all symmetric; *i*.*e*., they assume that the rate of dispersal from area *i* to area *j* is equal to the rate of dispersal from area *j* to area *i*.

The original method (1) assumes that instantaneous-rate matrix, *Q*, is symmetric, where *q*_*ij*_ = *qji* (*i*.*e*., *r*_*ij*_ = *r*_*ji*_ and *δ*_*ij*_ = *δ*_*ji*_). Accordingly, this model assumes that the instantaneous rate of dispersal from area *i* to area *j* is equal to the dispersal rate from area *j* to area *i*. For a dataset with *k* areas, the symmetric model has 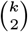 dispersal-route indicators and up to 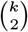 relative-rate parameters. A subsequent extension (2) allows the *Q* matrix to be asymmetric, *i*.*e*., *q*_*ij*_ and *q*_*ji*_ are not constrained to be equal. Accordingly, this model allows the rate of dispersal from area *i* to area *j* to be different from the rate of dispersal from area *j* to area *i*. For a dataset with *k* areas, the asymmetric model has *k* × (*k* −1) dispersal-route indicators and up to *k* × (*k*− 1) relative-rate parameters.

Lemey *et al*. (1) estimate the parameters of these geographic models in a Bayesian framework. Following Bayes’ theorem, the joint posterior probability distribution of the model parameters is (13):

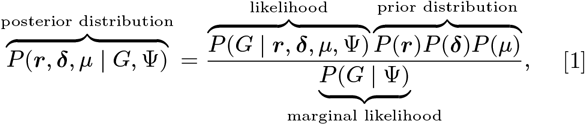

where ***r*** is a vector that contains all of the relative-rate parameters, ***δ*** is a vector that contains all of the dispersal-route indicators, *μ* is the average rate of dispersal, Ψ is the phylogeny, and *G* is the observed geographic data. The likelihood function is equal to the probability of the observed geographic data, *G*, given the geographic model, *Q*, and phylogeny, Ψ. The joint prior probability distribution reflects our beliefs about the model parameters before evaluating the geographic data at hand; the prior is updated by the information in the geographic data via the likelihood function to produce the joint posterior distribution, which reflects our beliefs about the model parameters after observing the geographic data. When the data contain limited information to update the assumed priors, posterior estimates may be sensitive to the assumed priors, a phenomenon known as *prior sensitivity*.

The denominator of Bayes theorem is the marginal likelihood (the likelihood function averaged over the parameter values, weighted by the prior probability of those parameter values), which represents the probability of observing our study data under the model. The joint posterior probability distribution is approximated using Markov chain Monte Carlo, which samples parameter values with a frequency proportional to their posterior probabilities.

### Prior on the Number of Dispersal Routes

Recall that each vector, ***δ***, specifies a unique configuration of dispersal routes, which corresponds to a unique geographic model. The total number of dispersal routes for a given geographic model is denoted Δ. For a given value of Δ, there may be multiple distinct geographic models (*e*.*g*., the three distinct symmetric models with Δ = 2 dispersal routes depicted in Figure 2). Lemey *et al*. (1) impose a prior on irreducible geographic models—where each area can be reached (either directly or indirectly) from any other area—by: (1) placing a prior on the total number of dispersal routes, Δ, and; (2) assuming that all irreducible geographic models with a given value of Δ are equiprobable. For example, the three distinct geographic models with Δ = 2 depicted in Figure 2 are assumed to have equal prior probability. Together, these assumptions induce a prior probability that a given dispersal route between areas *i* and *j* exists, *i*.*e*., that *δ*_*ij*_ = 1.

For the symmetric model, Lemey *et al*. (1) specify an *offset* Poisson prior on the total number of dispersal routes, Δ. That is, the prior on Δ assigns zero probability to all geographic models with fewer than *k*− 1 dispersal routes; this reflects the constraint that a dataset with *k* geographic areas cannot be realized under a CTMC with fewer than *k* − 1 non-zero *q*_*ij*_ values (*i*.*e*., dispersal routes).^*^ The prior on the number of dispersal routes *greater than k* −1 is described by a Poisson prior with rate parameter, *λ*. Lemey *et al*. (1) express an explicit prior preference for geographic models with the minimal number of dispersal routes. Specifically, by default, *λ* = ln(2),which places ∼40% of the prior mass on models with the absolute minimum number of dispersal routes (Δ = *k* − 1; Figure 3, left panel). For the asymmetric model, the number of dispersal routes is assumed to be drawn from a Poisson prior with rate *λ*. In this case, *λ* is specified such that the expected number of dispersal routes is *k* − 1 (Figure S1; note that this prior does not enforce a minimum number of dispersal routes).

**Fig. 3.**
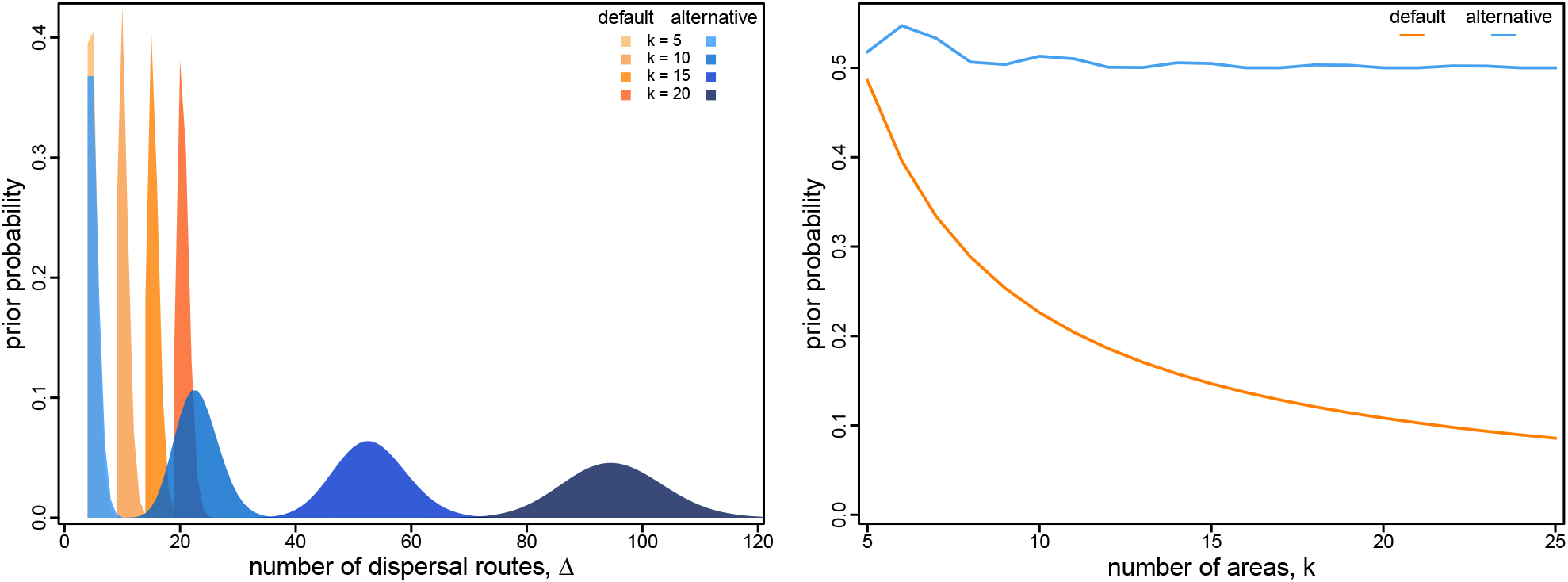
Prior on dispersal routes under the symmetric geographic model. The left panel illustrates the default (orange) and alternative (blue) prior distributions on the total number of dispersal routes, Δ, as a function of the number of areas, *k*. The default-prior distributions are highly focused on the minimal number of dispersal routes, *k* − 1, whereas the alternative-prior distributions are centered on an intermediate number of dispersal routes (*i*.*e*., the expected number of dispersal routes is about half the maximum number for a given value of *k*). The right panel illustrates the prior probability under the default (orange) and alternative (blue) prior models that a given dispersal route exists (*i*.*e*., that *δ*_*ij*_ = 1) as a function of the total number of geographic areas, *k*. Under the default-prior model, the probability that a given dispersal route exists drops rapidly for datasets with a moderately large (and common; *cf*. Figure 1) number of geographic areas; by contrast, under the alternative-prior model, this probability remains relatively constant for all values of *k*.

The number of dispersal-route indicators grows rapidly as a function of the number of areas, *k*; however, the prior expected number of dispersal routes grows linearly as a function of *k*. Consequently, the prior probability that any given dispersal route exists rapidly decreases as *k* increases (Figure 3, right). For inferences with large (and common; *cf*. Figure 1) values of *k*, the default prior on Δ results in an extremely informative prior on models with the minimum number of dispersal routes.

In the experiments below, we specify alternative and more diffuse priors on Δ, where the expected number of dispersal routes is about half the maximum possible number. We specify the prior mean to be half the maximum possible number so that the Poisson prior is relatively diffuse across all possible values of Δ (Figure 3, left) and this results in a relatively flat prior probability that any given dispersal route exists for all values of *k* (Figure 3, right). Specifically, for the symmetric model, we specify an offset (*i*.*e*., by *k* − 1) Poisson prior on Δ with λ specified so that the expected number of dispersal with *k* areas. For the asymmetric model, we specify a Poisson routes is about half of the maximum number, 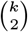, for a dataset belief that half of all possible dispersal routes are included in prior distribution on Δ with 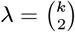, which represents a prior the geographic model.

### Prior on the Average Dispersal Rate

Recall that the rate matrix, *Q*, is rescaled so that the average rate of dispersal among all areas is *μ*. For a tree of length *T* (*i*.*e*., the sum of the durations of all branches in the tree), the expected number of dispersal events is *μ* × *T*. Therefore, the prior on *μ* is related to our prior belief about the number of dispersal events over the tree. By default, *μ* is assigned a gamma prior with shape parameter *α* = 0.5 and rate parameter *β* = *T*.^†^ The gamma distribution has a mean of *α/β*; therefore this prior expresses the belief that the average rate of dispersal is 0.5*/T* (Figure 4, left).

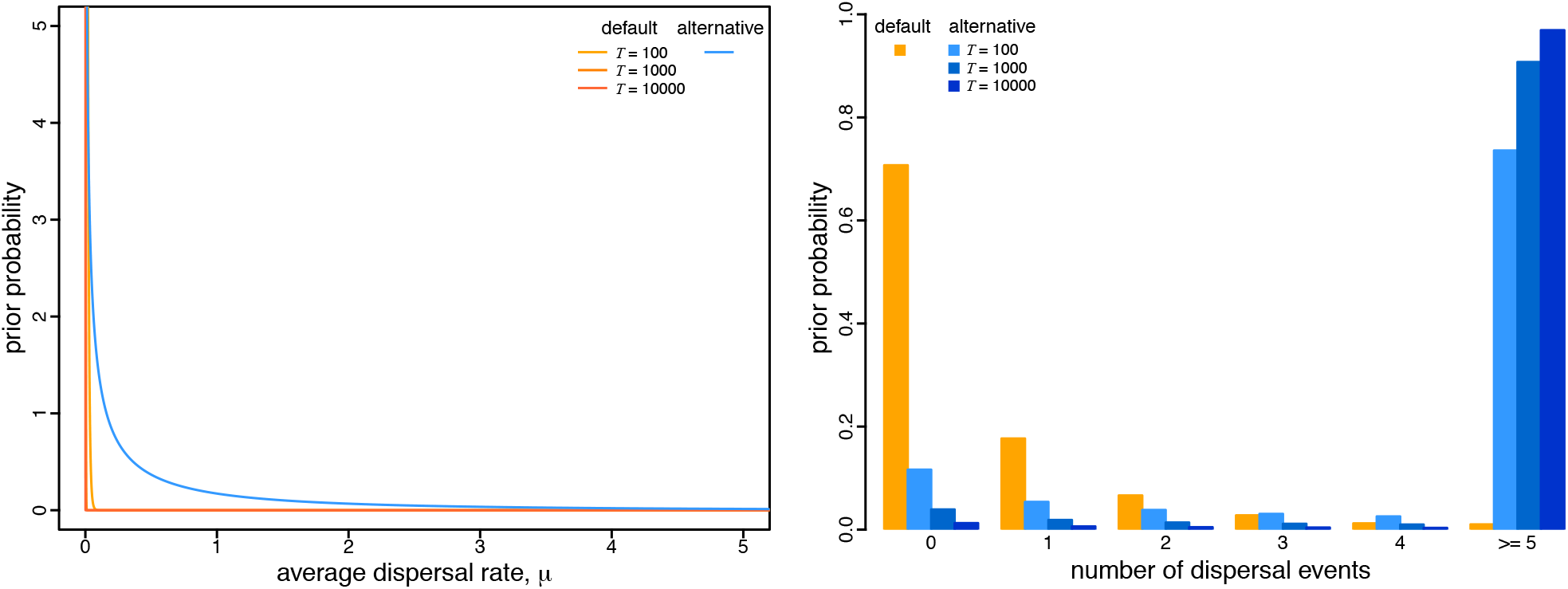

Because the expected number of dispersal events is *μ* × *T*, the prior expected number of dispersal events under this prior is 0.5, independent of the duration of the entire geographic history (*i*.*e*., the tree length, *T*), or the number of areas, *k*, in which the pathogen occurs. Similarly, the prior distribution on the number of dispersal events is independent of *T* and *k*: the 95% prior interval is [0, 3] dispersal events, which implies that we would be very surprised if a geographic history of *any* duration with *any* number of areas involved more than three dispersal events (see Figure 4, right panel). Logically, however, a geographic history that includes *k* areas minimally requires *k* −1 dispersal events. Therefore, this prior becomes increasingly unreasonable as *k* grows to large (and common; *cf*. Figure 1) values.

In our experiments below, we specify a more diffuse prior on the dispersal rate, *μ*. Specifically, we specify an exponential prior on *μ* with parameter *θ* (with a mean of 1/*θ*). To address concerns about the potential impact of assuming a fixed value of *θ* on posterior estimates, we treat the mean of the exponential prior, 1/*θ*, as a random variable to be estimated from the data. Specifically, we specify a gamma hyperprior on 1/*θ* with shape parameter, *α* = 0.5, and rate parameter, *β* = 0.5 (fixing the shape and rate parameters to be equal ensures that the resulting prior on *μ* is proper; *i*.*e*., that it integrates to one). The resulting prior—known as the *K*-distribution (14)—is more diffuse than the default prior on *μ* (Figure 4, right), as is the resulting prior distribution on the number of dispersal events (Figure 4, left). Importantly, this alternative prior distribution on the number of dispersal events sensibly scales with the duration of the entire geographic history, *T*.

### Empirical Consequences

In this section, we explore the empirical consequences of using the default priors on the number of dispersal routes and the average dispersal rate. We collected 14 datasets from published empirical studies, and reanalyzed each under a suite of geographic models, including all combinations of: (1) symmetric and asymmetric *Q* matrices; (2) default and alternative priors on the number of dispersal routes; and (3) default and alternative priors on the average dispersal rate. We first evaluated the relative and absolute fit of the eight candidate models to each empirical dataset to demonstrate that the default priors provide a poor description of the underlying geographic process. We then estimated the joint posterior distribution under each of the candidate models for each dataset to demonstrate how the strongly misinformative default priors adversely impact inferences about the geographic history of disease outbreaks. We detail these analyses in the Supplemental Material.

### The Impact of Prior Choice on Model Fit

Our concern regarding the default priors is that they represent strongly informative and biologically unrealistic beliefs about the geographic process that generates empirical data. Accordingly, we expect default priors to poorly fit empirical datasets compared to more biologically reasonable alternative priors.

Following Lemey *et al*. (1), we first tested this prediction by comparing the *relative* fit of the competing prior models to our empirical datasets. Specifically, we assessed the relative fit of each dataset to the eight candidate models using Bayes factors, which are computed as twice the difference in the log marginal likelihoods of the competing models (15). Bayes-factor comparisons indicate that the default prior on the number of dispersal routes *and* the average dispersal rate are both biologically unrealistic; the alternative priors for both parameters were significantly preferred compared to their default counterparts (Table S2).

In addition to assessing the *relative* fit of competing prior models to our empirical datasets, we also assessed the *absolute* fit of the prior models to these datasets using posterior-predictive simulation (16, 17). This approach is based on the following premise: if a given model provides an adequate description of the process that gave rise to our observed data, then new datasets simulated under that model should resemble our study data. Results of the posterior-predictive simulations corroborate our findings based on Bayes-factor comparisons: in all cases, the alternative priors provide an adequate fit to the empirical datasets, whereas the default priors are inadequate (Figures S2, S3; Tables S3, S4).

Both default priors—on the number of dispersal routes and the average dispersal rate—negatively impact the relative and absolute fit of geographic models to our empirical datasets, providing empirical evidence to support our premise that these default priors are strongly unrealistic. Nevertheless, it remains to be seen whether these unrealistic priors distort inferences about the geographic history of disease outbreaks. To this end, we inferred the joint posterior distribution for each dataset under two candidate models: one model with both default priors (‘default-prior model’), and one model with both alternative priors (‘alternative-prior model’). For both the default- and alternative-prior models, we specified the preferred *Q* matrix (*i*.*e*., symmetric or asymmetric). Note that—in all cases—the default-prior models are decisively rejected compared to the alternative-prior models (Table 1).

**Table 1.** **The relative fit of geographic models with default and alternative priors. We inferred marginal likelihoods for each dataset under two models: one using both default priors, the other both alternative priors. For each combination of priors, we assumed the preferred geographic model (*i*.*e*., with a symmetric or asymmetric rate matrix). Marginal-likelihood estimates for the default- and alternative-prior models are listed in the first two columns (**±**SD among four replicates);** 2 ln **BF between the two models are listed in the third column. The default-prior models are decisively rejected for all datasets (*i*.*e***., 2 ln **BF** ≫ 10).

| Dataset* | Default | Alternative | $2 \ln \text{BF}$ |
| --- | --- | --- | --- |
| 1 | $-187.65 \pm 0.12$ | $-147.32 \pm 0.11$ | 80.67 |
| 2 | $-142.52 \pm 0.04$ | $-128.76 \pm 0.04$ | 27.53 |
| 3 | $-214.89 \pm 0.12$ | $-174.02 \pm 0.07$ | 81.76 |
| 4 | $-106.20 \pm 0.03$ | $-91.47 \pm 0.12$ | 29.46 |
| 5 | $-1176.37 \pm 0.24$ | $-1037.50 \pm 0.04$ | 277.75 |
| 6 | $-1309.05 \pm 0.12$ | $-1164.72 \pm 0.04$ | 288.65 |
| 7 | $-835.94 \pm 0.30$ | $-726.80 \pm 0.15$ | 218.29 |
| 8 | $-2873.90 \pm 0.42$ | $-2275.48 \pm 0.04$ | 1196.85 |
| 9 | $-2333.80 \pm 0.69$ | $-1872.88 \pm 0.02$ | 921.84 |
| 10 | $-305.88 \pm 0.12$ | $-258.13 \pm 0.15$ | 95.79 |
| 11 | $-2519.33 \pm 0.12$ | $-2159.85 \pm 0.31$ | 718.97 |
| 12 | $-1983.57 \pm 0.35$ | $-1721.13 \pm 0.12$ | 524.88 |
| 13 | $-1754.35 \pm 0.04$ | $-1536.92 \pm 0.04$ | 434.88 |
| 14 | $-1372.08 \pm 0.09$ | $-1225.46 \pm 0.24$ | 293.24 |

\*Dataset sources: 1 (18); 2–4 (19); 5–7 (20); 8–9 (21); 10 (22); 11 (23); 12 (9), and; 13–14 (6).

### The Impact of Prior Choice on Pairwise Dispersal Rates

To explore the impact of prior (mis)specification on commonly reported geographic inferences, we first explored estimates of the *Q*-matrix parameters—*i*.*e*., ***r, δ***, and *μ*—under the default-prior model to those estimated under the alternative-prior model. Although these parameters are not usually reported in empirical studies, they are the actual basis of commonly reported aspects of geographic history, *i*.*e*., commonly reported inferences are a function of these *Q*-matrix parameters. The left two panels of Figure 5 compare posterior-mean estimates of *Q* under the default- and alternative-prior models for the deformed-wing virus dataset (19); the choice of prior model strongly impacts estimates of the dispersal rates between many areas. Perhaps unsurprisingly—given that the default priors imply fewer dispersal routes and a lower number of dispersal events—posterior-mean estimates under the default-prior models are systematically much lower than those inferred under the alternative-prior models.

**Fig. 5.**
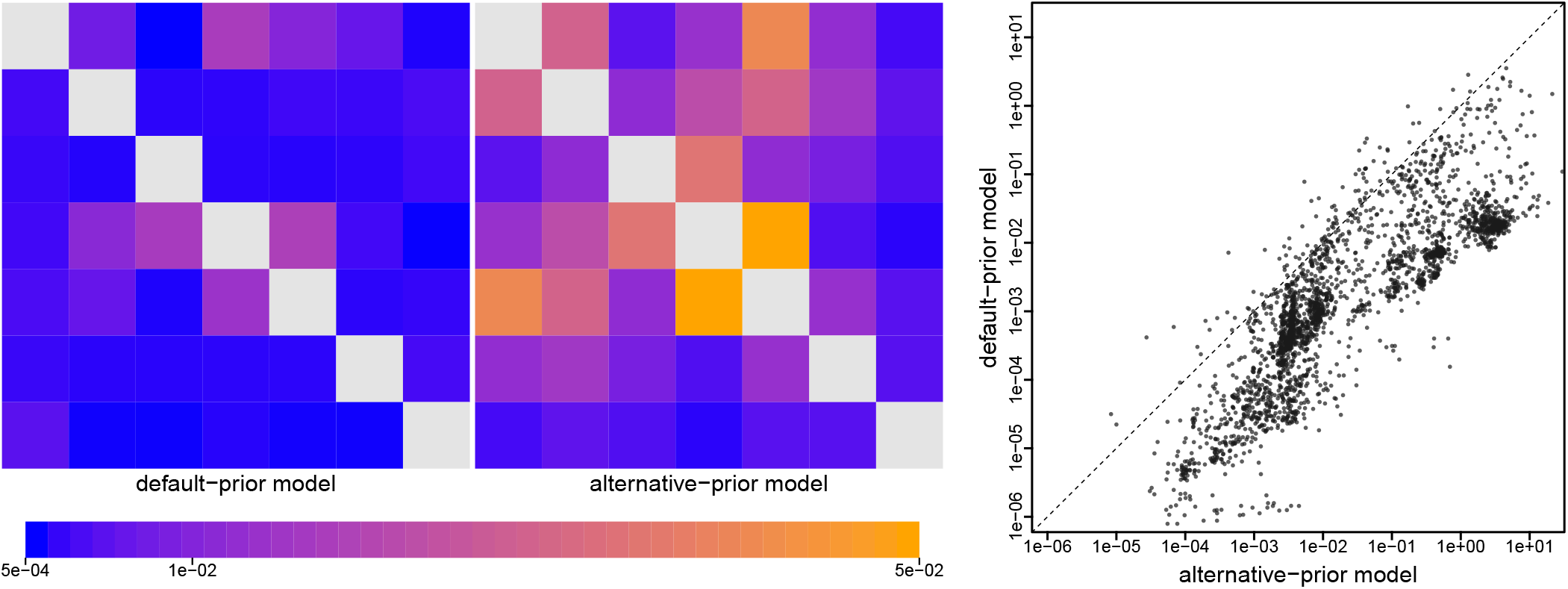
The impact of prior choice on estimates of pairwise dispersal rates. Heatmaps summarize posterior-mean estimates of the instantaneous rate of dispersal between each pair of geographic areas, *q*_*ij*_, estimated for the deformed-wing virus dataset (19) under the (disfavored) default (left panel) and (preferred) alternative (center panel) prior models. At right, we summarize dispersal-rate estimates for each pair of areas across all 14 empirical datasets. Note that dispersal-rate estimates under the default-prior model are consistently lower than those estimated under the alternative-prior model. (Uncertainty in these estimates is summarized in Figure S7.)

### The Impact of Prior Choice on Dispersal Routes

Empirical studies often focus on the evidential support for dispersal routes between each pair of geographic areas; these inferences are intended to identify dispersal routes that were important to the geographic spread of the disease. This involves computing Bayes factors for each of the dispersal-route indicators in the geographic model. Above, we computed Bayes factors for models as the difference in their log marginal likelihoods; an alternative (but equivalent) formulation is to compute the ratio of the posterior and prior odds for two competing models. For each dispersal-route indicator in the *Q* matrix, we compute the Bayes factor as:

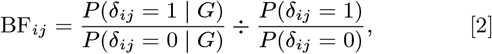

where *P* (*δ*_*ij*_ = 1) is the prior probability that a dispersal route between areas *i* and *j* exists, and *P* (*δ*_*ij*_ = 1| *G*) is the posterior probability that it exists (the latter is computed as the proportion of MCMC samples for which *δ*_*ij*_ = 1). This formulation of the Bayes factor captures the degree to which our beliefs (about the existence of a dispersal route) changed after observing the geographic data. Because the default-prior model favors geographic models with a small number of dispersal routes, the prior probability that each dispersal route exists is correspondingly small. As a result, we expect the default-prior model to increase the apparent Bayes-factor support for individual dispersal routes.

Our analyses of the H3N2 influenza virus dataset (21) illustrate the impact of the default- and alternative-prior models on the inferred support for dispersal routes (Figure 6, left panel). The default-prior model obscures our ability to identify the dispersal routes that played a potential role in the spread of this H3N2 influenza outbreak; *e*.*g*., 5 of 35 the dispersal routes decisively supported under the default-prior model (*i*.*e*. where 2 ln BF ≥ 10) appear to be spurious, and two decisively supported dispersal routes are not identified. Additionally, the rank order of these decisively supported dispersal routes differs markedly under the two prior models. The impact of prior choice on the estimated support for individual dispersal routes is pervasive across all the sampled empirical datasets (Figure 6, right panel). The scale of the Bayes factors inferred under the default-prior model is on average much higher than under the alternative-prior model.

**Fig. 6.**
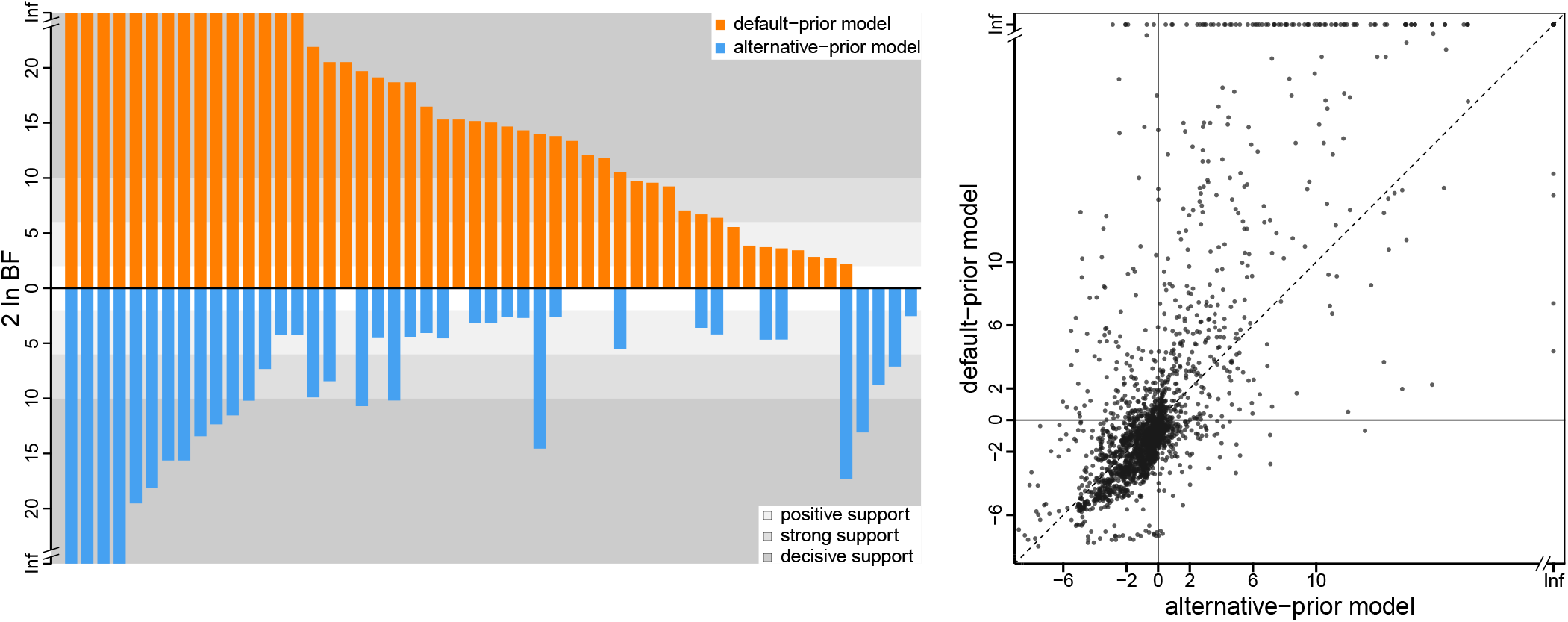
The impact of prior choice on the inferred support for dispersal routes. The left panel compares the evidential support for dispersal routes under the default (orange) and alternative (blue) prior models for the H3N2 influenza virus dataset (21). Each bar indicates the 2 ln BF (Bayes factor) for the corresponding dispersal route between two geographic areas; only “significant” dispersal routes (*i*.*e*., 2 ln BF > 2) are plotted. Background shading indicates the level of support; following Kass and Raftery (15), the support level is “positive” (light gray) when 2 < 2 ln BF ≤ 6, “strong” (gray) when 6 < 2 ln BF ≤ 10, and “decisive” (dark gray) when 2 ln BF > 10. Some dispersal routes identified as significant under the default-prior model have no support (*i*.*e*., 2 ln BF ≤ 2) under the alternative-prior model, and *vice versa*. Additionally, the rank order of dispersal routes according to their Bayes-factor support differs between the default- and alternative-prior models. The right panel plots the 2 ln BF for each dispersal route under the default (y-axis) and alternative (x-axis) prior models across all empirical datasets. Note that, under the alternative-prior model, many dispersal routes have equivocal Bayes-factor support (*i*.*e*., −2 ≤ 2 ln BF ≤ 2); conversely, Bayes factors under the default-prior model tend to be larger than those under the alternative-prior model (dots above the diagonal indicate greater support under the default-prior model compared to the alternative-prior model) (Uncertainty in these estimates is summarized in Figure S10.)

### The Impact of Prior Choice on Inferences of Geographic History

Empirical studies frequently report summaries that are based on the conditional probability distribution of geographic histories over the tree. The distribution of histories depends on—*i*.*e*., is *conditioned* on—the instantaneous-rate matrix, *Q*, the geographic data, *G*, and the phylogeny, Ψ. Conceptually, for a given tree and rate matrix, we imagine simulating a geographic history over the tree from root to tips, where the rate matrix specifies the waiting times between dispersal events. We can construct the conditional distribution of geographic histories by simulating many individual histories, and retaining only those histories that realize the observed geographic areas at the tips, *G*. This conditional distribution contains the information required to compute two commonly reported summaries: the ancestral areas at internal nodes of the tree, and the number of dispersal events between geographic areas. Because these summaries depend on the rate matrix, which in turn is sensitive to the choice of prior (Figure 5), we expect the prior to influence these summaries. We detail the impacts of default and alternative priors on each of these commonly reported summaries below.

#### Inferring ancestral areas

It is often critical to identify the point of origin for an outbreak. This involves inferring the probability that the corresponding internal node of the tree (including the root) occurred in each of the *k* geographic areas. The probability that a given node was in a particular area is simply the proportion of conditional histories for which the node is in that area. Our reanalysis of the SARS-CoV-2 Global dataset (23) reveals that the choice of prior model may exert a strong impact on estimates of ancestral areas. The first known outbreak of COVID-19 in North America occurred in the state of Washington. The origin of this ‘Washington Clade’ is therefore of considerable interest (5, 24); the default-prior model unequivocally identifies Western North America as the source of this outbreak (posterior probability 90.0%).

By contrast, the (preferred) alternative-prior model reveals Western North America (posterior probability 35.7%) and China (posterior probability 38.9% combining subareas) to be equiprobable sources of the Washington COVID-19 outbreak (Figure 7, left panel). The impact of prior models on ancestral-area estimates is prevalent across the 14 datasets; the choice of prior not only impacted the inferred probability of the most probable area at an internal node, but also changed the identity of the most probable (MAP) ancestral area for *≈*10% of the internal nodes (Figure 7, right panel). On average, the ancestral-area estimates tend to be more certain under the default-prior model—where the MAP ancestral area is generally inferred with a higher posterior probability compared to that under the alternative-prior model—which is consistent with our expectation given that the default priors are strongly informative.

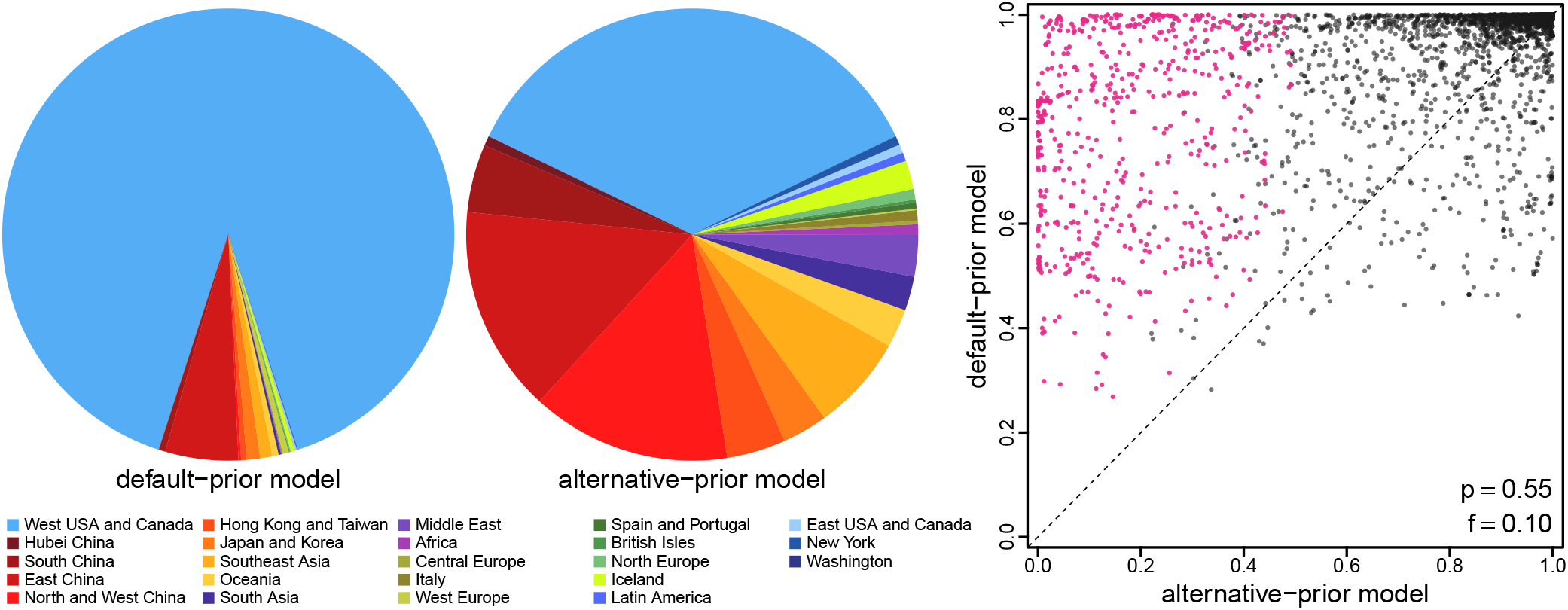

#### Inferring the number of dispersal events

Empirical phylodynamic studies often infer the number of dispersal events between each pair of areas, *e*.*g*., to understand whether a given area is a major source of disease outbreaks. A given conditional geographic history includes the number of dispersal events between each pair of areas; therefore, we can compute the average number of dispersal events between each pair of areas as the posterior-mean number of events over the conditional distribution of histories. The choice of prior model exerts a strong influence on estimates of the number of dispersal events. For example, our analyses of the SARS-CoV-2 Brazil dataset (6) under the default-prior model identified São Paulo as the single major source of SARS-CoV-2 dispersal within Brazil; 84.6% of the dispersal events within Brazil were inferred to have originated from this area (*cf*. the second-ranking area, Rio de Janeiro, was the source of only 3.7% domestic dispersal events). Analyses under the preferred alternative-prior model reveal a strikingly disparate history of SARS-CoV-2 spread within Brazil: these analyses identified six areas to be significant sources of domestic dispersal, with only 19.5% of all the domestic dispersal events stemming from São Paulo (Figure 8, left panel). The impact of prior choice on the inferred number of dispersal events was pervasive across all of our empirical datasets. As might be expected from the default prior on the number of events, we infer a larger number of dispersal events under the alternative-prior model (Figure 8, right panel).

**Fig. 8.**
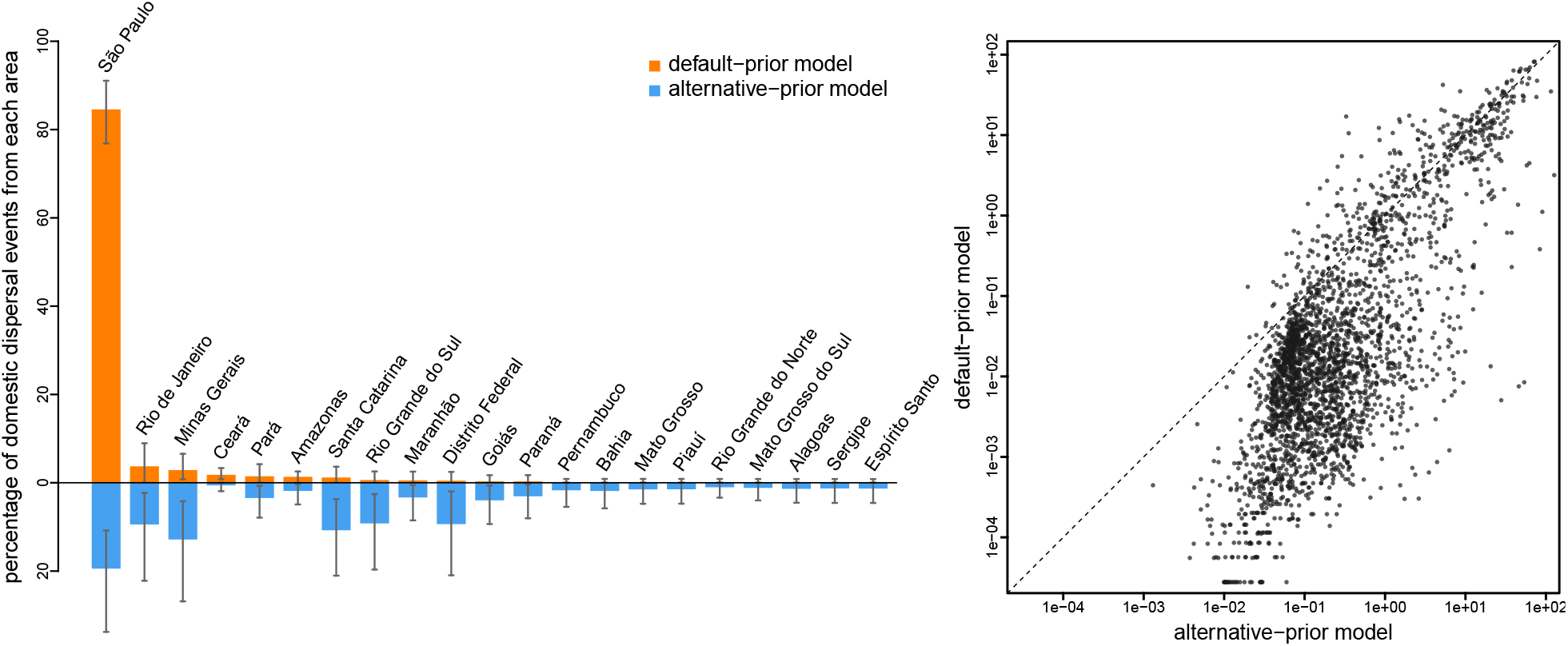
The impact of prior choice on the inferred number of dispersal events between areas. The left panel compares the number of dispersal events inferred under the default (orange) and alternative (blue) prior models for the SARS-CoV-2 Brazil dataset (6). Each bar indicates the estimated percentage of domestic dispersal events originating from each area within Brazil (mean [bar height] and 95% credible interval [whiskers]). Under the default-prior model, São Paulo is inferred to be the single major source of SARS-CoV-2 dispersal within Brazil, with 84.6% of all domestic dispersal events originating from this area. By contrast, our analyses of this dataset under the alternative-prior model reveals that only 19.5% of the domestic dispersal events originated from São Paulo, with five additional areas playing a significant role in domestic dispersal, including; two areas in Southeast Brazil (Minas Gerais [12.8%] and Rio de Janeiro [9.4%]), two areas in South Brazil (Santa Catarina [10.8%] and Rio Grande do Sul [9.2%]), and one area in Central-West Brazil (Distrito Federal [9.4%]). Note that the rank order of dispersal routes according to their inferred percentage of dispersal events differs between the default- and alternative-prior models. The right panel plots the number of dispersal events across each dispersal route inferred under the default (*y*-axis) and alternative (*x*-axis) prior models across all empirical datasets. (Uncertainty in these estimates is summarized in Figure S12.)

## Discussion

The development of Bayesian geographic models has the potential to transform our ability to study pathogen biology. The complexity of these geographic models is both an asset and a liability. It is an asset because it offers the potential to describe complex geographic processes. It is a liability because inference under these geographic models relies on minimal information (the geographic area in which each pathogen was sampled), rendering posterior estimates sensitive to the choice of priors. Moreover, the complexity of these geographic models obscures the biological interpretation of their parameters, making it difficult to formulate biologically sensible priors for those parameters. We suspect this underlies the fact that the vast majority of empirical phylodynamic geographic studies (≈93%) have assumed default priors.

In the present study, we have demonstrated that the default priors on the average dispersal rate and the number of dispersal routes implemented in `BEAST` imply biologically unrealistic assumptions about the geographic process (Figures 3 and 4). We have presented empirical evidence demonstrating that these default priors are in fact biologically unrealistic, *i*.*e*., they are strongly disfavored by all of the empirical datasets that we evaluated (Tables 1 and S2–S4). We have also demonstrated the consequences of these strongly misinformative priors; their use qualitatively changes our understanding of key aspects of pathogen geographic history, including inferences of relative dispersal rate between areas, the dispersal routes by which a disease spread across areas, the ancestral area in which an outbreak originated, and the number of dispersal events between areas (Figures 5–8).

Importantly, the unrealistic default priors not only distort inferences about key aspects of the geographic history of disease outbreaks, they also threaten to mislead public-health measures intended to mitigate these outbreaks. For example, inferences of the domestic spread of COVID-19 in Brazil under the (misspecified) default-prior model suggests that surveillance/testing and containment measures should be focused in a single area, whereas inferences under the (preferred) alternative-prior model reveal that effective mitigation requires deployment of these measures across multiple areas (Figure 8, left panel).

Our study highlights the need to develop and adopt best practices for empirical phylodynamic studies. All empirical datasets examined in our study (which are typical examples of empirical datasets, see Figure 1) decisively rejected the default priors in favor of the alternative priors (Tables 1, S2). Nevertheless, the alternative priors explored in our study are not intended as a panacea; that is, we are not advocating that the alternative priors explored herein be adopted indiscriminately in studies of discrete-geographic history. Rather, empirical studies should carefully consider the choice of priors and rigorously assess possible sensitivity of geographic inferences to those choices.

As illustrated in our study, numerous strategies are available to identify (and navigate) prior sensitivity. For example, robust Bayesian inference (25) and data cloning (26–29) can be used to identify when a given discrete-geographic inference is prior sensitive. Robust Bayesian inference involves performing a series of MCMC analyses—of the same dataset under the same inference model—where we iteratively change one (or more) priors of our discrete-geographic model: an analysis is prior sensitive when our posterior estimates differ for the candidate priors. Data cloning involves performing a series of MCMC analyses—under the same inference model with identical priors—where we iteratively increment the number of copies (“clones”) of our original dataset; these analyses can identify when the prior makes a relatively large contribution to the posterior. In cases where prior sensitivity is detected, we can adopt various approaches to navigate the choice of priors, including: (1) assessing the *relative* fit of candidate prior models (using Bayes factors), and; (2) assessing the *absolute* fit of candidate prior models (using posterior-predictive simulation). We have developed an interactive graphical utility, `PrioriTree` (https://github.com/jsigao/prioritree; 30), to facilitate adoption of these strategies, and thereby improve the reliability of geographic studies of disease outbreaks.

We are optimistic that rigorous empirical application of current phylodynamic models—with careful attention to identifying and navigating prior sensitivity—will greatly advance our understanding of pathogen biology and minimize the impact of infectious disease outbreaks.

## Materials and Methods

We provide details of the methods and analyses, as well as supplementary results, in the SI Appendix. We present supplementary figures and tables that directly complement the main text in SI Appendix section S1. We give a full description of the discrete-geographic model in SI Appendix section S2. We provide additional methodological details and supplementary results for our empirical analyses in SI Appendix section S3. All data, phylogenies, and code necessary to reproduce our results are available on Dryad (https://datadryad.org/stash/share/7Rd5kdTh7V66w9XefTuSeoui0LLv6LAwcY_5buMwUZU) and GitHub (https://github.com/jsigao/prior_misspecification_phylodynamic_biogeography).

## Data Availability

The sequence, sampling time and geography data used in this study, as well as the phylogenies we marginalized over or conditioned on in the biogeographic inference, are maintained in the GitHub repository (https://github.com/jsigao/prior_misspecification_phylodynamic_biogeography) and archived in the Dryad repository (https://datadryad.org/stash/share/vTbeDwLq2uSL9rL4NCe_Cocp2bY7BgWTI2tUgoNrLDA). Our repositories also contain BEAST XML scripts used to perform the phylodynamic analyses and R scripts used to post process the analyses and perform posterior-predictive simulation.

https://datadryad.org/stash/share/7Rd5kdTh7V66w9XefTuSeoui0LLv6LAwcY_5buMwUZU

## ACKNOWLEDGMENTS

This research was supported by the National Science Foundation grants DEB-0842181, DEB-0919529, DBI-1356737, and DEB-1457835 awarded to BRM, and the National Institutes of Health grant RO1GM123306-S awarded to BR.

## Footnotes

* The real constraint on the geographic model is that it must be *irreducible*. A model with fewer than *k* − 1 dispersal routes cannot be irreducible; however, a model with at least *k* − 1 dispersal routes is not guaranteed to be irreducible. See Supplemental Material for details.

† Note that the gamma prior on the average dispersal rate is referred to as the CTMC-rate reference prior in the BEAUti program used to generate input files for BEAST analyses.

## References

1. Lemey P, Rambaut A, Drummond AJ, Suchard MA (2009) Bayesian phylogeography finds its roots. PLoS Computational Biology 5(9):e1000520.

2. Edwards CJ, et al. (2011) Ancient hybridization and an Irish origin for the modern polar bear matriline. Current Biology 21(15):1251–1258.

3. Drummond AJ, Suchard MA, Xie D, Rambaut A (2012) Bayesian phylogenetics with BEAUti and the BEAST 1.7. Molecular Biology and Evolution 29(8):1969–1973.

4. Suchard MA, et al. (2018) Bayesian phylogenetic and phylodynamic data integration using BEAST 1.10. Virus Evolution 4(1):vey016.

5. Worobey M, et al. (2020) The emergence of SARS-CoV-2 in Europe and North America. Science 370(6516):564–570.

6. Candido DS, et al. (2020) Evolution and epidemic spread of SARS-CoV-2 in Brazil. Science 369(6508):1255–1260.

7. Lemey P, et al. (2021) Untangling introductions and persistence in COVID-19 resurgence in Europe. Nature 595(7869):713–717.

8. Kraemer MUG, et al. (2021) Spatiotemporal invasion dynamics of SARS-CoV-2 lineage B.1.1.7 emergence. Science 373(6557):889–895.

9. Alpert T, et al. (2021) Early introductions and transmission of SARS-CoV-2 variant B. 1.1. 7 in the United States. Cell 184(10):2595–2604.

10. Wilkinson E, et al. (2021) A year of genomic surveillance reveals how the SARS-CoV-2 pandemic unfolded in Africa. Science 374(6566):423–431.

11. du Plessis L, et al. (2021) Establishment and lineage dynamics of the SARS-CoV-2 epidemic in the UK. Science 371(6530):708–712.

12. Yang Z (2014) Molecular Evolution: a Statistical Approach. (Oxford University Press).

13. Bayes T (1763) LII. An essay towards solving a problem in the doctrine of chances. By the late Rev. Mr. Bayes, FRS communicated by Mr. Price, in a letter to John Canton, AMFR S. Philosophical Transactions of the Royal Society of London (53):370–418.

14. Jakeman E, Pusey P (1978) Significance of K distributions in scattering experiments. Physical Review Letters 40(9):546.

15. Kass RE, Raftery AE (1995) Bayes factors. Journal of the American Statistical Association 90(430):773–795.

16. Gelman A, Meng XL, Stern H (1996) Posterior predictive assessment of model fitness via realized discrepancies. Statistica Sinica pp. 733–760.

17. Bollback JP (2002) Bayesian model adequacy and choice in phylogenetics. Molecular Biology and Evolution 19(7):1171–1180.

18. Dash PK, et al. (2015) Complete genome sequencing and evolutionary phylogeography analysis of Indian isolates of Dengue virus type 1. Virus Research 195:124–134.

19. Wilfert L, et al. (2016) Deformed wing virus is a recent global epidemic in honeybees driven by Varroa mites. Science 351(6273):594–597.

20. Faria NR, et al. (2014) The early spread and epidemic ignition of HIV-1 in human populations. Science 346(6205):56–61.

21. Bedford T, et al. (2015) Global circulation patterns of seasonal influenza viruses vary with antigenic drift. Nature 523(7559):217.

22. Yao HW, et al. (2015) The spatiotemporal expansion of human Rabies and its probable explanation in mainland China, 2004-2013. PLoS Neglected Tropical Diseases 9(2):e0003502.

23. Gao J, May MR, Rannala B, Moore BR (2022) New phylogenetic models incorporating interval-specific dispersal dynamics improve inference of disease spread. Molecular Biology and Evolution 39(8):msac159.

24. Bedford T, et al. (2020) Cryptic transmission of SARS-CoV-2 in Washington state. Science 370(6516):571–575.

25. Berger JO (1994) An overview of robust Bayesian analysis. Test 3(1):5–124.

26. Robert CP (1993) Prior feedback: A Bayesian approach to maximum likelihood estimation. Computational Statistics 8:279–294.

27. Lele SR, Dennis B, Lutscher F (2007) Data cloning: easy maximum likelihood estimation for complex ecological models using Bayesian Markov chain Monte Carlo methods. Ecology Letters 10(7):551–563.

28. Ponciano JM, Taper ML, Dennis B, Lele SR (2009) Hierarchical models in ecology: confidence intervals, hypothesis testing, and model selection using data cloning. Ecology 90(2):356–362.

29. Ponciano JM, Burleigh JG, Braun EL, Taper ML (2012) Assessing parameter identifiability in phylogenetic models using data cloning. Systematic Biology 61(6):955–972.

30. Gao J, May MR, Rannala B, Moore BR (2022) PrioriTree: a utility for improving phylodynamic analyses in BEAST. bioRxiv.

