## Supplementary Material for "Model Misspecification Misleads Inference of the Spatial Dynamics of Disease Outbreaks"

### Supplemental Material for: Model Misspecification Misleads Inference of the Spatial Dynamics of Disease Outbreaks

#### Contents

**S1 Supplemental Figures and Tables for the Main Text** **S2**
S1.1 Priors on the Number of Dispersal Routes: Asymmetric Model . . . . . . . . . . . . . . . . S2
S1.2 Assessing the Fit of Default- and Alternative-Prior Models to Empirical Datasets . . . . . S3
S1.2.1 Using Bayes Factors to Assess Relative Model Fit . . . . . . . . . . . . . . . . . . . . S3
S1.2.2 Using Posterior-Predictive Simulation to Assess Absolute Model Fit . . . . . . . . S4
S1.3 The Impact of Prior Choice on the Inferred Biogeographic History . . . . . . . . . . . . . . S7

**S2 Additional Description of the Model** **S8**
S2.1 Irreducibility of the Discrete-Geographic Model . . . . . . . . . . . . . . . . . . . . . . . . S8
S2.1.1 Prior on the Dispersal-Route Indicator Vector, $P(\delta)$ . . . . . . . . . . . . . . . . . . S8
S2.1.2 The Conditional Prior on Each Dispersal Route . . . . . . . . . . . . . . . . . . . . . S11
S2.1.3 Consequences of Computing Bayes Factor with the Unconditional Prior . . . . . . S13
S2.1.4 Numerical Issue with Computing $\mathbb{1}_C(\delta)$ in BEAST . . . . . . . . . . . . . . . . . . . . . S14
S2.2 The Prior Probability of the Geographic Area at the Root . . . . . . . . . . . . . . . . . . . S16
S2.3 Joint and Sequential Bayesian Phylodynamic Inference . . . . . . . . . . . . . . . . . . . . S17

**S3 Analyses of Empirical Datasets** **S19**
S3.1 General Analysis Protocol . . . . . . . . . . . . . . . . . . . . . . . . . . . . . . . . . . . . S19
S3.2 Expanded Meta Summaries of Empirical Analyses . . . . . . . . . . . . . . . . . . . . . . . S24
S3.3 Expanded Dataset-Specific Summaries of Empirical Analyses . . . . . . . . . . . . . . . . . S31
S3.3.1 Dengue Virus . . . . . . . . . . . . . . . . . . . . . . . . . . . . . . . . . . . . . . . . S31
S3.3.2 Deformed Wing Virus . . . . . . . . . . . . . . . . . . . . . . . . . . . . . . . . . . . S42
S3.3.2.1 Lp fragment . . . . . . . . . . . . . . . . . . . . . . . . . . . . . . . . . . . . S43
S3.3.2.2 Rdrp fragment . . . . . . . . . . . . . . . . . . . . . . . . . . . . . . . . . . . S53
S3.3.2.3 Vp3 fragment . . . . . . . . . . . . . . . . . . . . . . . . . . . . . . . . . . . . S63
S3.3.3 HIV . . . . . . . . . . . . . . . . . . . . . . . . . . . . . . . . . . . . . . . . . . . . . S73
S3.3.3.1 Dataset A . . . . . . . . . . . . . . . . . . . . . . . . . . . . . . . . . . . . . . S74
S3.3.3.2 Dataset B . . . . . . . . . . . . . . . . . . . . . . . . . . . . . . . . . . . . . . S84
S3.3.3.3 Dataset C . . . . . . . . . . . . . . . . . . . . . . . . . . . . . . . . . . . . . . S94
S3.3.4 Influenza Virus . . . . . . . . . . . . . . . . . . . . . . . . . . . . . . . . . . . . . . . S104
S3.3.4.1 A/H3N2 Dataset . . . . . . . . . . . . . . . . . . . . . . . . . . . . . . . . . . S105
S3.3.4.2 B/Yamagata Dataset . . . . . . . . . . . . . . . . . . . . . . . . . . . . . . . . S115
S3.3.5 Rabies Virus . . . . . . . . . . . . . . . . . . . . . . . . . . . . . . . . . . . . . . . . S125
S3.3.6 SARS-CoV-2 Global . . . . . . . . . . . . . . . . . . . . . . . . . . . . . . . . . . . . S136
S3.3.7 SARS-CoV-2 B.1.1.7 US Dataset . . . . . . . . . . . . . . . . . . . . . . . . . . . . . S147
S3.3.8 SARS-CoV-2 Brazil . . . . . . . . . . . . . . . . . . . . . . . . . . . . . . . . . . . . . S158
S3.3.8.1 SchemeB Dataset . . . . . . . . . . . . . . . . . . . . . . . . . . . . . . . . . . S159
S3.3.8.2 SchemeC Dataset . . . . . . . . . . . . . . . . . . . . . . . . . . . . . . . . . . S169

### S1 Supplemental Figures and Tables for the Main Text

#### S1.1 Priors on the Number of Dispersal Routes: Asymmetric Model

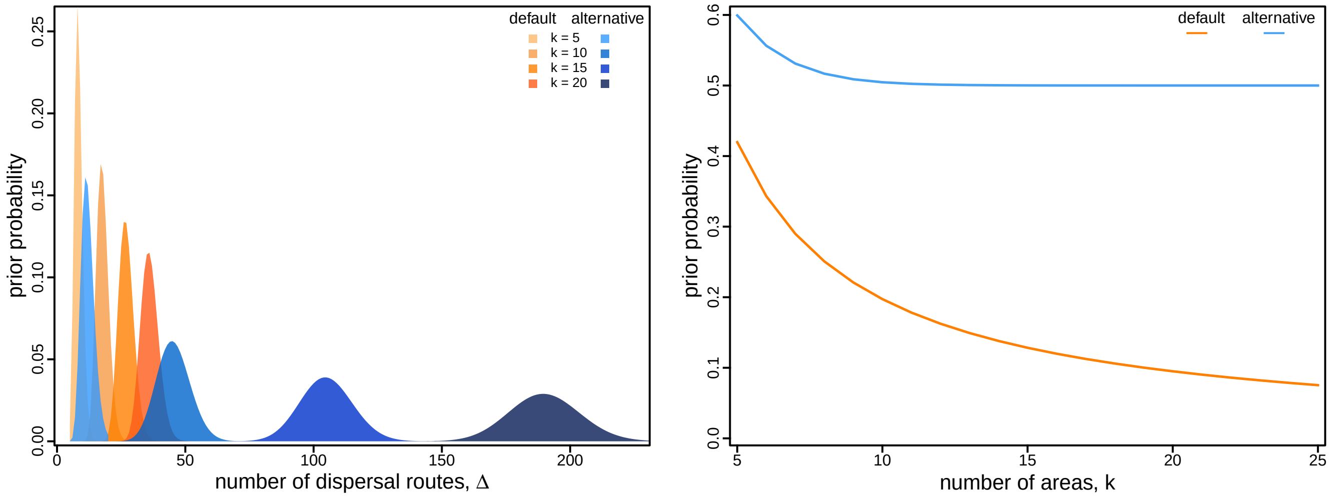


**Figure S1: Prior probability on dispersal routes under the asymmetric geographic model.** The left panel illustrates the default (orange) and alternative (blue) prior distributions on the total number of dispersal routes, $\Delta$, as a function of the number of areas, $k$. The default-prior distributions are highly focused on the minimal number of dispersal routes, $(k-1)$, whereas the alternative-prior distributions are centered on an intermediate number of dispersal routes (*i.e.*, the expected number of dispersal routes is half the maximum number). The right panel illustrates the prior probability under the default (orange) and alternative (blue) prior models that a given dispersal route exists (*i.e.*, $\delta_{ij} = 1$) as a function of the total number of areas, $k$. Under the default-prior model, the probability that a given dispersal route exists drops rapidly for moderately large (and common) values of $k$, whereas under the alternative-prior model, this probability remains constant for all values of $k$.

**Table S1: Default and alternative prior specifications.**

| Parameter | Model | Default | Alternative |
|---|---|---|---|
| Number of dispersal routes, $\Delta$ | Symmetric | $[\Delta - (k-1)] \sim \text{Pois}(\ln 2)$ | $[\Delta - (k-1)] \sim \text{Pois}(\lceil \frac{k(k-1)}{4} - (k-1) \rceil)$ |
| | Asymmetric | $\Delta \sim \text{Pois}(k-1)$ | $\Delta \sim \text{Pois}(\frac{k(k-1)}{2})$ |
| Average dispersal rate, $\mu$ | — | $\mu \sim \Gamma(0.5, T)$ (*i.e.*, CTMC-rate reference) | $\mu \sim \text{Exp}(1/\lambda)$ $\lambda \sim \Gamma(0.5, 0.5)$ |

#### S1.2 Assessing the Fit of Default- and Alternative-Prior Models to Empirical Datasets

##### S1.2.1 Using Bayes Factors to Assess Relative Model Fit

**Table S2: Assessing the relative fit of all eight candidate biogeographic models to the 14 empirical datasets.** For each dataset we computed the marginal likelihood for the eight candidate models corresponding to all possible combinations of: (1) default and alternative priors on the average rate of dispersal, $\mu$; (2) default and alternative priors on the number of dispersal routes, $\Delta$, and; (3) symmetric and asymmetric biogeographic models. For each dataset, we computed twice the log Bayes factors ($2 \ln \text{BF}$) between each model and the best model (*i.e.*, the model with the highest marginal likelihood; blue cells). For each dataset, $N$ is the number of sequences (*i.e.*, tips), and $k$ is the number of biogeographic areas.

| | | | $P(\mu)$ default | | | | $P(\mu)$ alternative | | | |
|---|---|---|---|---|---|---|---|---|---|---|
| | | | $P(\Delta)$ default | | $P(\Delta)$ alternative | | $P(\Delta)$ default | | $P(\Delta)$ alternative | |
| Dataset* | $N$ | $k$ | $Q_s$ | $Q_a$ | $Q_s$ | $Q_a$ | $Q_s$ | $Q_a$ | $Q_s$ | $Q_a$ |
| 1 | 62 | 23 | −86.33 | −80.67 | −47.80 | −46.60 | −10.54 | −10.93 | 0.00 | −1.81 |
| 2 | 209 | 8 | −32.61 | −27.53 | −26.00 | −23.16 | −9.22 | −1.48 | −1.98 | 0.00 |
| 3 | 183 | 7 | −86.43 | −95.38 | −83.16 | −90.38 | −20.13 | 0.00 | −20.21 | −4.68 |
| 4 | 96 | 7 | −29.79 | −29.46 | −25.20 | −25.10 | −2.72 | −1.88 | 0.00 | −0.96 |
| 5 | 792 | 8 | −277.75 | −279.98 | −275.74 | −278.64 | −36.55 | −5.69 | −26.30 | 0.00 |
| 6 | 927 | 10 | −343.40 | −288.65 | −340.35 | −291.39 | −46.33 | −3.86 | −42.45 | 0.00 |
| 7 | 466 | 8 | −264.34 | −218.29 | −262.26 | −216.95 | −6.45 | −8.62 | −3.26 | 0.00 |
| 8 | 1391 | 9 | −1223.33 | −1196.85 | −1125.69 | −1099.60 | −71.06 | −50.36 | −9.85 | 0.00 |
| 9 | 1240 | 9 | −957.61 | −921.84 | −870.86 | −840.76 | −114.79 | −49.62 | −51.45 | 0.00 |
| 10 | 141 | 18 | −122.47 | −95.49 | −82.89 | −77.24 | −22.36 | −8.27 | −0.88 | 0.00 |
| 11 | 1271 | 23 | −801.96 | −720.86 | −687.99 | −689.71 | −202.15 | 0.00 | −83.05 | −1.89 |
| 12 | 1908 | 22 | −659.04 | −524.88 | −515.28 | −492.04 | −248.4 | −16.66 | −140.19 | 0.00 |
| 13 | 1182 | 10 | −492.28 | −434.88 | −462.14 | −417.84 | −128.71 | −27.71 | −81.07 | 0.00 |
| 14 | 1182 | 22 | −316.08 | −294.59 | −275.22 | −272.16 | −145.04 | 0.00 | −50.15 | −1.35 |

* Dataset sources: 1) Dengue virus from Dash et al. (2015); 2–4) Deformed wing virus from Wilfert et al. (2016); 5–7) HIV from Faria et al. (2014); 8–9) Seasonal Influenza viruses from Bedford et al. (2015); 10) Rabies virus from Yao et al. (2015); 11) SARS-CoV-2 (Global) from Gao et al. (2022); 12) SARS-CoV-2 (B.1.1.7 USA) from Alpert et al. (2021), and; 13–14) SARS-CoV-2 (Brazil) from Candido et al. (2020).

##### S1.2.2 Using Posterior-Predictive Simulation to Assess Absolute Model Fit

**Table S3: Assessing the adequacy of all eight candidate biogeographic models for the 14 empirical datasets using the parsimony statistic.** Each row summarizes model adequacy for a given dataset (numbered as described in Table S2). For each dataset, $N$ is the number of sequences (*i.e.*, tips), and $k$ is the number of biogeographic areas. Each column lists the posterior-predictive $p$-values for a given prior model (notation follows Table S2) based on the parsimony summary statistic. Red $p$-values indicate that the model is inadequate at the 95% level.

| | | | $P(\mu)$ default | | | | $P(\mu)$ alternative | | | |
|---|---|---|---|---|---|---|---|---|---|---|
| | | | $P(\Delta)$ default | | $P(\Delta)$ alternative | | $P(\Delta)$ default | | $P(\Delta)$ alternative | |
| Dataset | $N$ | $k$ | $Q_s$ | $Q_a$ | $Q_s$ | $Q_a$ | $Q_s$ | $Q_a$ | $Q_s$ | $Q_a$ |
| 1 | 62 | 23 | **0.00** | **0.00** | **0.00** | **0.00** | 0.93 | 0.41 | 0.74 | 0.64 |
| 2 | 209 | 8 | **0.01** | **0.00** | **0.01** | **0.01** | 0.51 | 0.52 | 0.47 | 0.45 |
| 3 | 183 | 7 | **0.00** | **0.00** | **0.00** | **0.00** | 0.71 | 0.09 | 0.64 | 0.10 |
| 4 | 96 | 7 | **0.01** | **0.00** | **0.01** | **0.00** | 0.46 | 0.26 | 0.42 | 0.26 |
| 5 | 792 | 8 | **0.00** | **0.00** | **0.00** | **0.00** | 0.08 | 0.52 | 0.09 | 0.58 |
| 6 | 927 | 10 | **0.00** | **0.00** | **0.00** | **0.00** | 0.21 | 0.40 | 0.23 | 0.40 |
| 7 | 466 | 8 | **0.00** | **0.00** | **0.00** | **0.00** | 0.07 | 0.09 | 0.08 | 0.08 |
| 8 | 1391 | 9 | **0.00** | **0.00** | **0.00** | **0.00** | 0.68 | 0.14 | 0.47 | 0.11 |
| 9 | 1240 | 9 | **0.00** | **0.00** | **0.00** | **0.00** | 0.75 | 0.34 | 0.57 | 0.27 |
| 10 | 141 | 18 | **0.00** | **0.00** | **0.00** | **0.00** | 0.62 | 0.17 | 0.51 | 0.29 |
| 11 | 1271 | 23 | **0.00** | **0.00** | **0.00** | **0.00** | 0.50 | 0.44 | 0.14 | 0.44 |
| 12 | 1908 | 22 | **0.00** | **0.01** | **0.00** | **0.00** | 0.78 | 0.60 | 0.85 | 0.89 |
| 13 | 1182 | 10 | **0.00** | **0.00** | **0.00** | **0.00** | 0.42 | 0.53 | 0.31 | 0.59 |
| 14 | 1182 | 22 | **0.00** | **0.00** | **0.00** | **0.00** | 0.94 | 0.87 | 0.89 | 0.95 |

**Table S4: Assessing the adequacy of all eight candidate biogeographic models for the 14 empirical datasets using the tip-wise multinomial statistic.** Each row summarizes model adequacy for a given dataset (numbered as described in Table S2). For each dataset, $N$ is the number of sequences (*i.e.*, tips), and $k$ is the number of biogeographic areas. Each column lists the posterior-predictive $p$-values for a given prior model (notation follows Table S2) based on the tip-wise multinomial statistic. Red $p$-values indicate that the model is inadequate at the 95% level.

| | | | $P(\mu)$ default | | | | $P(\mu)$ alternative | | | |
|---|---|---|---|---|---|---|---|---|---|---|
| | | | $P(\Delta)$ default | | $P(\Delta)$ alternative | | $P(\Delta)$ default | | $P(\Delta)$ alternative | |
| Dataset | $N$ | $k$ | $Q_s$ | $Q_a$ | $Q_s$ | $Q_a$ | $Q_s$ | $Q_a$ | $Q_s$ | $Q_a$ |
| 1 | 62 | 23 | **1.00** | **1.00** | **1.00** | **1.00** | 0.46 | 0.94 | 0.44 | 0.59 |
| 2 | 209 | 8 | **0.98** | 0.97 | 0.96 | 0.96 | 0.70 | 0.75 | 0.59 | 0.57 |
| 3 | 183 | 7 | 0.91 | 0.95 | 0.88 | 0.94 | 0.18 | 0.89 | 0.21 | 0.87 |
| 4 | 96 | 7 | **1.00** | **1.00** | **1.00** | **1.00** | 0.92 | 0.96 | 0.89 | 0.94 |
| 5 | 792 | 8 | **0.98** | 0.93 | 0.97 | 0.93 | 0.87 | 0.76 | 0.81 | 0.60 |
| 6 | 927 | 10 | **1.00** | **1.00** | **1.00** | **1.00** | **0.98** | **0.99** | 0.96 | 0.94 |
| 7 | 466 | 8 | **1.00** | **0.99** | **1.00** | **0.99** | 0.96 | 0.95 | 0.96 | 0.95 |
| 8 | 1391 | 9 | **1.00** | **1.00** | **1.00** | **1.00** | **1.00** | **1.00** | **1.00** | **1.00** |
| 9 | 1240 | 9 | **1.00** | **1.00** | **1.00** | **1.00** | 0.39 | **0.99** | 0.40 | **0.99** |
| 10 | 141 | 18 | **1.00** | **1.00** | **1.00** | **1.00** | 0.97 | **1.00** | 0.92 | 0.96 |
| 11 | 1271 | 23 | **1.00** | **1.00** | **1.00** | **1.00** | 0.77 | **0.99** | 0.51 | 0.90 |
| 12 | 1908 | 22 | 0.97 | 0.96 | 0.96 | 0.95 | 0.43 | 0.69 | 0.19 | 0.19 |
| 13 | 1182 | 10 | 0.91 | 0.95 | 0.91 | 0.95 | 0.40 | 0.67 | 0.20 | 0.52 |
| 14 | 1182 | 22 | 0.89 | 0.95 | 0.84 | 0.89 | 0.06 | 0.20 | **0.02** | 0.10 |

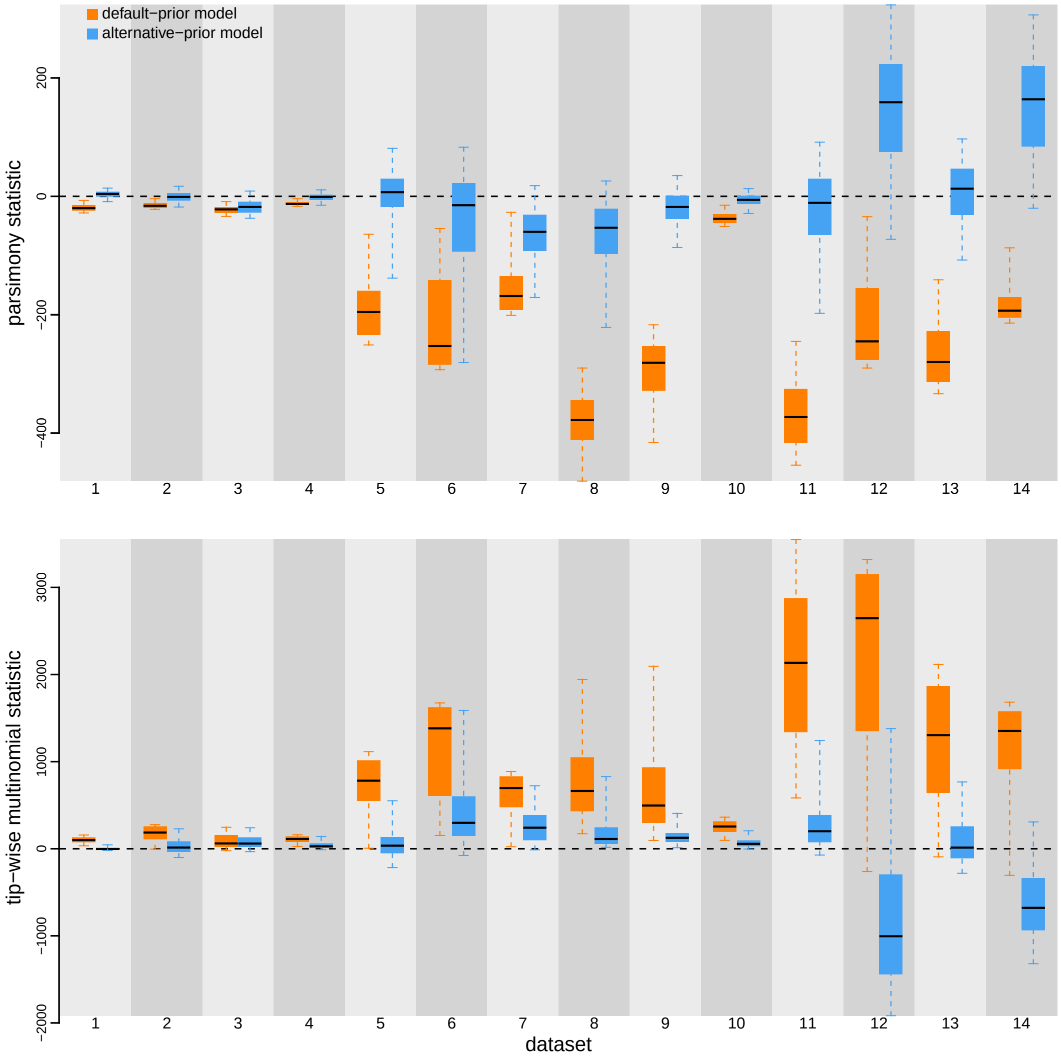


**Figure S2: Posterior-predictive distributions of the parsimony statistic (top panel) and the tip-wise multinomial statistic (bottom panel) under the preferred default- and alternative-prior models for all datasets.** Each column depicts estimates for one of the 14 datasets (numbered as described in Table S2). Within each column, the pair of boxplots depicts the posterior-predictive distributions of the summary statistic under the default (orange) and alternative (blue) prior models: the center of each box is the median predictive value of the summary statistic; the box and whiskers indicate the corresponding 50% and 95% posterior-predictive intervals, respectively. The horizontal dashed line indicates when the simulated and observed datasets produce identical value for the summary statistic. A model is judged to be inadequate (*i.e.*, incapable of generating geographic datasets that are similar to the observed data) if its 95% posterior-predictive interval does not overlap with the dashed line. Importantly, posterior-predictive simulation allows us to compare the absolute fit of the candidate models to the 14 geographic datasets: the preferred default prior models (orange) are always inadequate, whereas the preferred alternative prior models (blue) are almost always adequate.

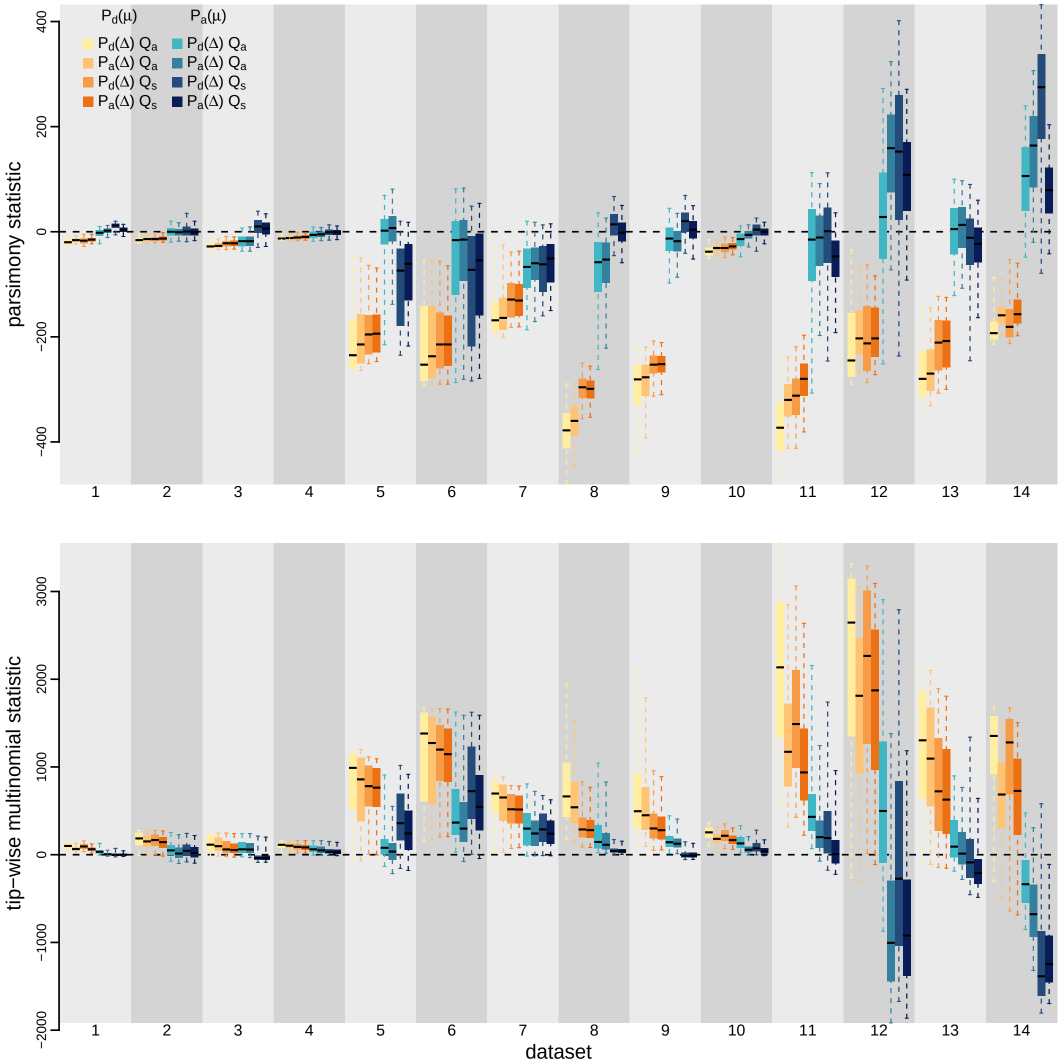


**Figure S3: Posterior-predictive distributions of the parsimony statistic (top panel) and the tip-wise multinomial statistic (bottom panel) under each of the eight prior models for all datasets.** Each column depicts estimates for one of the 14 datasets (numbered as described in Table S2). Within each column, the set of 8 boxplots depicts the posterior-predictive distributions of the summary statistic under each of the 8 prior models: the center of each box is the median predictive value of the summary statistic; the box and whiskers indicate the corresponding 50% and 95% posterior-predictive intervals, respectively. The horizontal dashed line indicates when the simulated and observed datasets produce identical value for the summary statistic. A model is judged to be inadequate (*i.e.*, incapable of generating geographic datasets that are similar to the observed data) if its 95% posterior-predictive interval does not overlap with the dashed line.

#### S1.3 The Impact of Prior Choice on the Inferred Biogeographic History

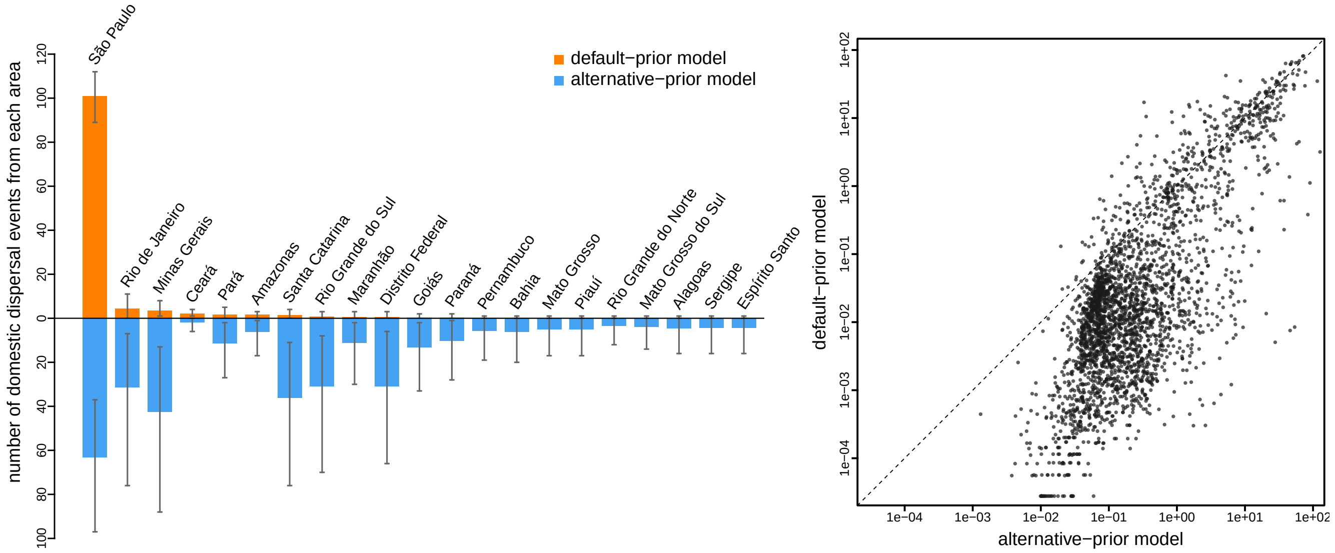


**Figure S4: The impact of prior choice on the inferred number of dispersal events between areas.** The left panel compares the number of dispersal events inferred under the default (orange) and alternative (blue) prior models for the SARS-CoV-2 Brazil dataset (Candido et al. 2020). Each bar indicates the estimated number of domestic dispersal events originating from each area within Brazil (mean [bar height] and 95% credible interval [whiskers]). Note that this figure complements Figure 8 in the main text as here we plot the absolute number of dispersal events instead of the percentage (as shown in the main text). Under the default-prior model, São Paulo is inferred to be the single major source of SARS-CoV-2 dispersal within Brazil, with 101.0 domestic dispersal events originating from this area, while merely 18.4 events originating from the other areas combined. By contrast, our analyses of this dataset under the alternative-prior model reveals that only 63.1 domestic dispersal events originated from São Paulo, with five additional areas playing a significant role in domestic dispersal, including; two areas in Southeast Brazil (Minas Gerais [42.5 events] and Rio de Janeiro [31.3 events]), two areas in South Brazil (Santa Catarina [36.1 events] and Rio Grande do Sul [31.0 events]), and one area in Central-West Brazil (Distrito Federal [30.9 events]). Note that the rank order of dispersal routes according to their inferred number of dispersal events differs between the default- and alternative-prior models. The right panel plots the number of dispersal events across each dispersal route inferred under the default ($y$-axis) and alternative ($x$-axis) prior models across all empirical datasets. On average, the inferred number of dispersal events under the alternative-prior model is larger than that inferred under the default-prior model.

### S2 Additional Description of the Model

#### S2.1 Irreducibility of the Discrete-Geographic Model

Discrete-geographic models describe the dispersal process over the phylogeny, $\Psi$, as a continuous-time Markov chain (CTMC). For a geographic history with $k$ discrete areas, the dispersal process is fully specified by a $k \times k$ instantaneous-rate matrix, $Q$, where an (off-diagonal) element of the matrix, $q_{ij}$, is the instantaneous rate of change between states $i$ and $j$. In the Bayesian approach developed by Lemey et al. (2009), $q_{ij}$ is specified as the product of a relative rate of dispersal between areas $i$ and $j$, $r_{ij}$, and a dispersal-route indicator variable $\delta_{ij}$ ($\delta_{ij} = 1$ or 0).

The possibility that any element of the $Q$ matrix can be zero under this model poses some computational challenges. Indeed, many properties of Markov chains (*e.g.*, irreducibility, ergodicity, and stationarity) assumed by conventional procedures for computing the phylogenetic likelihood only hold when the instantaneous rates are strictly positive (or at least some elements in each row and column of $Q$ are strictly positive). However, when any $q_{ij}$ can be zero, the Markov chain may not be irreducible; *i.e.*, some states in the Markov chain may be incapable of reaching (communicating with) some other states. For example, if $q_{i\bullet}$ (the i-th row of $Q$) is all zeros, then the rate of dispersal from area $i$ to any other area becomes zero, such that the pathogen can never disperse out of area $i$; alternatively, if $q_{\bullet j}$ (the j-th column of $Q$) is all zeros, then no pathogen can ever disperse into area $j$. In these cases, the corresponding Markov chain may not be stationary, and therefore some of the conventional steps we take in computing the phylogenetic likelihood—including rescaling $Q$ such that the average rate of dispersal between all areas is $\mu$ and using the stationary distribution $\pi$ as the prior probability that the dispersal process starts in each of the geographic areas at the root of the tree—become invalid.

Moreover, reducible geographic models are also biologically unreasonable. When modeling the dispersal process, we imagine the pathogen originates from one of the geographic areas at the root of the phylogeny and then disperses to the other geographic areas directly or indirectly over the phylogeny. A reducible model represents a process where there are some groups of areas that the pathogen can never reach (when the rates of dispersal into those groups of areas are all zero) from any area that is not in the same group, and/or a process where there are some groups of areas that the pathogen can never leave (when the rates of dispersal out of those groups of areas are all zero). Such a reducible model is unrealistic given that we have pathogen sequences sampled from each of the geographic areas in the model, and given that we always assume that the pathogen giving rise to the epidemic or pandemic under study should have originated only once and from one of the geographic areas where each area is associated with a positive probability *a priori*.

Accordingly, reducible geographic models should be avoided due to both practical and theoretical considerations. Indeed, `BEAST` does prevent sampling any reducible models in the MCMC. Below, we will first explain in theory how `BEAST` assigns zero prior probability on reducible models, and then describe how that is achieved in practice.

##### S2.1.1 Prior on the Dispersal-Route Indicator Vector, $P(\delta)$

For the ease of description, recall that (as described in Figure 2) a given dispersal-route indicator vector $\delta$ can also be represented as a graph where each vertex corresponds to a geographic area and each edge corresponds to a dispersal route. When the geographic model is symmetric, the graph is undirected so that each edge is bidirectional and there is at most one edge between areas $i$ and $j$ (Figure 2); for an asymmetric model, the corresponding graph is directed so that at most two directional edges may exist between each pair of areas. For the geographic model to be irreducible, the corresponding graph must be strongly connected; *i.e.*, every vertex must be reachable from every other vertex in the graph. Let $\boldsymbol{\mathcal{G}}$ denote the set of all possible graphs with $k$ vertices (*i.e.*, geographic areas), and $\boldsymbol{\mathcal{G}}_C$ denote the subset of $\boldsymbol{\mathcal{G}}$ formed by all the strongly connected graphs. Each element of $\boldsymbol{\mathcal{G}}$, denoted as $\mathcal{G}$, thus corresponds to a unique $\delta$, and *vice versa*. Let $C$ denotes the condition that $\mathcal{G}$ is strongly connected; *i.e.*, $C \equiv \mathcal{G} \in \boldsymbol{\mathcal{G}}_C$.

Lemey et al. (2009) impose a prior on $\delta$ (and therefore the corresponding geographic models) by: (1) placing a prior on the total number of dispersal routes, $\Delta$; (2) assuming that each $\delta$ configuration with a given value of $\Delta$ are equiprobable, and; (3) putting zero probability on each $\delta$ that corresponds to a reducible geographic model (*i.e.*, $P(\mathcal{G} \notin \boldsymbol{\mathcal{G}}_C) = 0$). Therefore, the prior probability on the dispersal-route indicator vector, $P(\delta)$, is computed as:

$$\begin{aligned}
P(\delta) &= \sum_{\Delta} \left[\mathbb{1}_C(\delta) P(\delta \mid \Delta) P(\Delta)\right] \\
P(\delta) &= \mathbb{1}_C(\delta) P(\delta \mid \Delta_\delta) P(\Delta_\delta) + \sum_{\Delta \neq \Delta_\delta} \left[\mathbb{1}_C(\delta) P(\delta \mid \Delta) P(\Delta)\right] \\
P(\delta) &= \mathbb{1}_C(\delta) P(\delta \mid \Delta_\delta) P(\Delta_\delta),
\end{aligned} \tag{S1}$$

where $\mathbb{1}_C(\delta)$ is an indicator function whose value is one when $\delta$ corresponds to a strongly connected graph (*i.e.*, an irreducible model) and zero otherwise, $\Delta_\delta$ is the number of dispersal routes indicated by $\delta$ (*i.e.*, $\Delta_\delta = \sum \delta$), $P(\delta \mid \Delta)$ is the probability of each $\delta$ with a given value of $\Delta$, and $P(\Delta)$ is the prior on the total number of dispersal routes (*e.g.*, the offset Poisson prior for the symmetric geographic model as described in the main text). Note that $\sum_{\Delta \neq \Delta_\delta} \left[P(\Delta) P(\delta \mid \Delta) \mathbb{1}_C(\delta)\right]$ disappears in the derivation as $P(\delta \mid \Delta) = 0$ when $\Delta \neq \Delta_\delta$; *i.e.*, there is no chance of getting $\delta$ as the dispersal-route indicator vector if the number of dispersal routes is not $\Delta_\delta$.

Below, we will describe how each of the three terms in Equation (S1) is computed. Before doing so, however, it is important to keep in mind that the exact full prior probability may not be computed in MCMC; instead, it is sufficient to compute the prior ratio $\frac{P(\delta')}{P(\delta)}$ whenever a new configuration of the dispersal-route indicator vector $\delta'$ is proposed during the MCMC. Specifically, the prior ratio is computed as:

$$\frac{P(\delta')}{P(\delta)} = \frac{\mathbb{1}_C(\delta')}{\mathbb{1}_C(\delta)} \times \frac{P(\delta' \mid \Delta_{\delta'})}{P(\delta \mid \Delta_\delta)} \times \frac{P(\Delta_{\delta'})}{P(\Delta_\delta)}. \tag{S2}$$

Therefore, it is possible that some terms of Equation (S1) do not need to be computed at some steps of the MCMC as the corresponding terms cancel out in Equation (S2). Nevertheless, computing the exact prior probability following Equation (S1) is still required when we calculate the Bayes factor support for each dispersal route. Accordingly, below when describing the computation of each of the three terms, we present both the full calculation—as shown in Equation (S1) to pave the way for describing the calculation of the Bayes factor—and also the ratio calculation as shown in Equation (S2) to establish a direct connection with the implementation and computation in `BEAST`.

##### Computing the indicator function $\mathbb{1}_C(\delta)$

By default, `BEAST` initializes the $Q$ matrix with each dispersal-route indicator $\delta_{ij}$ set to one. During the MCMC, `BEAST` employs the `bitFlipOperator` proposal to update the dispersal-route indicator vector $\delta$. This proposal randomly picks a $\delta_{ij}$ to flip—*i.e.*, to change its value to zero if its current value is one, and *vice versa*—so that in principle all possible configurations of $\delta$ can be visited in the MCMC.

Each time a new dispersal-route indicator vector $\delta'$ is proposed, `BEAST` checks whether the resulting $Q$ matrix is irreducible. Specifically, this is achieved by computing a transition-probability matrix $P$ as: $P = \exp(Qd)$, where the default value of $d$ is set to be one, and then checks if any element of the $P$ matrix, $p_{ij}$, is zero. This is effectively checking whether each area can reach every other area with positive probability, and if so, the geographic model determined by the proposed $\delta'$ is irreducible (*i.e.*, the corresponding graph is strongly connected). Otherwise, if the check fails, the proposed $\delta'$ will be rejected so that no reducible geographic model (*i.e.*, graph that is not strongly connected) will ever be sampled in the MCMC.

The indicator function $\mathbb{1}_C(\delta)$ can thus be defined as:

$$\mathbb{1}_C(\delta) = \begin{cases} 1 & \text{if } \mathcal{G} \in \boldsymbol{\mathcal{G}_C} \\ 0 & \text{if } \mathcal{G} \notin \boldsymbol{\mathcal{G}_C} \end{cases}. \tag{S3}$$

The prior ratio for the indicator-function term $\frac{\mathbb{1}_C(\delta')}{\mathbb{1}_C(\delta)}$ is either one (when $\mathcal{G}' \in \boldsymbol{\mathcal{G}_C}$, where $\mathcal{G}'$ denotes the graph corresponding to $\delta'$) or zero (when $\mathcal{G}' \notin \boldsymbol{\mathcal{G}_C}$).

**Computing the prior probability of $\delta$ given $\Delta$, $P(\delta \mid \Delta)$**

As all $\delta$ with a given value of $\Delta$ are assumed to be equiprobable, $P(\delta \mid \Delta)$ is simply computed as:

$$P(\delta \mid \Delta) = \frac{1}{|\boldsymbol{\mathcal{G}}^{\boldsymbol{\Delta}}|}, \tag{S4}$$

where $\boldsymbol{\mathcal{G}}^{\boldsymbol{\Delta}}$ denotes the subset of $\boldsymbol{\mathcal{G}}$ formed by all the graphs with $k$ vertices and $\Delta$ edges, and $|\boldsymbol{\mathcal{G}}^{\boldsymbol{\Delta}}|$ denotes the cardinality of that subset (*i.e.*, the number of graphs in that subset). $|\boldsymbol{\mathcal{G}}^{\boldsymbol{\Delta}}|$ is then computed as $\binom{\Delta_{\max}}{\Delta}$, where $\Delta_{\max}$ represents the number of possible dispersal routes (*i.e.*, the length of vector $\delta$, which is $\binom{k}{2}$ for the symmetric model and $k \times (k-1)$ for the asymmetric model).

Note that `BEAST` does not compute $P(\delta \mid \Delta)$ directly; instead, only the prior ratio $\frac{P(\delta' \mid \Delta')}{P(\delta \mid \Delta)}$ is computed upon a `bitFlipOperator` proposal that proposes a new dispersal-route indicator vector $\delta'$ which in turn is associated with a new number of dispersal routes $\Delta'$. Specifically, the prior ratio is $\frac{\Delta_{\max}-\Delta+1}{\Delta}$ when a dispersal route is proposed to be switched off, and $\frac{\Delta+1}{\Delta_{\max}-\Delta}$ when a dispersal route is proposed to be switched on. Specifically, the prior ratio is:

$$\frac{P(\delta' \mid \Delta')}{P(\delta \mid \Delta)} = \frac{\frac{1}{\binom{\Delta_{\max}}{\Delta'}}}{\frac{1}{\binom{\Delta_{\max}}{\Delta}}} = \begin{cases} \frac{\Delta_{\max}-\Delta+1}{\Delta} & \text{if } \Delta' = \Delta - 1; \textit{i.e.}, \text{ an indicator is proposed to be switched off} \\ \frac{\Delta+1}{\Delta_{\max}-\Delta} & \text{if } \Delta' = \Delta + 1; \textit{i.e.}, \text{ an indicator is proposed to be switched on} \end{cases}. \tag{S5}$$

**Computing the prior on the number of dispersal routes, $P(\Delta)$**

$P(\Delta)$ is the only term in Equation (S1) that is explicitly specified by the user and is one of the two focal priors in the geographic model that we have discussed extensively in the main text. In principle, this prior can take the form of any arbitrary discrete probability distribution; here we adopt a Poisson distribution, as used by empirical studies following Lemey et al. (2009).

To compute the full prior probability for the calculation of the Bayes factor support for each dispersal route, $P(\Delta)$ needs to be right truncated when $P(\Delta)$ is assumed to be a Poisson distribution. In principle, $\Delta$ can never take any value that is greater than the length of the indicator vector; *i.e.*, without considering the irreducibility and the specific form of the prior distribution, the range of possible values of $\Delta$ is $\{0, 1, \ldots, \binom{k}{2}\}$ when the model is symmetric and $\{0, 1, \ldots, k \times (k-1)\}$ when the model is asymmetric. Given that the range of a Poisson random variable is $[0, \infty)$, this Poisson distribution needs to be right truncated at the upper bound of the range of $\Delta$.

For example, for a symmetric model with two geographic areas, the default prior on the number of dispersal routes is an offset Poisson prior, where the offset—denoted $\beta$—is one ($\beta = k - 1$) and the rate of the Poisson $\lambda = \ln(2)$. In that case, at least one *and* at most one dispersal route $\delta$ must exist between the two areas, so the prior probability $P(\delta = 1)$ should be 1 instead of $f(\Delta = 1; \lambda = \ln(2), \beta = 1) = 0.5$. This discrepancy is reconciled when recognizing that the Poisson distribution is right truncated so $g(\Delta = 1; \lambda = \ln(2), \beta = 1) = 1$, where $g(\Delta; \lambda, \beta)$ denotes the right-truncated offset Poisson distribution.

More generally, $g(\Delta;\lambda,\beta)$ is computed as:

$$g(\Delta;\lambda,\beta) = \begin{cases} \frac{f(\Delta;\lambda,\beta)}{F(\Delta_{\max}-\beta;\lambda,\beta)} & \text{if } \Delta \in \boldsymbol{\Delta} = \{\beta, \beta+1, \ldots, \Delta_{\max}\} \\ 0 & \text{if } \Delta > \Delta_{\max} \text{ or } \Delta < \beta \end{cases}, \tag{S6}$$

where $\boldsymbol{\Delta}$ denotes the set of possible values of $\Delta$ and $F(\Delta_{\max} - \beta; \lambda, \beta)$ is the cumulative distribution function (CDF) of the offset Poisson distribution over $\boldsymbol{\Delta} - \beta$. Specifically, $\boldsymbol{\Delta} = \{k-1, k, \ldots, \binom{k}{2}\}$ for the symmetric model (as $\beta = k-1$) and $\boldsymbol{\Delta} = \{0, 1, \ldots, k \times (k-1)\}$ for the asymmetric model (as there is no offset; *i.e.*, $\beta = 0$; Edwards et al. 2011).

Note that as we specify $\lambda$ as a fixed variable in both the default and alternative Poisson priors on $\Delta$, the denominator in Equation (S6) (*i.e.*, the CDF) is a constant value for a given dataset and prior. Therefore, the denominator may be ignored (as is done in BEAST) when computing the prior ratio during the MCMC, which is computed as:

$$\frac{P(\Delta_{\delta'})}{P(\Delta_{\delta})} = \frac{f(\Delta';\lambda,\beta)}{f(\Delta;\lambda,\beta)}. \tag{S7}$$

##### S2.1.2 The Conditional Prior on Each Dispersal Route

Recall that for each dispersal-route indicator, the Bayes factor support is computed as:

$$\text{BF}_{ij} = \frac{P(\delta_{ij} = 1 \mid G)}{1 - P(\delta_{ij} = 1 \mid G)} \div \frac{P(\delta_{ij} = 1)}{1 - P(\delta_{ij} = 1)}, \tag{S8}$$

where $P(\delta_{ij} = 1 \mid G)$ is the posterior probability that the dispersal route from area $i$ to area $j$ exists; *i.e.*, the probability of $\delta_{ij} = 1$ given the observed geographic data $G$, which is computed as the proportion of MCMC samples for which $\delta_{ij} = 1$. $P(\delta_{ij} = 1)$ denotes the prior probability that the dispersal route exists, which—without conditioning on the irreducibility—can be derived as:

$$\begin{aligned} P(\delta_{ij} = 1) &= \sum_{\Delta} P(\delta_{ij} = 1 \mid \Delta) P(\Delta) \\ &= \sum_{\Delta} \frac{\Delta}{\Delta_{\max}} P(\Delta) \\ &= \frac{1}{\Delta_{\max}} \sum_{\Delta} \Delta P(\Delta) \\ &= \frac{\sum_{\Delta} \Delta f(\Delta;\lambda,\beta)}{\Delta_{\max}} \\ &= \frac{\mathbb{E}(\Delta)}{\Delta_{\max}} = \frac{\lambda+\beta}{\Delta_{\max}}. \end{aligned} \tag{S9}$$

A form of this equation for the symmetric model is presented as Equation (7) of Lemey et al. (2009), and is used in Spread (a helper program that most of the empirical biogeographic studies use to post process the output of BEAST biogeographic analyses; Bielejec et al. 2011, 2016) when computing the Bayes factor support for each dispersal route. Note that the last few steps of the above derivation, *e.g.*, $\mathbb{E}(\Delta) = \lambda + \beta$, make the implicit (and incorrect) assumption that $P(\Delta)$ is an offset Poisson distribution *without* the right truncation; *i.e.*, it assumes the upper bound of $\boldsymbol{\Delta}$ is $\infty$ instead of $\Delta_{\max}$.

Recall that $C \equiv \mathcal{G} \in \boldsymbol{\mathcal{G}}_C$; let $P_C(\delta_{ij} = 1) \equiv P(\delta_{ij} = 1 \mid C)$, denoting the prior probability that a given dispersal route exists conditioning on the irreducibility of the corresponding models (*i.e.*, the corresponding geographic graphs are always strongly connected). By replacing the unconditional prior probability in Equation (S8) with the conditional prior probability, the Bayes factor support conditioning on the graphs being always strongly connected, $\text{BF}_{ij}^C$, therefore is:

$$\text{BF}_{ij}^{C} = \frac{P(\delta_{ij} = 1 \mid G)}{1 - P(\delta_{ij} = 1 \mid G)} \div \frac{P_C(\delta_{ij} = 1)}{1 - P_C(\delta_{ij} = 1)}. \tag{S10}$$

Note that the posterior probability $P(\delta_{ij} = 1 \mid G) = P(\delta_{ij} = 1 \mid G, C)$, as the strongly connected condition is already implied by observing $G$; *i.e.*, the fact that we have observed all the $k$ areas defined in the geographic data $G$ implies that the geographic model must be irreducible, so the posterior probabilities used to compute the Bayes factor in Equations (S8) and (S10) are identical.

The conditional prior probability $P_C(\delta_{ij} = 1)$ can be computed similarly to Equation (S9) as:

$$P_C(\delta_{ij} = 1) = \frac{1}{\Delta_{\max}} \sum_{\Delta} \Delta P(\Delta \mid C), \tag{S11}$$

where $P(\Delta \mid C)$ represents the prior probability of $\Delta$ given the corresponding graphs are all strongly connected. This conditional probability can then be computed as:

$$\begin{aligned} P(\Delta \mid C) &= \frac{P(C \mid \Delta)P(\Delta)}{P(C)} \\ &= \frac{P(C \mid \Delta)P(\Delta)}{\sum_{\Delta} P(C \mid \Delta)P(\Delta)}, \end{aligned} \tag{S12}$$

where $P(C \mid \Delta)$ denotes the probability that a graph is strongly connected given $\Delta$ edges (and $k$ vertices). $P(C \mid \Delta)$ can then be computed as the ratio of the number of such strongly connected graphs, $|\boldsymbol{\mathcal{G}}_C^{\Delta}|$, over the number of all graphs with $\Delta$ edges (and $k$ vertices), $|\boldsymbol{\mathcal{G}}^{\Delta}|$, as each graph is assumed to be equiprobable *a priori*; *i.e.*, it is computed as:

$$P(C \mid \Delta) = \frac{|\boldsymbol{\mathcal{G}}_C^{\Delta}|}{|\boldsymbol{\mathcal{G}}^{\Delta}|}. \tag{S13}$$

$|\boldsymbol{\mathcal{G}}^{\Delta}|$ is $\binom{\Delta_{\max}}{\Delta}$. Combining the equations presented above, we obtain:

$$\begin{aligned} P_C(\delta_{ij} = 1) &= \frac{1}{\Delta_{\max}} \left( \frac{\sum_{\Delta} \Delta \frac{|\boldsymbol{\mathcal{G}}_C^{\Delta}|}{|\boldsymbol{\mathcal{G}}^{\Delta}|} g(\Delta; \lambda, \beta)}{\sum_{\Delta} \frac{|\boldsymbol{\mathcal{G}}_C^{\Delta}|}{|\boldsymbol{\mathcal{G}}^{\Delta}|} g(\Delta; \lambda, \beta)} \right) \\ &= \frac{1}{\Delta_{\max}} \left( \frac{\sum_{\Delta \in \boldsymbol{\Delta}} \Delta \frac{|\boldsymbol{\mathcal{G}}_C^{\Delta}|}{|\boldsymbol{\mathcal{G}}^{\Delta}|} f(\Delta; \lambda, \beta)}{\sum_{\Delta \in \boldsymbol{\Delta}} \frac{|\boldsymbol{\mathcal{G}}_C^{\Delta}|}{|\boldsymbol{\mathcal{G}}^{\Delta}|} f(\Delta; \lambda, \beta)} \right). \end{aligned} \tag{S14}$$

We provide functions that compute $P_C(\delta_{ij} = 1)$ following Equation (S14)—as well as the intermediate terms $P(C \mid \Delta)$ and $P(\Delta \mid C)$—in an R script available in our GitHub and Dryad repositories. The calculation of $|\boldsymbol{\mathcal{G}}_C^{\Delta}|$ is computationally demanding, especially when $k$ is relatively large. Specifically, to calculate $|\boldsymbol{\mathcal{G}}_C^{\Delta}|$, we implement the equation provided in Exercise 1.5(a) of Harary and Palmer (2014) for symmetric models (which is also provided in Entry A062734 in The On-Line Encyclopedia of Integer Sequences; OEIS Foundation Inc. 2022) and the recurrence formulae provided in Corollary 7 of Archer et al. (2020) for asymmetric models (see also Entry A057273; OEIS Foundation Inc. 2022). As these algorithms are both computationally and memory demanding for a relatively large number of areas (*e.g.*, $k > 30$), we provide pre-computed tables where each row contains $|\boldsymbol{\mathcal{G}}_C^{\Delta}|$ of each $\Delta \in \boldsymbol{\Delta}$ under a given value of $k$ (so the first row corresponds to $k = 1$, second row corresponds to $k = 2$, etc.) accompanying our implemented algorithms for computing $|\boldsymbol{\mathcal{G}}_C^{\Delta}|$ so that (by default) the desired value can be directly fetched from the tables to avoid the overhead at each computation. Currently, this implementation—accompanied by the pre-computed $|\boldsymbol{\mathcal{G}}_C^{\Delta}|$ tables—works for up to 100 areas (*cf.*, the maximum $k$ in the 749 published empirical studies is 79; input value of $k > 100$ results into an immediate error with message informing the user that the number of areas greater than 100 is currently not supported). We also provide `PARI/GP` (The PARI Group 2022) scripts in our supplementary repositories that can compute the number of strongly connected graphs with arbitrary $\Delta$ and $k$, which can then be used to expand the pre-computed table if the desired $k$ is greater than 100.

##### S2.1.3 Consequences of Computing Bayes Factor with the Unconditional Prior

The ratio of the Bayes factor computed with the incorrect unconditional prior, $P(\delta_{ij}=1)$, over the counterpart computed with the correct conditional prior, $P_C(\delta_{ij}=1)$, is:

$$\frac{\mathrm{BF}_{ij}}{\mathrm{BF}^C_{ij}} = \frac{\cancel{\frac{P(\delta_{ij}=1|G)}{1-P(\delta_{ij}=1|G)}} \div \frac{P(\delta_{ij}=1)}{1-P(\delta_{ij}=1)}}{\cancel{\frac{P(\delta_{ij}=1|G)}{1-P(\delta_{ij}=1|G)}} \div \frac{P_C(\delta_{ij}=1)}{1-P_C(\delta_{ij}=1)}} = \frac{P_C(\delta_{ij}=1)}{1-P_C(\delta_{ij}=1)} \div \frac{P(\delta_{ij}=1)}{1-P(\delta_{ij}=1)}, \tag{S15}$$

where $P(\delta_{ij}=1)$ and $P_C(\delta_{ij}=1)$ can be computed following Equations (S9) and (S14), respectively. It can be shown that, when $k \geq 4$, the prior probability that any dispersal route exists is always greater when conditioning on connectivity compared to the unconditional counterpart (*i.e.*, $P_C(\delta_{ij}=1) > P(\delta_{ij}=1)$); accordingly, the Bayes factor support for each dispersal route computed using the unconditional prior probability is always artificially inflated when there are more than three geographic areas in the study dataset (Figure S5). Unfortunately, we are unaware of any previous use of $P_C(\delta_{ij}=1)$ in calculating Bayes factors in any biogeographic study; therefore, the inferred Bayes factors reported in empirical studies are almost always inflated.

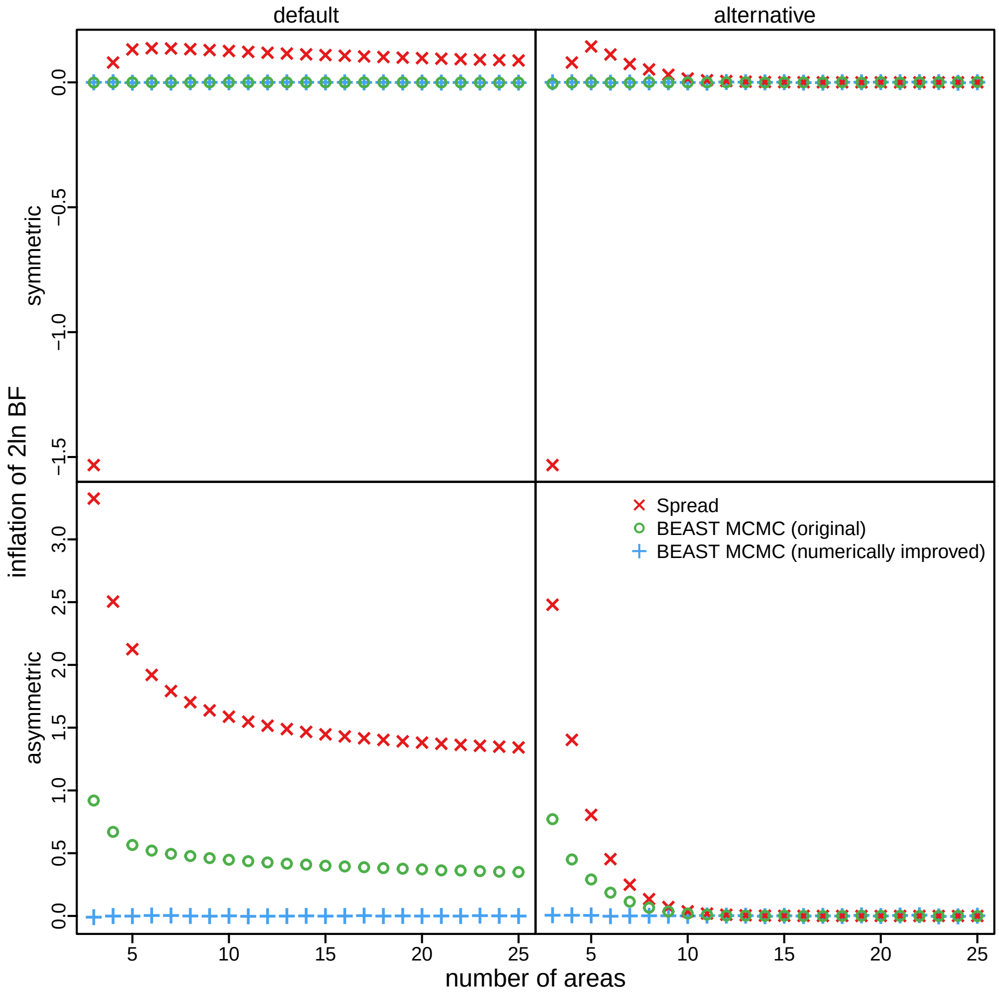


**Figure S5: The Bayes factor computed using the unconditional prior probability is inflated.** Each panel of the plot shows the difference between the computed 2 ln Bayes factor and $2\ln \mathrm{BF}^C_{ij}$ under a given geographic model (top row: symmetric; bottom row: asymmetric) and prior on $\Delta$ (left column: default; right column: alternative). The red *x*s in each panel indicate the inflation of 2 ln BF (*i.e.*, $2\ln \mathrm{BF}_{ij} - 2\ln \mathrm{BF}^C_{ij}$) under various numbers of geographic areas if the unconditional prior is used to compute the Bayes factor. The green circles reveal the inflation caused by the numerical issue in the original version of `BEAST`. The blue +s confirm that the theoretically derived conditional prior is always in accordance with the conditional behavior induced by the MCMC in the numerically improved version of `BEAST` (*i.e.*, the version that we modified to correct the numerical issue).

The inflation in the computed $2 \ln \mathrm{BF}$ (*i.e.*, $2 \ln \mathrm{BF}_{ij} - 2 \ln \mathrm{BF}^C_{ij}$) may be negligible ($< 0.1$ log unit) under the symmetric model (Figure S5, top row, red $x$s), but can be greater than 1.5 log unit under the asymmetric model (Figure S5, bottom row, red $x$s), especially under the default prior on the number of dispersal routes (Figure S5, bottom left panel, red $x$s). The inflation decreases rapidly as the number of areas $k$ increases under the alternative prior (Figure S5, bottom right panel, red $x$s), as it centers on intermediate number of dispersal routes (so that the prior mass on graphs that are not strongly connected becomes very small when $k > 10$). Accordingly, when the number of areas increases, the reported Bayes factors will become noticeably different between the default and alternative priors when the unconditional prior probability is used, even if the $2 \ln \mathrm{BF}^C_{ij}$s are identical.

To avoid confounding this artificial inflation with the real impacts of prior specification on the posterior estimates, we report the conditional Bayes factors in this study. In other words, the marked and pervasive differences in the supported dispersal routes between the default- and alternative-prior models across datasets that we have presented in this study do *not* result from a failure to condition on irreducibility when computing the Bayes factors; on the contrary, the differences would be even greater if we reported the incorrect unconditional Bayes factor.

##### S2.1.4 Numerical Issue with Computing $\mathbb{1}_C(\delta)$ in BEAST

To validate our theoretical derivation of the conditional prior probability and demonstrate that it is consistent with BEAST's implementation, we performed MCMC simulations in BEAST to approximate the prior distribution. This is achieved by specifying a discrete-geographic analysis in BEAST with the geographic data all set to "?". We performed such analyses under each value of $k$ to obtain the estimate of the conditional prior probability that each dispersal route exists, $\hat{P}_C(\delta_{ij} = 1)$, computed as the proportion of MCMC samples for which $\delta_{ij} = 1$. We then consider the ratio of $\widehat{\mathrm{BF}^C_{ij}}$ (computed with $\hat{P}_C(\delta_{ij} = 1)$) to the counterpart $\mathrm{BF}^C_{ij}$ (computed with $P_C(\delta_{ij} = 1)$) as:

$$\frac{\widehat{\mathrm{BF}^C_{ij}}}{\mathrm{BF}^C_{ij}} = \frac{P_C(\delta_{ij} = 1)}{1 - P_C(\delta_{ij} = 1)} \div \frac{\hat{P}_C(\delta_{ij} = 1)}{1 - \hat{P}_C(\delta_{ij} = 1)}. \tag{S16}$$

As expected, the difference in the computed $2 \ln \mathrm{BF}$ (*i.e.*, $2 \ln \widehat{\mathrm{BF}^C_{ij}} - 2 \ln \mathrm{BF}^C_{ij}$) was effectively zero when the model is symmetric (Figure S5, top row, green circles); to our surprise, however, the difference

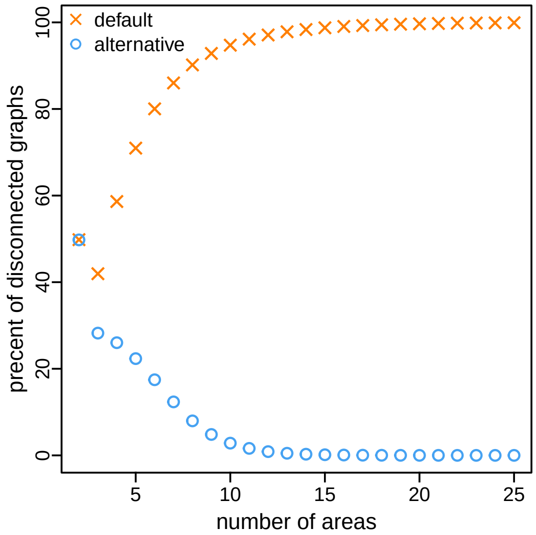


**Figure S6: BEAST by default samples graphs that are not strongly connected under prior for asymmetric models.** In theory, graphs that are not strongly connected ("disconnected") should never be sampled in the MCMC; in practice, however, due to the numerical issue with the implementation, BEAST does sample disconnected graphs under the prior (*i.e.*, without any data) when the geographic model is asymmetric. The percentage of sampled graphs that are disconnected is much higher under the default-prior model (orange $x$s) compared to the alternative-prior model (blue circles).

was always significantly greater than zero under the asymmetric model (Figure S5, bottom row, green circles). We identified a numerical issue with BEAST's implementation of the function that computes $\mathbb{1}_C(\delta)$ as the cause of this discrepancy.

Recall that BEAST determines whether the geographic model is irreducible (*i.e.*, $\mathbb{1}_C(\delta) = 1$) by computing a transition-probability matrix as $P = \exp(Qd)$ and checking if every entry of $P$, $p_{ij}$, is positive. In practice, BEAST checks whether $p_{ij} \in (\varepsilon, 1]$, where $\varepsilon$ represents a tolerance term whose default value is 1e−20. It turns out that this default value may be too small in almost all cases; when $Q$ is asymmetric, a value of $p_{ij}$ that in reality is exactly zero will very likely be on the same order as the machine precision, which is on the order of 1e−16 when double precision (likely to be the highest precision used for most BEAST analyses) is used. Therefore, some not strongly connected $\delta$ may actually be accepted as $\mathbb{1}_C(\delta)$ is incorrectly computed to be 1.

Indeed, when sampling under the prior (*i.e.*, without data), although the sampled graphs are all strongly connected under the symmetric model, many of the sampled graphs are not strongly connected when the model is asymmetric (Figure S6). The percentage of sampled graphs that are disconnected is much higher under the default-prior model (Figure S6, orange *x*s) compared to the alternative-prior model (Figure S6, blue circles), and increases as the number of areas $k$ increases; when $k > 20$, almost no graphs sampled under the default-prior model are strongly connected.

It is possible that this numerical issue can be alleviated by setting a larger $\varepsilon$ (*e.g.*, 1e−14), but as BEAST is sometimes also used on devices with lower precision (*e.g.*, single precision on GPU, where the machine precision is on the order of 1e−8) and setting an $\varepsilon$ that is too large may result in incorrectly rejecting strongly connected graphs, this solution may not be ideal. We implemented an additional function in BEAST using Kosaraju-Sharir's algorithm (Aho et al. 1983) to directly check whether the graph corresponding to $\delta$ is strongly connected.

We then reran the above analyses using this numerically improved version of BEAST to sample graphs under each prior model. As expected, the sampled graphs were *all* strongly connected (results not shown), regardless of the model symmetry, number of areas, or specified prior model. The difference in the computed $2 \ln \mathrm{BF}$ (*i.e.*, $2 \ln \widehat{\mathrm{BF}^C_{ij}} - 2 \ln \mathrm{BF}^C_{ij}$) is also always effectively zero (Figure S5, blue +s) after the numerical improvement, demonstrating that our theoretical derivation agrees with the BEAST results and the numerical issue has been resolved. We repeated all the analyses under the asymmetric model in this study using the numerically improved version of BEAST, although it appears that the impact is negligible as the results are effectively identical (results under the original version of BEAST not shown). We provide the numerically improved version of BEAST as an executable program in our our GitHub and Dryad repositories.

#### S2.2 The Prior Probability of the Geographic Area at the Root

The phylogenetic likelihood of the discrete-geographic data is computed exactly the same as the phylogenetic likelihood of nucleotide using the pruning algorithm (Felsenstein 1973, 1981), where a conditional probability vector—the probability of observing the geographic data at the descendant pathogen samples conditioning on the internal node being in each of the geographic areas—is computed for each node traversing down the tree until reaching the root of the tree. At the root, the conditional probability vector needs to be weighted by the (prior) probability that the pathogen originated in each of the geographic areas to obtain the unconditional likelihood of the geographic data. Let $\boldsymbol{\omega}$ denote this prior probability ("root-frequency") vector for the geographic model.

Following the convention for modeling the substitution process, Lemey et al. (2009) assign $\boldsymbol{\omega} = \boldsymbol{\pi}$ when the geographic model is symmetric, where $\boldsymbol{\pi}$ denotes the stationary distribution of the geographic process, so the prior probability of the geographic area at the root is uniform across all areas (*i.e.*, each $\omega_i$ is fixed as $\frac{1}{k}$). Conversely, when the geographic model is asymmetric, `BEAST` by default specifies $\boldsymbol{\omega}$ as an additional vector of free parameters to be estimated from the data; the default prior on $\boldsymbol{\omega}$ is a flat Dirichlet distribution, $\mathrm{Dir}(1, 1, \ldots, 1)$. This is similar with how the root-frequency vector is treated when nonstationary substitution models are used (*e.g.*, Yang and Roberts 1995; Galtier and Gouy 1998; Blanquart and Lartillot 2006), although the geographic model—even if asymmetric—is still stationary given its irreducibility (see Section S2.1).

We follow this default-model specification in our study to highlight the empirical consequences of the priors on the average dispersal rate and the number of dispersal routes, as presented in the main text. It appears that the impacts of alternative ways of specifying the root-frequency vector on the parameter estimates are relatively small in our preliminary analyses (results not shown).

#### S2.3 Joint and Sequential Bayesian Phylodynamic Inference

For simplicity, our description of the biogeographic model in the main text assumes that the phylogeny, $\Psi$, is known without error. However, empirical applications typically embed this biogeographic model in a larger "phylodynamic" model, which jointly models multiple aspects of epidemiological evolution, namely: (1) the *diversification model*, with parameters $\theta_\Psi$ that describe the branching process that generates the phylogeny; (2) the *substitution model*, with parameters $\theta_S$ that describe the process of molecular evolution over the branches of the tree, and; (3) the *geographic model*, with parameters $\theta_G = \{\boldsymbol{r}, \boldsymbol{\delta}, \mu\}$ that describe the dispersal of pathogens among areas. The resulting joint posterior density for the full phylodynamic model can be written as:

$$P(\Psi, \theta_\Psi, \theta_S, \theta_G \mid X, G) = \frac{P(X, G \mid \Psi, \theta_\Psi, \theta_S, \theta_G) P(\Psi, \theta_\Psi, \theta_S, \theta_G)}{P(X, G)}, \tag{S17}$$

where $X$ is an alignment of molecular sequence data and $G$ is the geographic data. Conditional on the phylogeny, the processes of molecular and geographic evolution are assumed to be independent. Combined with an assumption that the parameters of the three model components are independent *a priori*, Equation (S17) can be written as:

$$P(\Psi, \theta_\Psi, \theta_S, \theta_G \mid X, G) = \frac{P(X \mid \Psi, \theta_S) P(G \mid \Psi, \theta_G) P(\Psi \mid \theta_\Psi) P(\theta_\Psi) P(\theta_S) P(\theta_G)}{P(X, G)}. \tag{S18}$$

In principle, this joint posterior density can be approximated using Markov chain Monte Carlo (MCMC). However, owing to the complexity of the joint model, these MCMC analyses may perform poorly in practice. To simplify the MCMC, it is possible to perform a "sequential" analysis consisting of two steps that together are equivalent to a joint analysis. The first step estimates the joint posterior density of phylogenies, diversification-model parameters, and substitution-model parameters:

$$P(\Psi, \theta_\Psi, \theta_S \mid X) = \frac{P(X \mid \Psi, \theta_S) P(\Psi \mid \theta_\Psi) P(\theta_\Psi) P(\theta_S)}{P(X)}; \tag{S19}$$

this joint posterior density is approximated using MCMC. The second step uses the marginal posterior density of phylogenies from the first step as a prior to estimate the joint posterior distribution of the geographic-model parameters:

$$P(\Psi, \theta_G \mid G) = \frac{P(G \mid \Psi, \theta_G) P(\Psi) P(\theta_G)}{P(G)}, \tag{S20}$$

where $P(\Psi)$ corresponds to the marginal posterior distribution of phylogenies from the first step, $P(\Psi \mid X)$. Again, this joint posterior density is estimated using MCMC. Proposals for the phylogeny are made by drawing a new phylogeny, $\Psi'$, from the marginal prior distribution, $P(\Psi \mid X)$, and accepting the proposal with probability:

$$A = \min \left[ 1, \frac{P(G \mid \Psi', \theta_G) P(\Psi')}{P(G \mid \Psi, \theta_G) P(\Psi)} \times \frac{P(\Psi \mid X)}{P(\Psi' \mid X)} \right]. \tag{S21}$$

Recognizing that each sample from the joint posterior distribution in second step is associated with a sample of $\Psi$ from the first step, we can reconstitute the full joint posterior distribution (Equation (S17)). That is, for the $i^{\text{th}}$ sample from the posterior distribution of the second step with phylogeny $\Psi^i$, we can find the sample of the first step associated with $\Psi^i$, and "attach" the corresponding sample of parameters from the first step to the $i^{\text{th}}$ sample of the second step. The resulting distribution of samples is theoretically equivalent to the joint posterior distribution of the full model (*i.e.*, the sequential analysis is theoretically equivalent to the joint analysis).

We note that `BEAST` provides two options for performing sequential analysis through `empiricalTreeDistributionModel`. The first option, evoked with the argument `MetropolisHastings = "true"` of the `empiricalTreeDistributionOperator`, uses the proposal mechanism described above and uses Equation (S21) to accept or reject proposals on the tree. The second option, evoked with the argument `MetropolisHastings = "false"`, proposes new trees by drawing them from the marginal prior density, and then accepts the proposed tree with probability 1. The second option does not result in an ergodic Markov chain with a stationary distribution equivalent to Equation (S20), and therefore is not equivalent to a full joint phylodynamic analysis (Equation (S17)). For this reason, we use the argument `MetropolisHastings = "true"` for all empirical analyses described below.

### S3 Analyses of Empirical Datasets

#### S3.1 General Analysis Protocol

To explore the empirical consequences arising from our theoretical concerns with the informative default priors, we collected 14 datasets from published empirical studies (see Table S5), and reanalyzed each dataset using the sequential approach under a suite of biogeographic models, including all combinations of: (1) a symmetric and asymmetric rate matrix; (2) default and alternative priors on the number of dispersal routes; and (3) default and alternative priors on the average dispersal rate. We provide details of these two prior models in the Theoretical Concerns section of the main text and in Table S1.

**Table S5: Empirical datasets information.**

| Study | Virus | Dataset | $N$ | $k$ |
|---|---|---|---|---|
| Dash et al. (2015) | Dengue | — | 62 | 23 |
| Wilfert et al. (2016) | DWV | lp | 209 | 8 |
| | | rdrp | 183 | 7 |
| | | vp3 | 96 | 7 |
| Faria et al. (2014) | HIV | A | 792 | 8 |
| | | B | 927 | 10 |
| | | C | 466 | 8 |
| Bedford et al. (2015) | Influenza | H3 | 1391 | 9 |
| | | Yam | 1240 | 9 |
| Yao et al. (2015) | Rabies | — | 141 | 18 |
| Gao et al. (2022) | SARS-CoV-2 | Global | 1271 | 23 |
| Alpert et al. (2021) | SARS-CoV-2 | B.1.1.7 US | 1908 | 22 |
| Candido et al. (2020) | SARS-CoV-2 | Brazil SchemeB | 1182 | 10 |
| | | Brazil SchemeC | 1182 | 22 |

*Estimating the Marginal Posterior Distribution of Phylogenies from Molecular Sequence Data, $P(\Psi \mid X)$*

The marginal posterior distribution of trees inferred in three of the empirical studies (Faria et al. 2014; Bedford et al. 2015; Candido et al. 2020) were available directly; we used these posterior distributions of trees as the corresponding prior distributions for the second step of our sequential phylodynamic analyses. For the SARS-CoV-2 Global dataset (Gao et al. 2022), we conditioned the subsequent phylodynamic analyses on the maximum clade credibility (MCC) tree summarized from the marginal posterior distribution to ensure numerical stability of the analyses. We also conditioned on the MCC tree for the SARS-CoV-2 B.1.1.7 US dataset (Alpert et al. 2021) as original empirical study conditioned on that tree (instead of averaging over the marginal posterior distribution of trees).

The posterior distributions of trees were not published for the remainder of the empirical studies; accordingly, in these cases we first inferred the posterior distributions of trees from the corresponding sequence data, and then used the resulting posterior distributions of trees as the prior distributions in the second step of our sequential phylodynamic analyses. For each of these latter studies, we obtained the nucleotide sequences and sampling-time information from the original studies. When only the raw sequence data were available for a given study, we inferred the sequence alignment using `MUSCLE` version 3.8 (Edgar 2004). For each of the five (published or inferred) sequence alignments, we inferred the posterior probability density of trees under the identical diversification and substitution models as those used in the original studies, and then performed MCMC simulations to approximate the joint posterior distribution using `BEAST` version 1.8.2 (Drummond et al. 2012) (with `BEAGLE` version 3.1.2 [Ayres et al.

2019] enabled). Details of these analyses are available in the XML scripts included in our GitHub and Dryad repositories. For each dataset, we performed four replicate MCMC simulations; we set the length (100–200 million generations) and sampling frequency of each simulation to values that provided an adequate approximation of the posterior distribution of model parameters. We combined the posterior samples of trees from the four replicate simulations (after discarding burnin samples from each simulation) using `LogCombiner` version 1.8.2. We then subsampled the resulting composite posterior sample of trees to retain a total of 500–1000 trees (available in our GitHub and Dryad repositories); we used this posterior sample of trees as the prior distribution for the second step of the corresponding sequential analyses (detailed below).

*Estimating the Joint Posterior Distribution of Geographic-Model Parameters,* $P(\boldsymbol{r}, \boldsymbol{\delta}, \mu, \Psi \mid G)$

For each empirical dataset, we performed MCMC simulations to infer the joint posterior probability distribution under the symmetric geographic models using `BEAST` version 1.8.2 (Drummond et al. 2012), with `BEAGLE` version 3.1.2 (Ayres et al. 2012, 2019) enabled (except the SARS-CoV-2 datasets, for which we used `BEAST` version 1.10.5 (Suchard et al. 2018) with `BEAGLE` version 3.2.0). For the analyses under the asymmetric geographic model, we used our modified version of `BEAST` (with `BEAGLE` version 3.2.0) to avoid the numerical issue with determining the irreducibility of a geographic model (see Section S2.1.4). For each candidate model, we performed 4–16 replicate MCMC simulations; we set the length (5–50 million generations) and sampling frequency of each MCMC simulation to values that provided an adequate approximation of the posterior distribution of the geographic-model parameters. Details of these analyses are available in the XML scripts included in our GitHub and Dryad repositories. In these repositories, we also provide `R` scripts that can be used to generate the XML scripts. We then discarded burnin samples drawn from the first 5–20% of each MCMC simulation, and then combined the remaining samples from the four replicate MCMC simulations using `LogCombiner` version 1.8.2. Finally, we generated the MCC tree from the composite posterior sample for each unique analysis using `TreeAnnotator` version 1.8.2.

*Estimating the Joint Prior distribution of Geographic model parameters,* $P(\boldsymbol{r}, \boldsymbol{\delta}, \mu, \Psi)$

For each empirical dataset, we also run MCMC simulations in `BEAST` (Drummond et al. 2012; Suchard et al. 2018) to approximate the joint prior distribution under each prior model. This is achieved by setting all the geographic data to "?" in the XML scripts so that the data contain no information at all. Details of these analyses are available in the XML scripts included in our GitHub and Dryad repositories. We run 2–16 independent MCMC replicates (2.5–20 million generations each) under each model, and then we combined the samples of parameter estimates from each independent replicate with the first 5–20% discarded as the burn-in using `LogCombiner` version 1.8.2.

*Estimating the Posterior Distribution of Biogeographic History*

We estimated the ancestral area at each internal node using the ancestral-state estimation algorithm (Yang 2014) implemented in `BEAST`. We calculated the expected number of dispersal events between each pair of areas using the "fast stochastic-mapping algorithm" developed by Minin and Suchard (2008a, see also Minin and Suchard 2008b, O'Brien et al. 2009) implemented in `BEAST`. The exceptions are SARS-CoV-2 datasets, where we inferred the number of dispersal events between each pair of areas by simulating the full biogeographic history using the stochastic-mapping algorithm Nielsen (2002); Rodrigue et al. (2007); Hobolth and Stone (2009) implemented in `BEAST`. These two statistics were computed during the MCMC simulation used to infer the joint posterior distribution of geographic-model parameters, $P(\boldsymbol{r}, \boldsymbol{\delta}, \mu, \Psi \mid G)$ (*i.e.*, in the second step of our sequential analyses).

*Estimating Marginal Likelihoods, $P(G)$*

We used Bayes factors to evaluate the relative fit of each candidate prior model to each of the biogeographic datasets; to this end, we estimated the marginal likelihood for each prior model using both thermodynamic integration (Lartillot and Philippe 2006) and stepping-stone sampling (Xie et al. 2011; Baele et al. 2012). We ran four independent series of power-posterior simulations to estimate the marginal likelihood of each prior model. We set the chain length and sampling frequency of the power-posterior analysis at each stone, as well as the number of stones, to achieve stable marginal-likelihood estimates (both among the four replicates and also between thermodynamic integration and stepping-stone sampling estimators). Details of these analyses are available in the XML scripts included in our GitHub and Dryad repositories.

*Posterior-Predictive Simulations*

We used posterior-predictive simulation to evaluate the absolute fit of each candidate prior model to each of the biogeographic datasets (Gelman et al. 1996; Bollback 2002). For each model and dataset combination, we combined the four MCMC replicates and simulated $m = 2500$ predictive datasets. For each predictive dataset, $G_i^{\text{sim}}$, we drew a vector of parameters, $\theta_i = \{\Psi_i, \boldsymbol{r_i}, \boldsymbol{\delta_i}, \mu_i\}$, at random from the combined MCMC samples, and simulated a dataset conditional on those parameters using the `sim.history()` function in the `R` package `phytools` (Revell 2012). We then calculated a difference statistic for the $i^{\text{th}}$ simulated dataset as:

$$D_i = T(G_i^{\text{sim}} \mid \theta_i) - T(G^{\text{obs}} \mid \theta_i),$$

where $G^{\text{obs}}$ is the observed biogeographic dataset, and $T(\cdot \mid \theta_i)$ is a summary statistic (detailed below). For the $m$ predictive datasets for a given model and dataset combination, we calculated the posterior-predictive $p$-value as:

$$P = \left[\frac{1}{m}\sum_{i=1}^{m} D_i > 0\right] + \left[\frac{1}{2}\frac{1}{m}\sum_{i=1}^{m} D_i = 0\right],$$

where the first term measures the fraction of simulated statistics that are more extreme than the observed statistic, and the second term measures *half* the fraction of simulated statistics that are equal to the observed statistic (to accommodate discrete summary statistics, as described by Gelman et al. 2013). Posterior-predictive $p$-values between 0.025 and 0.975 indicating that the model is adequate and cannot be rejected (*i.e.*, the observed statistic is within the 95% posterior-predictive interval).

We used two summary statistics to assess model adequacy: (1) the *parsimony statistic*, and; (2) the *tip-wise multinomial statistic*. We calculated the posterior-predictive $p$-value for both of these statistics for each model and dataset combination. For the parsimony statistic, we simply calculated the parsimony score for the given simulated or observed dataset, conditional on the sampled tree, $\Psi_i$, using the `parsimony()` function in `R` package `phangorn` (Schliep 2010). The tip-wise multinomial statistic is similar to the multinomial statistic introduced by Goldman (1993) and used in posterior-predictive simulation by Bollback (2002), which treats the sites (columns) in a molecular alignment as outcomes of a multinomial trial. Our tip-wise statistic is similar, but treats the states at the tips of the tree for the single geographic character (*i.e.*, site) as the outcomes of the multinomial trial. For the tip-wise multinomial statistic, we calculated:

$$T(G \mid \theta_i) = \sum_{i=1}^{k} n_i \ln(n_i/n),$$

where $n$ is the number of tips, and $n_i$ is the number of tips in state $i$. (Note that this statistic is also similar to the entropy statistic used to assess genetic variability along sequences; Shannon 1948; Schneider et al. 1986). Details about the computation of these two summary statistics are available in the `R` script included in our GitHub and Dryad repositories.

*Data Cloning*

We explored the use of a computational technique called *data cloning* to understand the sensitivity of posterior estimates to the choice of prior. Originally developed as a tool for using MCMC to perform maximum-likelihood inference (Robert 1993), and later used as a tool for understanding model identifiability for complex Bayesian models (Lele et al. 2007; Ponciano et al. 2009, 2012), data cloning involves performing a sequence of MCMC analyses with an increasing number of duplicates of the observed data. A particular MCMC in the sequence is defined by the number of duplicated datasets, $\beta_i \geq 1$, with the resulting posterior distribution being:

$$P(\theta \mid X)_{\beta_i} \propto P(X \mid \theta)^{\beta_i} P(\theta).$$

As $\beta_i \to \infty$ (assuming the model is identifiable), the joint posterior distribution converges to a point that corresponds to the joint maximum-likelihood estimate (MLE); if the joint posterior distribution does not converge to a point, then the model is non-identifiable (*i.e.*, the MLE may not be unique). When the model is identifiable, the rate at which the joint posterior distribution converges to the MLE is proportional to the amount of information available in the data relative to the strength of the prior, *i.e.*, when the prior is extremely (mis)informative, convergence to the MLE will be very slow.

We used data cloning to analyze each of the biogeographic datasets under the symmetric default- and alternative-prior models, with $\beta = \{1, 5, 10, 20\}$. This is achieved by duplicating the discrete-geography data in the `BEAST` XML scripts. These XML scripts are available in our GitHub and Dryad repositories. In these repositories, we also provide `R` scripts that can be used to generate the XML scripts.

To ensure good MCMC performance, we conducted the biogeographic inferences conditioning on the MCC tree inferred using the sequence data. In all cases, the inferred posterior distributions shrink as $\beta$ increases. Under the alternative-prior model, the posterior-mean estimates remain mostly constant as $\beta$ increases (*i.e.*, the posterior-mean estimates are almost identical to the MLEs). By contrast, the posterior-mean estimates under the default-prior model change drastically as $\beta$ increases, converging to the MLEs very slowly (Figures S15 and S16). These results indicate that the default-prior models exert much stronger influence on posterior estimates relative to the alternative-prior models.

*MCMC Diagnosis*

After initial inspection of the output log files using `Tracer` version 1.7.1 (Rambaut et al. 2018), we assessed MCMC performance using the `coda` package (Plummer et al. 2006) in `R` (R Core Team 2020). Specifically, we assess mixing and adequacy within each MCMC replicate by calculating the effective sample size (ESS) diagnostic for each continuous parameter (ensuring ESS values $\gg 100$) after discarding the first 5–20% of samples from each replicate simulation as the burn-in. We assessed convergence among replicate MCMC simulations by calculating the potential scale reduction factor (PSRF Gelman and Rubin 1992) diagnostic for each continuous parameter (ensuring $\text{R} \approx 1$). We also assessed the convergence among replicates by calculating the ESS for each continuous parameter for each combined MCMC chain (independent replicates combined after discarding the burn-in), ensuring the ESS values $\gg 200$.

*Parameter Summaries*

As described in the Empirical Consequences section of the main text, for each dataset we summarized the following statistics: (1) marginal likelihood; (2) posterior-predictive summary statistics; (3) rate of dispersal between each pair of areas; (4) support for dispersal routes between each pair of areas; (5) average dispersal rate among all areas; (6) ancestral area at each internal node of the phylogeny; (7) total number of dispersal events among all areas, and; (8) number of dispersal events between each pair of areas. The `R` scripts used to summarize these statistics are available in our GitHub and Dryad repositories. We compared estimates of these statistics across all candidate models to assess the impact of the default and alternative priors. We report the meta summaries of these statistics across all the empirical datasets in Section S3.2, and provide these summaries for each dataset in detail in Section S3.3.

*Data and Code Availability*

The sequence, sampling time and geography data used in this study, as well as the phylogenies we marginalized over or conditioned on in the biogeographic inference, are maintained in the GitHub repository (https://github.com/jsigao/prior_misspecification_phylodynamic_biogeography) and archived in the Dryad repository (https://datadryad.org/stash/share/7Rd5kdTh7V66w9XefTuSeoui0LLv6LAwcY_5buMwUZU). Our repositories also contain BEAST XML scripts used to perform the phylodynamic analyses and R scripts used to post process the analyses and perform posterior-predictive simulation.

##### S3.2 Expanded Meta Summaries of Empirical Analyses

In this section, we provide various summaries across all the empirical datasets. In the main text, we focus on comparisons between the preferred default-prior model and the preferred alternative-prior model; here, we provide results for pairwise comparisons between all the eight candidate prior models.

*The Impact of Prior Choice on Pairwise Dispersal Rates*

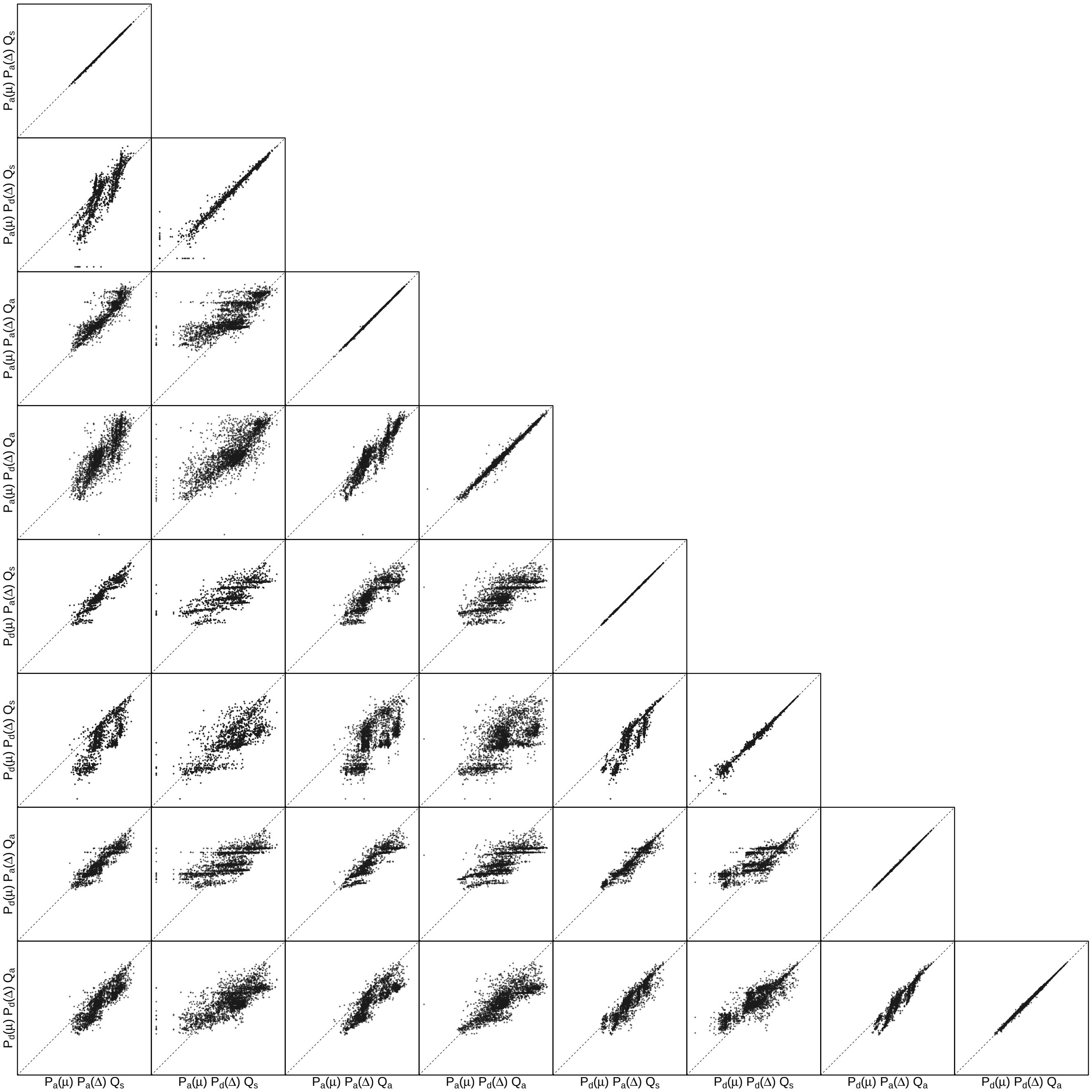


**Figure S7: The impact of prior choice on pairwise dispersal rates.** Each cell of the plot compares posterior-mean estimate of the rate of dispersal between each pair of geographic areas, $q_{ij}$, between each pair of prior models, summarized across all datasets. $x$- and $y$-axis of each cell are both on log scale. Diagonal cells are comparisons between half and the other half of the replicates under the same prior model, assessing the convergence of MCMC simulations; off-diagonal cells are comparisons between different prior models, demonstrating the impact of the prior model on the estimates of pairwise dispersal rates. Axis label notation for the prior models follows Table S2.

*The Impact of Prior Choice on Average Dispersal Rate*

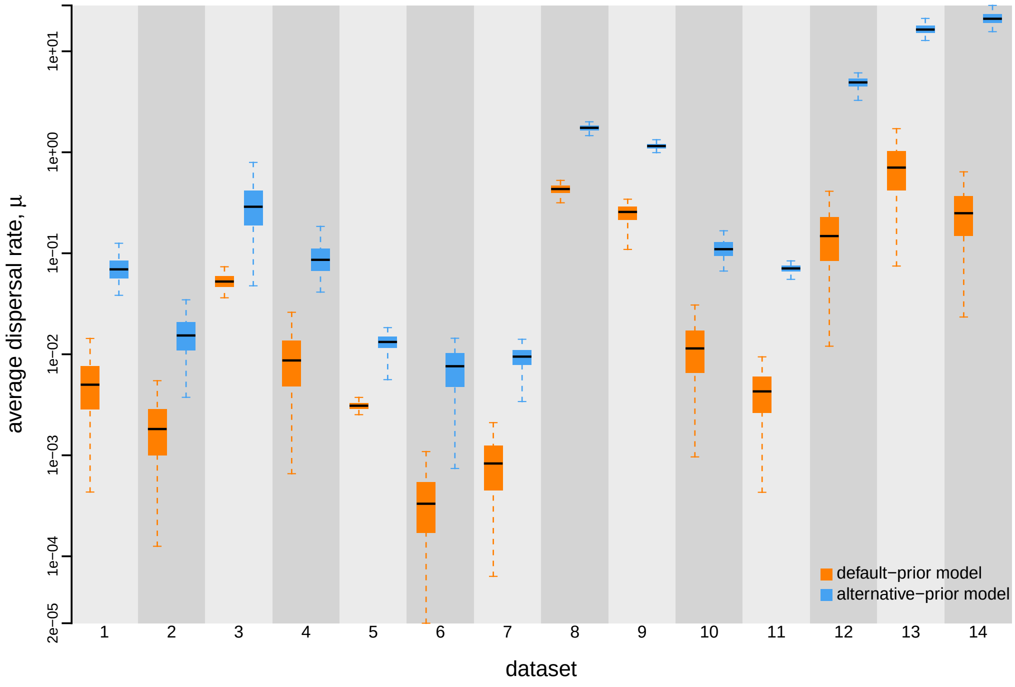


**Figure S8: The impact of prior choice on the average dispersal rate.** Each column depicts estimates for one empirical dataset (see Table S5 for the description of datasets). Within each column, each pair of boxplots depicts posterior estimates of the average dispersal rate, $\mu$, under the default (orange) and alternative (blue) prior models: the center of each box indicates the posterior-median rate; the box and whiskers indicate the corresponding 50% and 95% credible intervals, respectively.

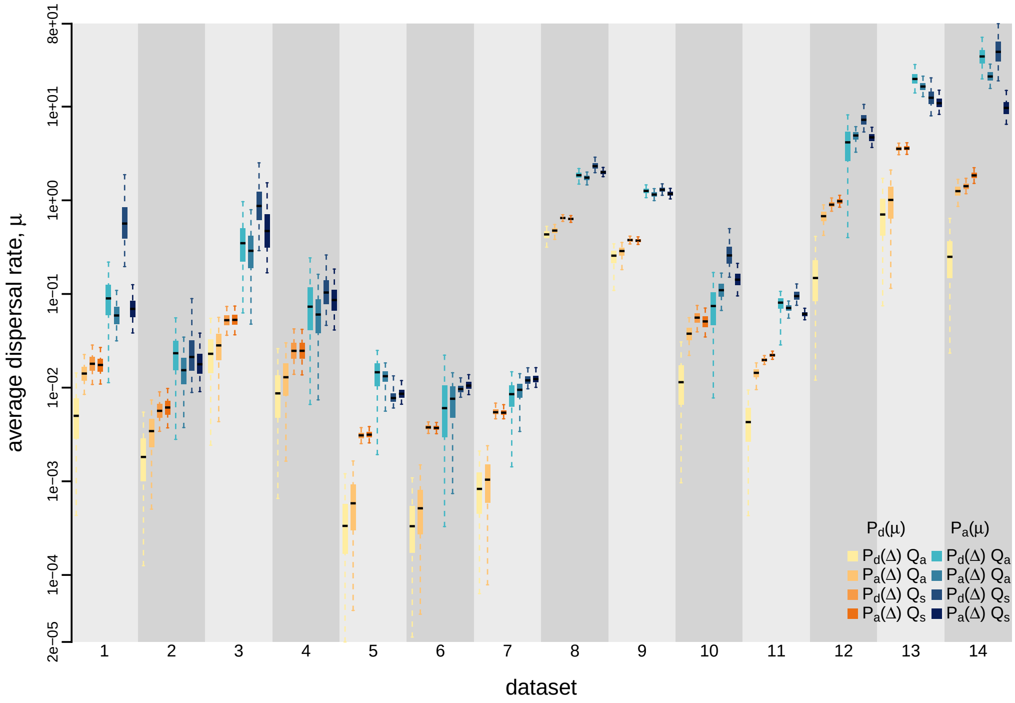


**Figure S9: The impact of prior choice on the average dispersal rate.** Each column depicts estimates for one of the 14 datasets (description of datasets see Table S5). Within each column, the set of eight boxplots depicts posterior estimates of the average dispersal rate, $\mu$, under the prior models: the center of each box indicates the posterior-median rate; the box and whiskers indicate the corresponding 50% and 95% credible intervals.

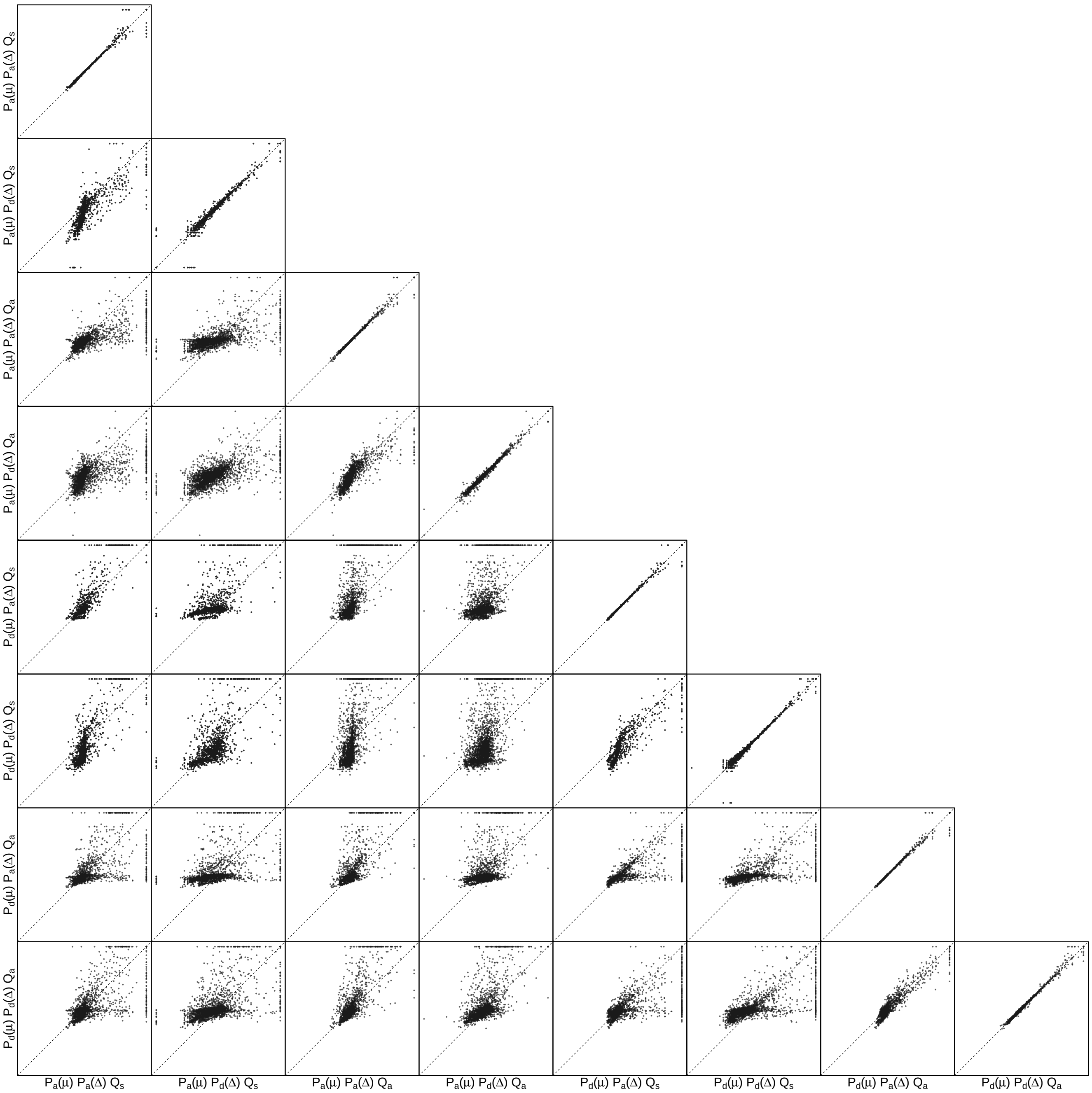


**Figure S10: The impact of prior choice on the inferred support for dispersal routes.** Each cell of the plot compares the inferred support ($2 \ln \mathrm{BF}$) for pairwise dispersal routes between each pair of prior models, summarized across all datasets. Diagonal cells are comparisons between half and the other half of the replicates under the same prior model, assessing the convergence of MCMC simulations; off-diagonal cells are comparisons between different prior models, demonstrating the impact of the prior model on the inferred support for pairwise dispersal routes. Axis label notation for the prior models follows Table S2.

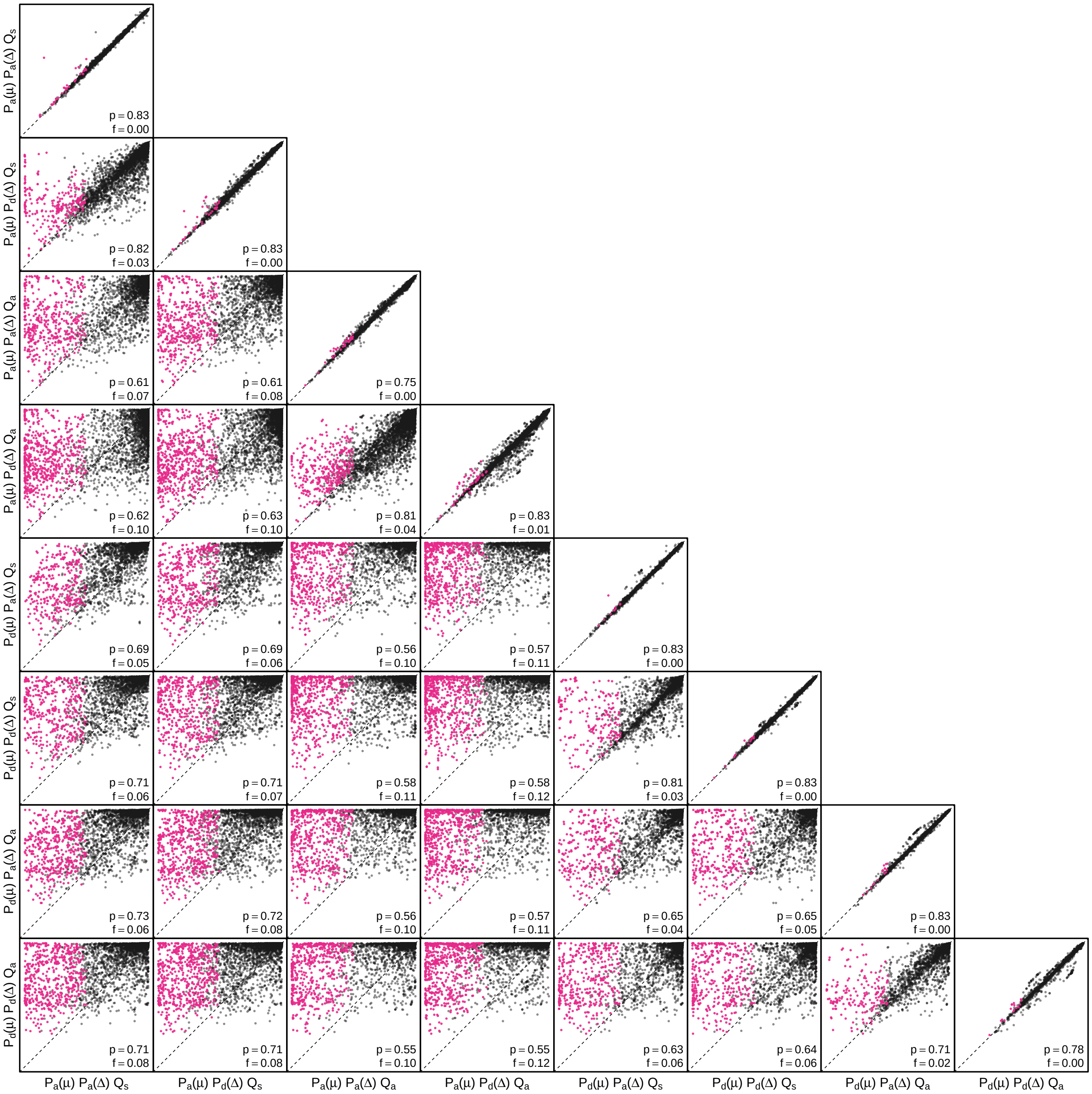


**Figure S11: The impact of prior choice on ancestral-area estimates.** Each cell of the plot compares the estimated posterior probability of the maximum a posteriori (MAP) ancestral area between each pair of prior models, summarized across all datasets. Diagonal cells are comparisons between two replicates under the same prior model, assessing the convergence of MCMC simulations; off-diagonal cells are comparisons between different prior models, demonstrating the impact of the prior model on both the posterior probability and the identity of the MAP ancestral-area estimates at internal nodes. Pink dots represent the internal nodes where the MAP ancestral area inferred under the default-prior model differs from that inferred under the alternative-prior models. The statistic $p$ denotes the fraction of internal nodes that are shared under the default- and alternative-prior models; $f$ is the fraction of shared nodes where the MAP ancestral area differs under the default- and alternative-prior models. Axis label notation for the prior models follows Table S2.

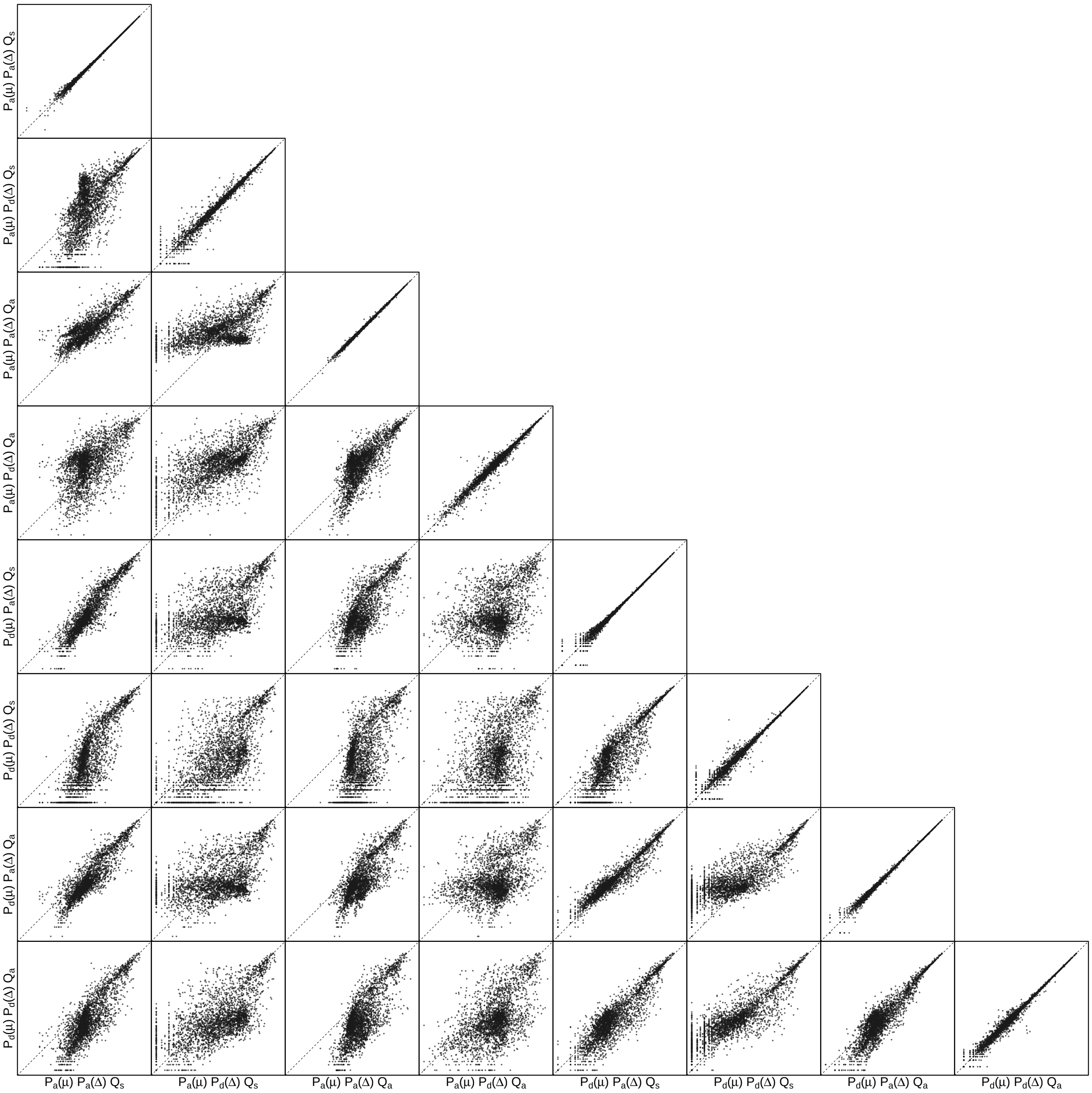


**Figure S12: The impact of prior choice on the inferred number of dispersal events between each pair of areas.** Each cell of the plot compares the inferred number of pairwise dispersal events between each pair of prior models, summarized across all datasets. $x$- and $y$-axis of each cell are both on log scale. Diagonal cells are comparisons between half and the other half of the replicates under the same prior model, assessing the convergence of MCMC simulations; off-diagonal cells are comparisons between different prior models, demonstrating the impact of the prior model on the inferred number of pairwise dispersal events. Model notation on the axis follows Table S2.

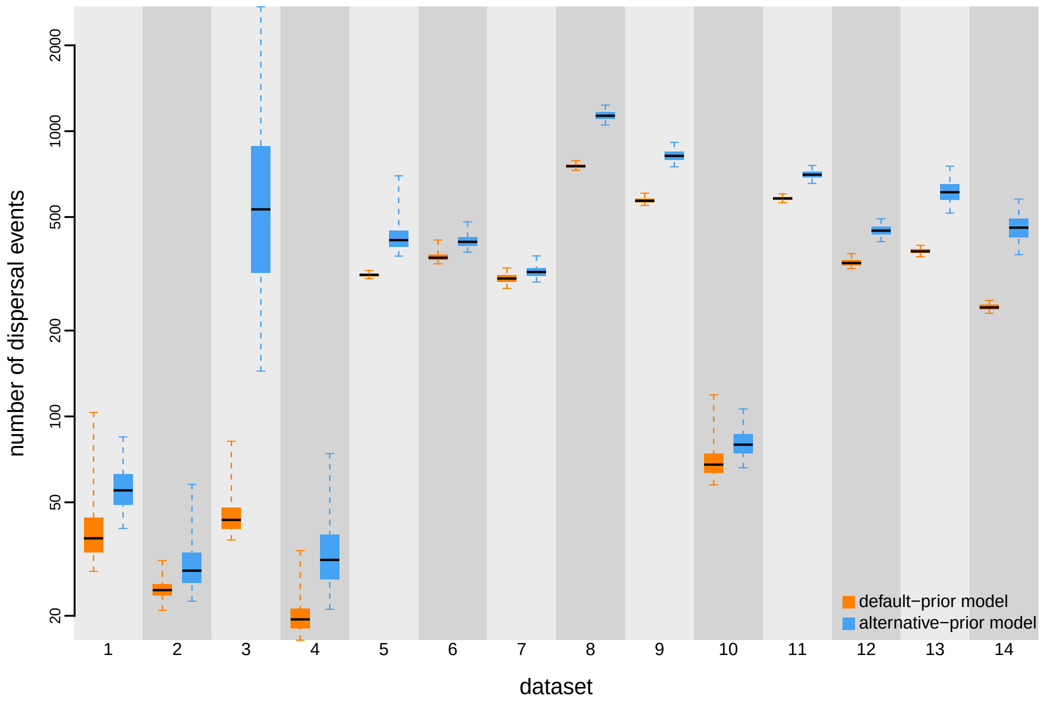


**Figure S13: The impact of prior choice on the inferred total number of dispersal events between all areas.** Each column depicts estimates for one of the 14 datasets (description of datasets see Table S5). Within each column, the pair of boxplots depicts posterior estimates of the total number of dispersal events under the default (orange) and alternative (blue) prior models: the center of each box indicates the posterior-median number of dispersal events; the box and whiskers indicate the corresponding 50% and 95% credible intervals, respectively.

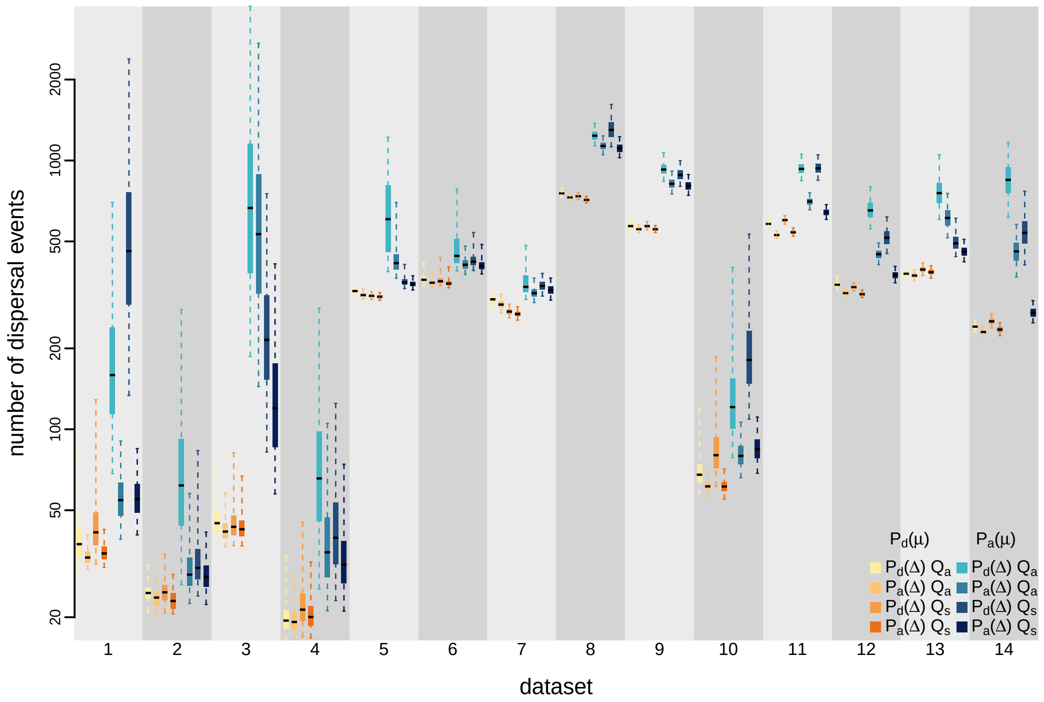


**Figure S14: The impact of prior choice on the inferred total number of dispersal events between all areas.** Each column depicts estimates for one of the 14 datasets (description of datasets see Table S5). Within each column, the set of eight boxplots depicts posterior estimates of the total number of dispersal events under each of the prior models: the center of each box indicates the posterior-median number of dispersal events; the box and whiskers indicate the corresponding 50% and 95% credible intervals, respectively.

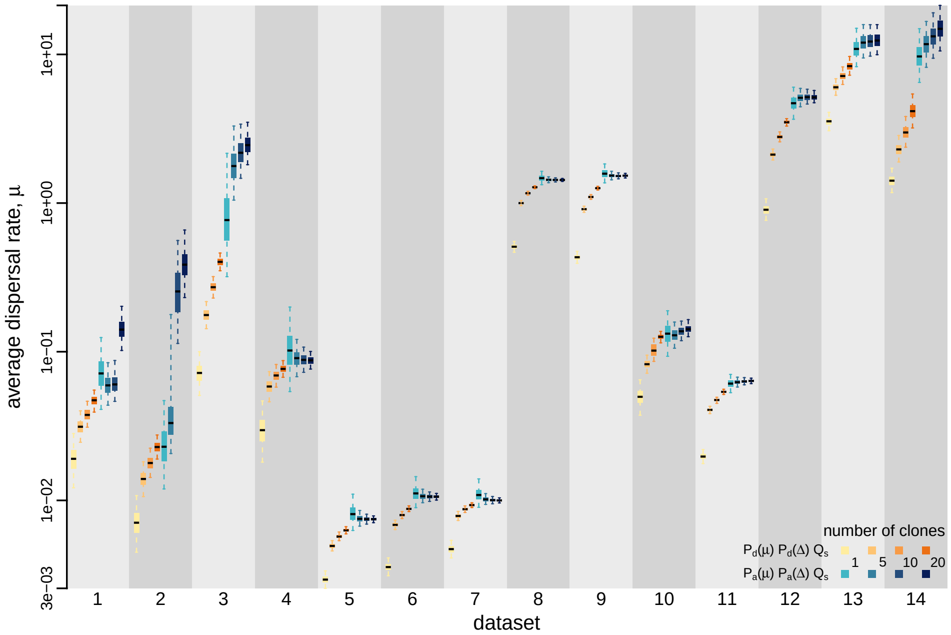


**Figure S15: Using data cloning to explore the impact of prior choice on posterior estimates of the average dispersal rate.** Each column depicts estimates for one of the 14 datasets (description of datasets see Table S5). Within each column, the set of eight boxplots depicts posterior estimates inferred from the associated dataset that has been cloned 1, 5, 10, 20 times: the center of each box indicates the posterior-median average dispersal rate; the box and whiskers indicate the corresponding 50% and 95% credible intervals, respectively.

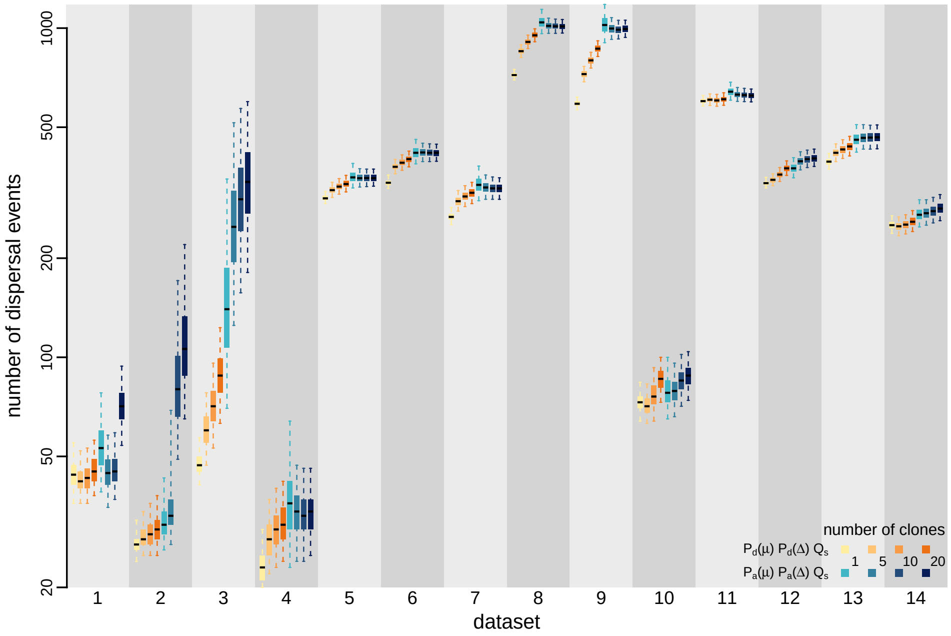


**Figure S16: Exploring the impact of prior choice on posterior estimates of the number of dispersal events via data cloning.** Each column depicts estimates for one of the 14 datasets (description of datasets see Table S5). Within each column, the set of eight boxplots depicts posterior estimates inferred from the associated dataset that has been cloned 1, 5, 10, 20 times: the center of each box indicates the posterior-median number of dispersal events; the box and whiskers indicate the corresponding 50% and 95% credible intervals, respectively.

##### S3.3 Expanded Dataset-Specific Summaries of Empirical Analyses

In this section, we provide dataset-specific summaries of the statistics we reported in the section above. These results reveal a highly consistent pattern across all the datasets, further demonstrating the widespread impact of prior misspecification in biogeographic inferences and the universality of this issue.

###### S3.3.1 Dengue Virus

Dash et al. (2015) studied the geographic dynamics of Dengue virus type 1 (DENV-1) in India and inferred the history by which this virus dispersed throughout the world. This study contains a single dataset comprised of sequences of (part of) the envelope gene, sampled from across a large number of distant geographic areas over a protracted sampling interval (1956–2011). We acquired the sampling time and location data, as well as the GenBank accession numbers from the sequence names in the MCC tree figured in Dash et al. (2015, Fig. 3), and then obtained the nucleotide sequences from GenBank. This dataset has 62 sequences distributed among 23 defined geographic areas. We aligned the nucleotide sequences using `MUSCLE` version 3.8 (Edgar 2004). The files containing the GenBank accession numbers, the sequence alignment, and the sampling time and location data are available in our GitHub and Dryad repositories.

To infer the marginal posterior distribution of phylogenies given the sequence alignment, we specified a phylogenetic model with the following components: (1) the GTR+I+$\Gamma_4$ substitution model (Tavaré 1986; Yang 1994; Gu et al. 1995); (2) the uncorrelated lognormal (UCLN) branch-rate prior model (Drummond et al. 2006; Rannala and Yang 2007), and; (3) the Gaussian Markov Random Field (GMRF) Bayesian Skyride coalescent node-age model (Minin et al. 2008). Details of these analyses are available in the XML scripts included in our GitHub and Dryad repositories.

We ran four independent MCMC simulations in `BEAST` version 1.8.2 for 200 million generations each, sampling every 15000 generations. We first assessed the performance of each MCMC simulation using `Tracer` version 1.7.1 (Rambaut et al. 2018), removed the first 10% of samples from each chain as the burn-in, and then combined the remaining posterior samples of trees from the replicate simulations using `LogCombiner` version 1.8.2. This resulted in a posterior sample of 1200 trees (available in our GitHub and Dryad repositories), which we then used as the prior distribution of phylogenies for the second step of our sequential analyses.

The MCMC simulations used in the second step of our sequential analyses and of the analyses used to estimate marginal likelihoods under each prior model are described above in Section S3.1. Details of these analyses are available in the XML scripts included in our GitHub and Dryad repositories.

**Table S6: Marginal-likelihood estimates of the eight prior models for the Dengue virus dataset.** Columns 2–5 list marginal likelihoods inferred from four replicate analyses. Columns 6–7 list the mean and standard deviation of these marginal-likelihood estimates. The last column lists the marginal-likelihood estimates computed by combining the samples from the replicate power-posterior MCMC simulations. Candidate models are listed in rows and include all possible combinations of: (1) instantaneous-rate matrices (symmetric, $Q_s$ or asymmetric, $Q_a$); (2) priors on the average dispersal rate [default, $P_d(\mu)$ or alternative, $P_a(\mu)$], and; (3) priors on the number of dispersal routes [default, $P_d(\Delta)$ or alternative, $P_a(\Delta)$]. The preferred default- and alternative-prior models are indicated in bold text.

| Model | replicate1 | replicate2 | replicate3 | replicate4 | mean | sd | combined |
|---|---|---|---|---|---|---|---|
| $\boldsymbol{P_d(\mu) Q_a P_d(\Delta)}$ | **-187.73** | **-187.55** | **-187.55** | **-187.78** | **-187.65** | **0.12** | **-187.65** |
| $P_d(\mu) Q_a P_a(\Delta)$ | -170.68 | -170.55 | -170.47 | -170.77 | -170.62 | 0.13 | -170.61 |
| $P_d(\mu) Q_s P_d(\Delta)$ | -190.44 | -190.62 | -190.47 | -190.39 | -190.48 | 0.10 | -190.46 |
| $P_d(\mu) Q_s P_a(\Delta)$ | -170.91 | -171.57 | -171.24 | -171.15 | -171.22 | 0.27 | -172.21 |
| $P_a(\mu) Q_a P_d(\Delta)$ | -152.74 | -152.89 | -152.72 | -152.76 | -152.78 | 0.07 | -152.78 |
| $P_a(\mu) Q_a P_a(\Delta)$ | -148.27 | -148.18 | -148.17 | -148.27 | -148.22 | 0.05 | -148.22 |
| $P_a(\mu) Q_s P_d(\Delta)$ | -152.76 | -152.36 | -152.54 | -152.69 | -152.59 | 0.18 | -152.57 |
| $\boldsymbol{P_a(\mu) Q_s P_a(\Delta)}$ | **-147.24** | **-147.46** | **-147.24** | **-147.33** | **-147.32** | **0.11** | **-147.30** |

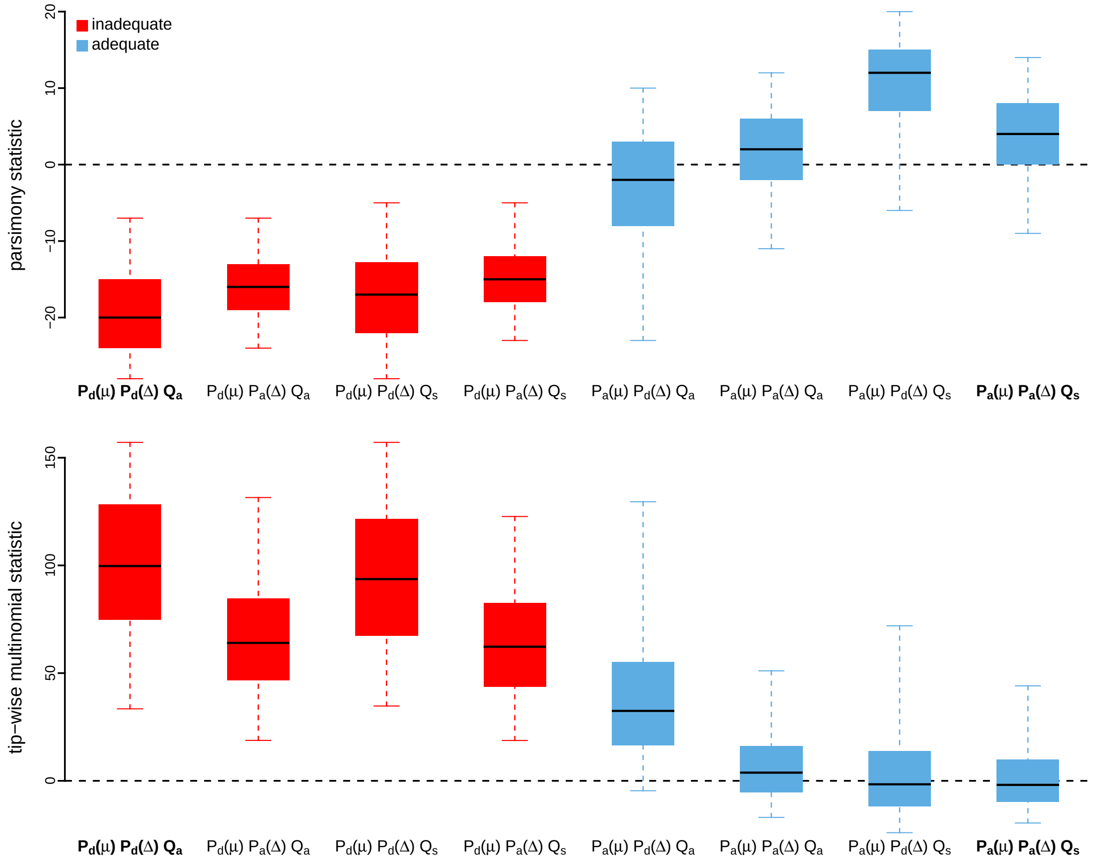


**Figure S17: Posterior-predictive distributions of the parsimony statistic (top panel) and the tip-wise multinomial statistic (bottom panel) under each of the eight prior models for the Dengue virus dataset.** Boxplots depict the posterior-predictive distributions of the statistic under each of the eight candidate prior models: the center of each box is the median predictive value of the summary statistic; the box and whiskers indicate the corresponding 50% and 95% posterior-predictive intervals, respectively. The horizontal dashed line indicates when the simulated and observed datasets produce identical value for the summary statistic. A model is judged to be inadequate (*i.e.*, incapable of generating geographic datasets that are similar to the observed data) if its 95% posterior-predictive interval does not overlap with the dashed line. Here the preferred default prior model is inadequate, whereas the preferred alternative prior model is adequate. Notation for the candidate models is described in Table S6; the preferred default- and alternative-prior models are indicated in bold text.

*The Impact of Prior Choice on Average Dispersal Rate*

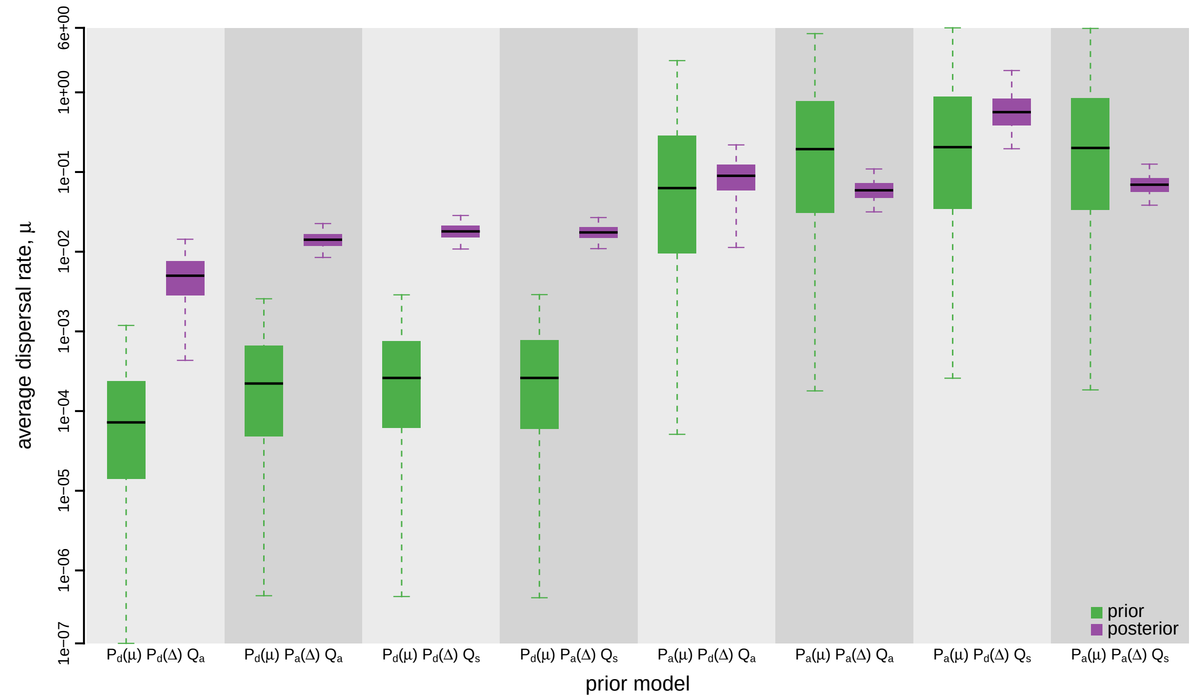


**Figure S18: The impact of prior choice on the average rate of dispersal for the Dengue virus dataset.** Each pair of boxplots depicts the specified prior (green) and corresponding posterior estimate (purple) of the average dispersal rate, $\mu$, under each prior model: the center of each box indicates the median rate; the box and whiskers indicate the corresponding 50% and 95% credible intervals, respectively. Notation for the prior models is as described in Table S6.

*The Impact of Prior Choice on Pairwise Dispersal Rates*

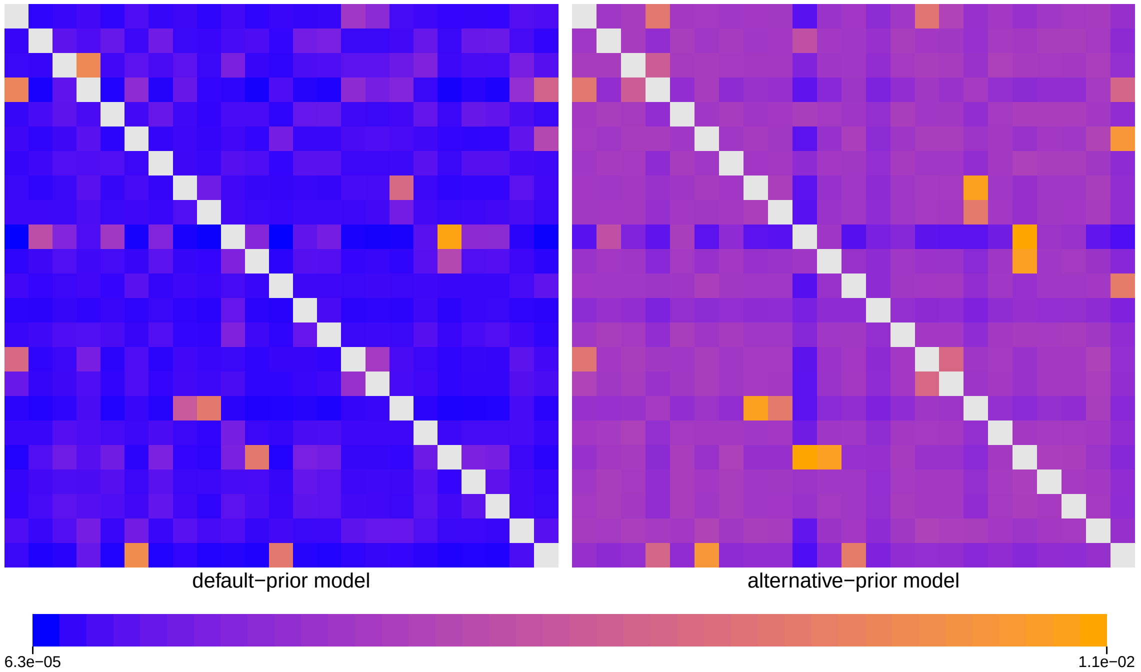


**Figure S19: The impact of prior choice on pairwise dispersal rates for the Dengue virus dataset.** Heatmaps summarize posterior-mean estimates of the instantaneous rate of dispersal between each pair of geographic areas, $q_{ij}$, under the default (left) and alternative (right) prior models.

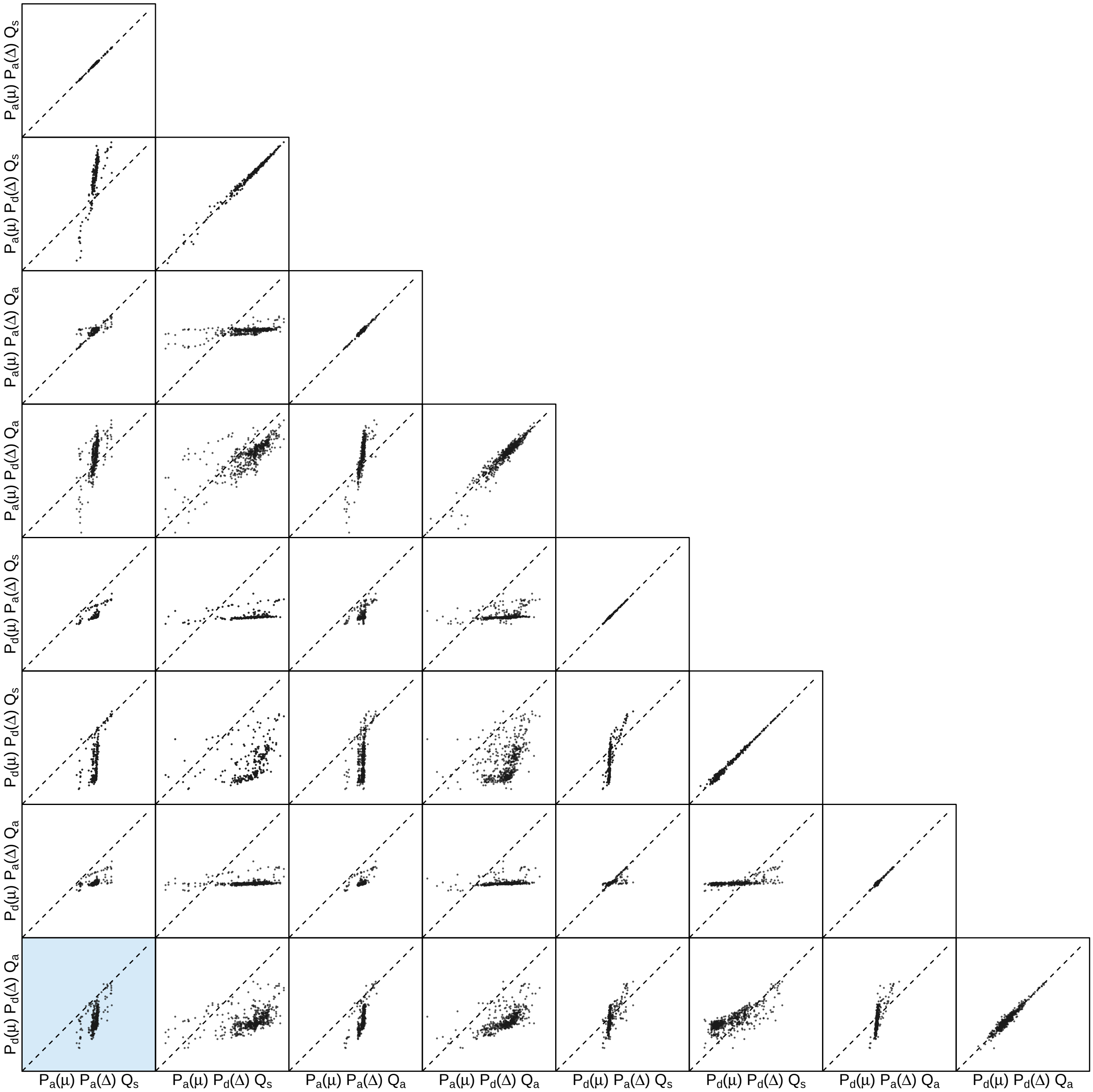


**Figure S20: The impact of prior choice on pairwise dispersal rates for the Dengue virus dataset.** Each cell of the plot compares posterior-mean estimate of the rate of dispersal between each pair of geographic areas, $q_{ij}$, between each pair of prior models. $x$- and $y$-axis of each cell are both on log scale. Diagonal cells are comparisons between half and the other half of the replicates under the same prior model, assessing the convergence of MCMC simulations; off-diagonal cells are comparisons between different prior models, demonstrating the impact of the prior model on the estimates of pairwise dispersal rates. The shaded cell corresponds to the comparison between the preferred default- and the preferred alternative-prior models (*c.f.*, Figure S19). Notation for the prior models is as described in Table S6.

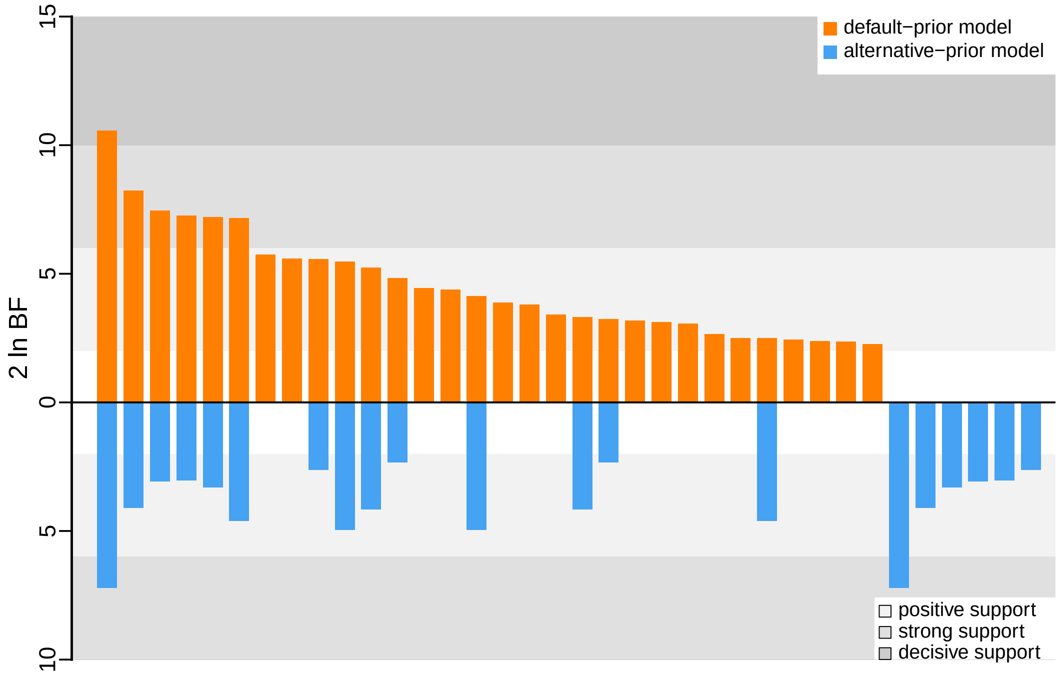


**Figure S21: The impact of prior choice on the inferred support for dispersal routes for the Dengue virus dataset.** We compare the evidential support for each dispersal route for the Dengue virus dataset under the default (orange) and alternative (blue) prior models. Each bar indicates the $2 \ln \mathrm{BF}$ for the corresponding dispersal route between two areas; only supported dispersal routes (*i.e.*, $2 \ln \mathrm{BF} > 2$) are plotted.

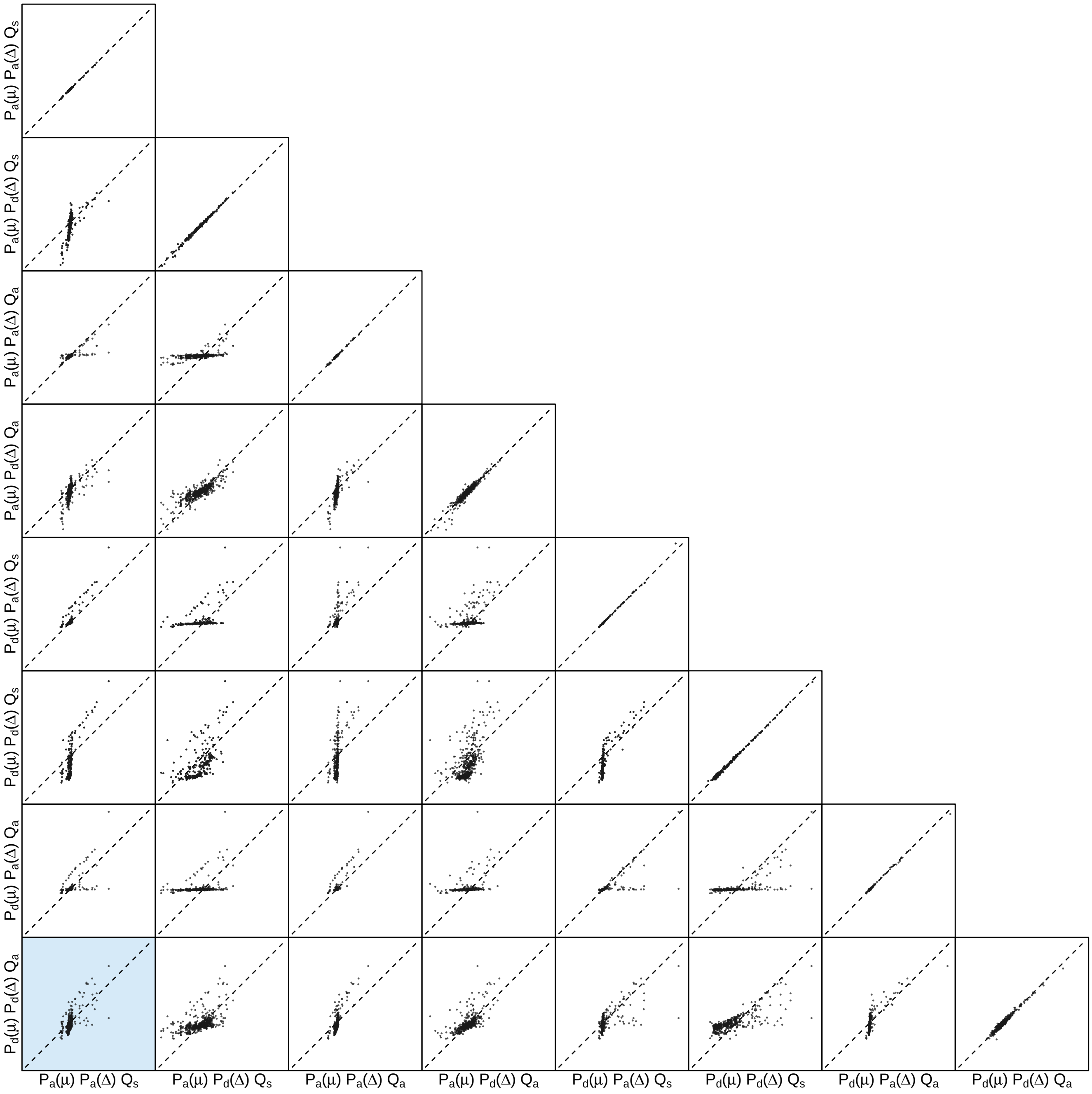


**Figure S22: The impact of prior choice on the inferred support for dispersal routes for the Dengue virus dataset.** Each cell of the plot compares the inferred support ($2 \ln \mathrm{BF}$) for pairwise dispersal routes between each pair of prior models. Diagonal cells are comparisons between half and the other half of the replicates under the same prior model, assessing the convergence of MCMC simulations; off-diagonal cells are comparisons between different prior models, demonstrating the impact of the prior model on the inferred support for pairwise dispersal routes. The shaded cell corresponds to the comparison between the preferred default- and the preferred alternative-prior models (*c.f.*, Figure S21). Notation for the prior models is as described in Table S6.

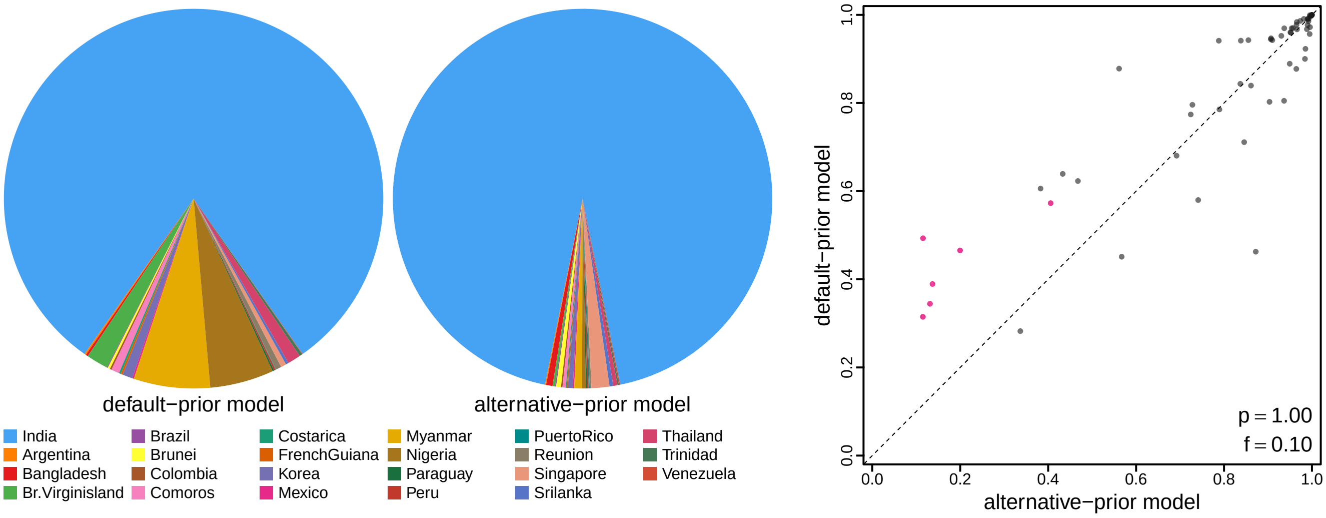


**Figure S23: The impact of prior choice on ancestral-area estimates for the Dengue virus dataset.** The left panel compares the posterior probability of each ancestral area at the root node under the default- and alternative-prior models for the Dengue virus dataset. The right panel plots the posterior probability of the most probable ancestral area under the default-prior model for each node in the MCC tree (y-axis) against the corresponding posterior probability of that area under the alternative-prior model (x-axis). Pink dots represent the internal nodes where the MAP ancestral area inferred under the default-prior model differs from that inferred under the alternative-prior models. The statistic $p$ denotes the fraction of internal nodes that are shared under the default- and alternative-prior models; $f$, is the fraction of shared nodes where the MAP ancestral area differs under the default- and alternative-prior models. Note that the posterior probabilities of the MAP ancestral area under the default-prior model are generally higher than those under the alternative-prior model (*i.e.*, the default-prior model tends to mask uncertainty in the ancestral-area estimates).

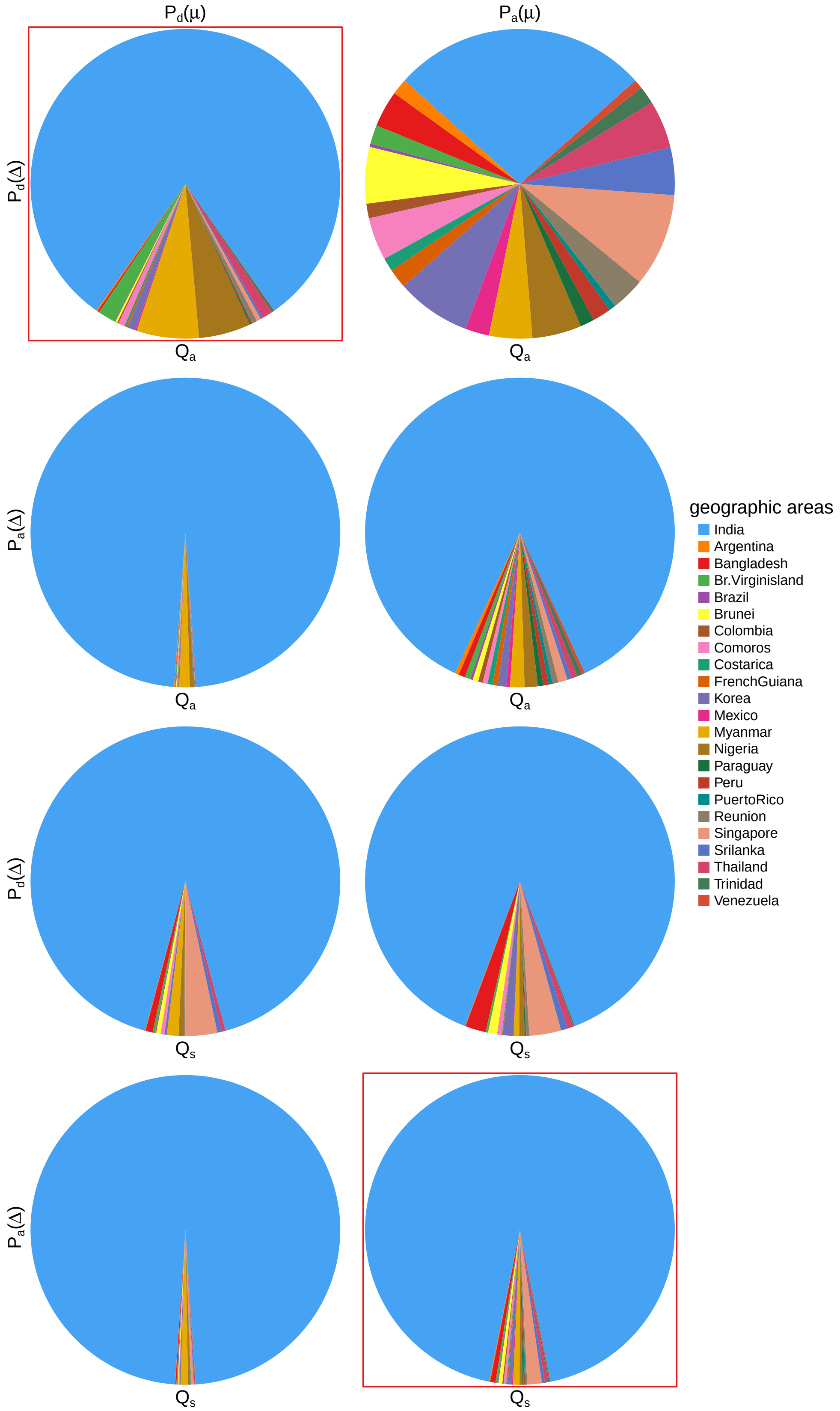


**Figure S24: The impact of prior choice on ancestral-area estimates at the root node for the Dengue virus dataset.** The posterior probability of each ancestral area at the root node under each prior model for the Dengue virus dataset. The pie chart enclosed in red corresponds to the comparison between the preferred default- and the preferred alternative-prior models (*c.f.*, Figure S23, left panel). Model notation follows the description in Table S6.

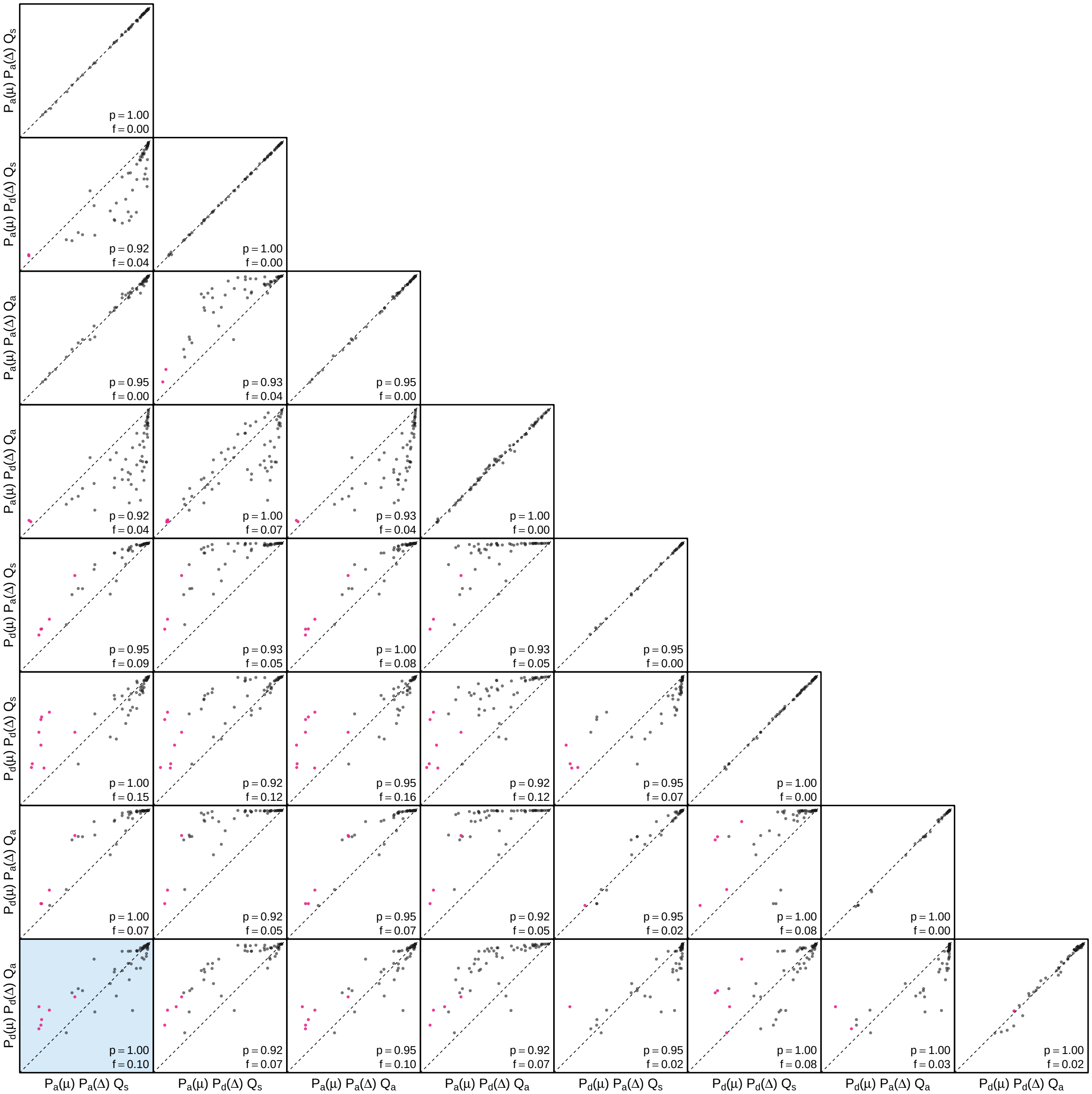


**Figure S25: The impact of prior choice on the MAP ancestral-area estimates at internal nodes for the Dengue virus dataset.** Pairwise scatter plots compare the posterior probability of the MAP ancestral area inferred under each combination of prior models. Diagonal cells are comparisons between two replicates under the same prior model (assessing the convergence of MCMC simulations); off-diagonal cells are comparisons between different prior models, demonstrating the impact of the prior model on both the posterior probability and the identity of the MAP ancestral-area estimates at internal nodes. Pink dots represent the internal nodes where the MAP ancestral area inferred under the default-prior model differs from that inferred under the alternative-prior models. The statistic $p$ denotes the fraction of internal nodes that are shared under the default- and alternative-prior models; $f$ is the fraction of shared nodes where the MAP ancestral area differs under the default- and alternative-prior models. The shaded cell corresponds to the comparison between the preferred default- and the preferred alternative-prior models (*c.f.*, Figure S23, right panel). Model notation follows the description in Table S6.

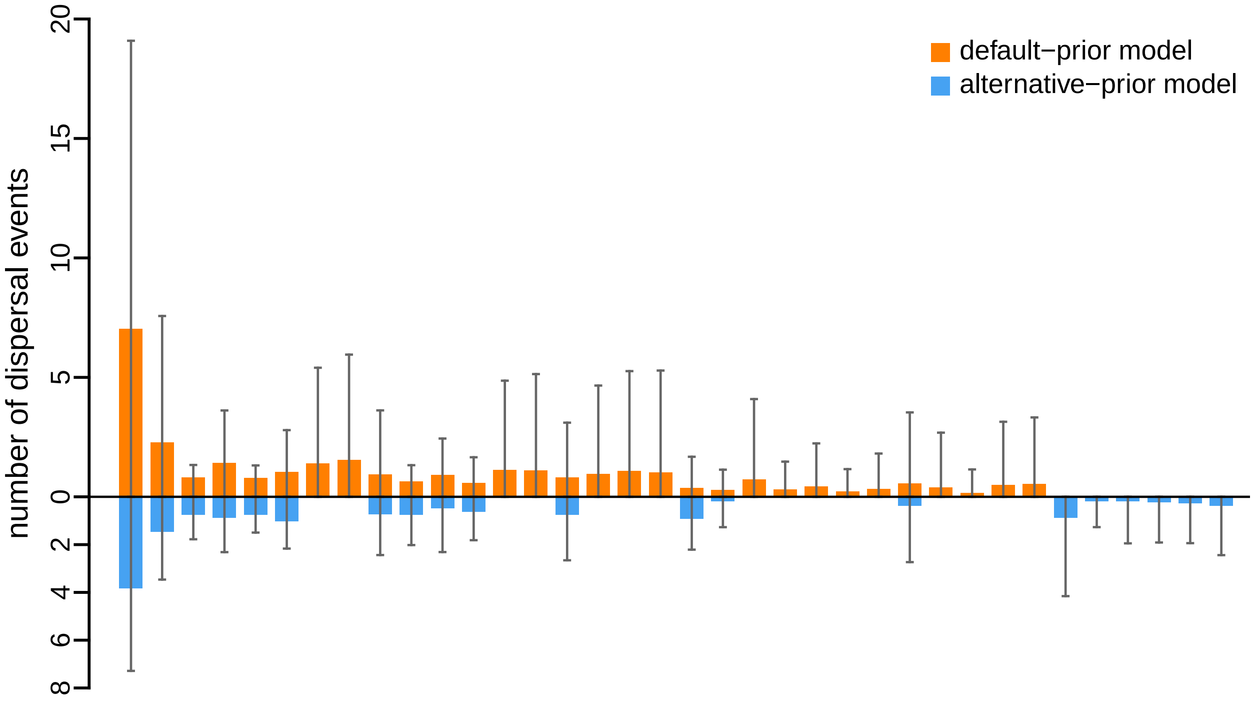


**Figure S26: The impact of prior choice on the inferred number of dispersal events between each pair of areas for the Dengue virus dataset.** The reflected bar plot depicts the number of dispersal events inferred under the default (orange) and alternative (blue) prior models for the Dengue virus dataset. Each bar indicates the posterior-mean number of dispersal events between a pair of areas; whiskers indicate the 95% credible interval. Note that only the number of dispersal events over the "significant" dispersal routes (*i.e.*, $2 \ln \mathrm{BF} > 2$) are figured.

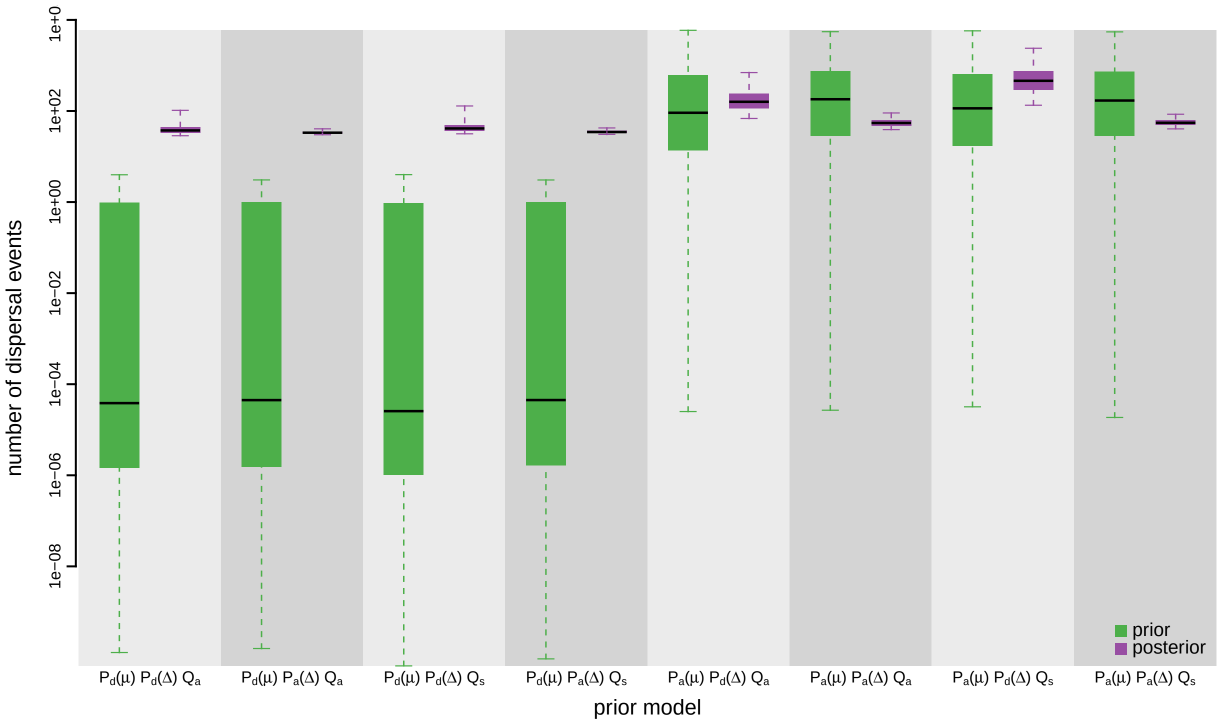


**Figure S27: The impact of prior choice on the inferred total number of dispersal events between all areas for the Dengue virus dataset.** Each pair of boxplots depicts the specified prior (green) and corresponding posterior estimate (purple) of the number of dispersal events between all areas under each prior model: the center of each box indicates the posterior-median number of dispersal events; the box and whiskers indicate the corresponding 50% and 95% credible intervals, respectively. Model notation follows the description in Table S6.

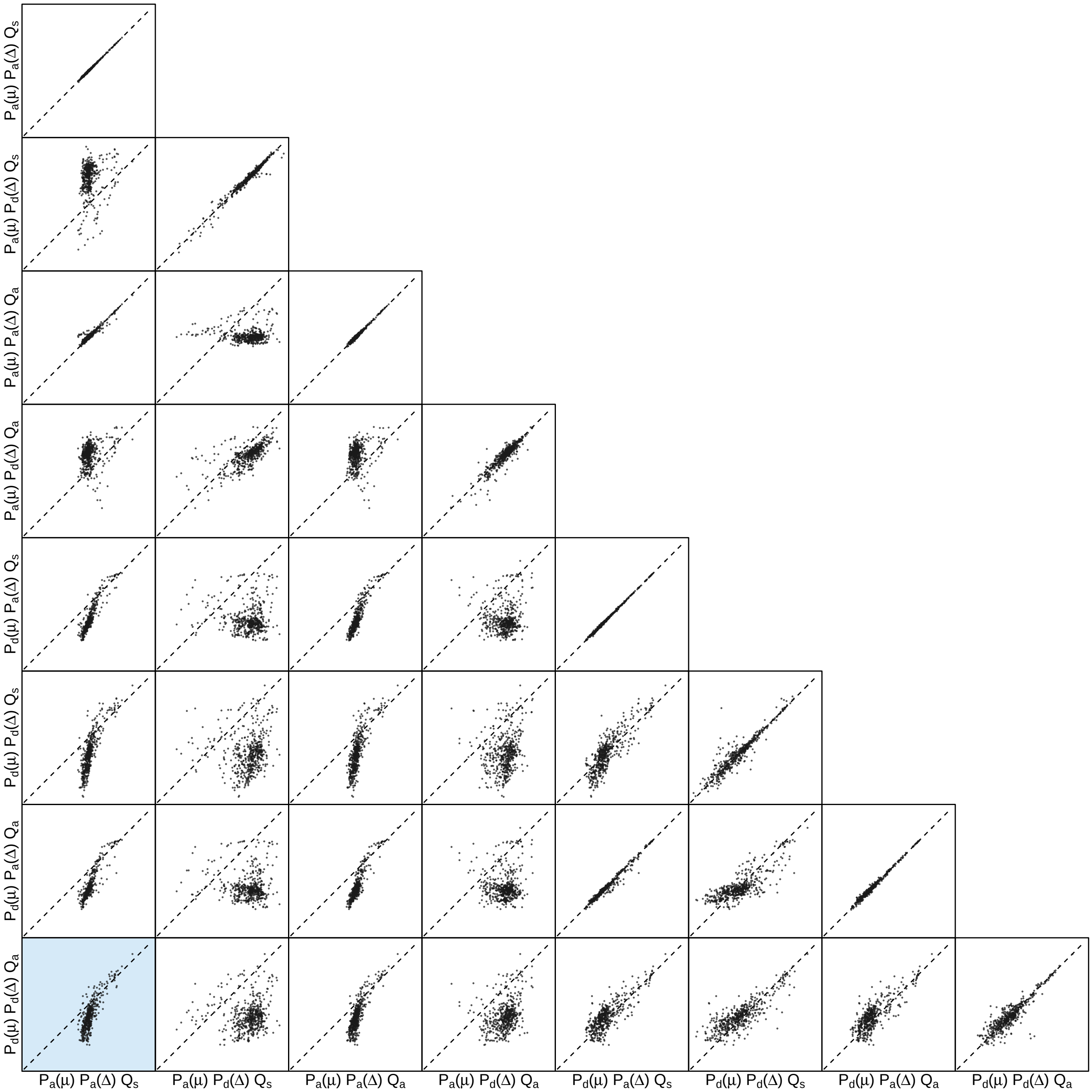


**Figure S28: The impact of prior choice on the inferred number of dispersal events between each pair of areas for the Dengue virus dataset.** Each cell of the plot compares the inferred number of pairwise dispersal events between each pair of prior models. *x*- and *y*-axis of each cell are both on log scale. Diagonal cells are comparisons between half and the other half of the replicates under the same prior model, assessing the convergence of MCMC simulations; off-diagonal cells are comparisons between different prior models, demonstrating the impact of the prior model on the inferred number of pairwise dispersal events. The shaded cell corresponds to the comparison between the preferred default- and the preferred alternative-prior models (*c.f.*, Figure S26). Model notation follows the description in Table S6.

###### S3.3.2 Deformed Wing Virus

Wilfert et al. (2016) explored the role of Varroa in the spread of DWV in honeybees (*i.e.*, whether they were the source of the virus or merely facilitated its spread among honeybees), and also identified significant dispersal routes of DWV between geographic populations. This study contains six datasets, including three molecular sequence alignments (lp, rdrp, and vp3) and two discrete-trait datasets (geographic areas and host species). We re-inferred the biogeographic history for each of the three molecular datasets (see Wilfert et al. (2016) for details about these datasets).

We acquired the `BEAST` XML scripts used in the original study—containing both the sequence alignment and sampling time and geographic location data—directly from the authors. For each gene region, we inferred the marginal posterior distribution of phylogenies under the phylogenetic models identical to those specified in Wilfert et al. (2016). Specifically, for each gene region we partitioned the alignments into two subsets (where the first subset included sites at the first and second codon positions, and the second subset included sites at the third codon position), and specified independent substitution models for each of these two partitions. Our phylogenetic models assume that the two data partitions share the same uncorrelated exponential (UCED) branch-rate model (Drummond et al. 2006; Rannala and Yang 2007), and the same exponential coalescent node-age model, but we specified a rate multiplier for each data subset to allow the average substitution rate to vary among data partitions. We constrained the mean of these rate multipliers to one so that these rates are identifiable under the uncorrelated branch-rate model. The phylogenetic models were identical for all three gene regions except for the substitution model specified for the partitions of each alignment. Following Wilfert et al. (2016), we specified independent TN93+I+$\Gamma_4$ model (Tamura and Nei 1993; Gu et al. 1995; Yang 1994) for the two partitions of the lp fragment, and specified independent HKY+$\Gamma_4$ model (Hasegawa et al. 1984, 1985) for the two partitions of the rdrp fragment, and specified independent HKY+I model for the two partitions of the vp3 fragment. Details of these analyses are available in the XML scripts included in our GitHub and Dryad repositories.

For each gene region, we ran four independent MCMC simulations in `BEAST` version 1.8.2 for 100 million generations each, sampling every 10000 generations (except for the rdrp fragment, where we it was necessary to run simulations for 200 million cycles and sample every 20000 generations to achieve adequate MCMC performance). We first assessed the performance of each MCMC simulation, removed the first 10% of samples from each chain as the burn-in, and then combined the remaining posterior samples of trees from the replicate simulations using `LogCombiner` version 1.8.2. This resulted in a posterior sample of 360 trees (available in our GitHub and Dryad repositories), which we then used as the prior distribution of phylogenies.

The MCMC simulations used in the second step of our sequential analyses and of the analyses used to estimate marginal likelihoods under each prior model are described above in Section S3.1. Details of these analyses are available in the XML scripts included in our GitHub and Dryad repositories.

###### S3.3.2.1 Lp fragment

*The Impact of Prior Choice on Biogeographic Model Fit*

**Table S7: Marginal-likelihood estimates of each of the eight prior models for the Deformed wing virus lp fragment dataset.** Columns 2–5 list marginal likelihoods inferred from four replicate analyses. Columns 6–7 list the mean and standard deviation of these marginal-likelihood estimates. The last column lists the marginal-likelihood estimates computed by combining the samples from the replicate power-posterior MCMC simulations. Candidate models are listed in rows and include all possible combinations of: (1) instantaneous-rate matrices (symmetric, $Q_s$ or asymmetric, $Q_a$); (2) priors on the average dispersal rate [default, $P_d(\mu)$ or alternative, $P_a(\mu)$], and; (3) priors on the number of dispersal routes [default, $P_d(\Delta)$ or alternative, $P_a(\Delta)$]. The preferred default- and alternative-prior models are indicated in bold text.

| Model | replicate1 | replicate2 | replicate3 | replicate4 | mean | sd | combined |
|---|---|---|---|---|---|---|---|
| $\boldsymbol{P_d(\mu)Q_aP_d(\Delta)}$ | **-142.48** | **-142.51** | **-142.53** | **-142.58** | **-142.52** | **0.04** | **-142.52** |
| $P_d(\mu)Q_aP_a(\Delta)$ | -140.30 | -140.30 | -140.42 | -140.32 | -140.34 | 0.06 | -140.33 |
| $P_d(\mu)Q_sP_d(\Delta)$ | -145.12 | -145.14 | -145.03 | -144.95 | -145.06 | 0.09 | -145.07 |
| $P_d(\mu)Q_sP_a(\Delta)$ | -141.90 | -141.88 | -141.62 | -141.63 | -141.76 | 0.15 | -141.72 |
| $P_a(\mu)Q_aP_d(\Delta)$ | -129.61 | -129.38 | -129.51 | -129.48 | -129.50 | 0.09 | -129.49 |
| $\boldsymbol{P_a(\mu)Q_aP_a(\Delta)}$ | **-128.73** | **-128.82** | **-128.75** | **-128.73** | **-128.76** | **0.04** | **-128.76** |
| $P_a(\mu)Q_sP_d(\Delta)$ | -133.11 | -133.43 | -133.43 | -133.48 | -133.36 | 0.17 | -133.39 |
| $P_a(\mu)Q_sP_a(\Delta)$ | -129.85 | -129.66 | -129.76 | -129.71 | -129.74 | 0.08 | -129.74 |

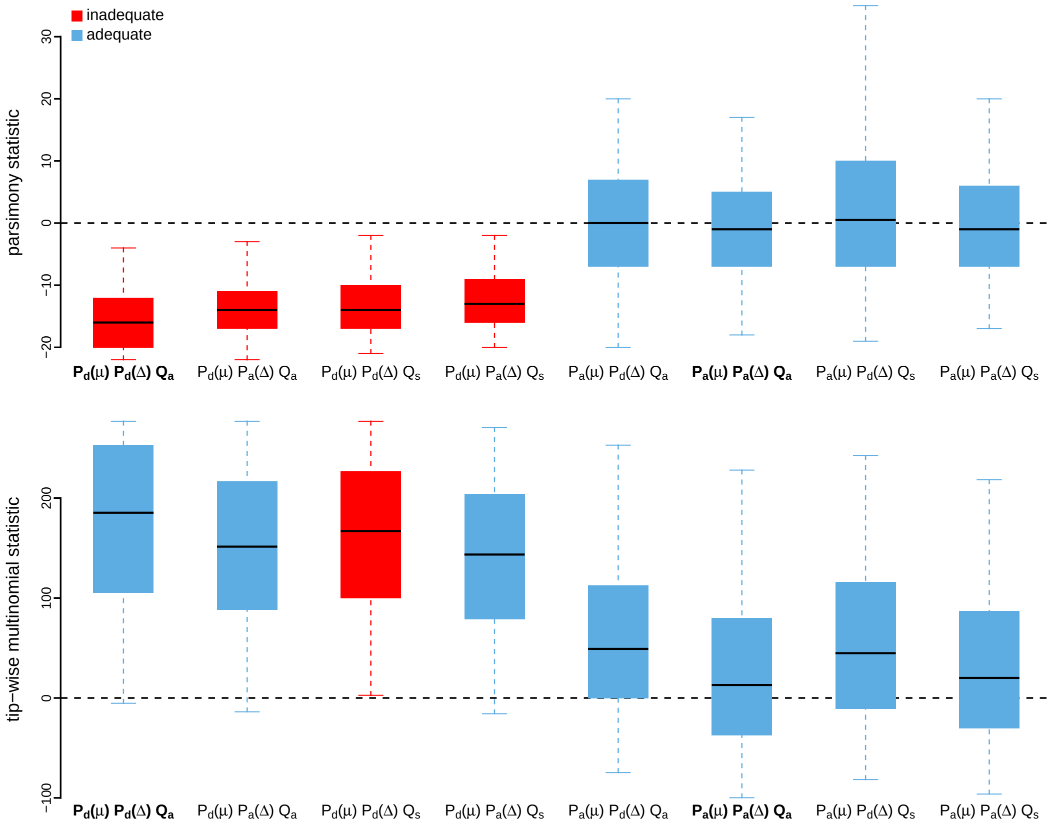


**Figure S29: Posterior-predictive distributions of the parsimony statistic (top panel) and the tip-wise multinomial statistic (bottom panel) under each of the eight prior models for the Deformed wing virus lp fragment dataset.** Boxplots depict the posterior-predictive distributions of the statistic under each of the eight candidate prior models: the center of each box is the median predictive value of the summary statistic; the box and whiskers indicate the corresponding 50% and 95% posterior-predictive intervals, respectively. The horizontal dashed line indicates when the simulated and observed datasets produce identical value for the summary statistic. A model is judged to be inadequate (*i.e.*, incapable of generating geographic datasets that are similar to the observed data) if its 95% posterior-predictive interval does not overlap with the dashed line. Here the preferred default prior model is inadequate, whereas the preferred alternative prior model is adequate. Notation for the candidate models is described in Table S7; the preferred default- and alternative-prior models are indicated in bold text.

*The Impact of Prior Choice on Average Dispersal Rate*

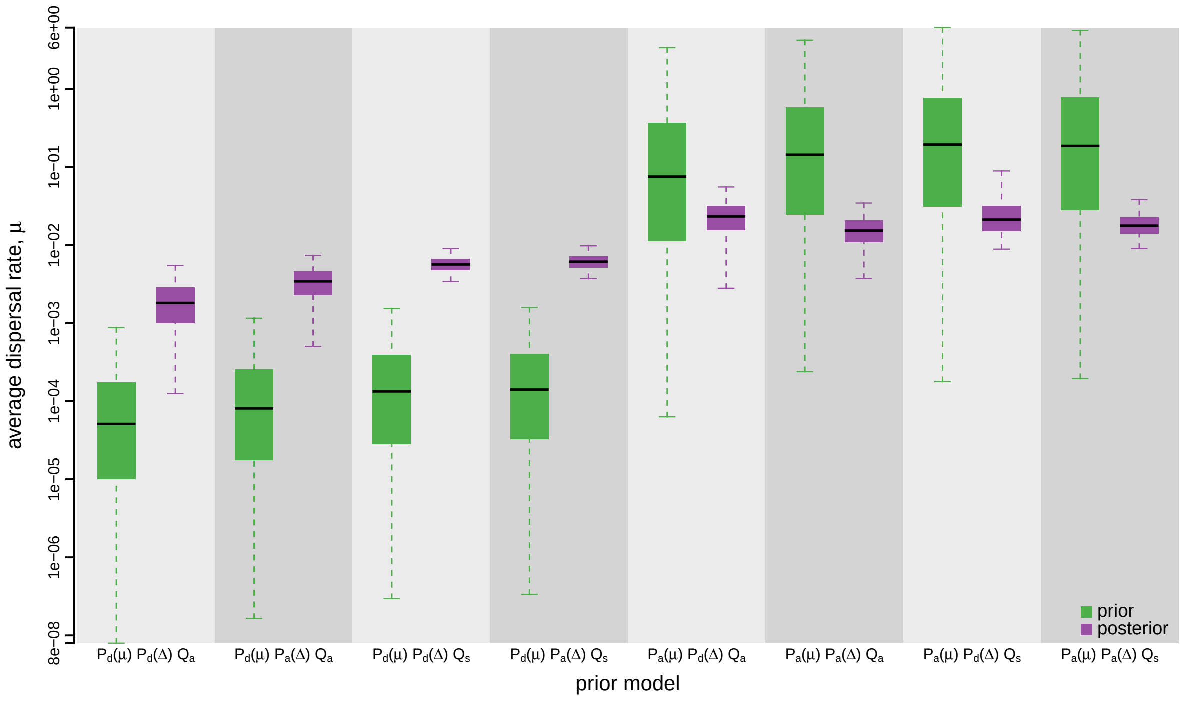


**Figure S30: The impact of prior choice on the average dispersal rate for the Deformed wing virus lp fragment dataset.** Each pair of boxplots depicts the specified prior (green) and corresponding posterior estimate (purple) of the average dispersal rate, $\mu$, under each prior model: the center of each box indicates the median rate; the box and whiskers indicate the corresponding 50% and 95% credible intervals, respectively. Notation for the prior models is as described in Table S7.

*The Impact of Prior Choice on Pairwise Dispersal Rates*

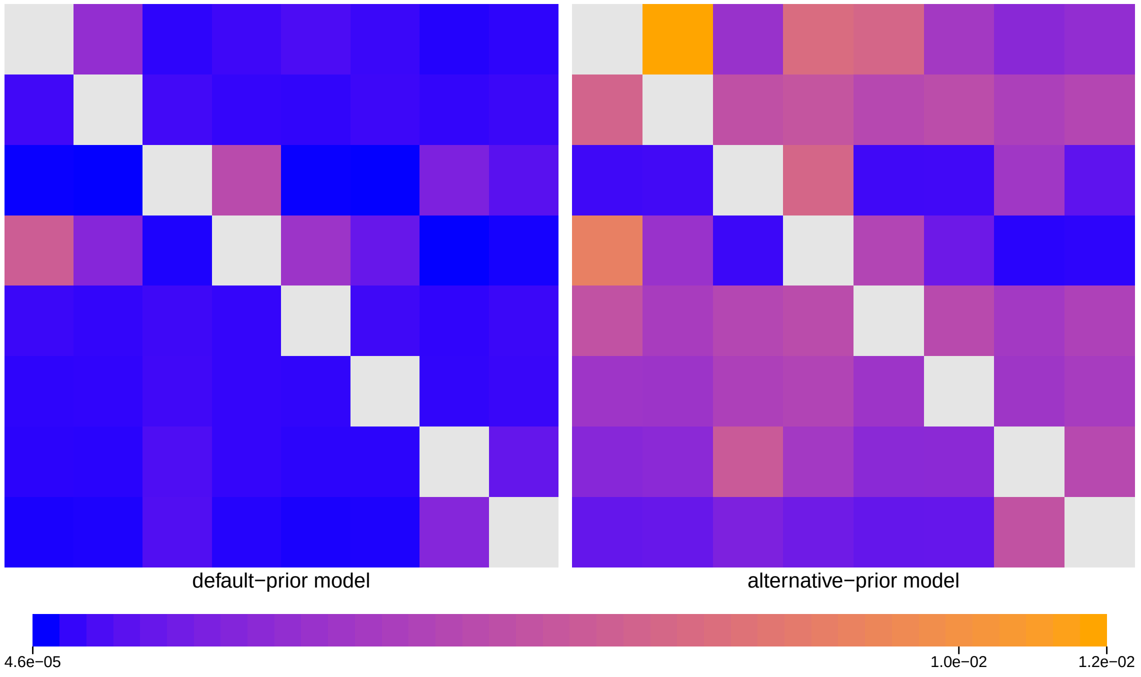


**Figure S31: The impact of prior choice on pairwise dispersal rates for the Deformed wing virus lp fragment dataset.** Heatmaps summarize posterior-mean estimates of the instantaneous rate of dispersal between each pair of geographic areas, $q_{ij}$, under the default (left) and alternative (right) prior models.

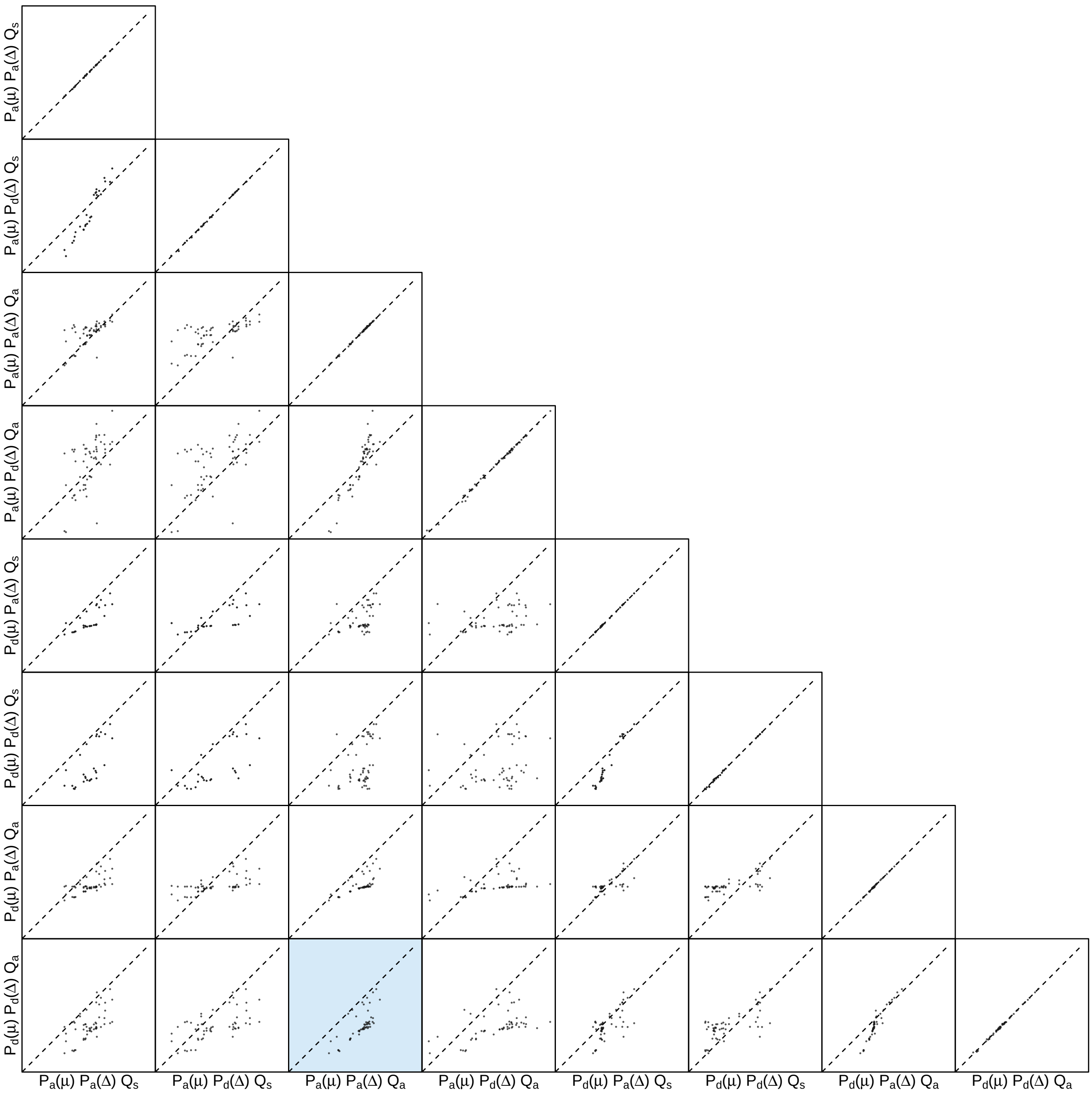


**Figure S32: The impact of prior choice on pairwise dispersal rates for the Deformed wing virus lp fragment dataset.** Each cell of the plot compares posterior-mean estimate of the rate of dispersal between each pair of geographic areas, $q_{ij}$, between each pair of prior models. $x$- and $y$-axis of each cell are both on log scale. Diagonal cells are comparisons between half and the other half of the replicates under the same prior model, assessing the convergence of MCMC simulations; off-diagonal cells are comparisons between different prior models, demonstrating the impact of the prior model on the estimates of pairwise dispersal rates. The shaded cell corresponds to the comparison between the preferred default- and the preferred alternative-prior models (*c.f.*, Figure S31). Notation for the prior models is as described in Table S7.

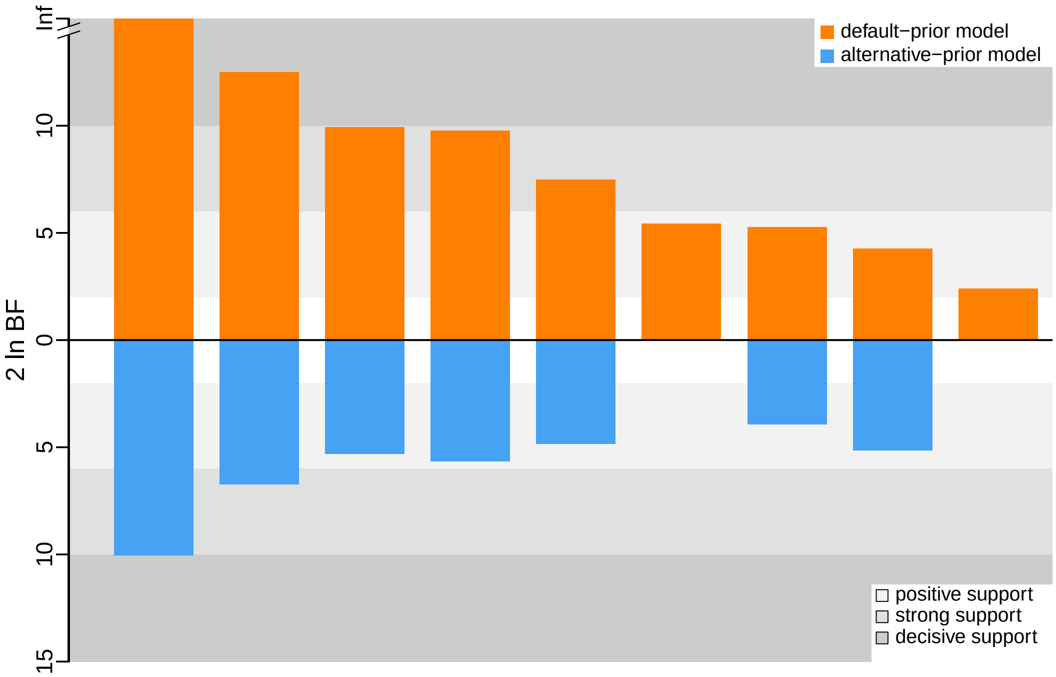


**Figure S33: The impact of prior choice on the inferred support for dispersal routes for the Deformed wing virus lp fragment dataset.** We compare the evidential support for each dispersal route for the Deformed wing virus lp fragment dataset under the default (orange) and alternative (blue) prior models. Each bar indicates the $2 \ln \text{BF}$ for the corresponding dispersal route between two areas; only supported dispersal routes (*i.e.*, $2 \ln \text{BF} > 2$) are plotted.

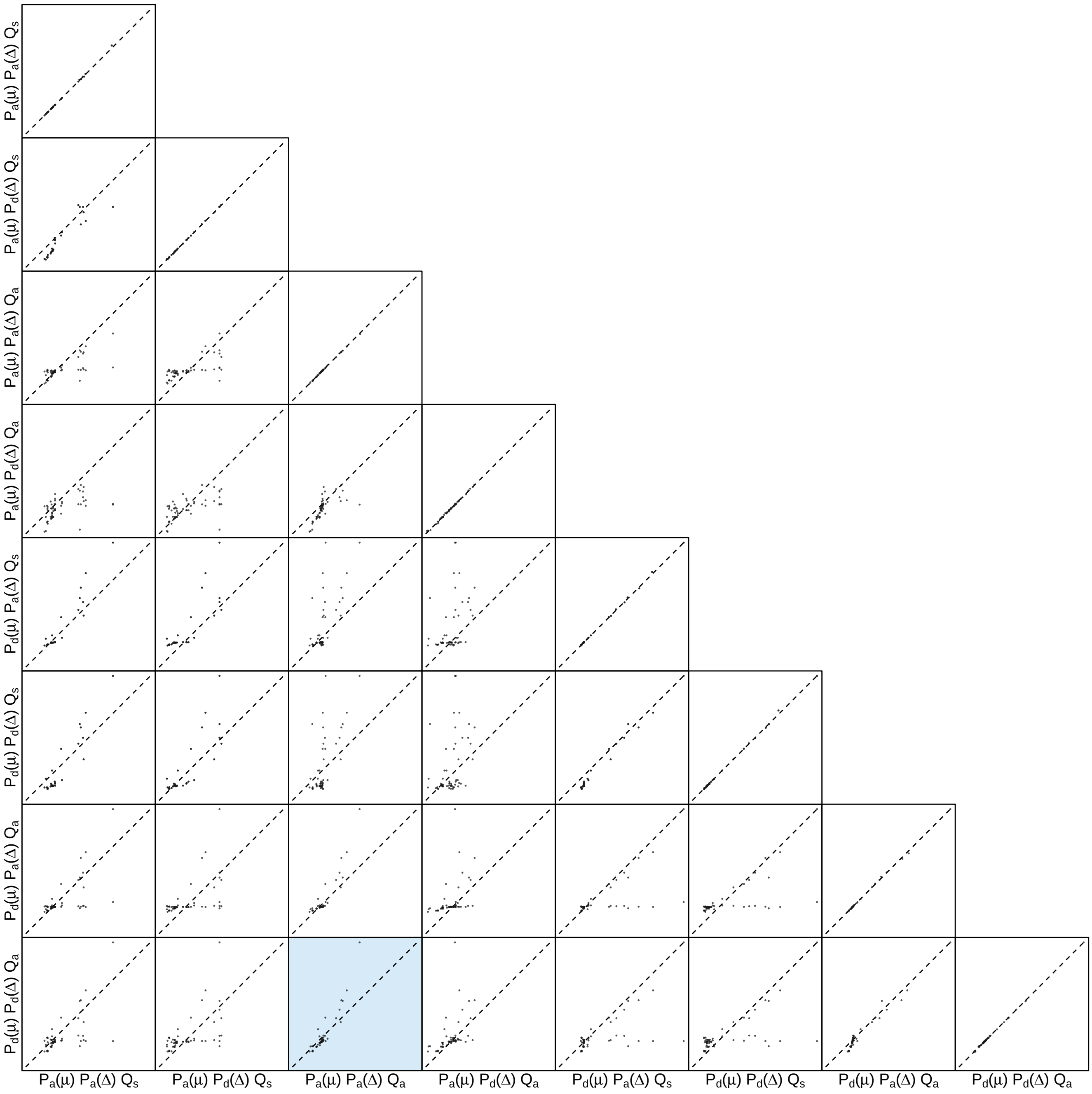


**Figure S34: The impact of prior choice on the inferred support for dispersal routes for the Deformed wing virus lp fragment dataset.** Each cell of the plot compares the inferred support ($2 \ln \mathrm{BF}$) for pairwise dispersal routes between each pair of prior models. Diagonal cells are comparisons between half and the other half of the replicates under the same prior model, assessing the convergence of MCMC simulations; off-diagonal cells are comparisons between different prior models, demonstrating the impact of the prior model on the inferred support for pairwise dispersal routes. The shaded cell corresponds to the comparison between the preferred default- and the preferred alternative-prior models (*c.f.*, Figure S33). Notation for the prior models is as described in Table S7.

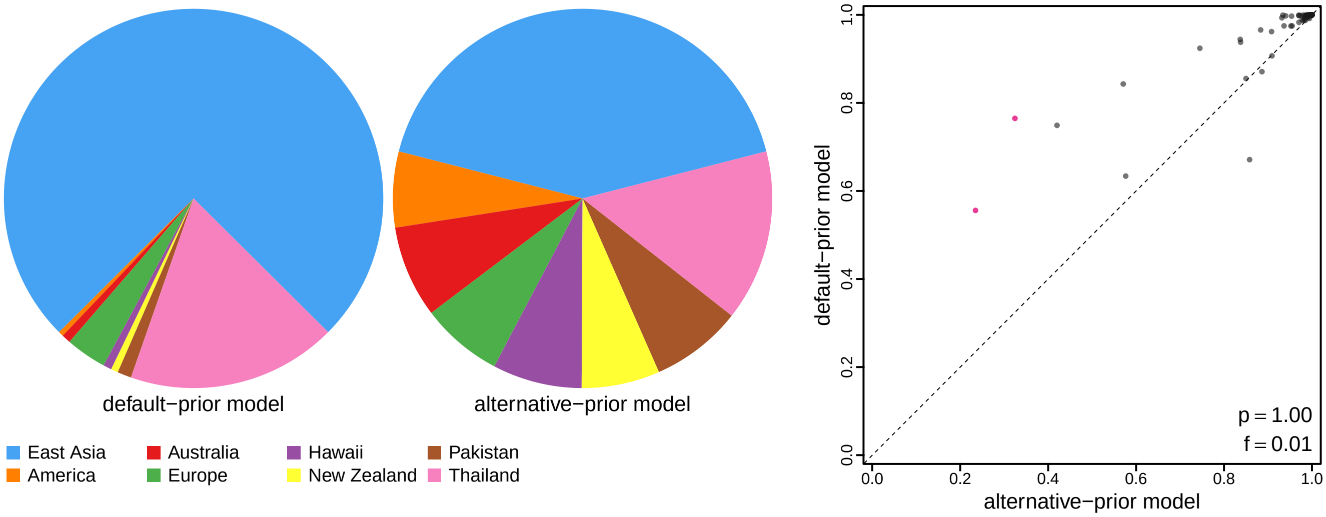


**Figure S35: The impact of prior choice on ancestral-area estimates for the Deformed wing virus lp fragment dataset.** The left panel compares the posterior probability of each ancestral area at the root node under the default- and alternative-prior models for the Deformed wing virus lp fragment dataset. The right panel plots the posterior probability of the most probable ancestral area under the default-prior model for each node in the MCC tree (y-axis) against the corresponding posterior probability of that area under the alternative-prior model (x-axis). Pink dots represent the internal nodes where the MAP ancestral area inferred under the default-prior model differs from that inferred under the alternative-prior models. The statistic $p$ denotes the fraction of internal nodes that are shared under the default- and alternative-prior models; $f$, is the fraction of shared nodes where the MAP ancestral area differs under the default- and alternative-prior models. Note that the posterior probabilities of the MAP ancestral area under the default-prior model are generally higher than those under the alternative-prior model (*i.e.*, the default-prior model tends to mask uncertainty in the ancestral-area estimates).

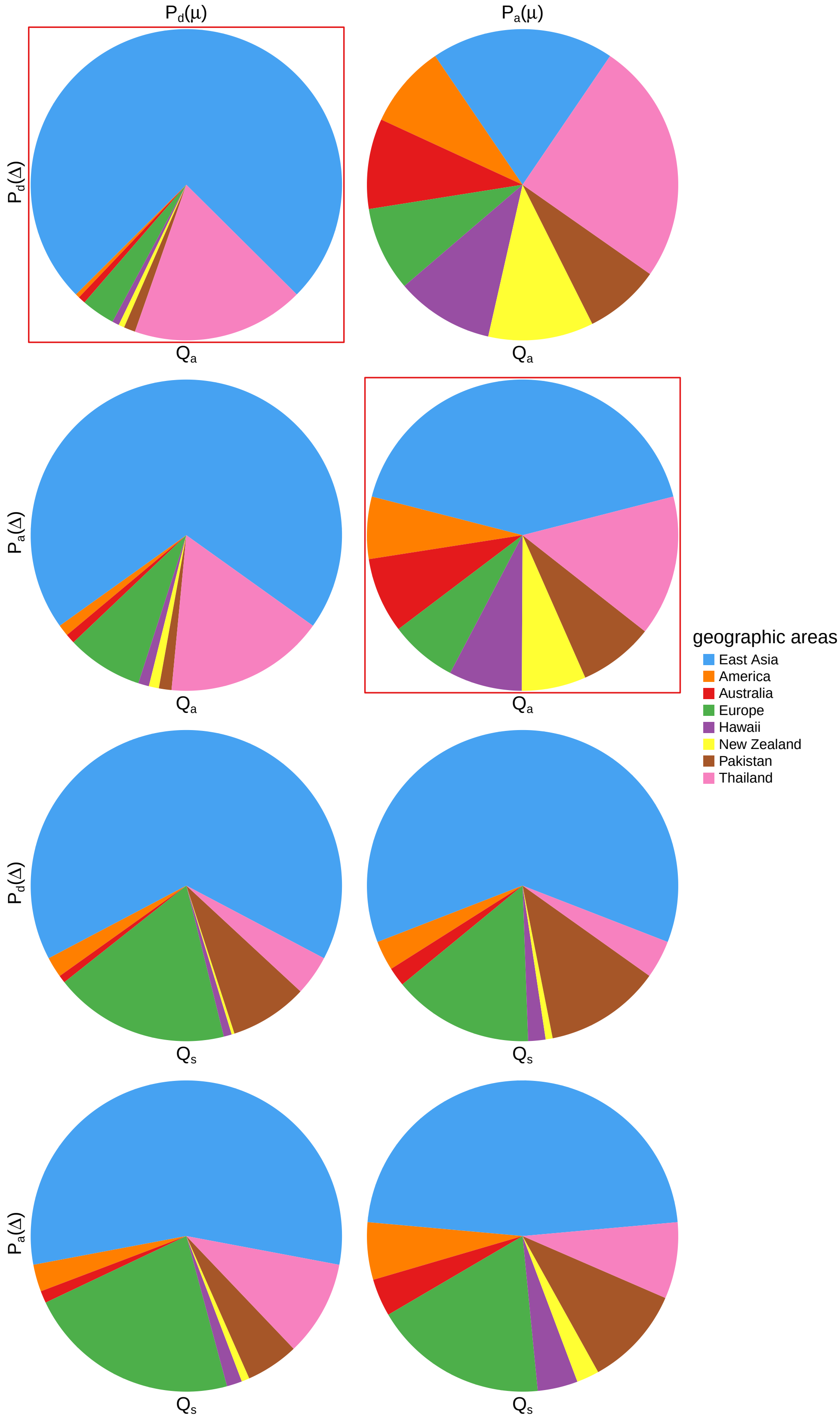


**Figure S36: The impact of prior choice on ancestral-area estimates at the root node for the Deformed wing virus lp fragment dataset.** The posterior probability of each ancestral area at the root node under each prior model for the Deformed wing virus lp fragment dataset. The pie chart enclosed in red corresponds to the comparison between the preferred default- and the preferred alternative-prior models (*c.f.*, Figure S35, left panel). Model notation follows the description in Table S7.

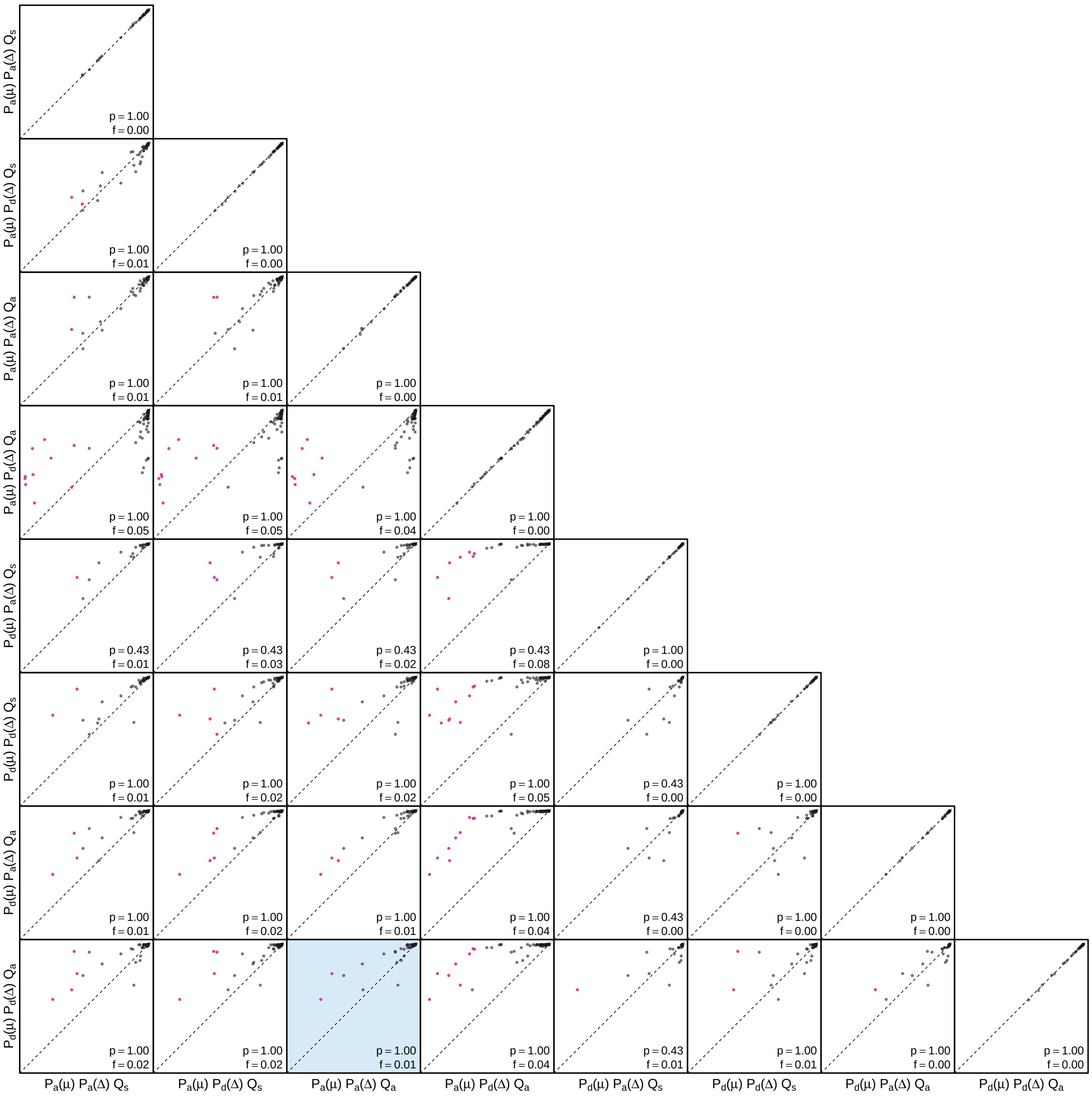


**Figure S37: The impact of prior choice on the MAP ancestral-area estimates at internal nodes for the Deformed wing virus lp fragment dataset.** Pairwise scatter plots compare the posterior probability of the MAP ancestral area inferred under each combination of prior models. Diagonal cells are comparisons between two replicates under the same prior model (assessing the convergence of MCMC simulations); off-diagonal cells are comparisons between different prior models, demonstrating the impact of the prior model on both the posterior probability and the identity of the MAP ancestral-area estimates at internal nodes. Pink dots represent the internal nodes where the MAP ancestral area inferred under the default-prior model differs from that inferred under the alternative-prior models. The statistic $p$ denotes the fraction of internal nodes that are shared under the default- and alternative-prior models; $f$ is the fraction of shared nodes where the MAP ancestral area differs under the default- and alternative-prior models. The shaded cell corresponds to the comparison between the preferred default- and the preferred alternative-prior models (*c.f.*, Figure S35, right panel). Model notation follows the description in Table S7.

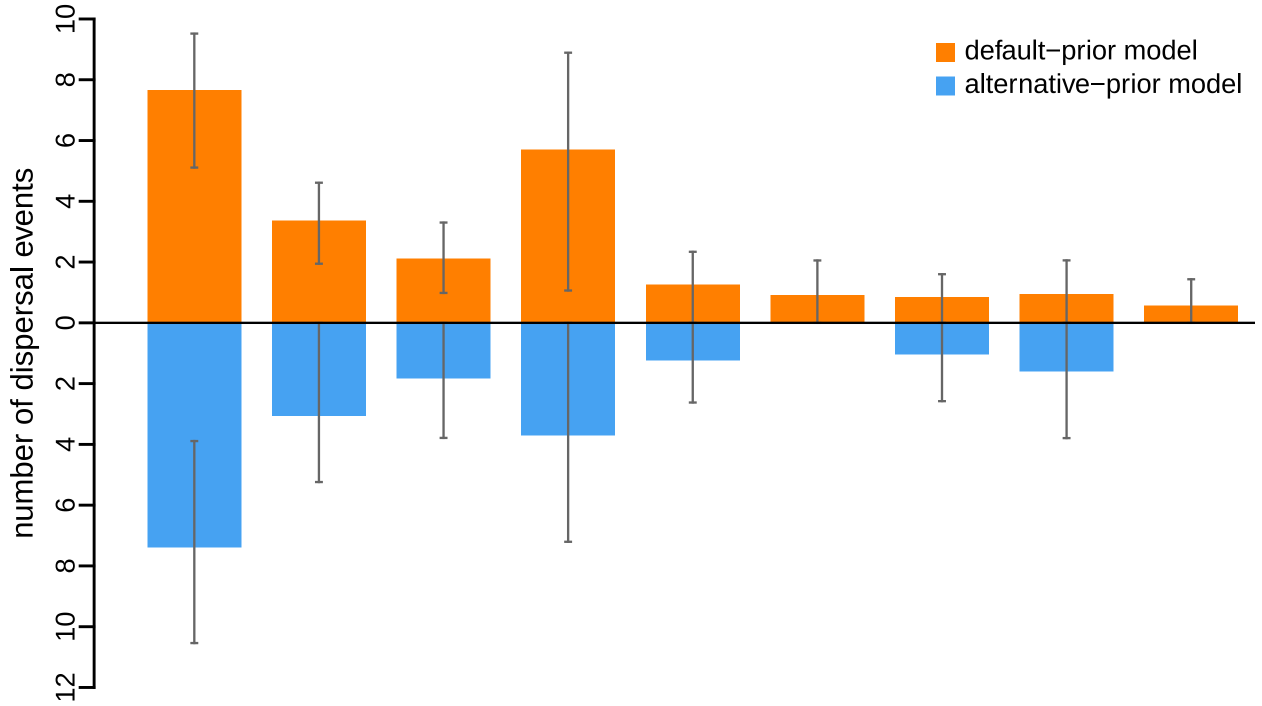


**Figure S38: The impact of prior choice on the inferred number of dispersal events between each pair of areas for the Deformed wing virus lp fragment dataset.** The reflected bar plot depicts the number of dispersal events inferred under the default (orange) and alternative (blue) prior models for the Deformed wing virus lp fragment dataset. Each bar indicates the posterior-mean number of dispersal events between a pair of areas; whiskers indicate the 95% credible interval. Note that only the number of dispersal events over the "significant" dispersal routes (*i.e.*, $2 \ln \mathrm{BF} > 2$) are figured.

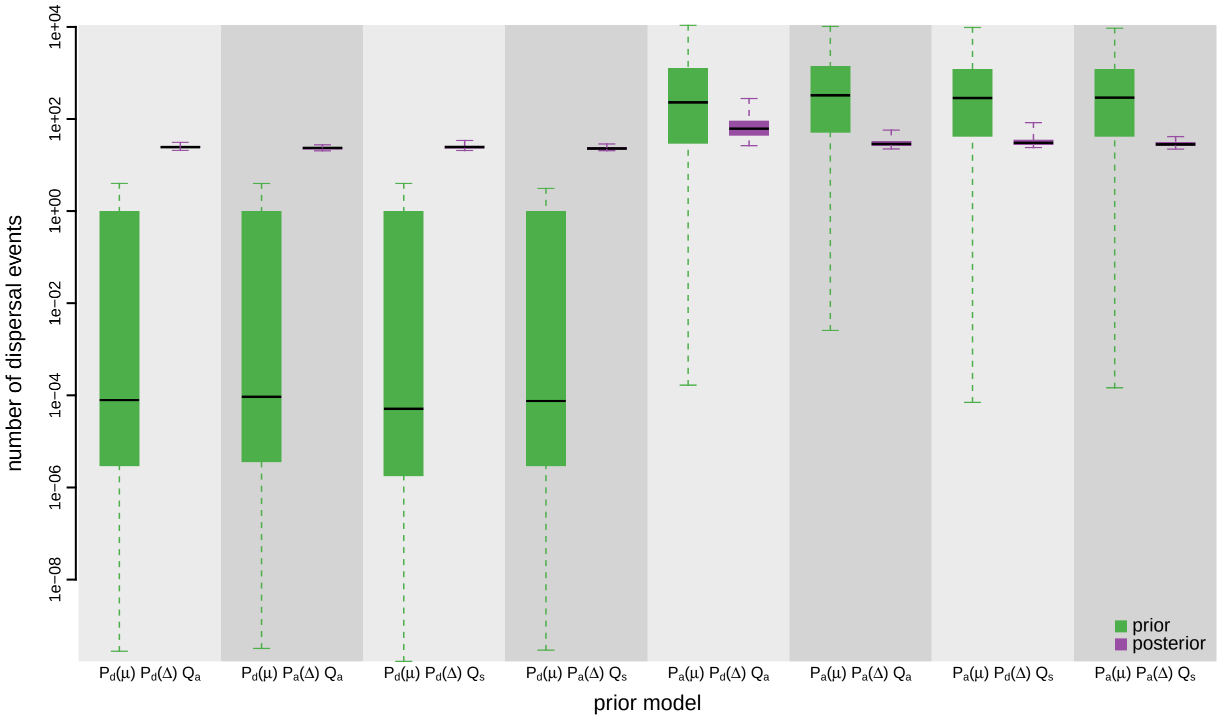


**Figure S39: The impact of prior choice on the inferred total number of dispersal events between all areas for the Deformed wing virus lp fragment dataset.** Each pair of boxplots depicts the specified prior (green) and corresponding posterior estimate (purple) of the number of dispersal events between all areas under each prior model: the center of each box indicates the posterior-median number of dispersal events; the box and whiskers indicate the corresponding 50% and 95% credible intervals, respectively. Model notation follows the description in Table S7.

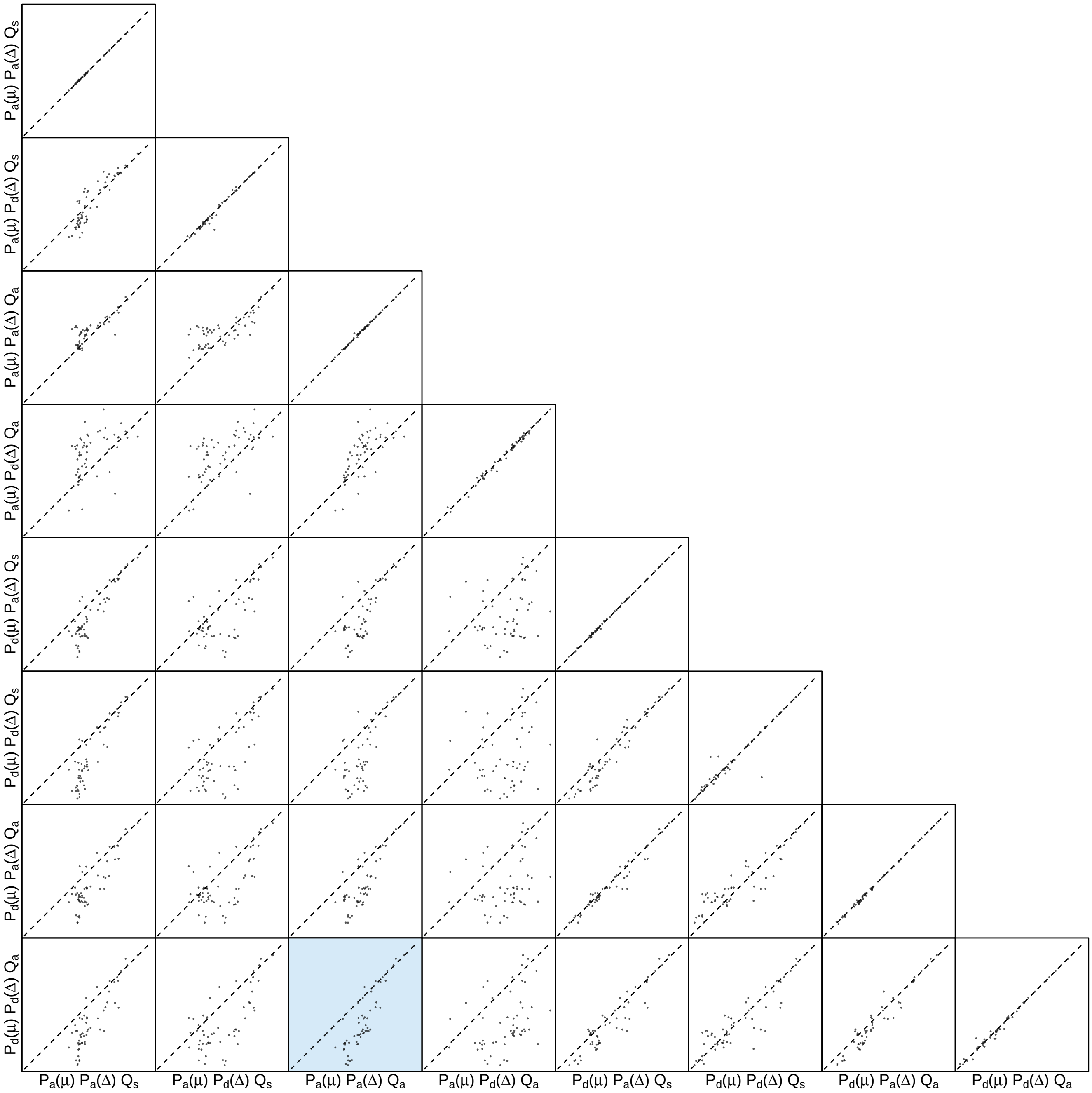


**Figure S40: The impact of prior choice on the inferred number of dispersal events between each pair of areas for the Deformed wing virus lp fragment dataset.** Each cell of the plot compares the inferred number of pairwise dispersal events between each pair of prior models. *x*- and *y*-axis of each cell are both on log scale. Diagonal cells are comparisons between half and the other half of the replicates under the same prior model, assessing the convergence of MCMC simulations; off-diagonal cells are comparisons between different prior models, demonstrating the impact of the prior model on the inferred number of pairwise dispersal events. The shaded cell corresponds to the comparison between the preferred default- and the preferred alternative-prior models (*c.f.*, Figure S38). Model notation follows the description in Table S7.

##### S3.3.2.2 Rdrp fragment

*The Impact of Prior Choice on Biogeographic Model Fit*

**Table S8: Marginal-likelihood estimates of each of the eight prior models for the Deformed wing virus rdrp fragment dataset.** Columns 2–5 list marginal likelihoods inferred from four replicate analyses. Columns 6–7 list the mean and standard deviation of these marginal-likelihood estimates. The last column lists the marginal-likelihood estimates computed by combining the samples from the replicate power-posterior MCMC simulations. Candidate models are listed in rows and include all possible combinations of: (1) instantaneous-rate matrices (symmetric, $Q_s$ or asymmetric, $Q_a$); (2) priors on the average dispersal rate [default, $P_d(\mu)$ or alternative, $P_a(\mu)$], and; (3) priors on the number of dispersal routes [default, $P_d(\Delta)$ or alternative, $P_a(\Delta)$]. The preferred default- and alternative-prior models are indicated in bold text.

| Model | replicate1 | replicate2 | replicate3 | replicate4 | mean | sd | combined |
|---|---|---|---|---|---|---|---|
| $P_d(\mu)Q_aP_d(\Delta)$ | -219.38 | -219.44 | -219.34 | -219.31 | -219.37 | 0.06 | -219.37 |
| $P_d(\mu)Q_aP_a(\Delta)$ | -216.87 | -216.89 | -216.82 | -216.88 | -216.87 | 0.03 | -216.87 |
| $\boldsymbol{P_d(\mu)Q_sP_d(\Delta)}$ | **-214.77** | **-215.04** | **-214.84** | **-214.93** | **-214.89** | **0.12** | **-214.83** |
| $P_d(\mu)Q_sP_a(\Delta)$ | -213.43 | -213.19 | -213.25 | -213.16 | -213.26 | 0.12 | -213.21 |
| $P_a(\mu)Q_aP_d(\Delta)$ | -171.71 | -171.78 | -171.66 | -171.56 | -171.68 | 0.09 | -171.67 |
| $\boldsymbol{P_a(\mu)Q_aP_a(\Delta)}$ | **-174.03** | **-173.98** | **-173.94** | **-174.11** | **-174.02** | **0.07** | **-174.01** |
| $P_a(\mu)Q_sP_d(\Delta)$ | -181.72 | -181.87 | -181.81 | -181.56 | -181.74 | 0.13 | -181.72 |
| $P_a(\mu)Q_sP_a(\Delta)$ | -181.86 | -181.55 | -181.81 | -181.91 | -181.78 | 0.16 | -181.82 |

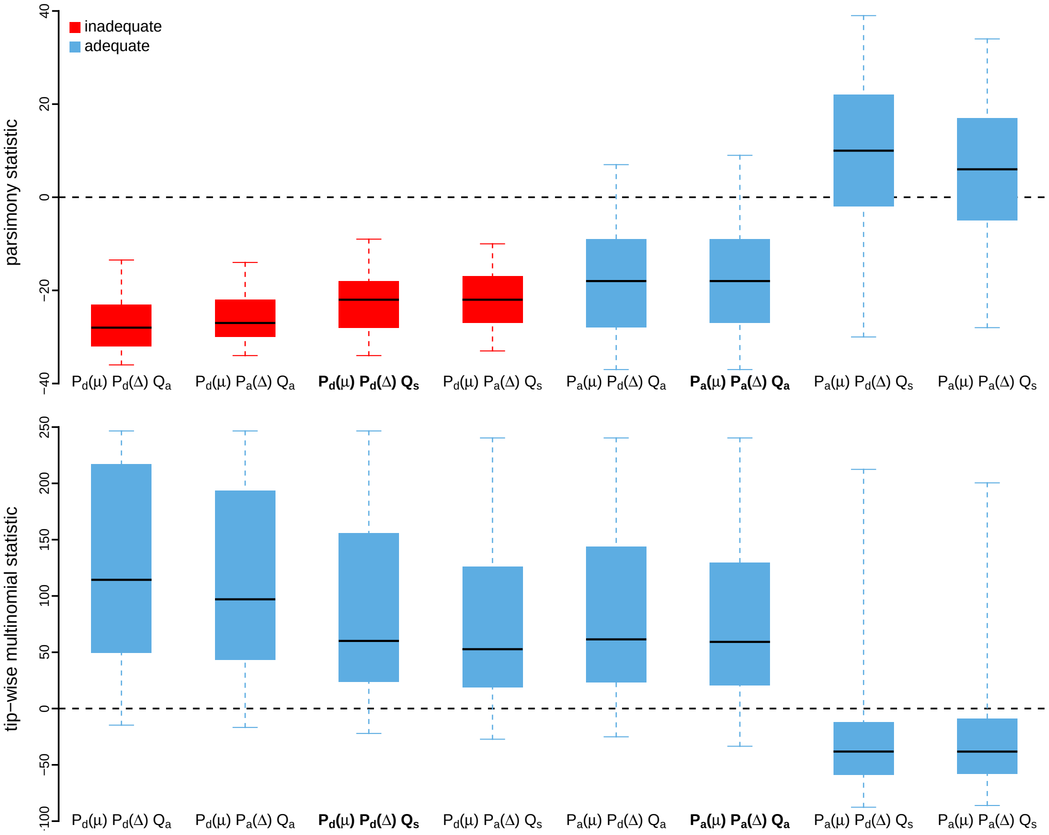


**Figure S41: Posterior-predictive distributions of the parsimony statistic (top panel) and the tip-wise multinomial statistic (bottom panel) under each of the eight prior models for the Deformed wing virus rdrp fragment dataset.** Boxplots depict the posterior-predictive distributions of the statistic under each of the eight candidate prior models: the center of each box is the median predictive value of the summary statistic; the box and whiskers indicate the corresponding 50% and 95% posterior-predictive intervals, respectively. The horizontal dashed line indicates when the simulated and observed datasets produce identical value for the summary statistic. A model is judged to be inadequate (*i.e.*, incapable of generating geographic datasets that are similar to the observed data) if its 95% posterior-predictive interval does not overlap with the dashed line. Here the preferred default prior model is inadequate, whereas the preferred alternative prior model is adequate. Notation for the candidate models is described in Table S8; the preferred default- and alternative-prior models are indicated in bold text.

*The Impact of Prior Choice on Average Dispersal Rate*

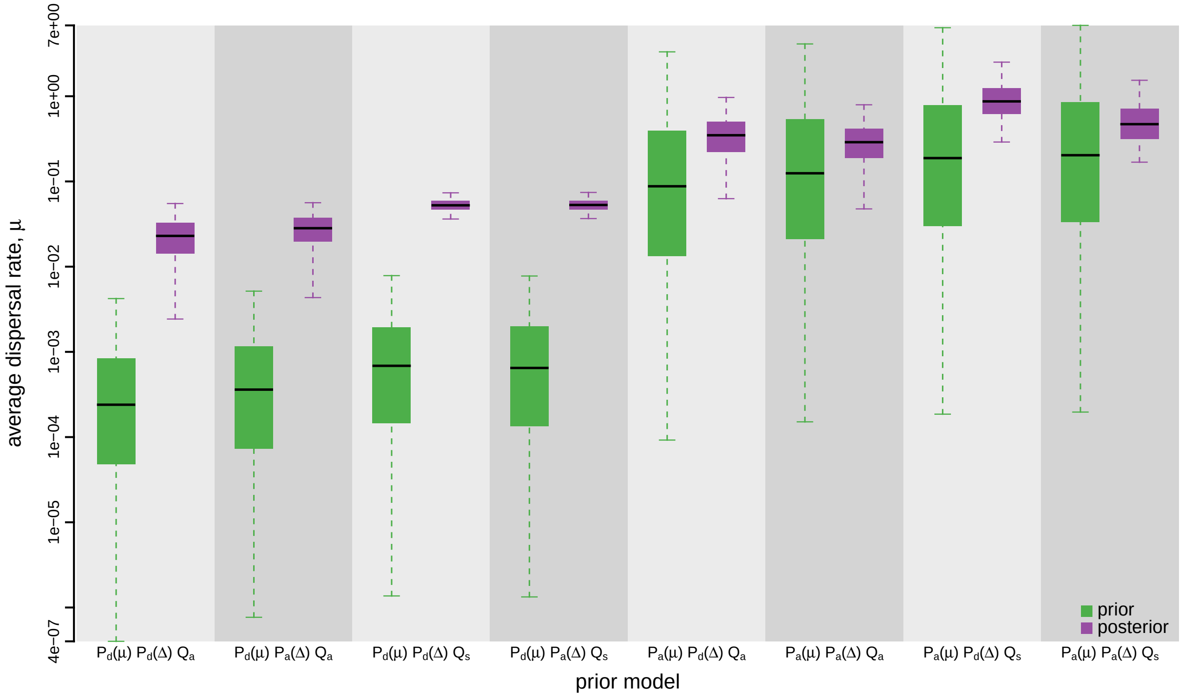


**Figure S42: The impact of prior choice on the average dispersal rate for the Deformed wing virus rdrp fragment dataset.** Each pair of boxplots depicts the specified prior (green) and corresponding posterior estimate (purple) of the average dispersal rate, $\mu$, under each prior model: the center of each box indicates the median rate; the box and whiskers indicate the corresponding 50% and 95% credible intervals, respectively. Notation for the prior models is as described in Table S8.

*The Impact of Prior Choice on Pairwise Dispersal Rates*

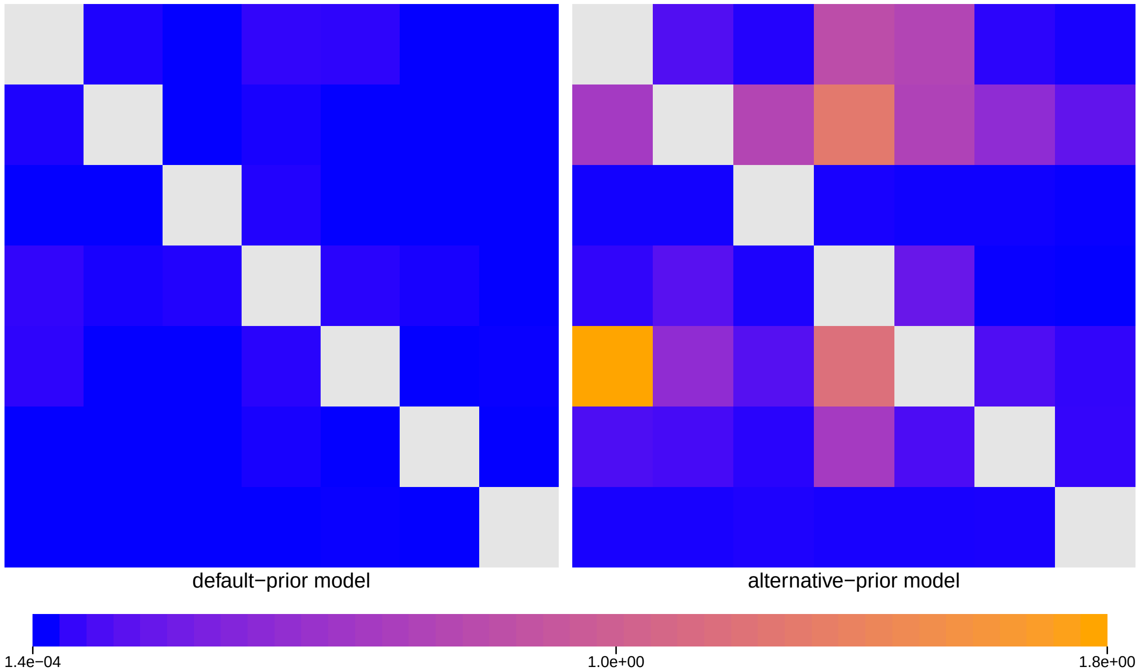


**Figure S43: The impact of prior choice on pairwise dispersal rates for the Deformed wing virus rdrp fragment dataset.** Heatmaps summarize posterior-mean estimates of the instantaneous rate of dispersal between each pair of geographic areas, $q_{ij}$, under the default (left) and alternative (right) prior models.

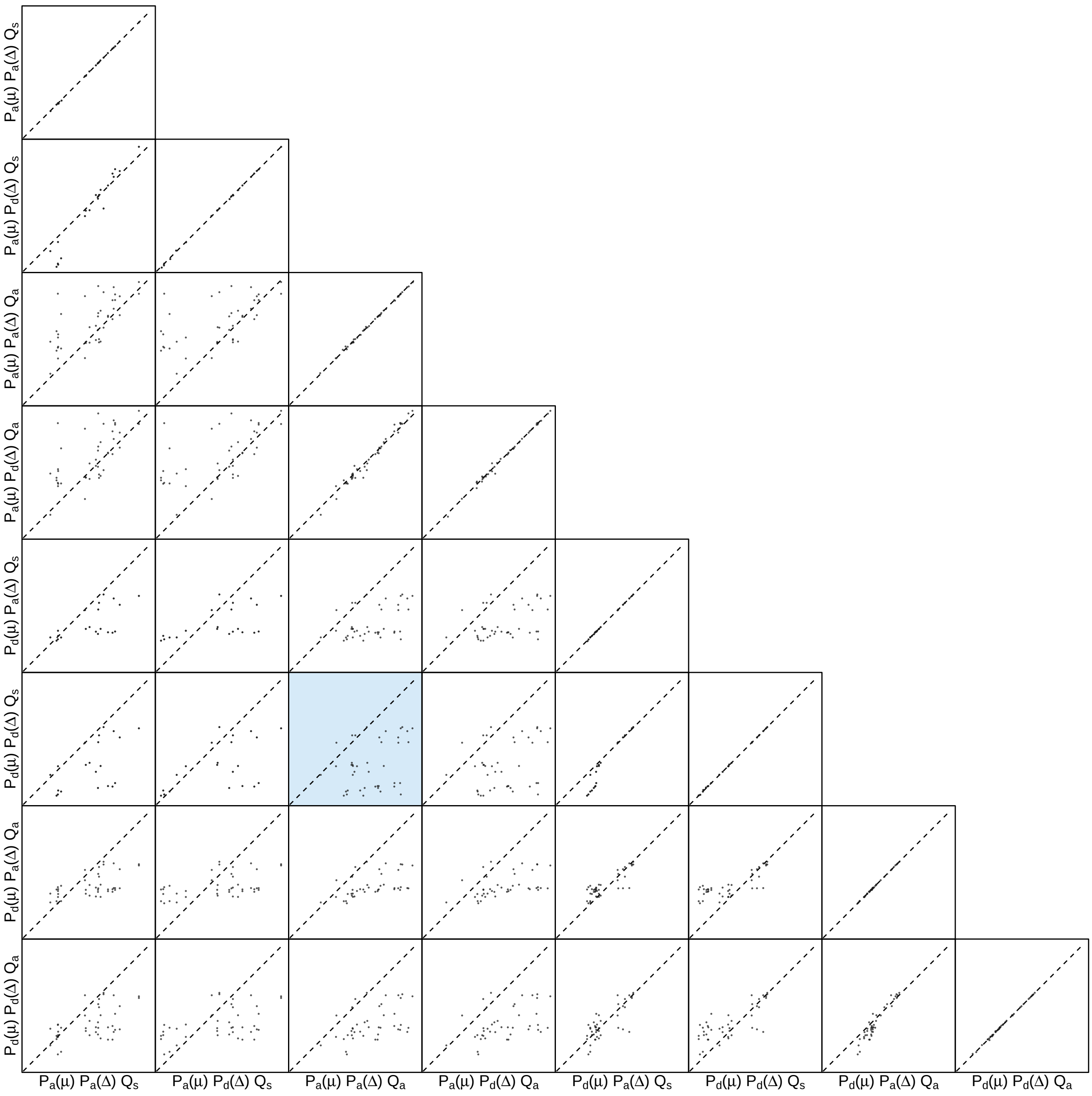


**Figure S44: The impact of prior choice on pairwise dispersal rates for the Deformed wing virus rdrp fragment dataset.** Each cell of the plot compares posterior-mean estimate of the rate of dispersal between each pair of geographic areas, $q_{ij}$, between each pair of prior models. $x$- and $y$-axis of each cell are both on log scale. Diagonal cells are comparisons between half and the other half of the replicates under the same prior model, assessing the convergence of MCMC simulations; off-diagonal cells are comparisons between different prior models, demonstrating the impact of the prior model on the estimates of pairwise dispersal rates. The shaded cell corresponds to the comparison between the preferred default- and the preferred alternative-prior models (*c.f.*, Figure S43). Notation for the prior models is as described in Table S8.

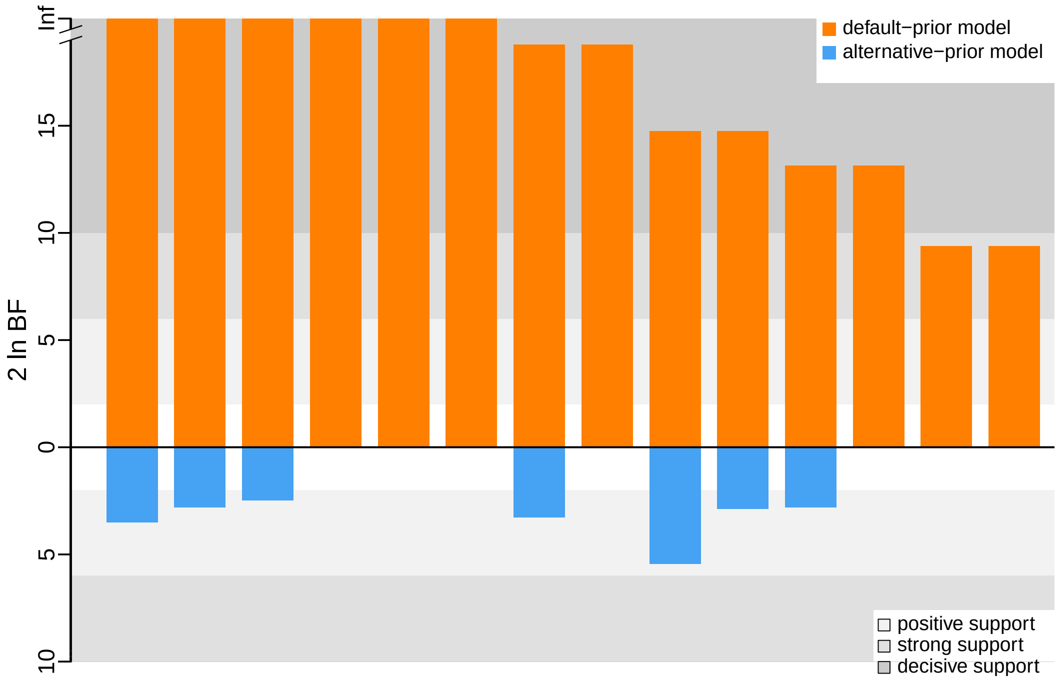


**Figure S45: The impact of prior choice on the inferred support for dispersal routes for the Deformed wing virus rdrp fragment dataset.** We compare the evidential support for each dispersal route for the Deformed wing virus rdrp fragment dataset under the default (orange) and alternative (blue) prior models. Each bar indicates the $2 \ln \text{BF}$ for the corresponding dispersal route between two areas; only supported dispersal routes (*i.e.*, $2 \ln \text{BF} > 2$) are plotted.

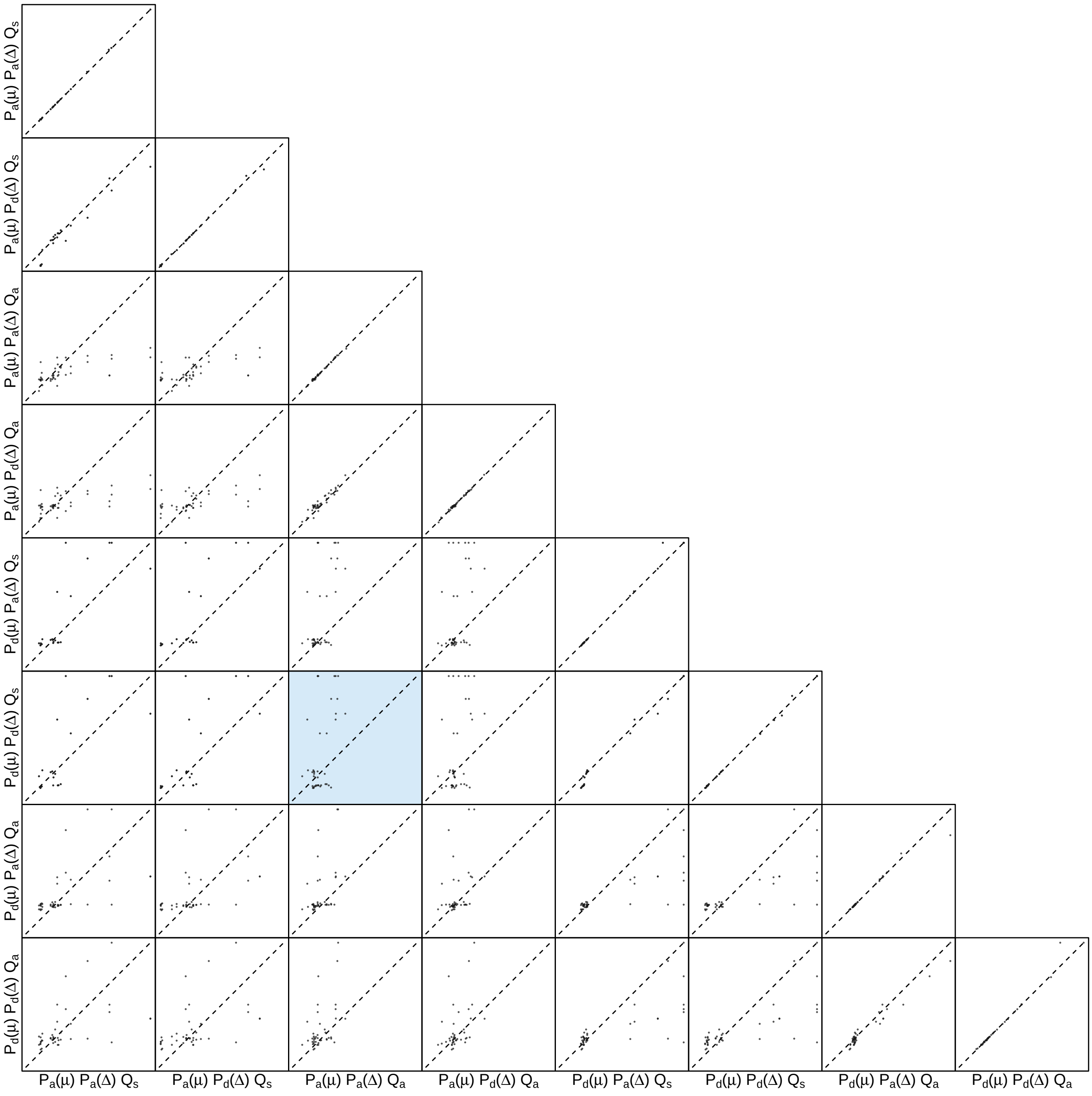


**Figure S46: The impact of prior choice on the inferred support for dispersal routes for the Deformed wing virus rdrp fragment dataset.** Each cell of the plot compares the inferred support (2 ln BF) for pairwise dispersal routes between each pair of prior models. Diagonal cells are comparisons between half and the other half of the replicates under the same prior model, assessing the convergence of MCMC simulations; off-diagonal cells are comparisons between different prior models, demonstrating the impact of the prior model on the inferred support for pairwise dispersal routes. The shaded cell corresponds to the comparison between the preferred default- and the preferred alternative-prior models (*c.f.*, Figure S45). Notation for the prior models is as described in Table S8.

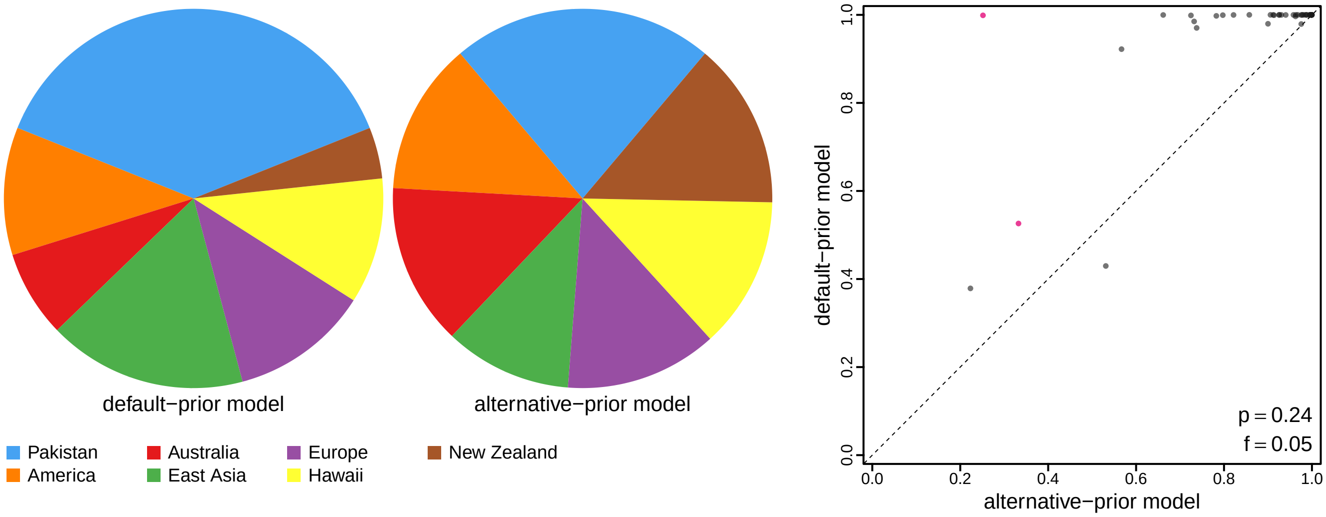


**Figure S47: The impact of prior choice on ancestral-area estimates for the Deformed wing virus rdrp fragment dataset.** The left panel compares the posterior probability of each ancestral area at the root node under the default- and alternative-prior models for the Deformed wing virus rdrp fragment dataset. The right panel plots the posterior probability of the most probable ancestral area under the default-prior model for each node in the MCC tree (y-axis) against the corresponding posterior probability of that area under the alternative-prior model (x-axis). Pink dots represent the internal nodes where the MAP ancestral area inferred under the default-prior model differs from that inferred under the alternative-prior models. The statistic $p$ denotes the fraction of internal nodes that are shared under the default- and alternative-prior models; $f$, is the fraction of shared nodes where the MAP ancestral area differs under the default- and alternative-prior models. Note that the posterior probabilities of the MAP ancestral area under the default-prior model are generally higher than those under the alternative-prior model (*i.e.*, the default-prior model tends to mask uncertainty in the ancestral-area estimates).

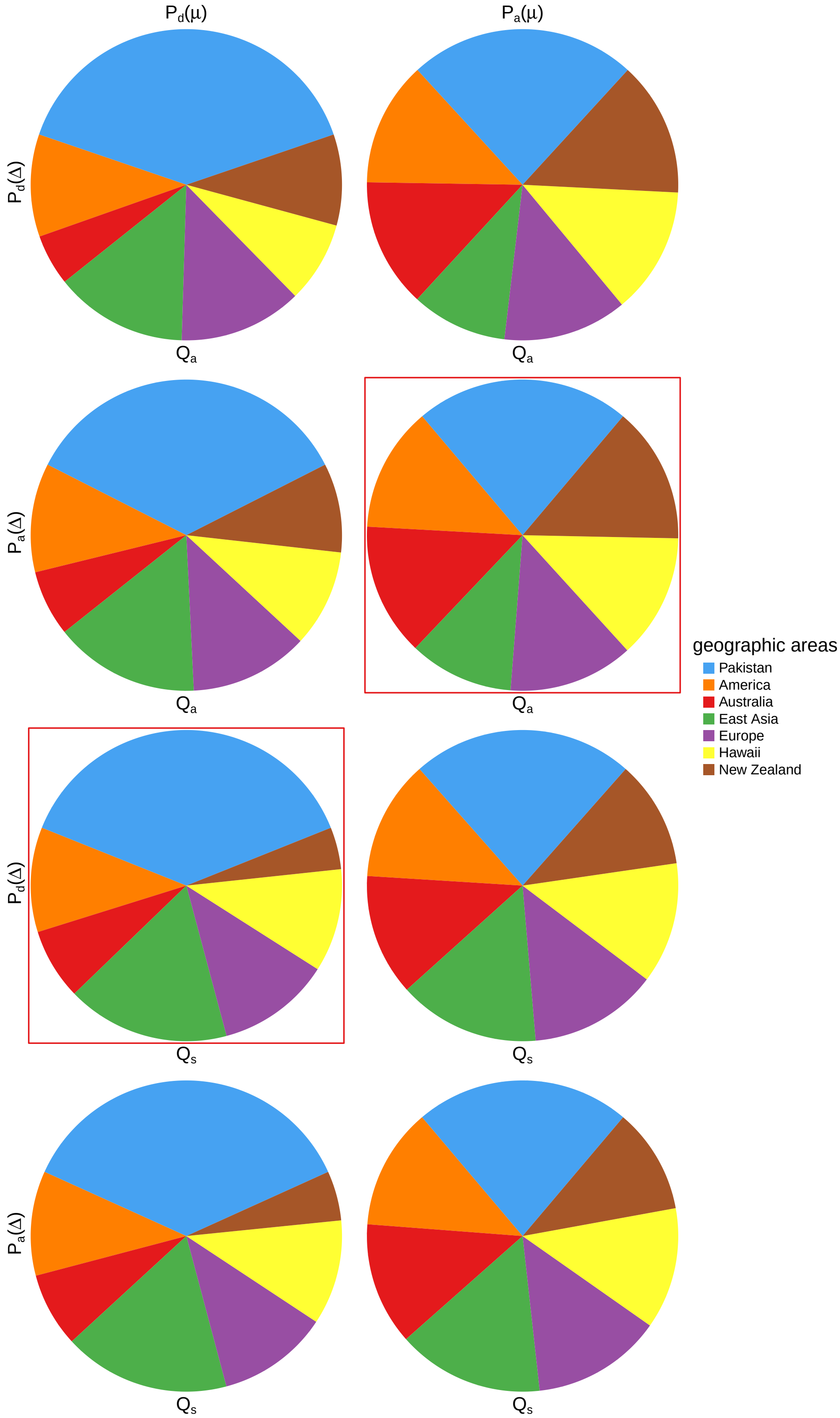


**Figure S48: The impact of prior choice on ancestral-area estimates at the root node for the Deformed wing virus rdrp fragment dataset.** The posterior probability of each ancestral area at the root node under each prior model for the Deformed wing virus rdrp fragment dataset. The pie chart enclosed in red corresponds to the comparison between the preferred default- and the preferred alternative-prior models (*c.f.*, Figure S47, left panel). Model notation follows the description in Table S8.

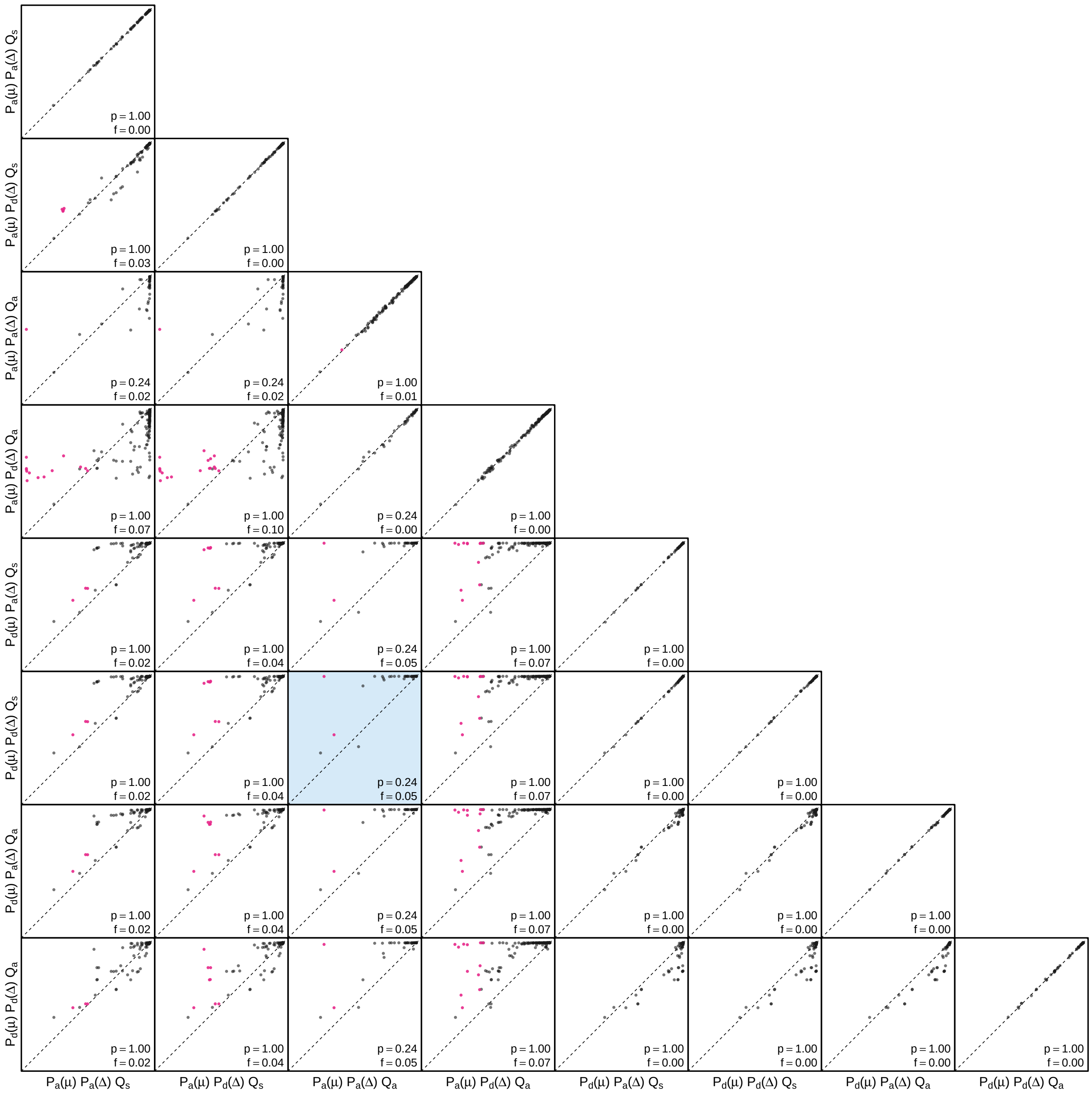


**Figure S49: The impact of prior choice on the MAP ancestral-area estimates at internal nodes for the Deformed wing virus rdrp fragment dataset.** Pairwise scatter plots compare the posterior probability of the MAP ancestral area inferred under each combination of prior models. Diagonal cells are comparisons between two replicates under the same prior model (assessing the convergence of MCMC simulations); off-diagonal cells are comparisons between different prior models, demonstrating the impact of the prior model on both the posterior probability and the identity of the MAP ancestral-area estimates at internal nodes. Pink dots represent the internal nodes where the MAP ancestral area inferred under the default-prior model differs from that inferred under the alternative-prior models. The statistic $p$ denotes the fraction of internal nodes that are shared under the default- and alternative-prior models; $f$ is the fraction of shared nodes where the MAP ancestral area differs under the default- and alternative-prior models. The shaded cell corresponds to the comparison between the preferred default- and the preferred alternative-prior models (*c.f.*, Figure S47, left panel). Model notation follows the description in Table S8.

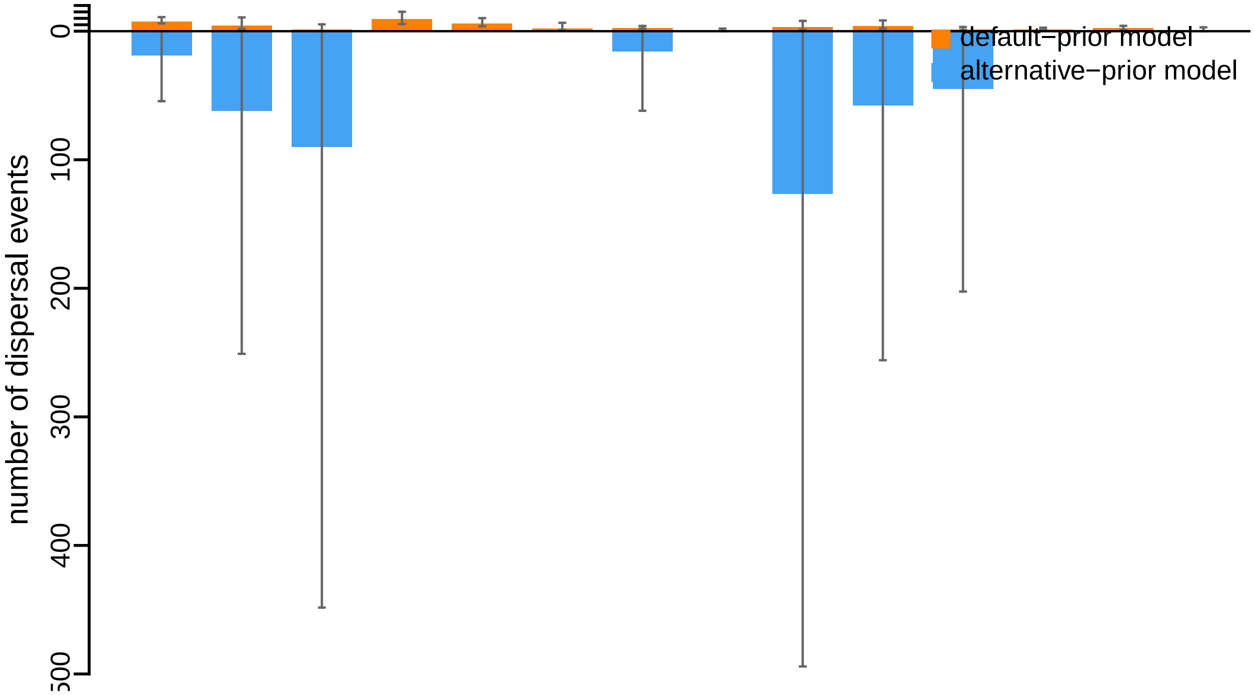


**Figure S50: The impact of prior choice on the inferred number of dispersal events between each pair of areas for the Deformed wing virus rdrp fragment dataset.** The reflected bar plot depicts the number of dispersal events inferred under the default (orange) and alternative (blue) prior models for the Deformed wing virus rdrp fragment dataset. Each bar indicates the posterior-mean number of dispersal events between a pair of areas; whiskers indicate the 95% credible interval. Note that only the number of dispersal events over the "significant" dispersal routes (*i.e.*, $2 \ln \mathrm{BF} > 2$) are figured.

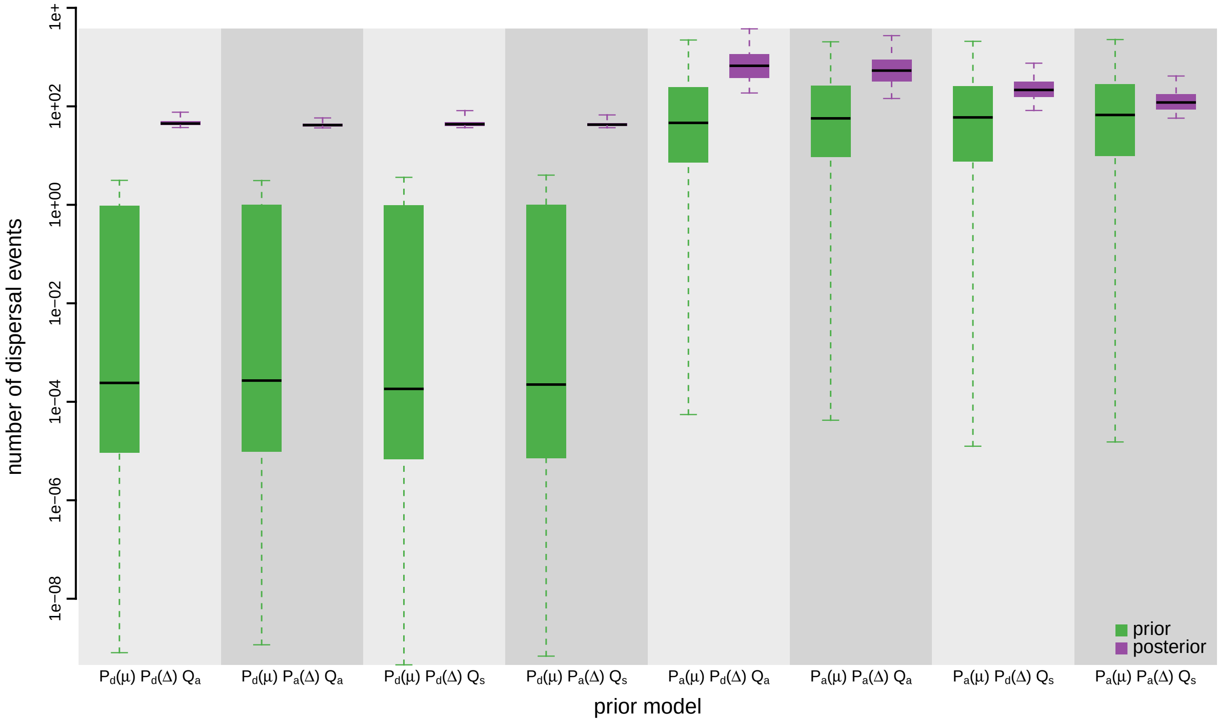


**Figure S51: The impact of prior choice on the inferred total number of dispersal events between all areas for the Deformed wing virus rdrp fragment dataset.** Each pair of boxplots depicts the specified prior (green) and corresponding posterior estimate (purple) of the number of dispersal events between all areas under each prior model: the center of each box indicates the posterior-median number of dispersal events; the box and whiskers indicate the corresponding 50% and 95% credible intervals, respectively. Model notation follows the description in Table S8.

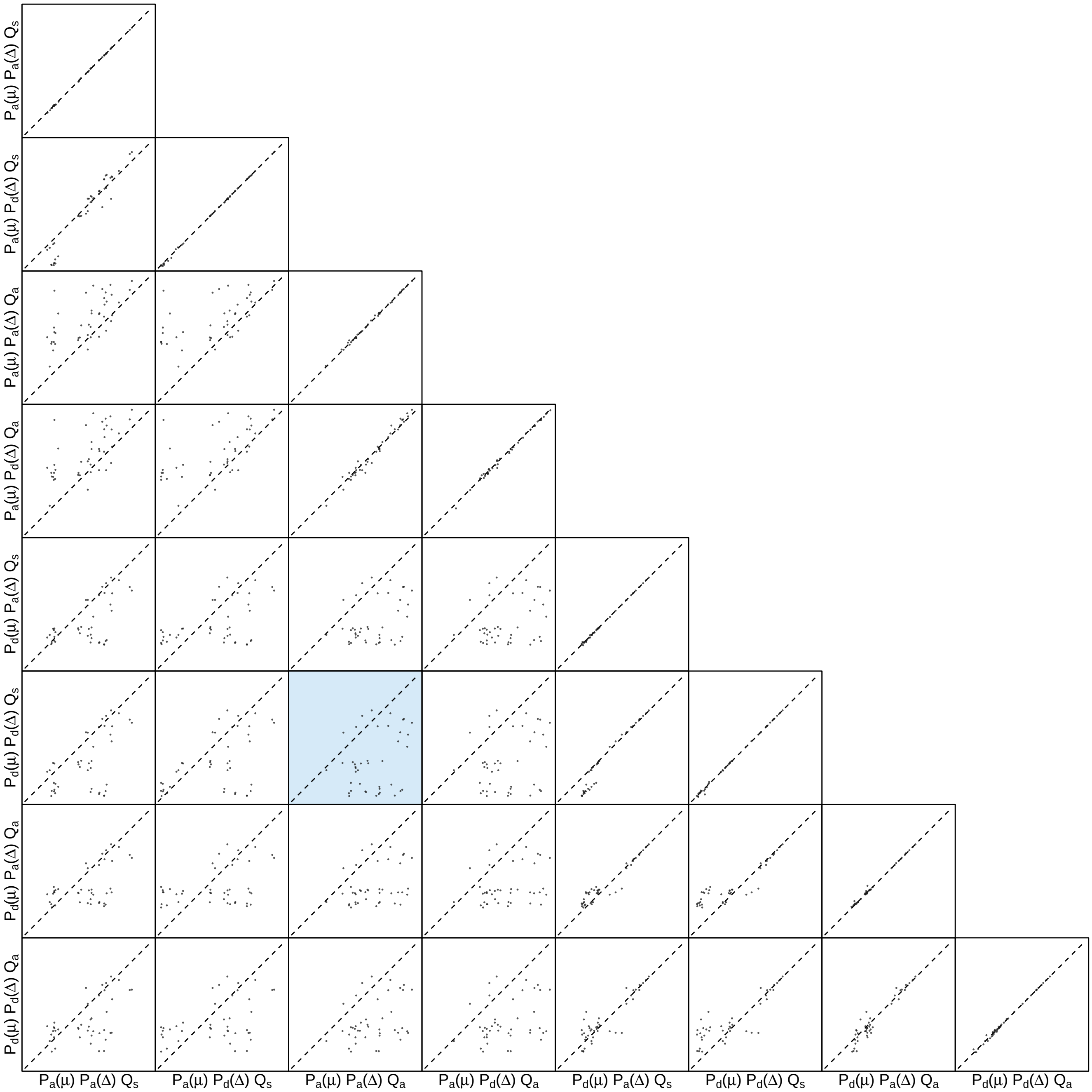


**Figure S52: The impact of prior choice on the inferred number of dispersal events between each pair of areas for the Deformed wing virus rdrp fragment dataset.** Each cell of the plot compares the inferred number of pairwise dispersal events between each pair of prior models. *x*- and *y*-axis of each cell are both on log scale. Diagonal cells are comparisons between half and the other half of the replicates under the same prior model, assessing the convergence of MCMC simulations; off-diagonal cells are comparisons between different prior models, demonstrating the impact of the prior model on the inferred number of pairwise dispersal events. The shaded cell corresponds to the comparison between the preferred default- and the preferred alternative-prior models (*c.f.*, Figure S50). Model notation follows the description in Table S8.

##### S3.3.2.3 Vp3 fragment

*The Impact of Prior Choice on Biogeographic Model Fit*

**Table S9: Marginal-likelihood estimates of each of the eight prior models for the Deformed wing virus vp3 fragment dataset.** Columns 2–5 list marginal likelihoods inferred from four replicate analyses. Columns 6–7 list the mean and standard deviation of these marginal-likelihood estimates. The last column lists the marginal-likelihood estimates computed by combining the samples from the replicate power-posterior MCMC simulations. Candidate models are listed in rows and include all possible combinations of: (1) instantaneous-rate matrices (symmetric, $Q_s$ or asymmetric, $Q_a$); (2) priors on the average dispersal rate [default, $P_d(\mu)$ or alternative, $P_a(\mu)$], and; (3) priors on the number of dispersal routes [default, $P_d(\Delta)$ or alternative, $P_a(\Delta)$]. The preferred default- and alternative-prior models are indicated in bold text.

| Model | replicate1 | replicate2 | replicate3 | replicate4 | mean | sd | combined |
|---|---|---|---|---|---|---|---|
| $\boldsymbol{P_d(\mu)Q_aP_d(\Delta)}$ | **-106.16** | **-106.21** | **-106.20** | **-106.23** | **-106.20** | **0.03** | **-106.20** |
| $P_d(\mu)Q_aP_a(\Delta)$ | -104.06 | -104.02 | -104.05 | -103.98 | -104.03 | 0.04 | -104.03 |
| $P_d(\mu)Q_sP_d(\Delta)$ | -106.43 | -106.45 | -106.25 | -106.35 | -106.37 | 0.09 | -106.31 |
| $P_d(\mu)Q_sP_a(\Delta)$ | -104.08 | -104.15 | -103.95 | -104.11 | -104.07 | 0.09 | -104.07 |
| $P_a(\mu)Q_aP_d(\Delta)$ | -92.46 | -92.33 | -92.45 | -92.42 | -92.41 | 0.06 | -92.41 |
| $P_a(\mu)Q_aP_a(\Delta)$ | -91.99 | -91.94 | -91.95 | -91.96 | -91.96 | 0.02 | -91.96 |
| $P_a(\mu)Q_sP_d(\Delta)$ | -93.00 | -92.86 | -92.76 | -92.72 | -92.84 | 0.12 | -92.80 |
| $\boldsymbol{P_a(\mu)Q_sP_a(\Delta)}$ | **-91.56** | **-91.30** | **-91.53** | **-91.51** | **-91.47** | **0.12** | **-91.49** |

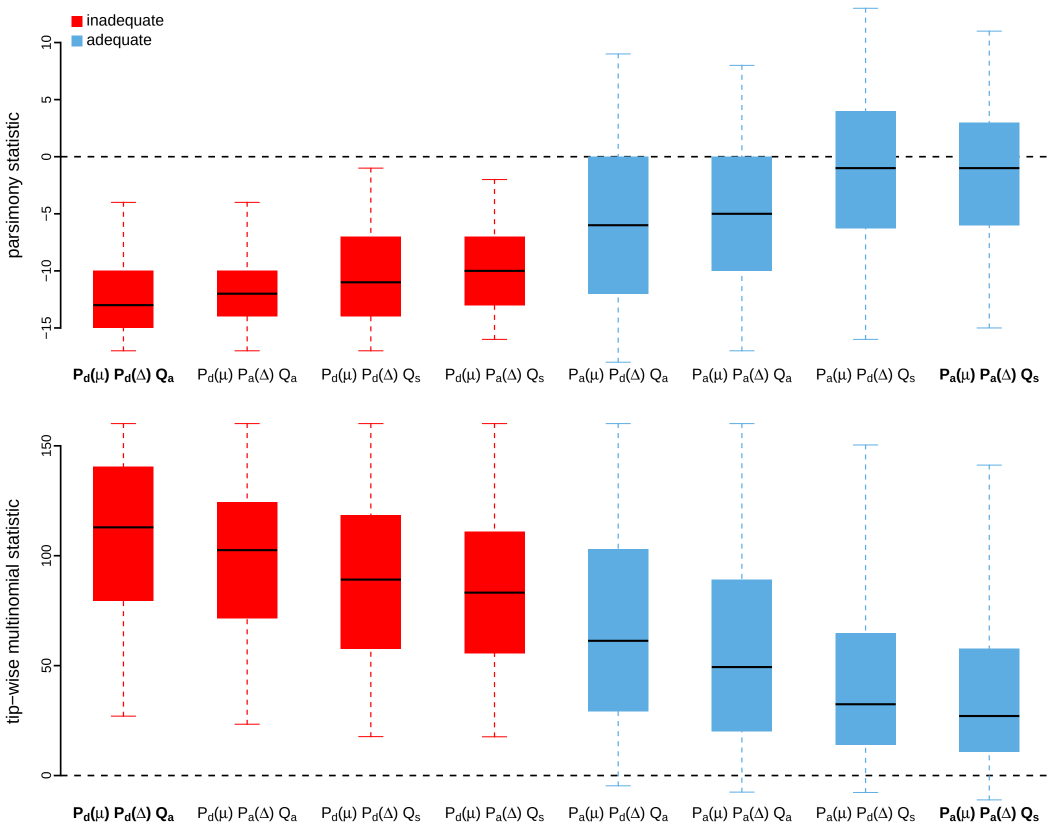


**Figure S53: Posterior-predictive distributions of the parsimony statistic (top panel) and the tip-wise multinomial statistic (bottom panel) under each of the eight prior models for the Deformed wing virus vp3 fragment dataset.** Boxplots depict the posterior-predictive distributions of the statistic under each of the eight candidate prior models: the center of each box is the median predictive value of the summary statistic; the box and whiskers indicate the corresponding 50% and 95% posterior-predictive intervals, respectively. The horizontal dashed line indicates when the simulated and observed datasets produce identical value for the summary statistic. A model is judged to be inadequate (*i.e.*, incapable of generating geographic datasets that are similar to the observed data) if its 95% posterior-predictive interval does not overlap with the dashed line. Here the preferred default prior model is inadequate, whereas the preferred alternative prior model is adequate. Notation for the candidate models is described in Table S9; the preferred default- and alternative-prior models are indicated in bold text.

*The Impact of Prior Choice on Average Dispersal Rate*

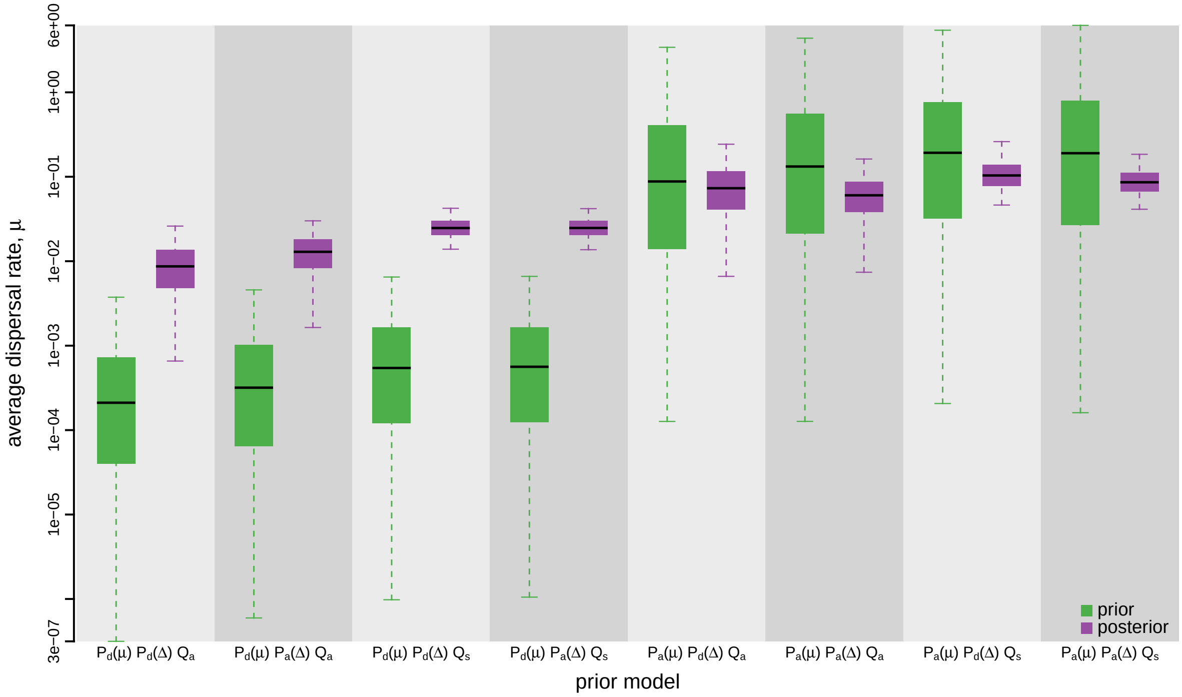


**Figure S54: The impact of prior choice on the average dispersal rate for the Deformed wing virus vp3 fragment dataset.** Each pair of boxplots depicts the specified prior (green) and corresponding posterior estimate (purple) of the average dispersal rate, $\mu$, under each prior model: the center of each box indicates the median rate; the box and whiskers indicate the corresponding 50% and 95% credible intervals, respectively. Notation for the prior models is as described in Table S9.

*The Impact of Prior Choice on Pairwise Dispersal Rates*

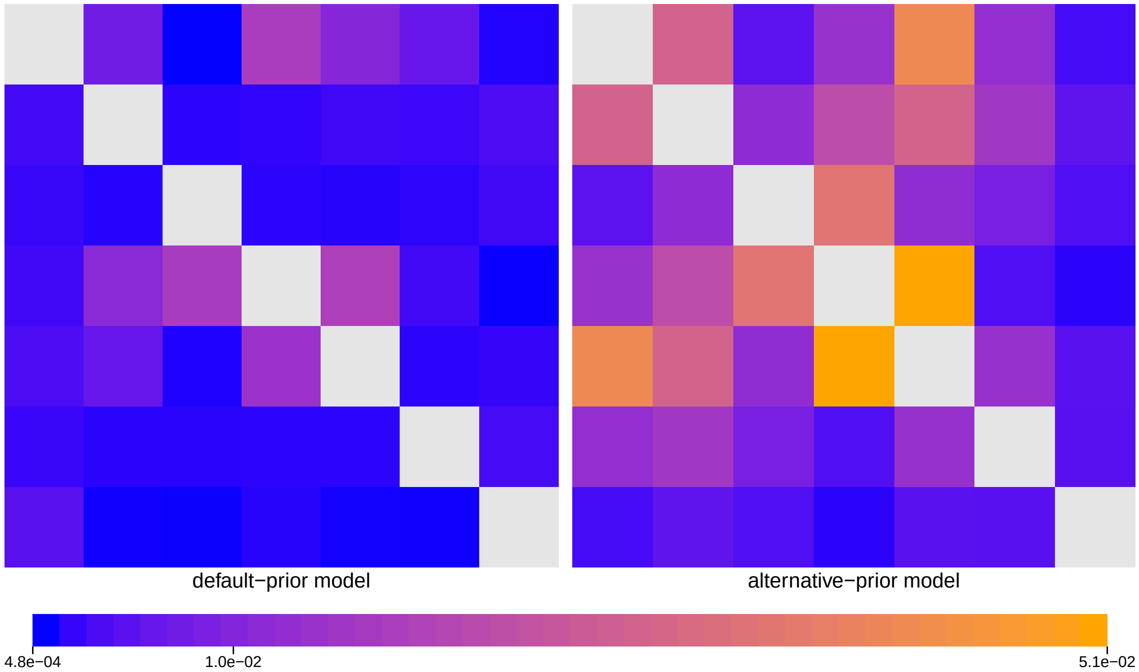


**Figure S55: The impact of prior choice on pairwise dispersal rates for the Deformed wing virus vp3 fragment dataset.** Heatmaps summarize posterior-mean estimates of the instantaneous rate of dispersal between each pair of geographic areas, $q_{ij}$, under the default (left) and alternative (right) prior models.

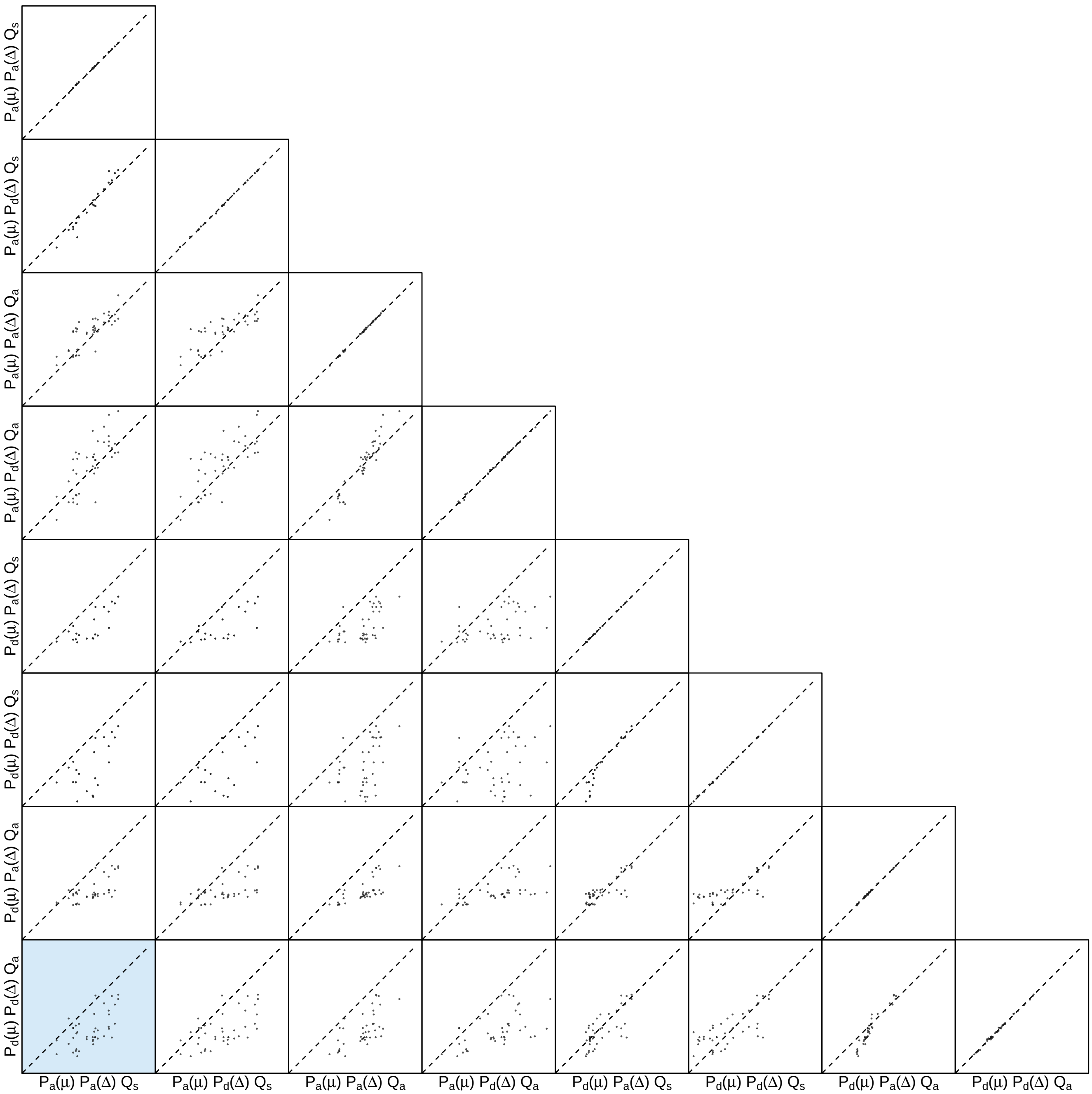


**Figure S56: The impact of prior choice on pairwise dispersal rates for the Deformed wing virus vp3 fragment dataset.** Each cell of the plot compares posterior-mean estimate of the rate of dispersal between each pair of geographic areas, $q_{ij}$, between each pair of prior models. $x$- and $y$-axis of each cell are both on log scale. Diagonal cells are comparisons between half and the other half of the replicates under the same prior model, assessing the convergence of MCMC simulations; off-diagonal cells are comparisons between different prior models, demonstrating the impact of the prior model on the estimates of pairwise dispersal rates. The shaded cell corresponds to the comparison between the preferred default- and the preferred alternative-prior models (*c.f.*, Figure S55). Notation for the prior models is as described in Table S9.

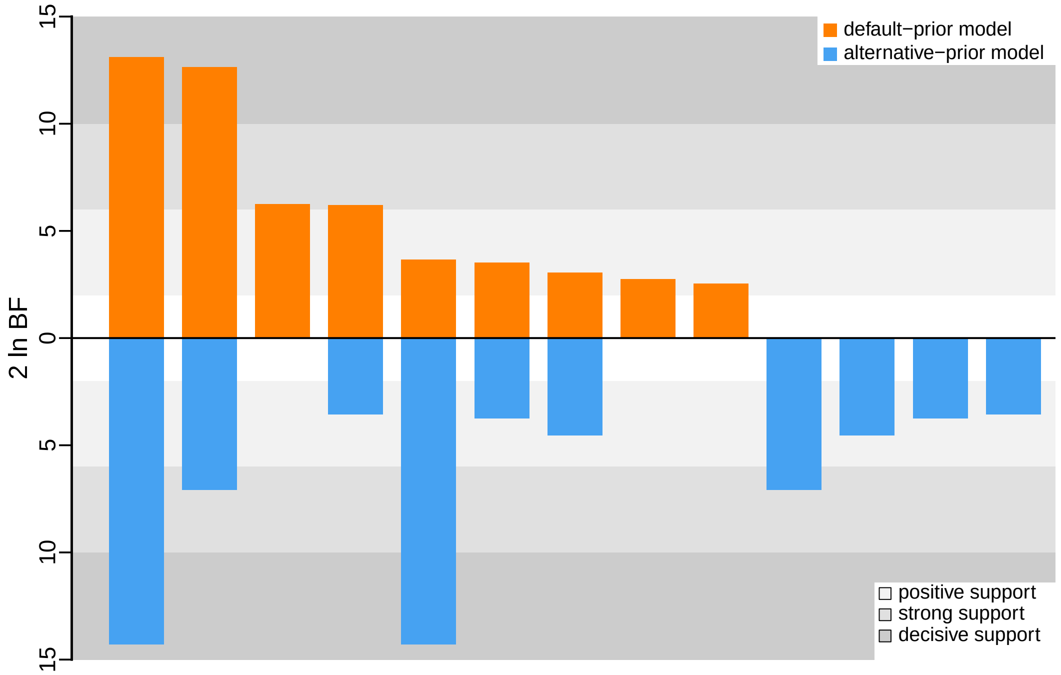


**Figure S57: The impact of prior choice on the inferred support for dispersal routes for the Deformed wing virus vp3 fragment dataset.** We compare the evidential support for each dispersal route for the Deformed wing virus vp3 fragment dataset under the default (orange) and alternative (blue) prior models. Each bar indicates the $2 \ln \text{BF}$ for the corresponding dispersal route between two areas; only supported dispersal routes (*i.e.*, $2 \ln \text{BF} > 2$) are plotted.

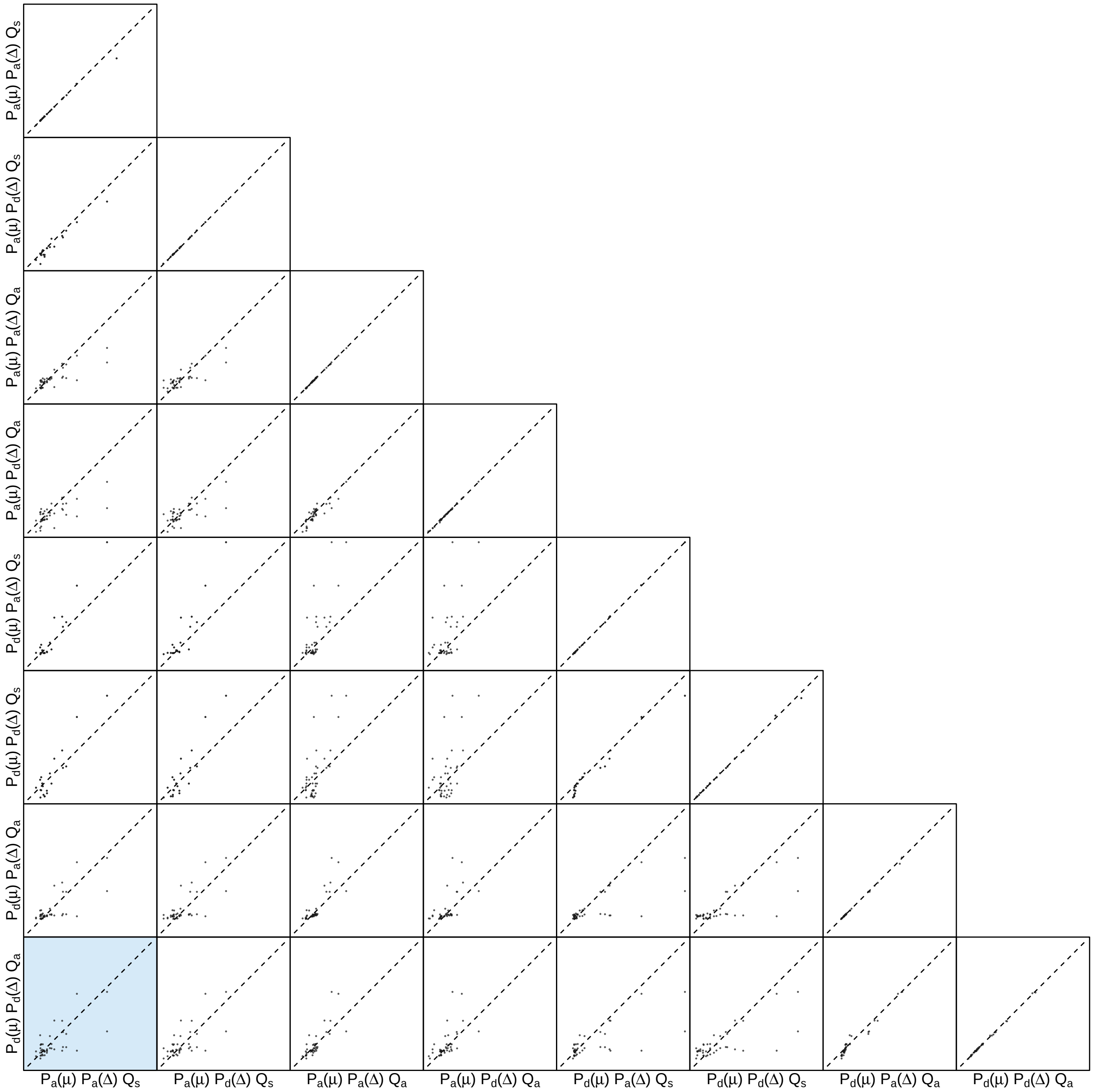


**Figure S58: The impact of prior choice on the inferred support for dispersal routes for the Deformed wing virus vp3 fragment dataset.** Each cell of the plot compares the inferred support (2 ln BF) for pairwise dispersal routes between each pair of prior models. Diagonal cells are comparisons between half and the other half of the replicates under the same prior model, assessing the convergence of MCMC simulations; off-diagonal cells are comparisons between different prior models, demonstrating the impact of the prior model on the inferred support for pairwise dispersal routes. The shaded cell corresponds to the comparison between the preferred default- and the preferred alternative-prior models (*c.f.*, Figure S57). Notation for the prior models is as described in Table S9.

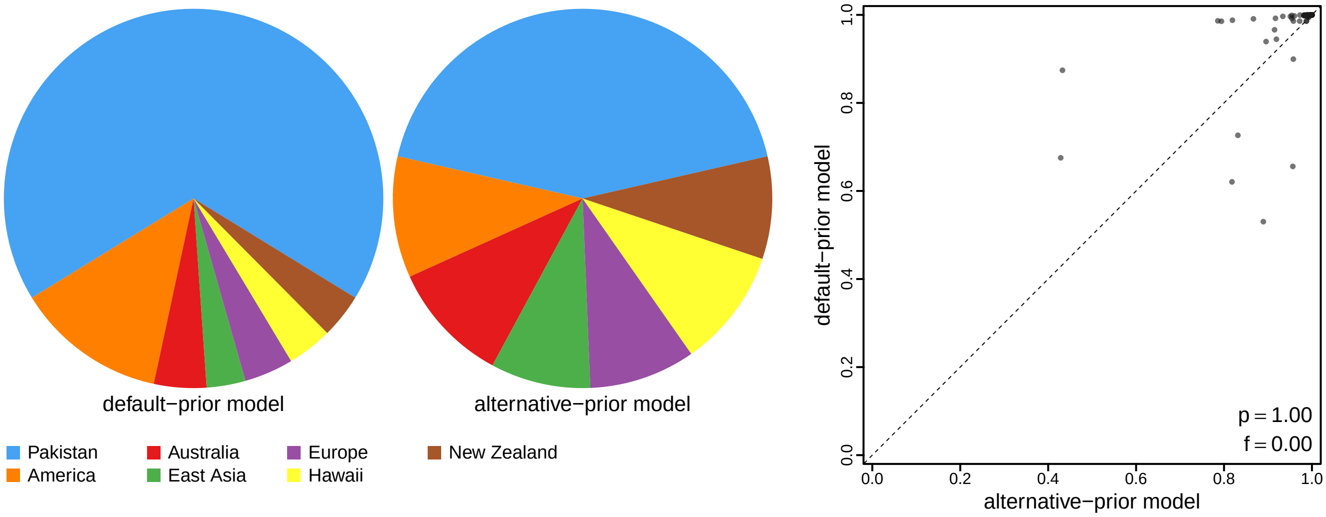


**Figure S59: The impact of prior choice on ancestral-area estimates for the Deformed wing virus vp3 fragment dataset.** The left panel compares the posterior probability of each ancestral area at the root node under the default- and alternative-prior models for the Deformed wing virus vp3 fragment dataset. The right panel plots the posterior probability of the most probable ancestral area under the default-prior model for each node in the MCC tree (y-axis) against the corresponding posterior probability of that area under the alternative-prior model (x-axis). Pink dots represent the internal nodes where the MAP ancestral area inferred under the default-prior model differs from that inferred under the alternative-prior models. The statistic $p$ denotes the fraction of internal nodes that are shared under the default- and alternative-prior models; $f$, is the fraction of shared nodes where the MAP ancestral area differs under the default- and alternative-prior models. Note that the posterior probabilities of the MAP ancestral area under the default-prior model are generally higher than those under the alternative-prior model (*i.e.*, the default-prior model tends to mask uncertainty in the ancestral-area estimates).

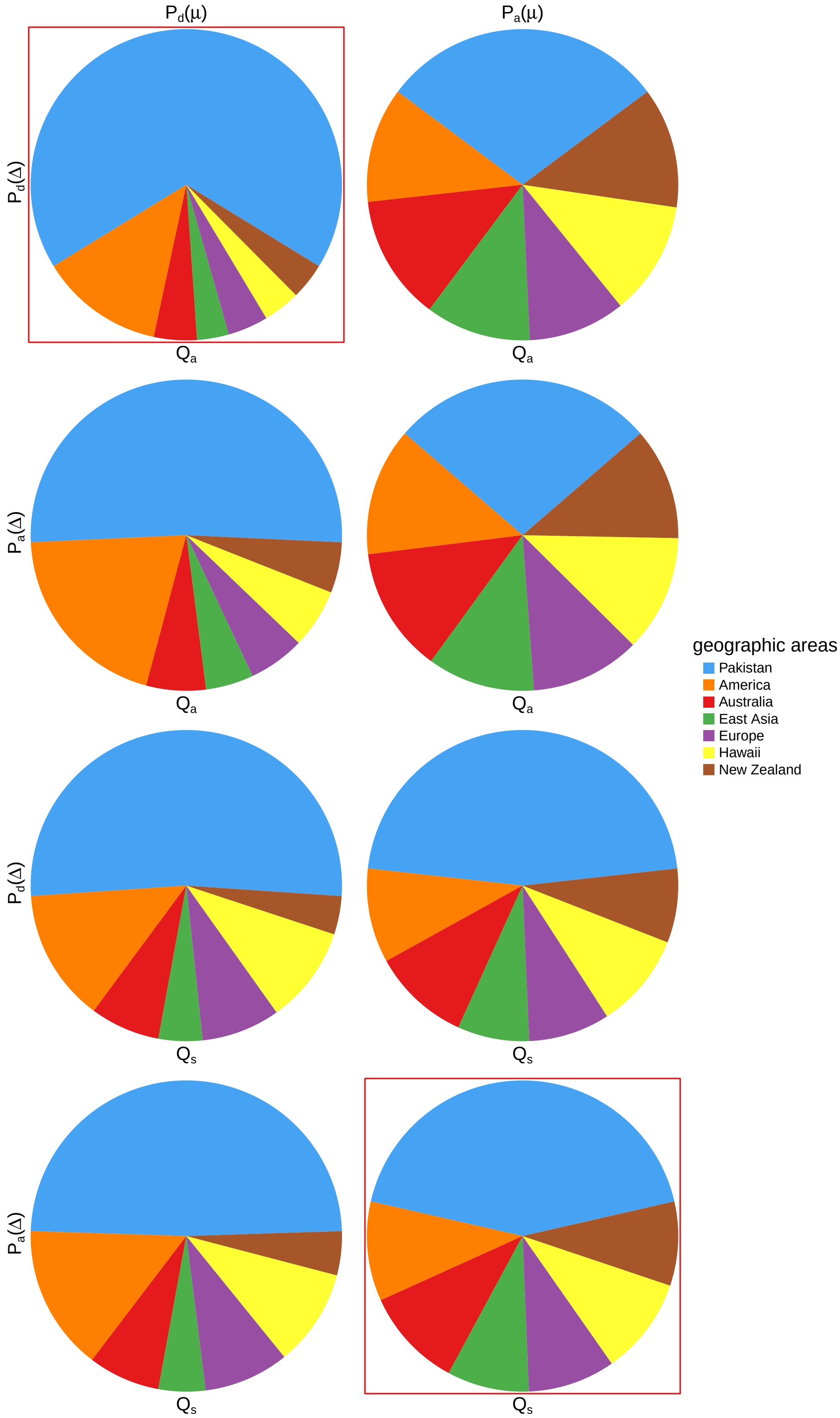


**Figure S60: The impact of prior choice on ancestral-area estimates at the root node for the Deformed wing virus vp3 fragment dataset.** The posterior probability of each ancestral area at the root node under each prior model for the Deformed wing virus vp3 fragment dataset. The pie chart enclosed in red corresponds to the comparison between the preferred default- and the preferred alternative-prior models (*c.f.*, Figure S59, left panel). Model notation follows the description in Table S9.

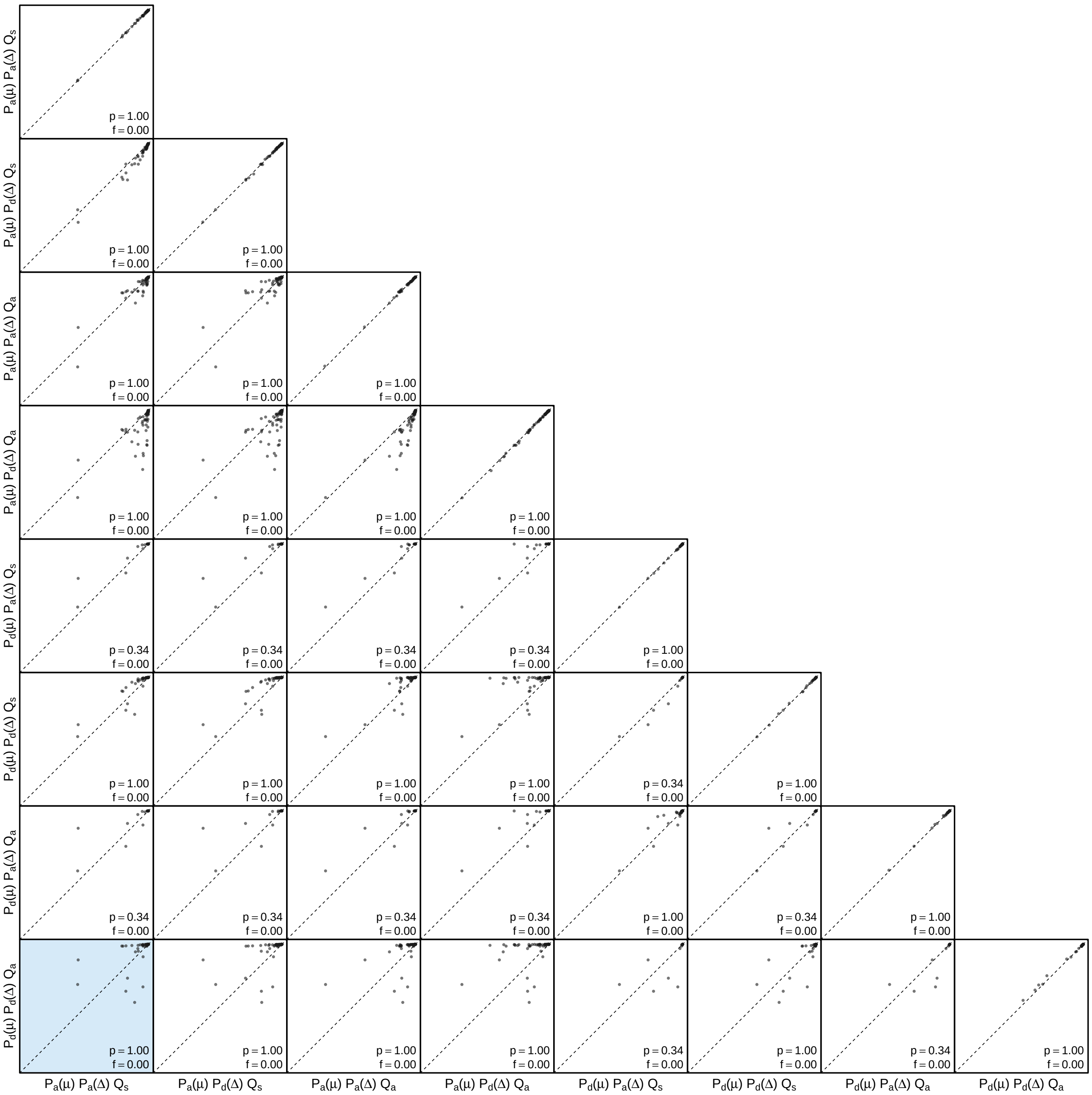


**Figure S61: The impact of prior choice on the MAP ancestral-area estimates at internal nodes for the Deformed wing virus vp3 fragment dataset.** Pairwise scatter plots compare the posterior probability of the MAP ancestral area inferred under each combination of prior models. Diagonal cells are comparisons between two replicates under the same prior model (assessing the convergence of MCMC simulations); off-diagonal cells are comparisons between different prior models, demonstrating the impact of the prior model on both the posterior probability and the identity of the MAP ancestral-area estimates at internal nodes. Pink dots represent the internal nodes where the MAP ancestral area inferred under the default-prior model differs from that inferred under the alternative-prior models. The statistic $p$ denotes the fraction of internal nodes that are shared under the default- and alternative-prior models; $f$ is the fraction of shared nodes where the MAP ancestral area differs under the default- and alternative-prior models. The shaded cell corresponds to the comparison between the preferred default- and the preferred alternative-prior models (*c.f.*, Figure S59, left panel). Model notation follows the description in Table S9.

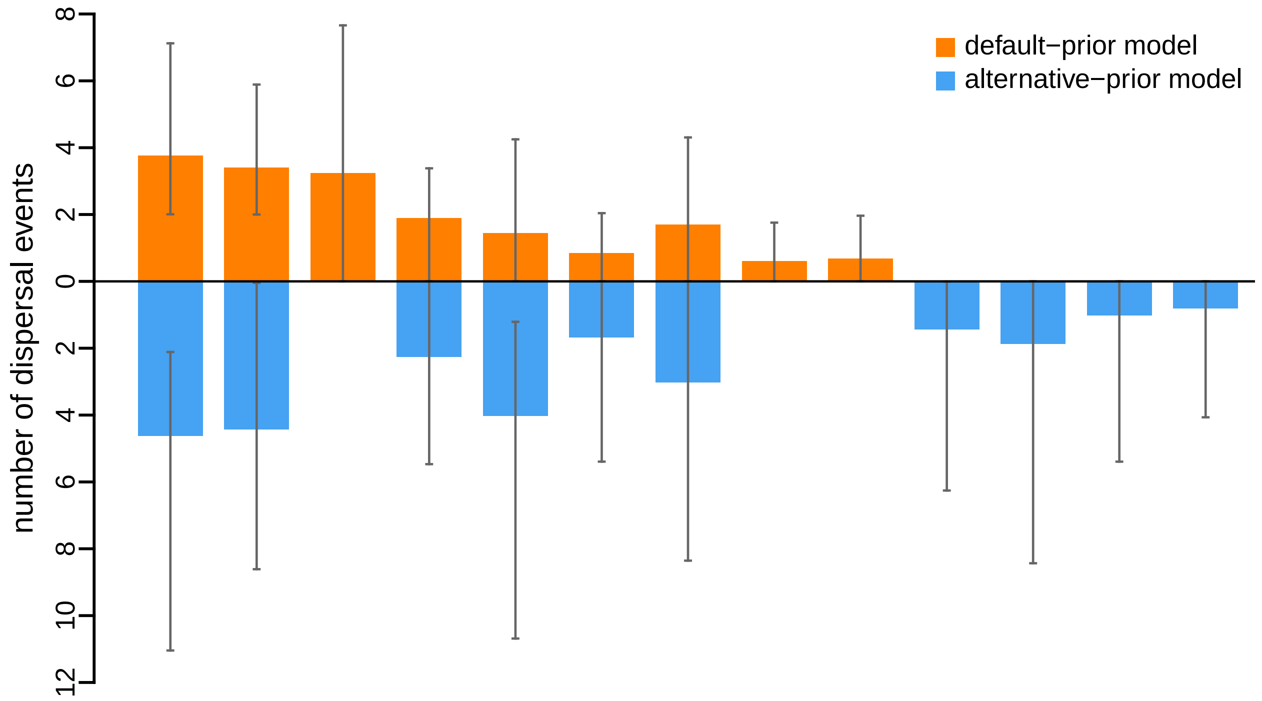


**Figure S62: The impact of prior choice on the inferred number of dispersal events between each pair of areas for the Deformed wing virus vp3 fragment dataset.** The reflected bar plot depicts the number of dispersal events inferred under the default (orange) and alternative (blue) prior models for the Deformed wing virus vp3 fragment dataset. Each bar indicates the posterior-mean number of dispersal events between a pair of areas; whiskers indicate the 95% credible interval. Note that only the number of dispersal events over the "significant" dispersal routes (*i.e.*, $2 \ln \mathrm{BF} > 2$) are figured.

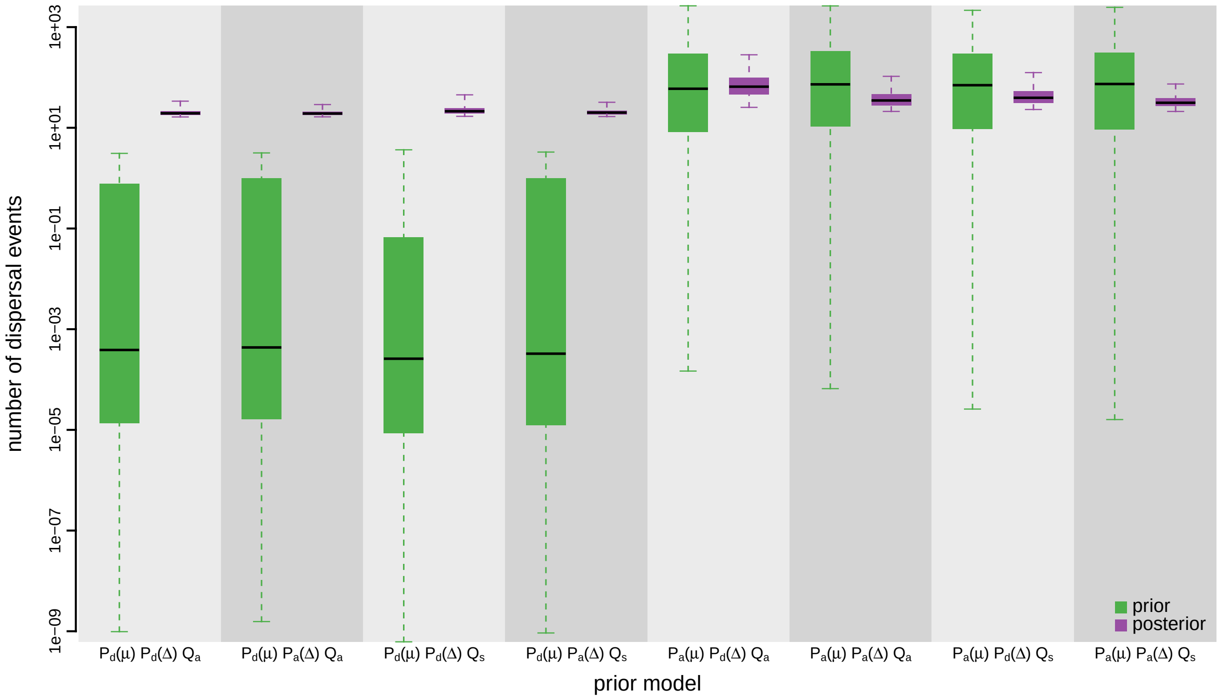


**Figure S63: The impact of prior choice on the inferred total number of dispersal events between all areas for the Deformed wing virus vp3 fragment dataset.** Each pair of boxplots depicts the specified prior (green) and corresponding posterior estimate (purple) of the number of dispersal events between all areas under each prior model: the center of each box indicates the posterior-median number of dispersal events; the box and whiskers indicate the corresponding 50% and 95% credible intervals, respectively. Model notation follows the description in Table S9.

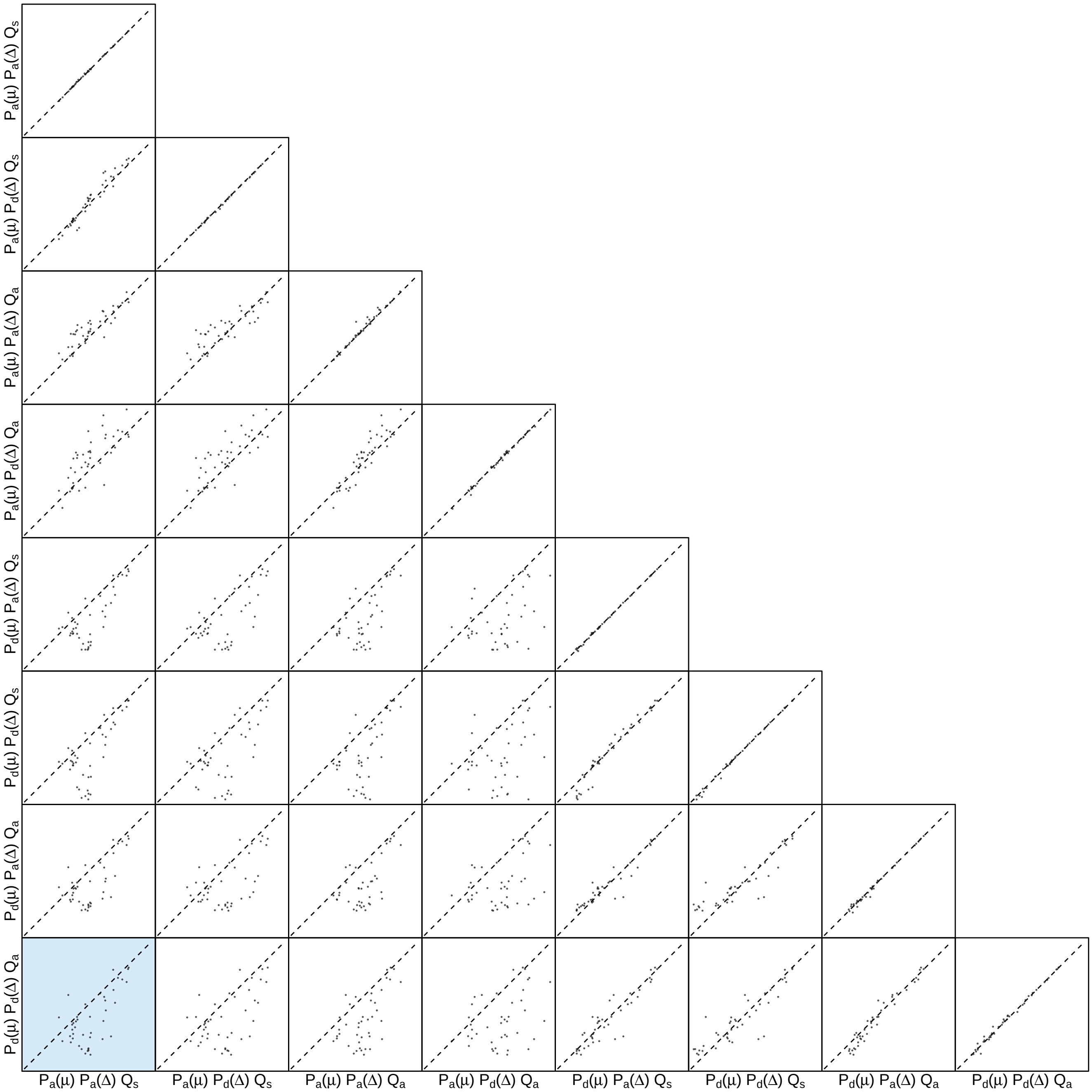


**Figure S64: The impact of prior choice on the inferred number of dispersal events between each pair of areas for the Deformed wing virus vp3 fragment dataset.** Each cell of the plot compares the inferred number of pairwise dispersal events between each pair of prior models. *x*- and *y*-axis of each cell are both on log scale. Diagonal cells are comparisons between half and the other half of the replicates under the same prior model, assessing the convergence of MCMC simulations; off-diagonal cells are comparisons between different prior models, demonstrating the impact of the prior model on the inferred number of pairwise dispersal events. The shaded cell corresponds to the comparison between the preferred default- and the preferred alternative-prior models (*c.f.*, Figure S62). Model notation follows the description in Table S9.

###### S3.3.3 HIV

Faria et al. (2014) explored the origin and early spread of HIV-1 in human populations by analyzing sequences collected from central and southeast Africa and America between the 1980s and early 2000s. The authors used a down-sampling scheme of the complete dataset to verify the robustness of the main conclusions in the original study: this involved the creation of four data(sub)sets. Specifically, Dataset A includes 792 envelope C2V3 sequences collected between 1985–2004 from eight cities in the Democratic Republic of the Congo and the Republic of the Congo. Dataset B includes 927 sequences, with the addition of 67 subtype C sequences from southeast Africa (Zambia, Botswana, Tanzania, Kenya, Uganda, Burundi, Ethiopia and South Africa) sampled between 1986–2005, 67 sequences from the Americas (Haiti, Trinidad and Tobago and the USA) sampled between 1978–1997, and the ZR59 isolate obtained in 1959 from blood collected in Kinshasa. Dataset C includes 466 sequences that were down-sampled from Dataset A; Dataset D includes 601 sequences that were down-sampled from Dataset B; this down-sampling was motivated to decrease the representation of sequences sampled from Kinshasa (see (Faria et al. 2014) for details).

We acquired the sampling geographic location data of Datasets A, B, and C from the BEAST XML scripts provided by the original study; this sampling-area data are available in our GitHub and Dryad repositories.

The posterior distribution of phylogenies (used to perform sequential analyses in Faria et al. 2014) was obtained directly from the Dryad repository of the original study (also available in our GitHub and Dryad repositories).

We reanalyzed Datasets A, B, and C. The MCMC simulations used in the second step of our sequential analyses and of the analyses used to estimate marginal likelihoods under each prior model are described above in Section S3.1. Details of these analyses are available in the XML scripts included in our GitHub and Dryad repositories.

###### S3.3.3.1 Dataset A

*The Impact of Prior Choice on Biogeographic Model Fit*

**Table S10: Marginal-likelihood estimates of each of the eight prior models for HIV dataset A.** Columns 2–5 list marginal likelihoods inferred from four replicate analyses. Columns 6–7 list the mean and standard deviation of these marginal-likelihood estimates. The last column lists the marginal-likelihood estimates computed by combining the samples from the replicate power-posterior MCMC simulations. Candidate models are listed in rows and include all possible combinations of: (1) instantaneous-rate matrices (symmetric, $Q_s$ or asymmetric, $Q_a$); (2) priors on the average dispersal rate [default, $P_d(\mu)$ or alternative, $P_a(\mu)$], and; (3) priors on the number of dispersal routes [default, $P_d(\Delta)$ or alternative, $P_a(\Delta)$]. The preferred default- and alternative-prior models are indicated in bold text.

| Model | replicate1 | replicate2 | replicate3 | replicate4 | mean | sd | combined |
|---|---|---|---|---|---|---|---|
| $P_d(\mu)Q_aP_d(\Delta)$ | -1177.56 | -1177.33 | -1177.40 | -1177.66 | -1177.49 | 0.15 | -1177.48 |
| $P_d(\mu)Q_aP_a(\Delta)$ | -1176.89 | -1176.77 | -1176.84 | -1176.78 | -1176.82 | 0.05 | -1176.81 |
| $\mathbf{P_d(\mu)Q_sP_d(\Delta)}$ | **-1176.34** | **-1176.65** | **-1176.07** | **-1176.43** | **-1176.37** | **0.24** | **-1176.36** |
| $P_d(\mu)Q_sP_a(\Delta)$ | -1175.19 | -1175.21 | -1175.62 | -1175.43 | -1175.36 | 0.20 | -1175.44 |
| $P_a(\mu)Q_aP_d(\Delta)$ | -1040.35 | -1040.39 | -1040.31 | -1040.32 | -1040.34 | 0.04 | -1040.34 |
| $\mathbf{P_a(\mu)Q_aP_a(\Delta)}$ | **-1037.46** | **-1037.5** | **-1037.48** | **-1037.56** | **-1037.50** | **0.04** | **-1037.49** |
| $P_a(\mu)Q_sP_d(\Delta)$ | -1055.82 | -1055.68 | -1055.75 | -1055.83 | -1055.77 | 0.07 | -1055.76 |
| $P_a(\mu)Q_sP_a(\Delta)$ | -1050.73 | -1050.38 | -1050.69 | -1050.79 | -1050.65 | 0.19 | -1050.67 |

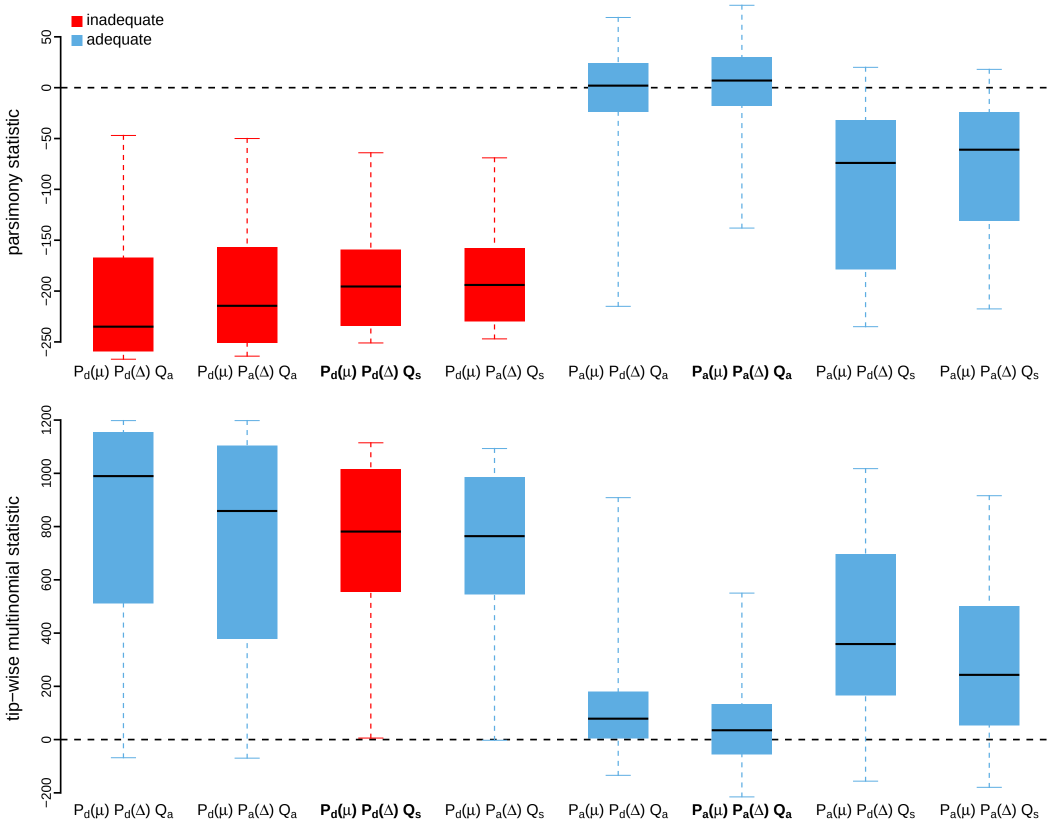


**Figure S65: Posterior-predictive distributions of the parsimony statistic (top panel) and the tip-wise multinomial statistic (bottom panel) under each of the eight prior models for HIV dataset A.** Boxplots depict the posterior-predictive distributions of the statistic under each of the eight candidate prior models: the center of each box is the median predictive value of the summary statistic; the box and whiskers indicate the corresponding 50% and 95% posterior-predictive intervals, respectively. The horizontal dashed line indicates when the simulated and observed datasets produce identical value for the summary statistic. A model is judged to be inadequate (*i.e.*, incapable of generating geographic datasets that are similar to the observed data) if its 95% posterior-predictive interval does not overlap with the dashed line. Here the preferred default prior model is inadequate, whereas the preferred alternative prior model is adequate. Notation for the candidate models is described in Table S12; the preferred default- and alternative-prior models are indicated in bold text.

*The Impact of Prior Choice on Average Dispersal Rate*

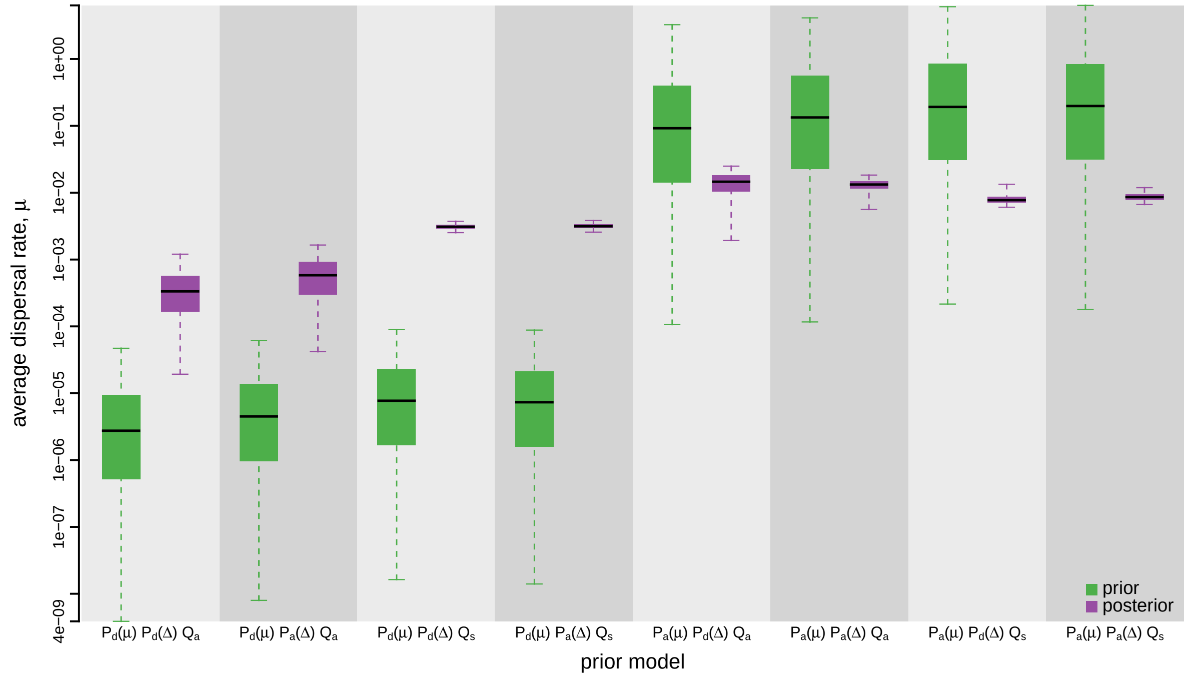


**Figure S66: The impact of prior choice on the average dispersal rate for HIV dataset A.** Each pair of boxplots depicts the specified prior (green) and corresponding posterior estimate (purple) of the average dispersal rate, $\mu$, under each prior model: the center of each box indicates the median rate; the box and whiskers indicate the corresponding 50% and 95% credible intervals, respectively. Notation for the prior models is as described in Table S10.

*The Impact of Prior Choice on Pairwise Dispersal Rates*

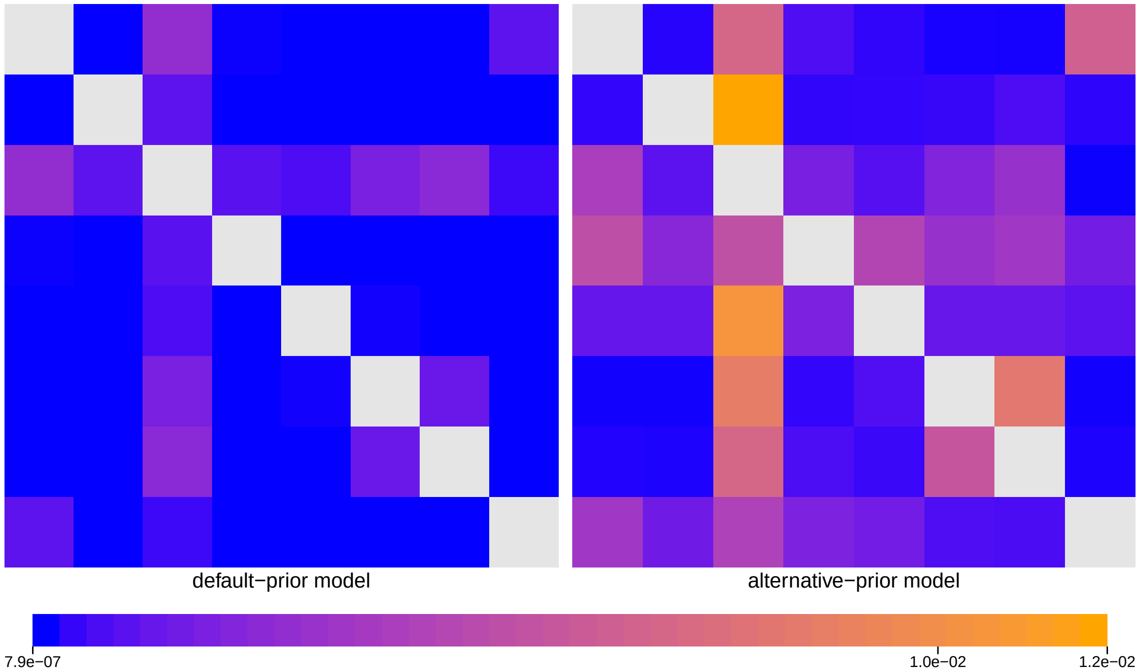


**Figure S67: The impact of prior choice on pairwise dispersal rates for HIV dataset A.** Heatmaps summarize posterior-mean estimates of the instantaneous rate of dispersal between each pair of geographic areas, $q_{ij}$, under the default (left) and alternative (right) prior models.

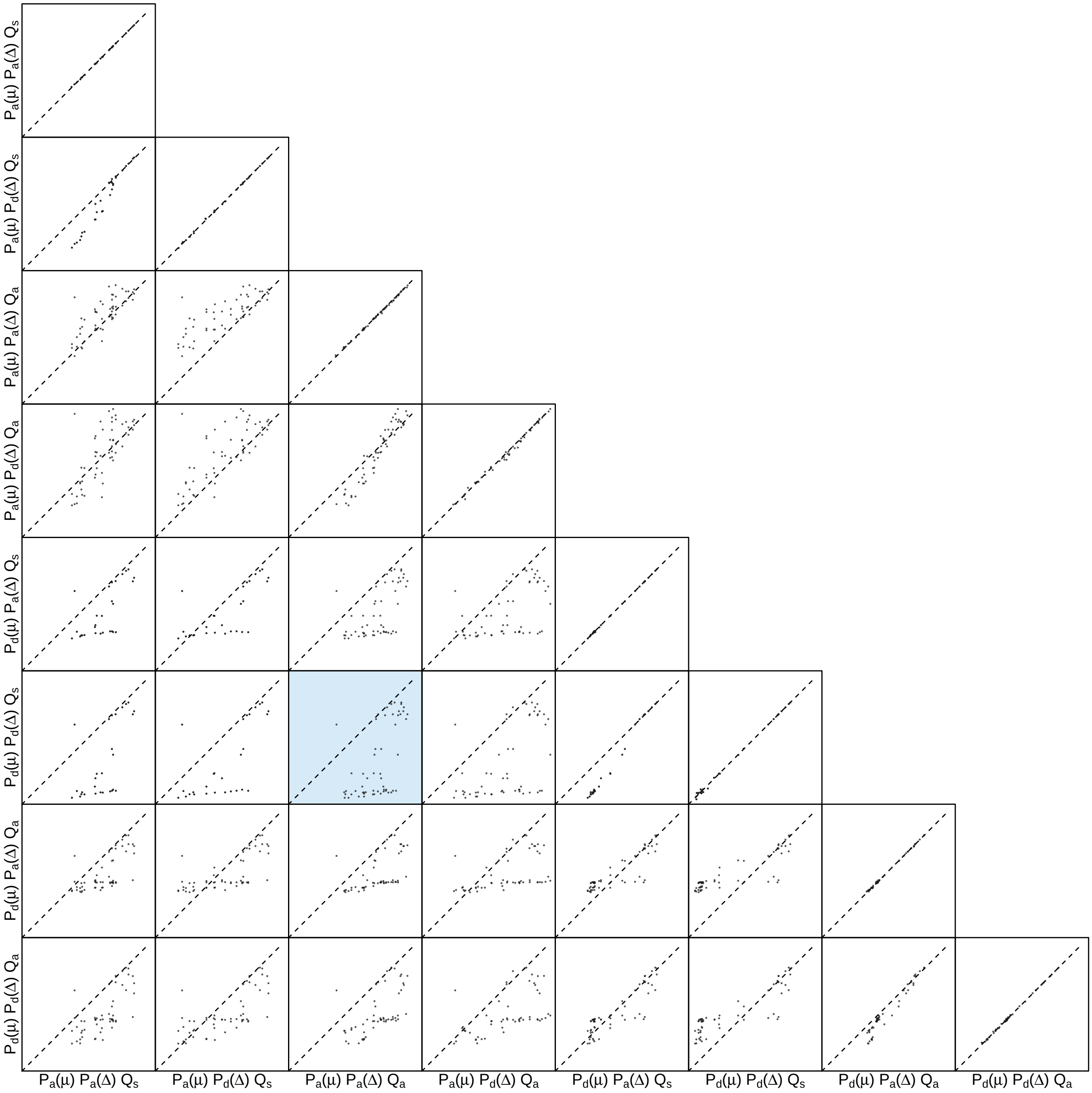


**Figure S68: The impact of prior choice on pairwise dispersal rates for HIV dataset A.** Each cell of the plot compares posterior-mean estimate of the rate of dispersal between each pair of geographic areas, $q_{ij}$, between each pair of prior models. $x$- and $y$-axis of each cell are both on log scale. Diagonal cells are comparisons between half and the other half of the replicates under the same prior model, assessing the convergence of MCMC simulations; off-diagonal cells are comparisons between different prior models, demonstrating the impact of the prior model on the estimates of pairwise dispersal rates. The shaded cell corresponds to the comparison between the preferred default- and the preferred alternative-prior models (*c.f.*, Figure S67). Notation for the prior models is as described in Table S10.

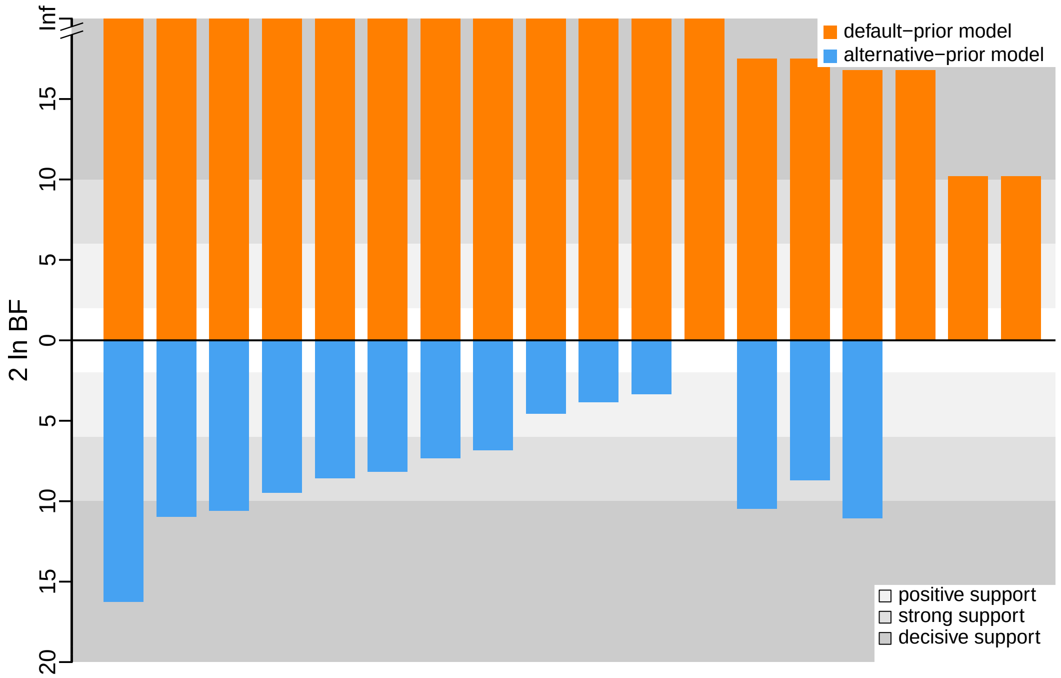


**Figure S69: The impact of prior choice on the inferred support for dispersal routes for HIV dataset A.** We compare the evidential support for each dispersal route for HIV dataset A under the default (orange) and alternative (blue) prior models. Each bar indicates the $2 \ln \mathrm{BF}$ for the corresponding dispersal route between two areas; only supported dispersal routes (*i.e.*, $2 \ln \mathrm{BF} > 2$) are plotted.

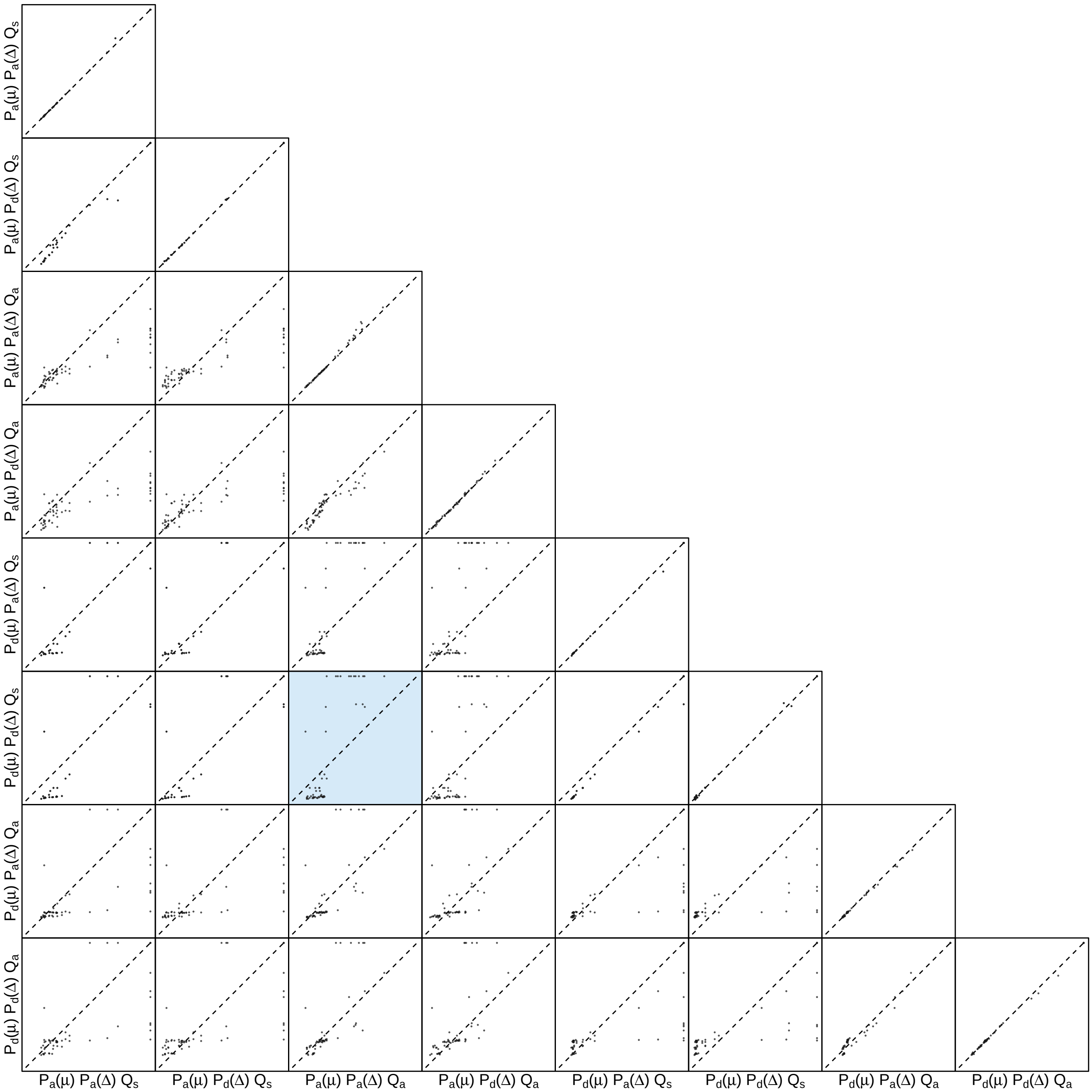


**Figure S70: The impact of prior choice on the inferred support for dispersal routes for HIV dataset A.** Each cell of the plot compares the inferred support (2 ln BF) for pairwise dispersal routes between each pair of prior models. Diagonal cells are comparisons between half and the other half of the replicates under the same prior model, assessing the convergence of MCMC simulations; off-diagonal cells are comparisons between different prior models, demonstrating the impact of the prior model on the inferred support for pairwise dispersal routes. The shaded cell corresponds to the comparison between the preferred default- and the preferred alternative-prior models (*c.f.*, Figure S69). Notation for the prior models is as described in Table S10.

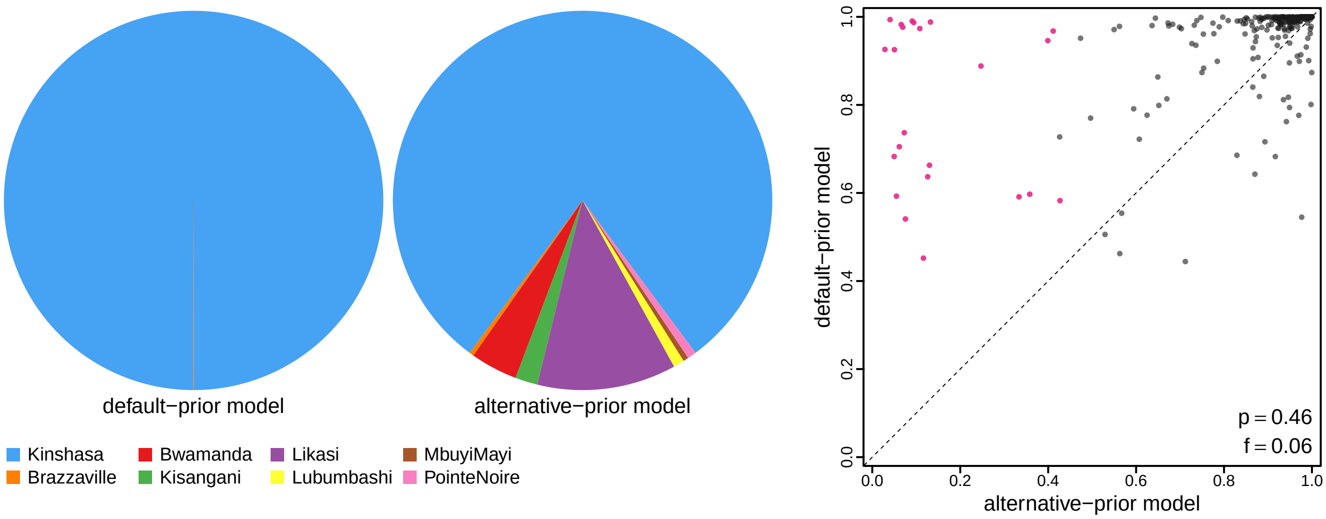


**Figure S71: The impact of prior choice on ancestral-area estimates for HIV dataset A.** The left panel compares the posterior probability of each ancestral area at the root node under the default- and alternative-prior models for HIV dataset A. The right panel plots the posterior probability of the most probable ancestral area under the default-prior model for each node in the MCC tree (y-axis) against the corresponding posterior probability of that area under the alternative-prior model (x-axis). Pink dots represent the internal nodes where the MAP ancestral area inferred under the default-prior model differs from that inferred under the alternative-prior models. The statistic $p$ denotes the fraction of internal nodes that are shared under the default- and alternative-prior models; $f$, is the fraction of shared nodes where the MAP ancestral area differs under the default- and alternative-prior models. Note that the posterior probabilities of the MAP ancestral area under the default-prior model are generally higher than those under the alternative-prior model (*i.e.*, the default-prior model tends to mask uncertainty in the ancestral-area estimates).

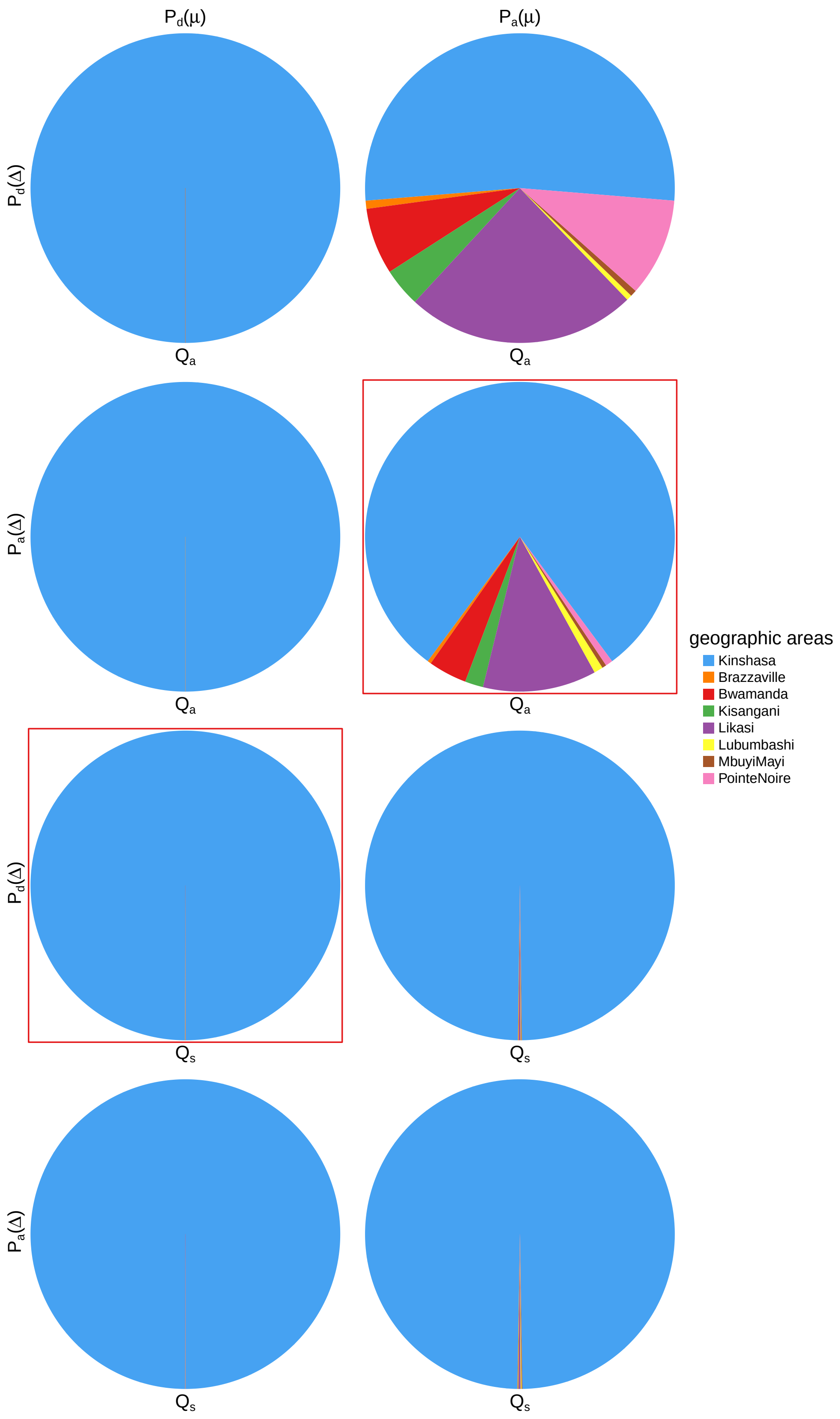


**Figure S72: The impact of prior choice on ancestral-area estimates at the root node for HIV dataset A.** The posterior probability of each ancestral area at the root node under each prior model for HIV dataset A. The pie chart enclosed in red corresponds to the comparison between the preferred default- and the preferred alternative-prior models (*c.f.*, Figure S71, left panel). Model notation follows the description in Table S10.

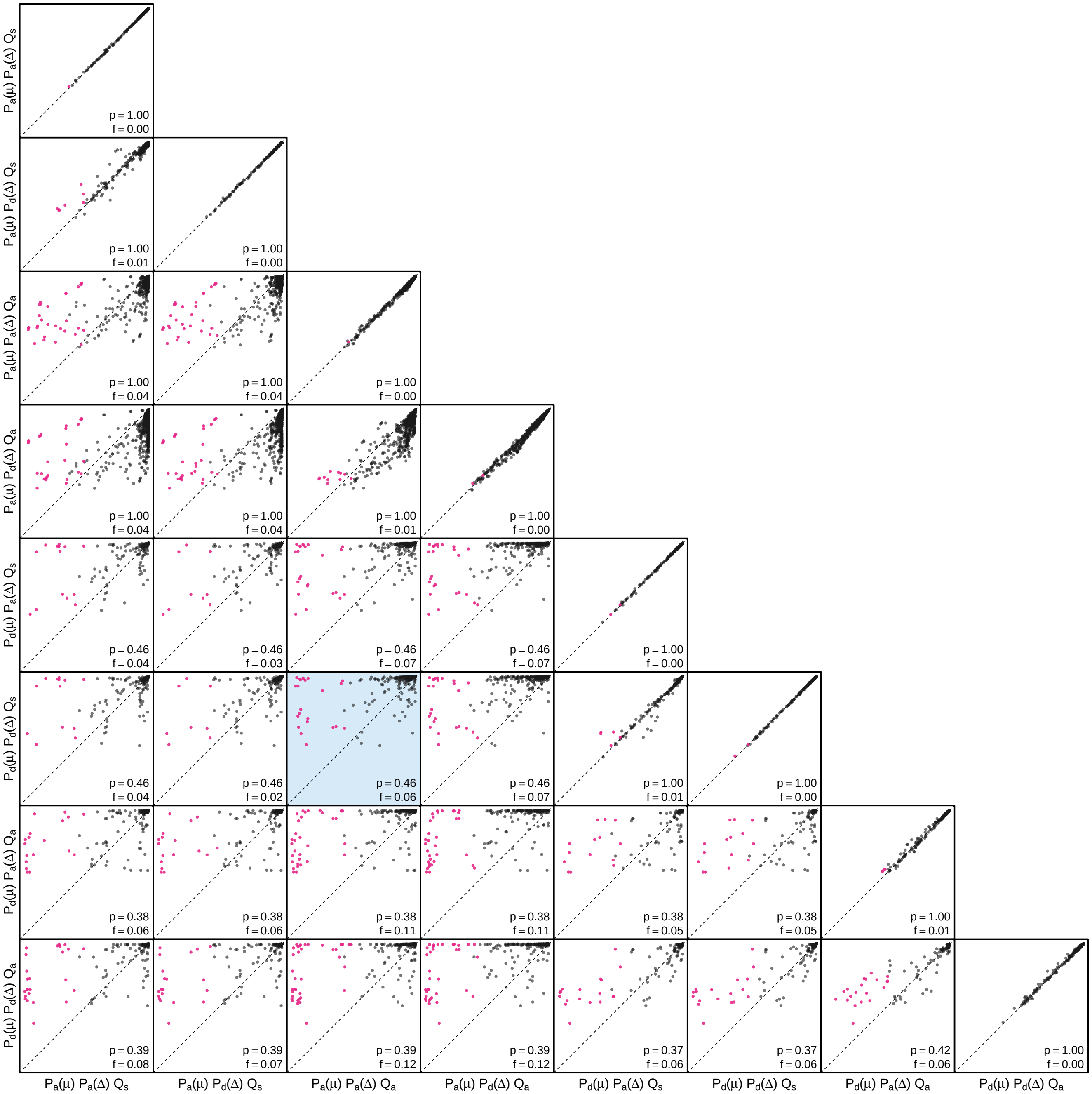


**Figure S73: The impact of prior choice on the MAP ancestral-area estimates at internal nodes for HIV dataset A.** Pairwise scatter plots compare the posterior probability of the MAP ancestral area inferred under each combination of prior models. Diagonal cells are comparisons between two replicates under the same prior model (assessing the convergence of MCMC simulations); off-diagonal cells are comparisons between different prior models, demonstrating the impact of the prior model on both the posterior probability and the identity of the MAP ancestral-area estimates at internal nodes. Pink dots represent the internal nodes where the MAP ancestral area inferred under the default-prior model differs from that inferred under the alternative-prior models. The statistic $p$ denotes the fraction of internal nodes that are shared under the default- and alternative-prior models; $f$ is the fraction of shared nodes where the MAP ancestral area differs under the default- and alternative-prior models. The shaded cell corresponds to the comparison between the preferred default- and the preferred alternative-prior models (*c.f.*, Figure S71, left panel). Model notation follows the description in Table S10.

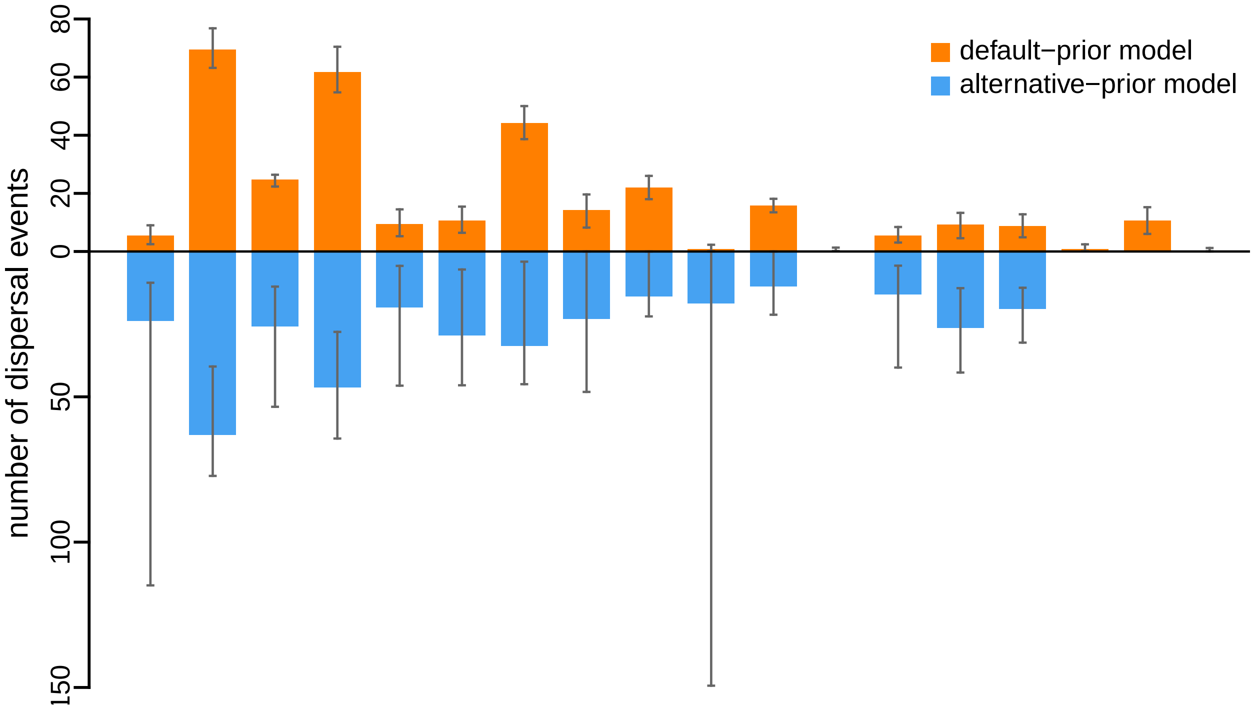


**Figure S74: The impact of prior choice on the inferred number of dispersal events between each pair of areas for HIV dataset A.** The reflected bar plot depicts the number of dispersal events inferred under the default (orange) and alternative (blue) prior models for HIV dataset A. Each bar indicates the posterior-mean number of dispersal events between a pair of areas; whiskers indicate the 95% credible interval. Note that only the number of dispersal events over the "significant" dispersal routes (*i.e.*, $2 \ln \mathrm{BF} > 2$) are figured.

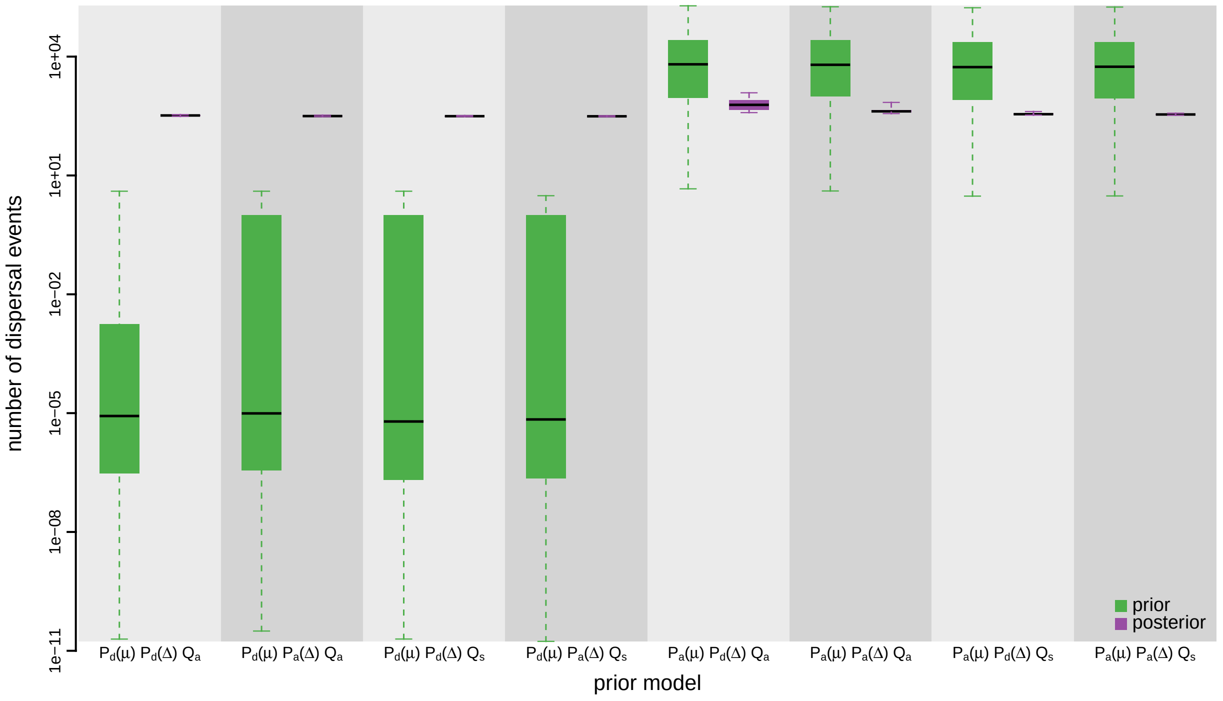


**Figure S75: The impact of prior choice on the inferred total number of dispersal events between all areas for HIV dataset A.** Each pair of boxplots depicts the specified prior (green) and corresponding posterior estimate (purple) of the number of dispersal events between all areas under each prior model: the center of each box indicates the posterior-median number of dispersal events; the box and whiskers indicate the corresponding 50% and 95% credible intervals, respectively. Model notation follows the description in Table S10.

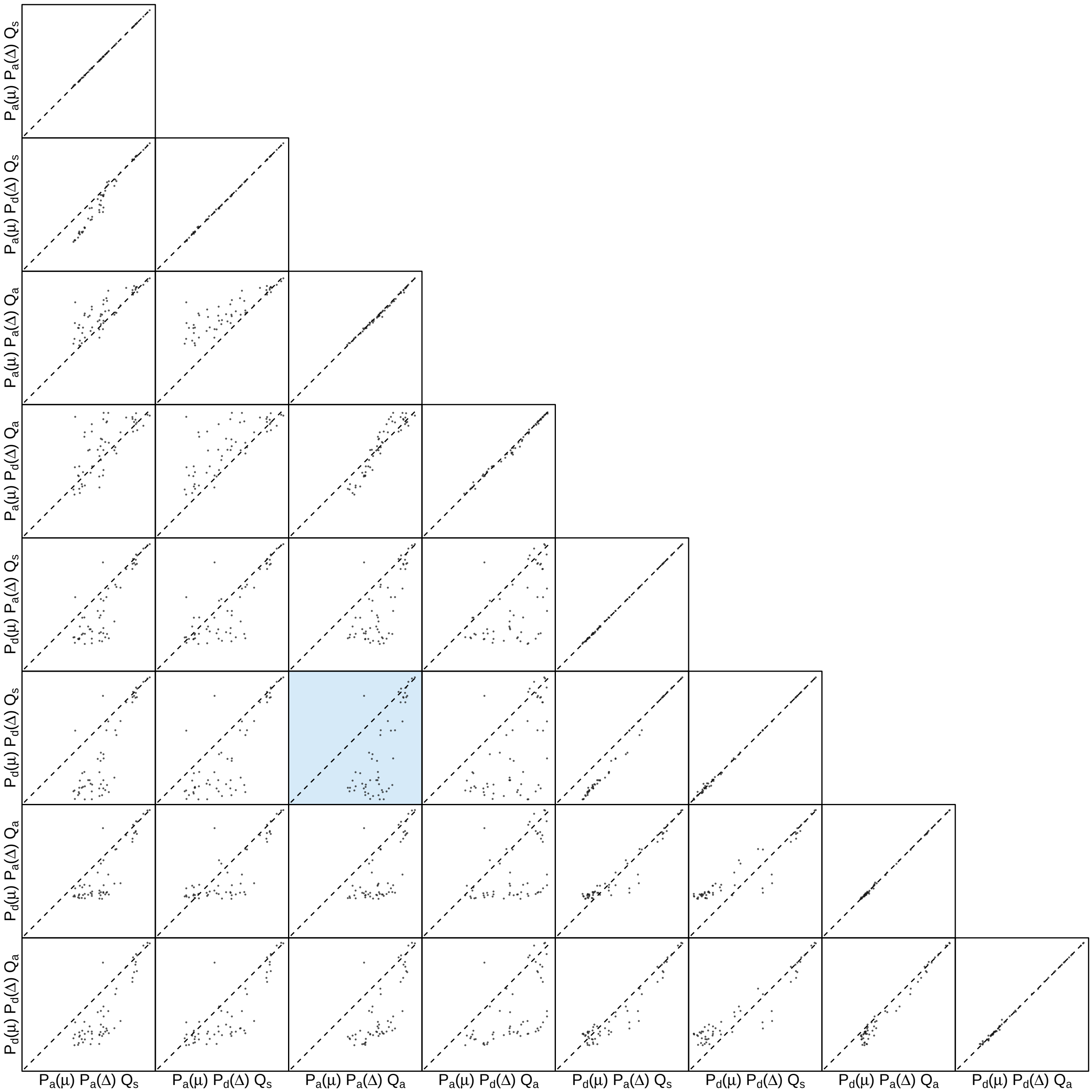


**Figure S76: The impact of prior choice on the inferred number of dispersal events between each pair of areas for HIV dataset A.** Each cell of the plot compares the inferred number of pairwise dispersal events between each pair of prior models. *x*- and *y*-axis of each cell are both on log scale. Diagonal cells are comparisons between half and the other half of the replicates under the same prior model, assessing the convergence of MCMC simulations; off-diagonal cells are comparisons between different prior models, demonstrating the impact of the prior model on the inferred number of pairwise dispersal events. The shaded cell corresponds to the comparison between the preferred default- and the preferred alternative-prior models (*c.f.*, Figure S74). Model notation follows the description in Table S10.

##### S3.3.3.2 Dataset B

*The Impact of Prior Choice on Biogeographic Model Fit*

**Table S11: Marginal-likelihood estimates of each of the eight prior models for HIV dataset B.** Columns 2–5 list marginal likelihoods inferred from four replicate analyses. Columns 6–7 list the mean and standard deviation of these marginal-likelihood estimates. The last column lists the marginal-likelihood estimates computed by combining the samples from the replicate power-posterior MCMC simulations. Candidate models are listed in rows and include all possible combinations of: (1) instantaneous-rate matrices (symmetric, $Q_s$ or asymmetric, $Q_a$); (2) priors on the average dispersal rate [default, $P_d(\mu)$ or alternative, $P_a(\mu)$], and; (3) priors on the number of dispersal routes [default, $P_d(\Delta)$ or alternative, $P_a(\Delta)$]. The preferred default- and alternative-prior models are indicated in bold text.

| Model | replicate1 | replicate2 | replicate3 | replicate4 | mean | sd | combined |
|---|---|---|---|---|---|---|---|
| $\boldsymbol{P_d(\mu)Q_aP_d(\Delta)}$ | **-1309.09** | **-1309.08** | **-1309.13** | **-1308.88** | **-1309.05** | **0.12** | **-1309.04** |
| $P_d(\mu)Q_aP_a(\Delta)$ | -1310.43 | -1310.40 | -1310.46 | -1310.37 | -1310.42 | 0.04 | -1310.41 |
| $P_d(\mu)Q_sP_d(\Delta)$ | -1335.72 | -1336.59 | -1336.31 | -1337.06 | -1336.42 | 0.56 | -1336.43 |
| $P_d(\mu)Q_sP_a(\Delta)$ | -1335.18 | -1334.56 | -1334.92 | -1334.93 | -1334.90 | 0.26 | -1334.86 |
| $P_a(\mu)Q_aP_d(\Delta)$ | -1166.55 | -1166.43 | -1166.88 | -1166.75 | -1166.65 | 0.20 | -1166.64 |
| $\boldsymbol{P_a(\mu)Q_aP_a(\Delta)}$ | **-1164.69** | **-1164.76** | **-1164.68** | **-1164.76** | **-1164.72** | **0.04** | **-1164.72** |
| $P_a(\mu)Q_sP_d(\Delta)$ | -1188.41 | -1187.71 | -1187.91 | -1187.51 | -1187.89 | 0.39 | -1187.82 |
| $P_a(\mu)Q_sP_a(\Delta)$ | -1185.73 | -1186.09 | -1186.03 | -1185.93 | -1185.94 | 0.16 | -1185.90 |

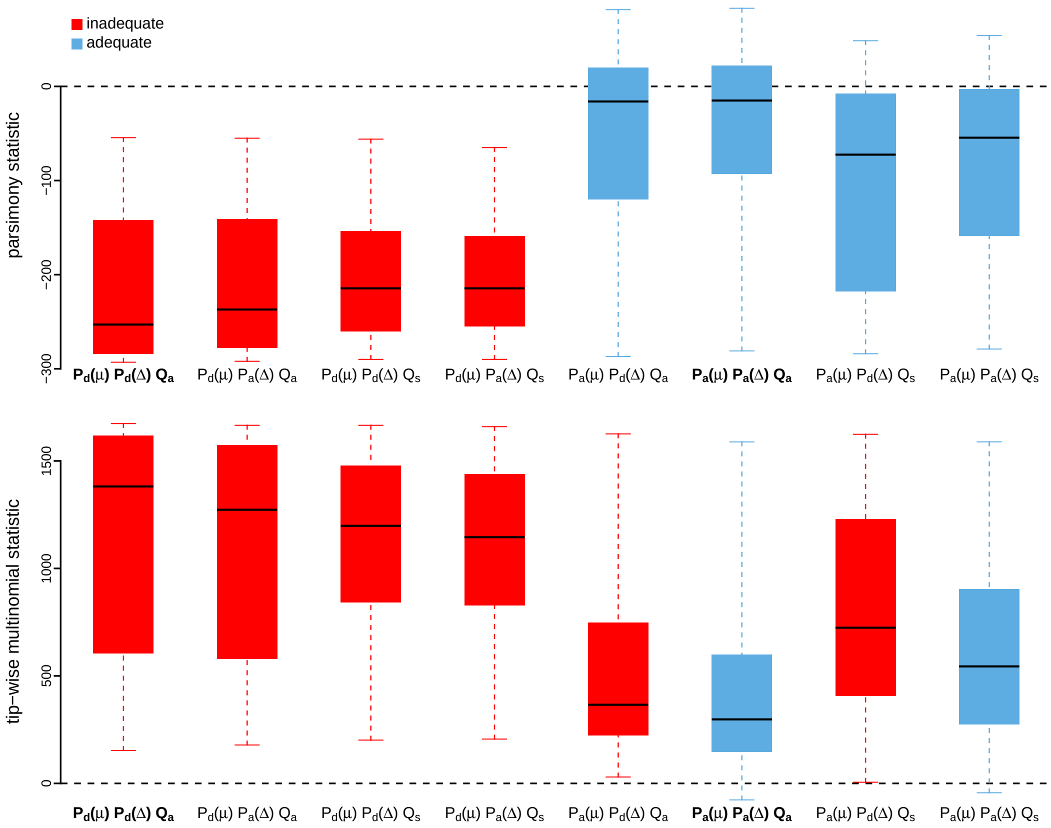


**Figure S77: Posterior-predictive distributions of the parsimony statistic (top panel) and the tip-wise multinomial statistic (bottom panel) under each of the eight prior models for HIV dataset B.** Boxplots depict the posterior-predictive distributions of the statistic under each of the eight candidate prior models: the center of each box is the median predictive value of the summary statistic; the box and whiskers indicate the corresponding 50% and 95% posterior-predictive intervals, respectively. The horizontal dashed line indicates when the simulated and observed datasets produce identical value for the summary statistic. A model is judged to be inadequate (*i.e.*, incapable of generating geographic datasets that are similar to the observed data) if its 95% posterior-predictive interval does not overlap with the dashed line. Here the preferred default prior model is inadequate, whereas the preferred alternative prior model is adequate. Notation for the candidate models is described in Table S12; the preferred default- and alternative-prior models are indicated in bold text.

*The Impact of Prior Choice on Average Dispersal Rate*

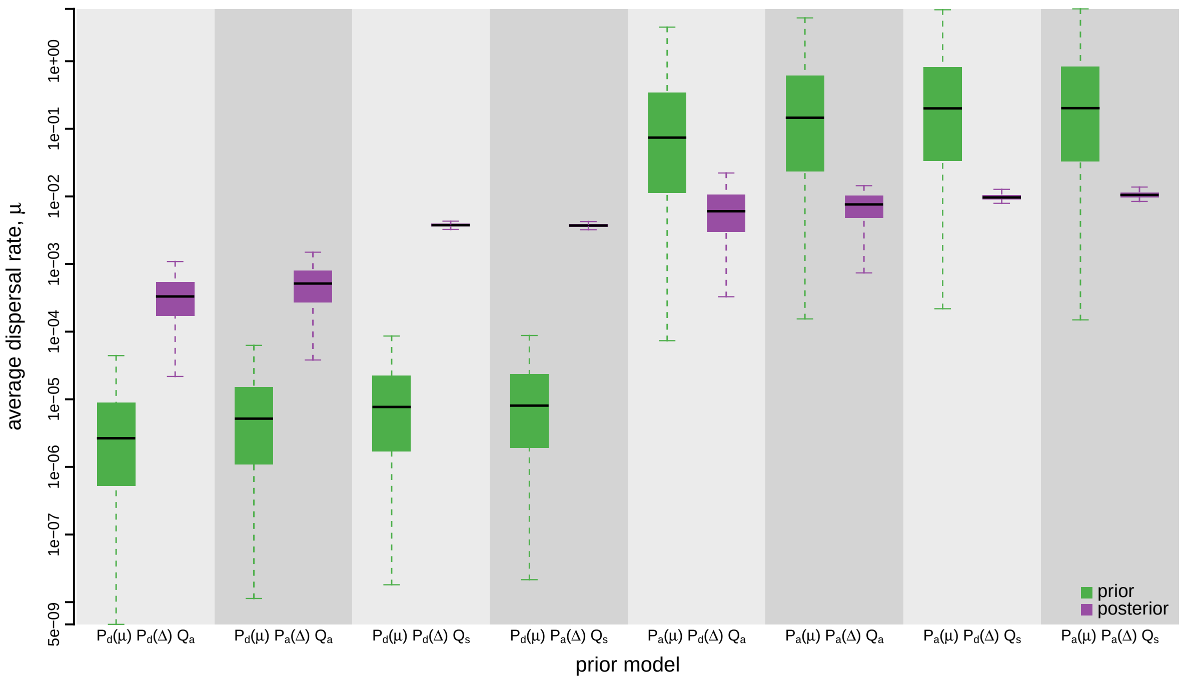


**Figure S78: The impact of prior choice on the average dispersal rate for HIV dataset B.** Each pair of boxplots depicts the specified prior (green) and corresponding posterior estimate (purple) of the average dispersal rate, $\mu$, under each prior model: the center of each box indicates the median rate; the box and whiskers indicate the corresponding 50% and 95% credible intervals, respectively. Notation for the prior models is as described in Table S11.

*The Impact of Prior Choice on Pairwise Dispersal Rates*

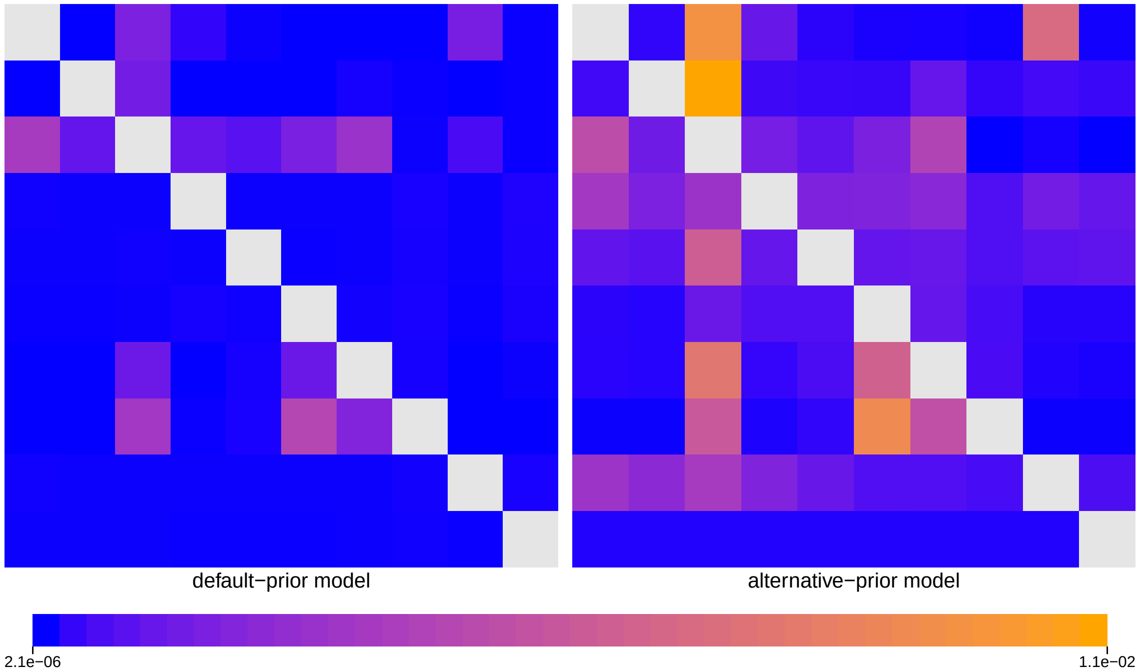


**Figure S79: The impact of prior choice on pairwise dispersal rates for HIV dataset B.** Heatmaps summarize posterior-mean estimates of the instantaneous rate of dispersal between each pair of geographic areas, $q_{ij}$, under the default (left) and alternative (right) prior models.

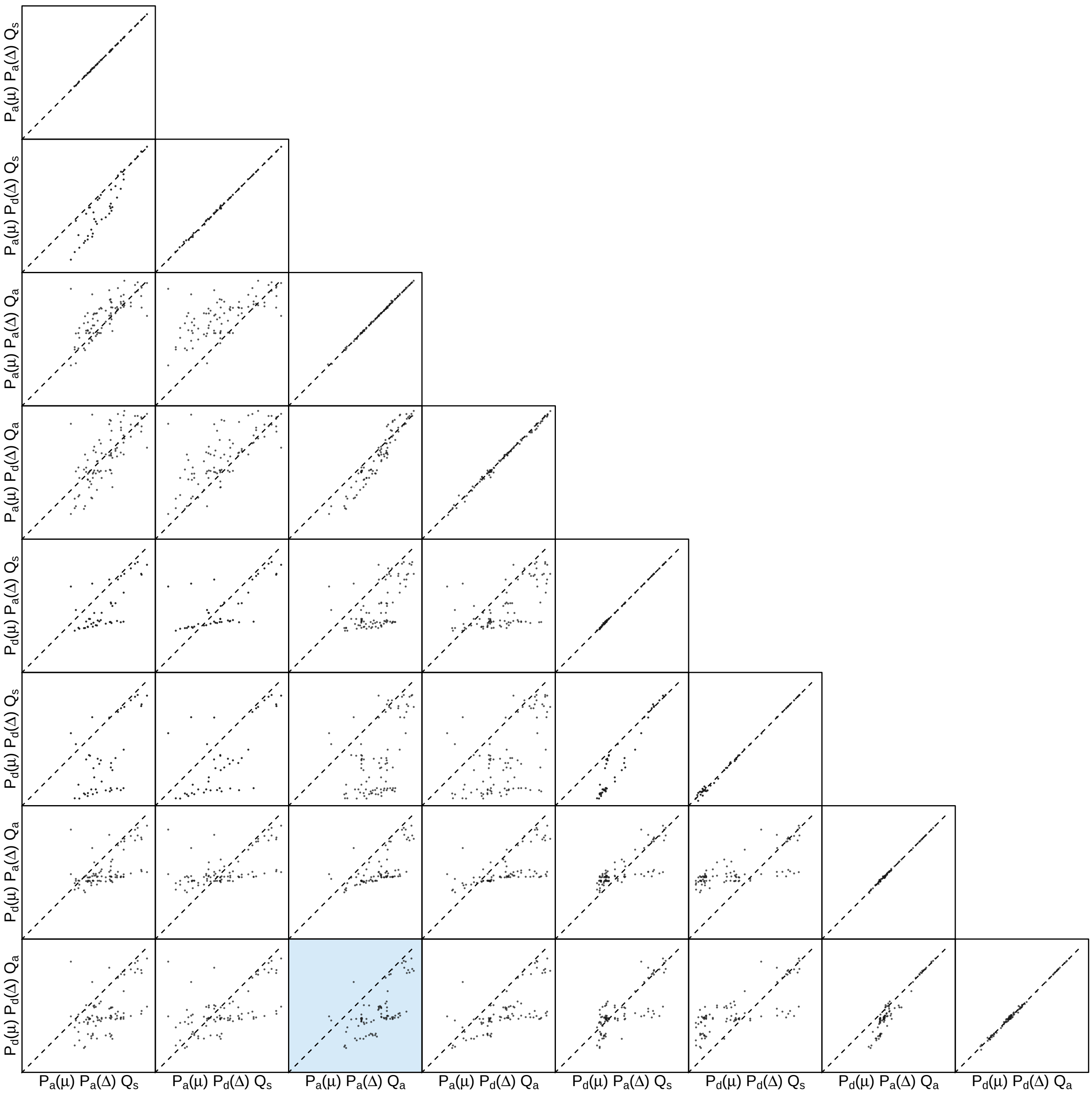


**Figure S80: The impact of prior choice on pairwise dispersal rates for HIV dataset B.** Each cell of the plot compares posterior-mean estimate of the rate of dispersal between each pair of geographic areas, $q_{ij}$, between each pair of prior models. $x$- and $y$-axis of each cell are both on log scale. Diagonal cells are comparisons between half and the other half of the replicates under the same prior model, assessing the convergence of MCMC simulations; off-diagonal cells are comparisons between different prior models, demonstrating the impact of the prior model on the estimates of pairwise dispersal rates. The shaded cell corresponds to the comparison between the preferred default- and the preferred alternative-prior models (*c.f.*, Figure S79). Notation for the prior models is as described in Table S11.

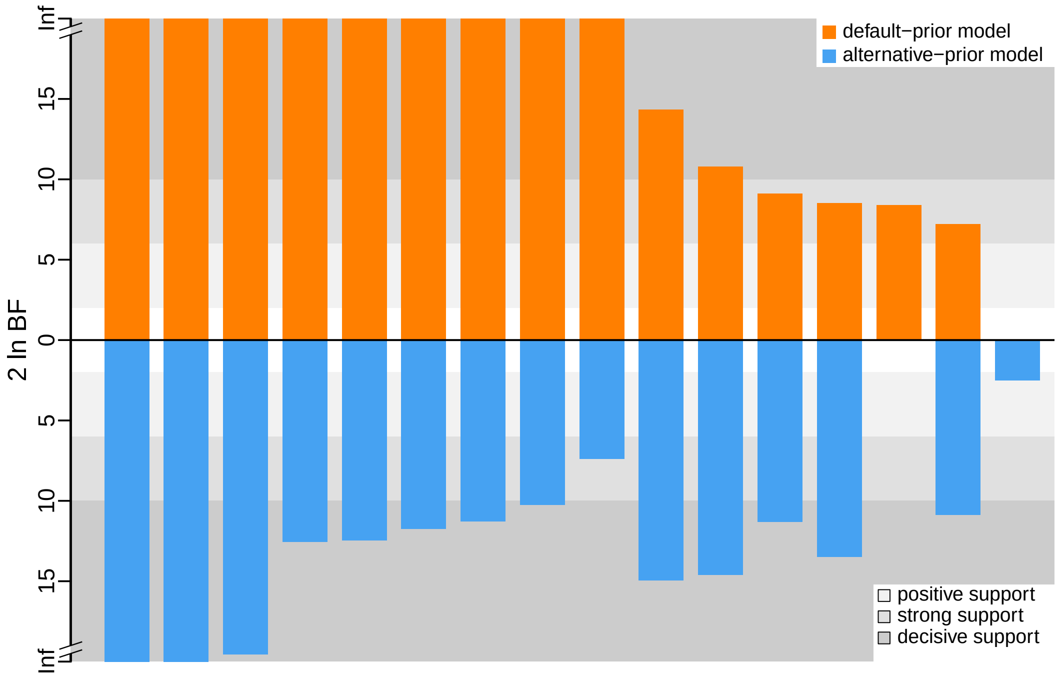


**Figure S81: The impact of prior choice on the inferred support for dispersal routes for HIV dataset B.** We compare the evidential support for each dispersal route for HIV dataset B under the default (orange) and alternative (blue) prior models. Each bar indicates the $2 \ln \mathrm{BF}$ for the corresponding dispersal route between two areas; only supported dispersal routes (*i.e.*, $2 \ln \mathrm{BF} > 2$) are plotted.

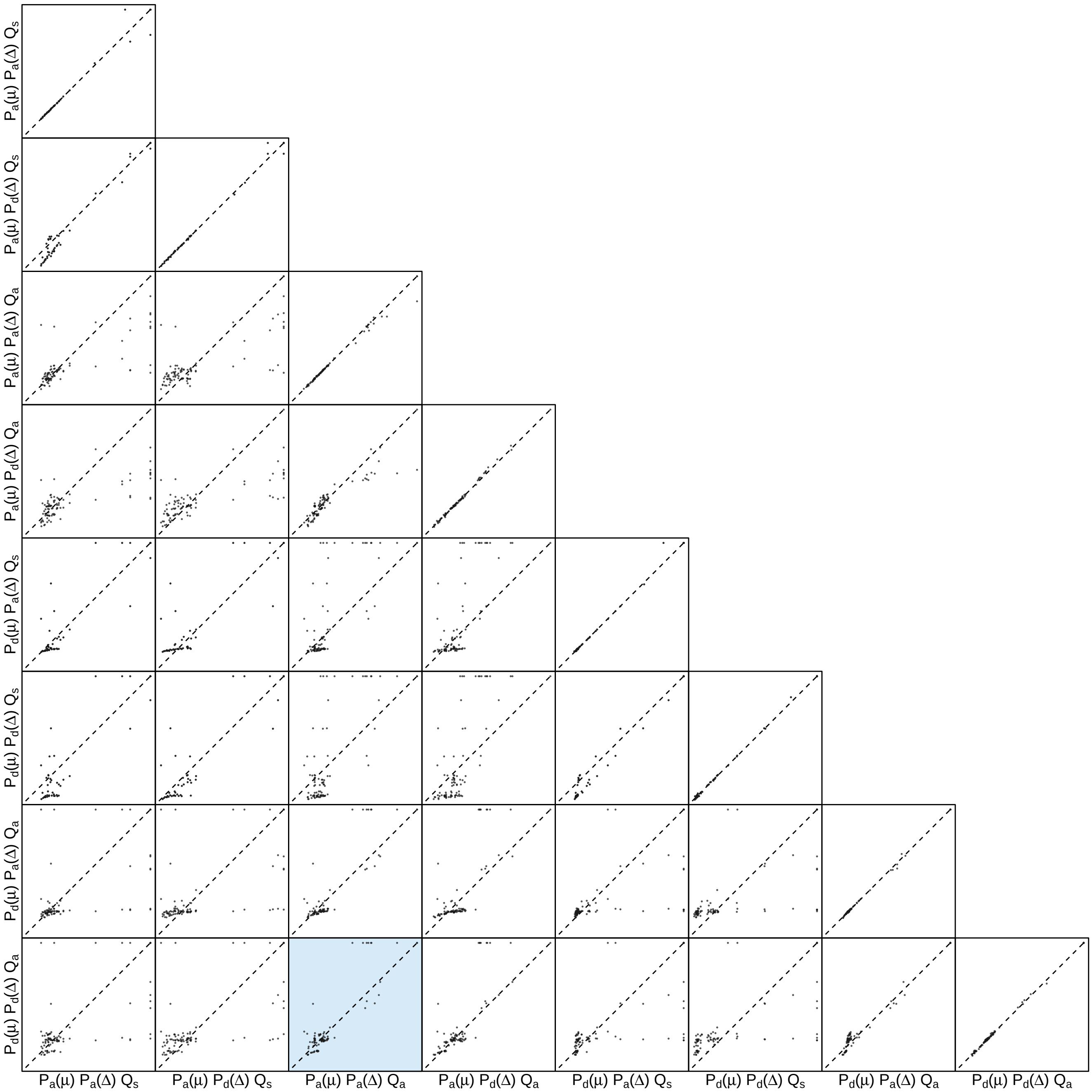


**Figure S82: The impact of prior choice on the inferred support for dispersal routes for HIV dataset B.** Each cell of the plot compares the inferred support (2 ln BF) for pairwise dispersal routes between each pair of prior models. Diagonal cells are comparisons between half and the other half of the replicates under the same prior model, assessing the convergence of MCMC simulations; off-diagonal cells are comparisons between different prior models, demonstrating the impact of the prior model on the inferred support for pairwise dispersal routes. The shaded cell corresponds to the comparison between the preferred default- and the preferred alternative-prior models (*c.f.*, Figure S81). Notation for the prior models is as described in Table S11.

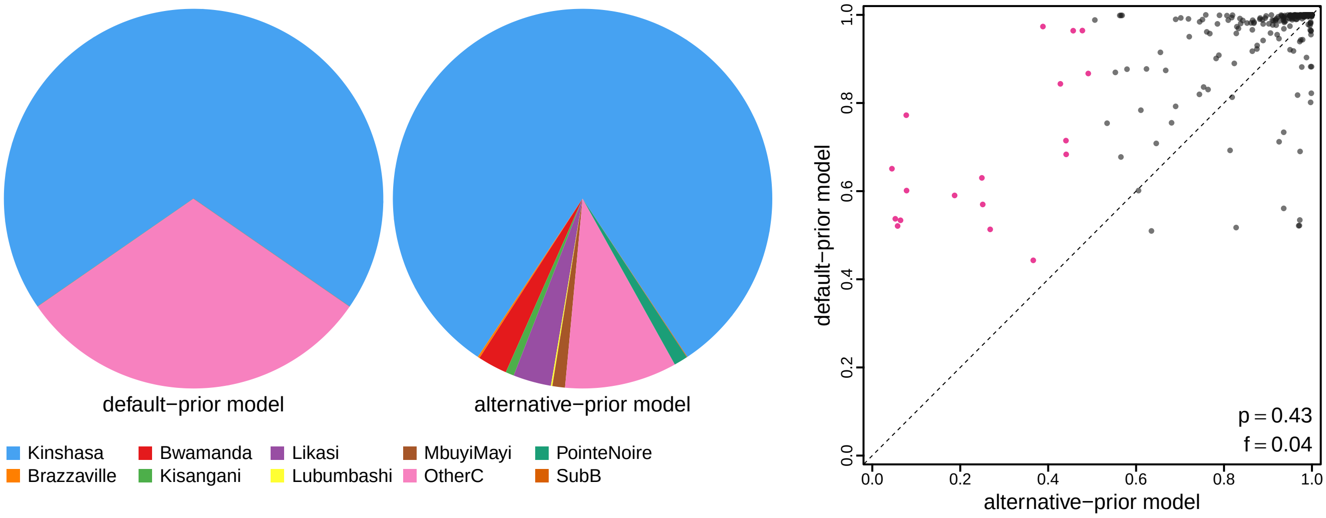


**Figure S83: The impact of prior choice on ancestral-area estimates for HIV dataset B.** The left panel compares the posterior probability of each ancestral area at the root node under the default- and alternative-prior models for HIV dataset B. The right panel plots the posterior probability of the most probable ancestral area under the default-prior model for each node in the MCC tree (y-axis) against the corresponding posterior probability of that area under the alternative-prior model (x-axis). Pink dots represent the internal nodes where the MAP ancestral area inferred under the default-prior model differs from that inferred under the alternative-prior models. The statistic $p$ denotes the fraction of internal nodes that are shared under the default- and alternative-prior models; $f$, is the fraction of shared nodes where the MAP ancestral area differs under the default- and alternative-prior models. Note that the posterior probabilities of the MAP ancestral area under the default-prior model are generally higher than those under the alternative-prior model (*i.e.*, the default-prior model tends to mask uncertainty in the ancestral-area estimates).

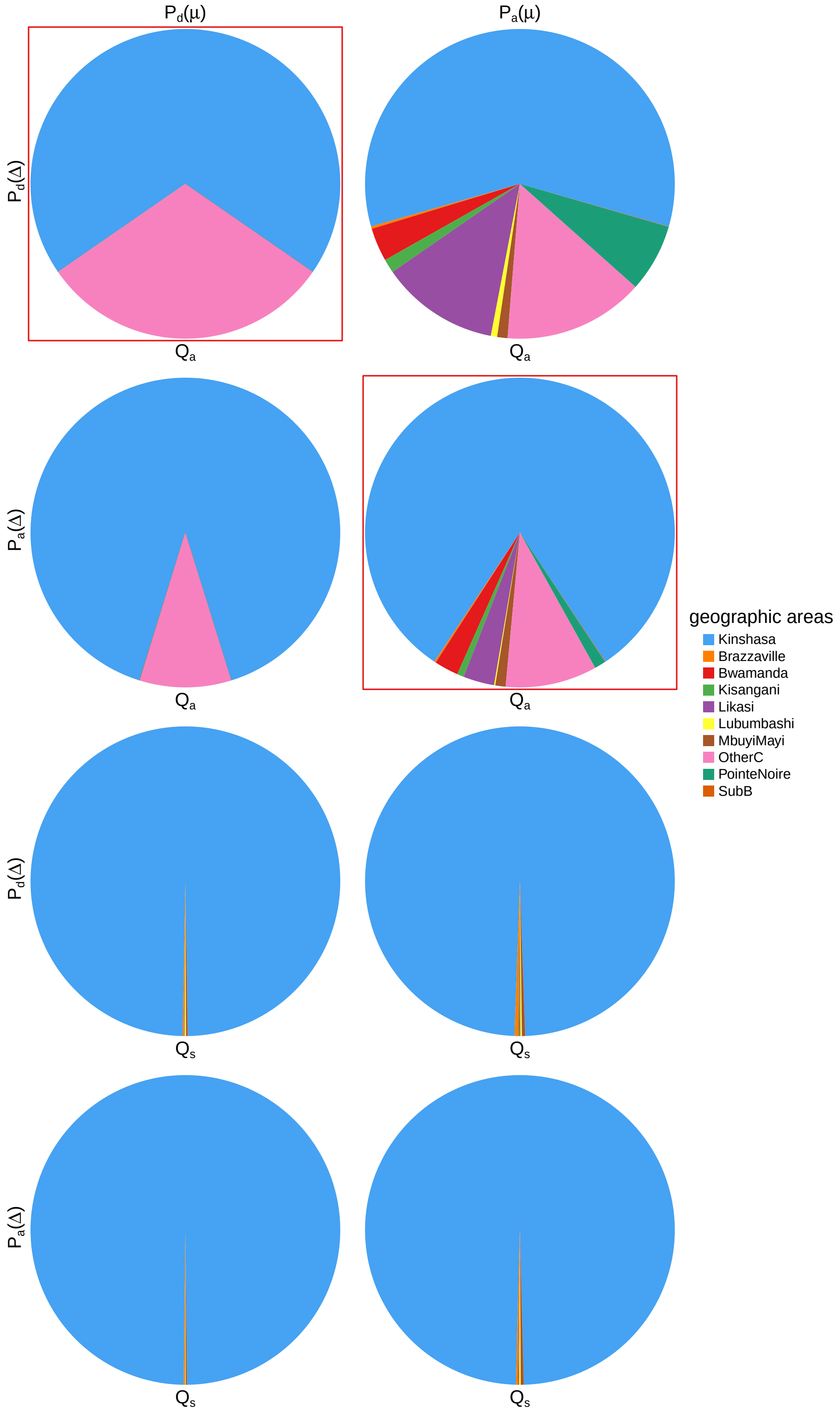


**Figure S84: The impact of prior choice on ancestral-area estimates at the root node for HIV dataset B.** The posterior probability of each ancestral area at the root node under each prior model for HIV dataset B. The pie chart enclosed in red corresponds to the comparison between the preferred default- and the preferred alternative-prior models (*c.f.*, Figure S83, left panel). Model notation follows the description in Table S11.

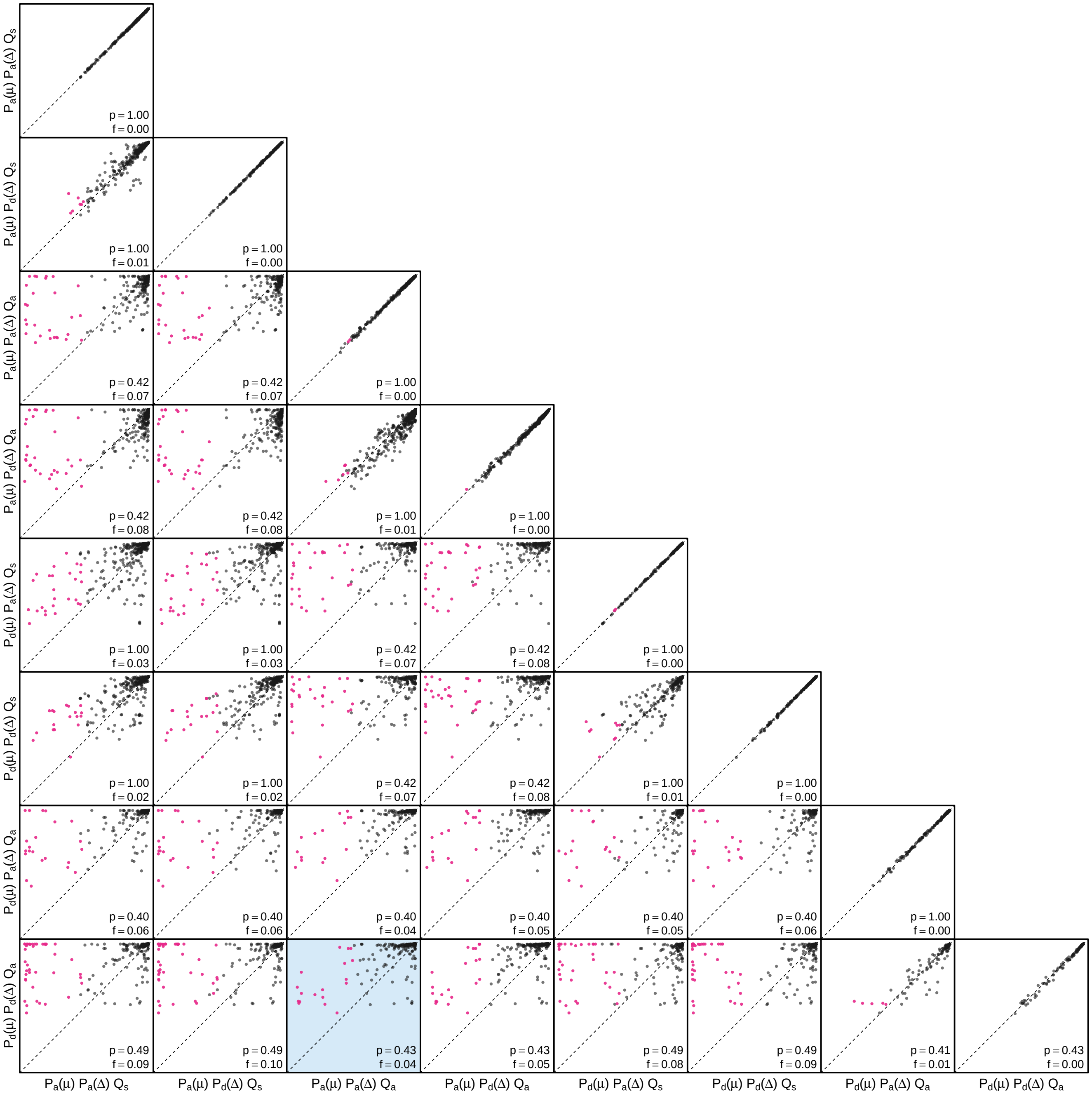


**Figure S85: The impact of prior choice on the MAP ancestral-area estimates at internal nodes for HIV dataset B.** Pairwise scatter plots compare the posterior probability of the MAP ancestral area inferred under each combination of prior models. Diagonal cells are comparisons between two replicates under the same prior model (assessing the convergence of MCMC simulations); off-diagonal cells are comparisons between different prior models, demonstrating the impact of the prior model on both the posterior probability and the identity of the MAP ancestral-area estimates at internal nodes. Pink dots represent the internal nodes where the MAP ancestral area inferred under the default-prior model differs from that inferred under the alternative-prior models. The statistic $p$ denotes the fraction of internal nodes that are shared under the default- and alternative-prior models; $f$ is the fraction of shared nodes where the MAP ancestral area differs under the default- and alternative-prior models. The shaded cell corresponds to the comparison between the preferred default- and the preferred alternative-prior models (*c.f.*, Figure S83, left panel). Model notation follows the description in Table S11.

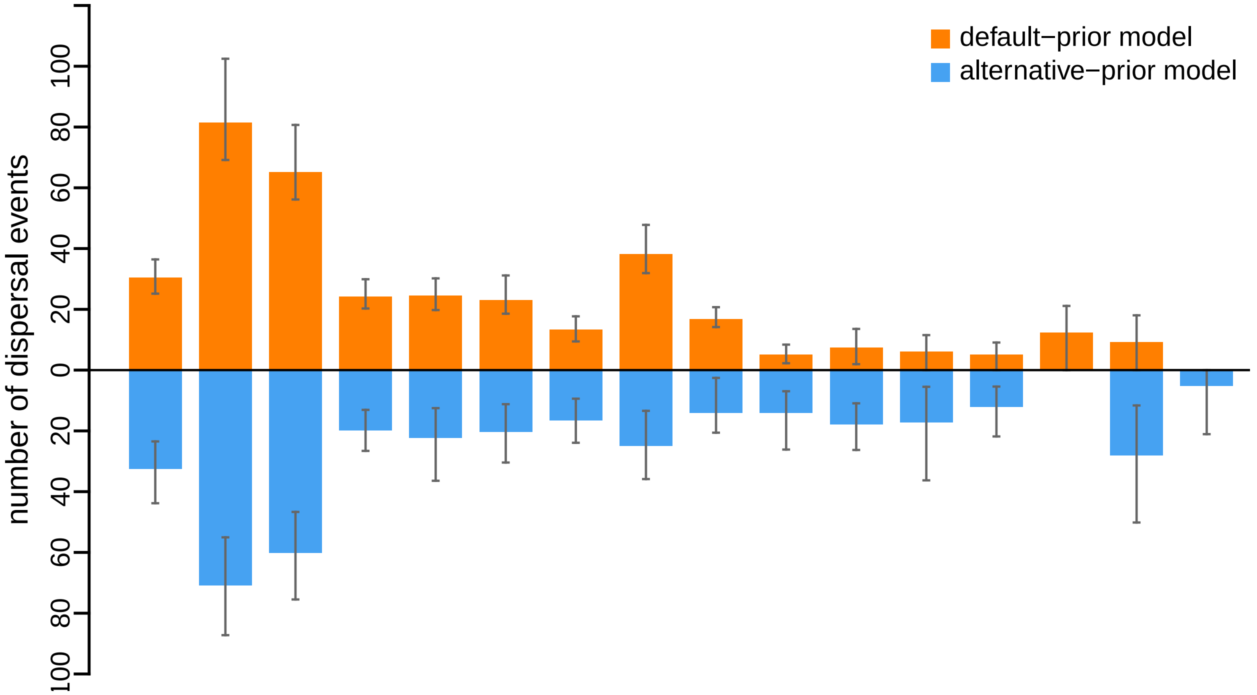


**Figure S86: The impact of prior choice on the inferred number of dispersal events between each pair of areas for HIV dataset B.** The reflected bar plot depicts the number of dispersal events inferred under the default (orange) and alternative (blue) prior models for HIV dataset B. Each bar indicates the posterior-mean number of dispersal events between a pair of areas; whiskers indicate the 95% credible interval. Note that only the number of dispersal events over the "significant" dispersal routes (*i.e.*, $2 \ln \mathrm{BF} > 2$) are figured.

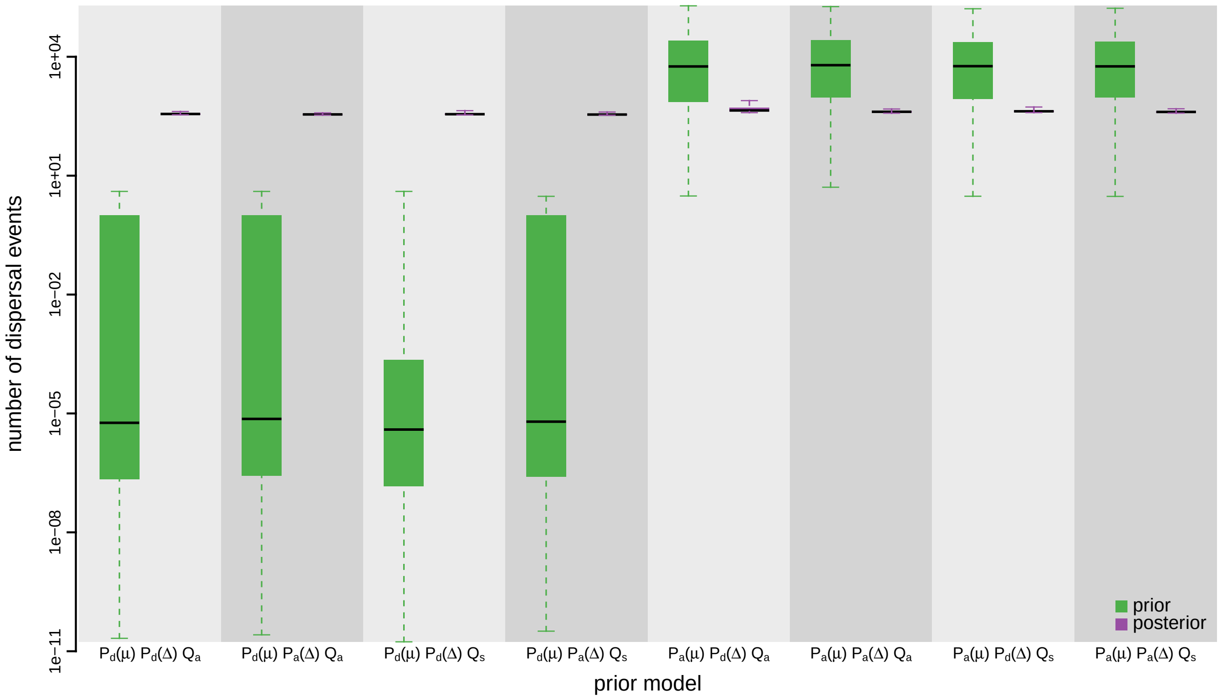


**Figure S87: The impact of prior choice on the inferred total number of dispersal events between all areas for HIV dataset B.** Each pair of boxplots depicts the specified prior (green) and corresponding posterior estimate (purple) of the number of dispersal events between all areas under each prior model: the center of each box indicates the posterior-median number of dispersal events; the box and whiskers indicate the corresponding 50% and 95% credible intervals, respectively. Model notation follows the description in Table S11.

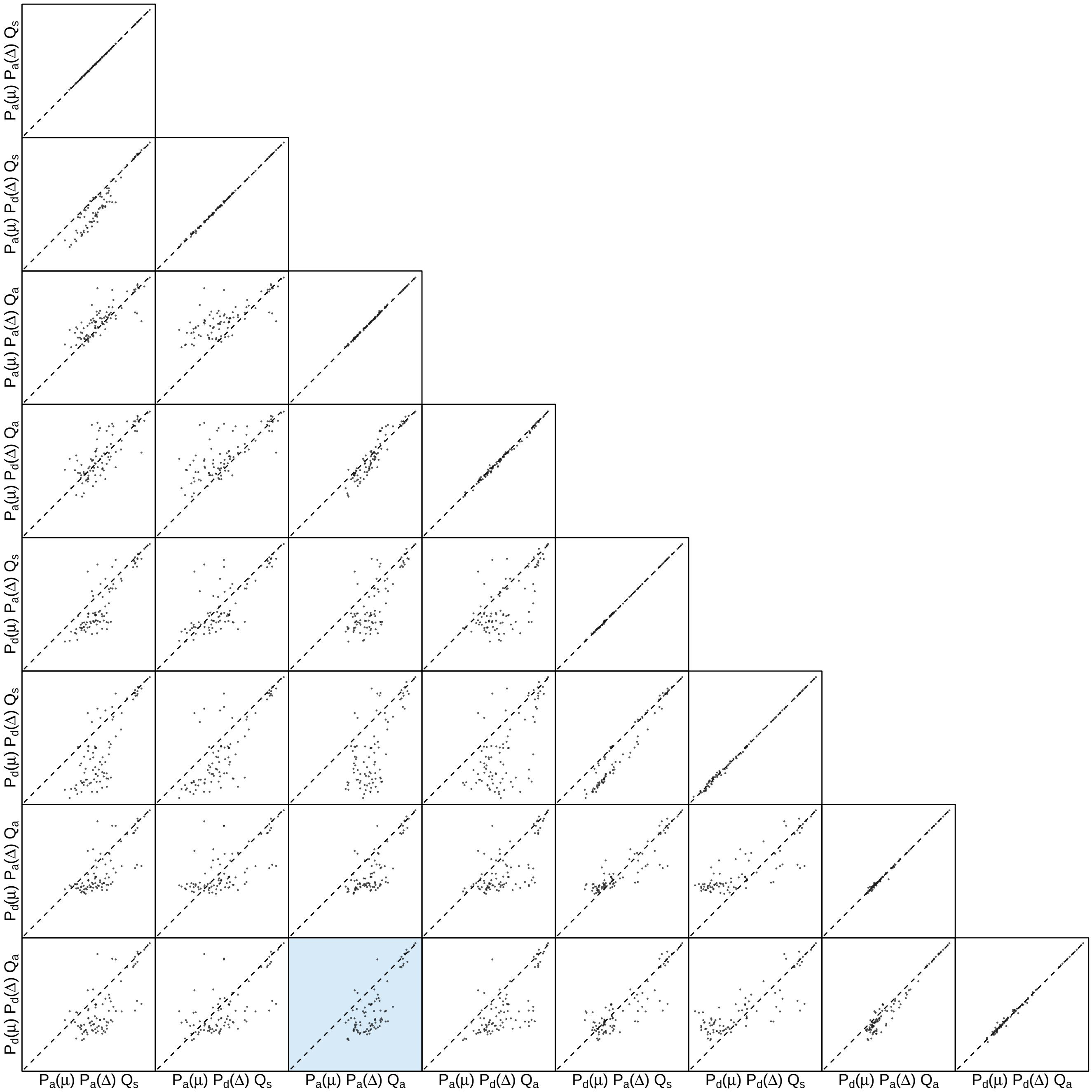


**Figure S88: The impact of prior choice on the inferred number of dispersal events between each pair of areas for HIV dataset B.** Each cell of the plot compares the inferred number of pairwise dispersal events between each pair of prior models. *x*- and *y*-axis of each cell are both on log scale. Diagonal cells are comparisons between half and the other half of the replicates under the same prior model, assessing the convergence of MCMC simulations; off-diagonal cells are comparisons between different prior models, demonstrating the impact of the prior model on the inferred number of pairwise dispersal events. The shaded cell corresponds to the comparison between the preferred default- and the preferred alternative-prior models (*c.f.*, Figure S86). Model notation follows the description in Table S11.

###### S3.3.3.3 Dataset C

*The Impact of Prior Choice on Biogeographic Model Fit*

**Table S12: Marginal-likelihood estimates of each of the eight prior models for HIV dataset C.** Columns 2–5 list marginal likelihoods inferred from four replicate analyses. Columns 6–7 list the mean and standard deviation of these marginal-likelihood estimates. The last column lists the marginal-likelihood estimates computed by combining the samples from the replicate power-posterior MCMC simulations. Candidate models are listed in rows and include all possible combinations of: (1) instantaneous-rate matrices (symmetric, $Q_s$ or asymmetric, $Q_a$); (2) priors on the average dispersal rate [default, $P_d(\mu)$ or alternative, $P_a(\mu)$], and; (3) priors on the number of dispersal routes [default, $P_d(\Delta)$ or alternative, $P_a(\Delta)$]. The preferred default- and alternative-prior models are indicated in bold text.

| Model | replicate1 | replicate2 | replicate3 | replicate4 | mean | sd | combined |
|---|---|---|---|---|---|---|---|
| $\boldsymbol{P_d(\mu)Q_aP_d(\Delta)}$ | **-835.93** | **-835.83** | **-835.65** | **-836.36** | **-835.94** | **0.30** | **-835.91** |
| $P_d(\mu)Q_aP_a(\Delta)$ | -835.36 | -835.01 | -835.17 | -835.56 | -835.27 | 0.24 | -835.23 |
| $P_d(\mu)Q_sP_d(\Delta)$ | -859.10 | -859.03 | -858.77 | -858.98 | -858.97 | 0.14 | -858.83 |
| $P_d(\mu)Q_sP_a(\Delta)$ | -858.01 | -858.00 | -858.05 | -857.66 | -857.93 | 0.18 | -857.87 |
| $P_a(\mu)Q_aP_d(\Delta)$ | -731.09 | -731.05 | -731.08 | -731.22 | -731.11 | 0.07 | -731.11 |
| $\boldsymbol{P_a(\mu)Q_aP_a(\Delta)}$ | **-726.85** | **-726.65** | **-726.71** | **-726.98** | **-726.80** | **0.15** | **-726.80** |
| $P_a(\mu)Q_sP_d(\Delta)$ | -730.16 | -730.01 | -730.04 | -729.88 | -730.02 | 0.11 | -729.97 |
| $P_a(\mu)Q_sP_a(\Delta)$ | -728.56 | -728.34 | -728.44 | -728.38 | -728.43 | 0.09 | -728.43 |

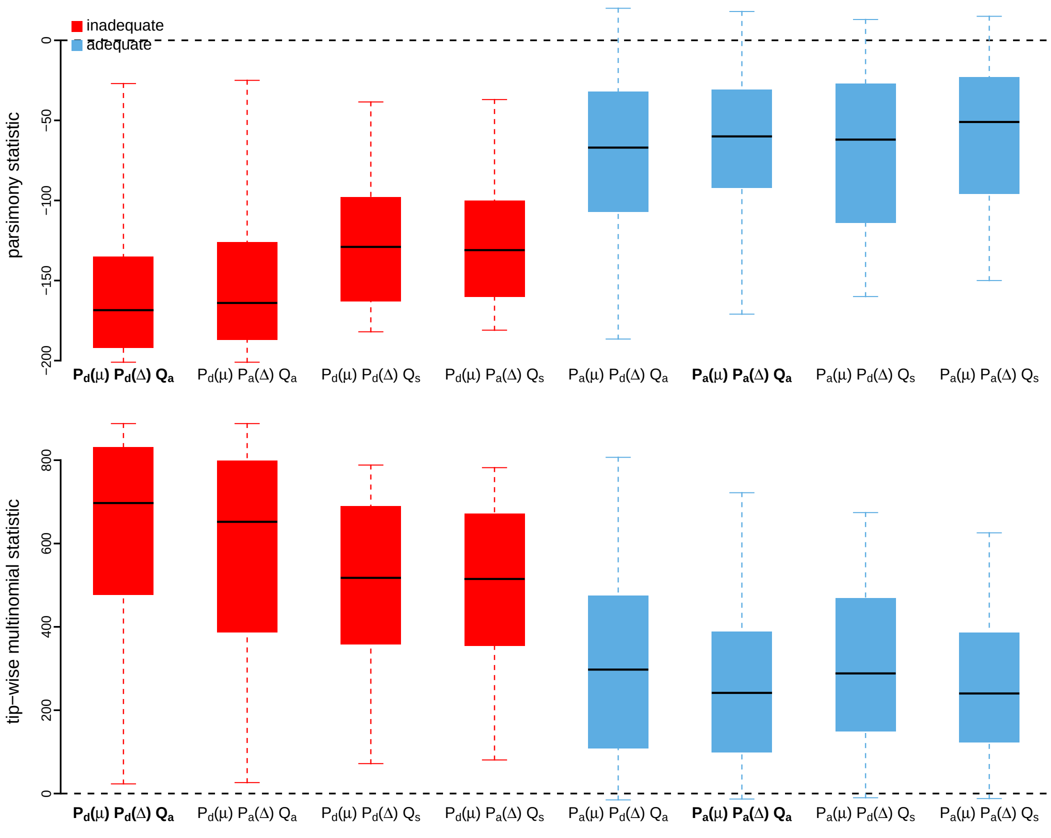


**Figure S89: Posterior-predictive distributions of the parsimony statistic (top panel) and the tip-wise multinomial statistic (bottom panel) under each of the eight prior models for HIV dataset C.** Boxplots depict the posterior-predictive distributions of the statistic under each of the eight candidate prior models: the center of each box is the median predictive value of the summary statistic; the box and whiskers indicate the corresponding 50% and 95% posterior-predictive intervals, respectively. The horizontal dashed line indicates when the simulated and observed datasets produce identical value for the summary statistic. A model is judged to be inadequate (*i.e.*, incapable of generating geographic datasets that are similar to the observed data) if its 95% posterior-predictive interval does not overlap with the dashed line. Here the preferred default prior model is inadequate, whereas the preferred alternative prior model is adequate. Notation for the candidate models is described in Table S12; the preferred default- and alternative-prior models are indicated in bold text.

*The Impact of Prior Choice on Average Dispersal Rate*

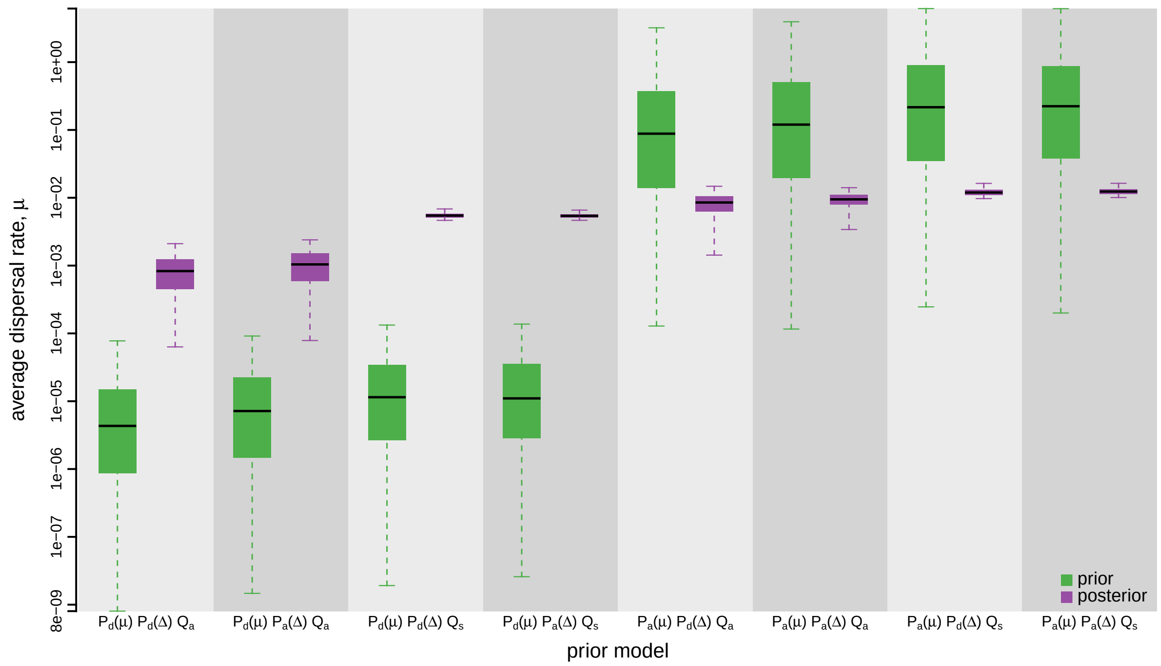


**Figure S90: The impact of prior choice on the average dispersal rate for HIV dataset C.** Each pair of boxplots depicts the specified prior (green) and corresponding posterior estimate (purple) of the average dispersal rate, $\mu$, under each prior model: the center of each box indicates the median rate; the box and whiskers indicate the corresponding 50% and 95% credible intervals, respectively. Notation for the prior models is as described in Table S12.

*The Impact of Prior Choice on Pairwise Dispersal Rates*

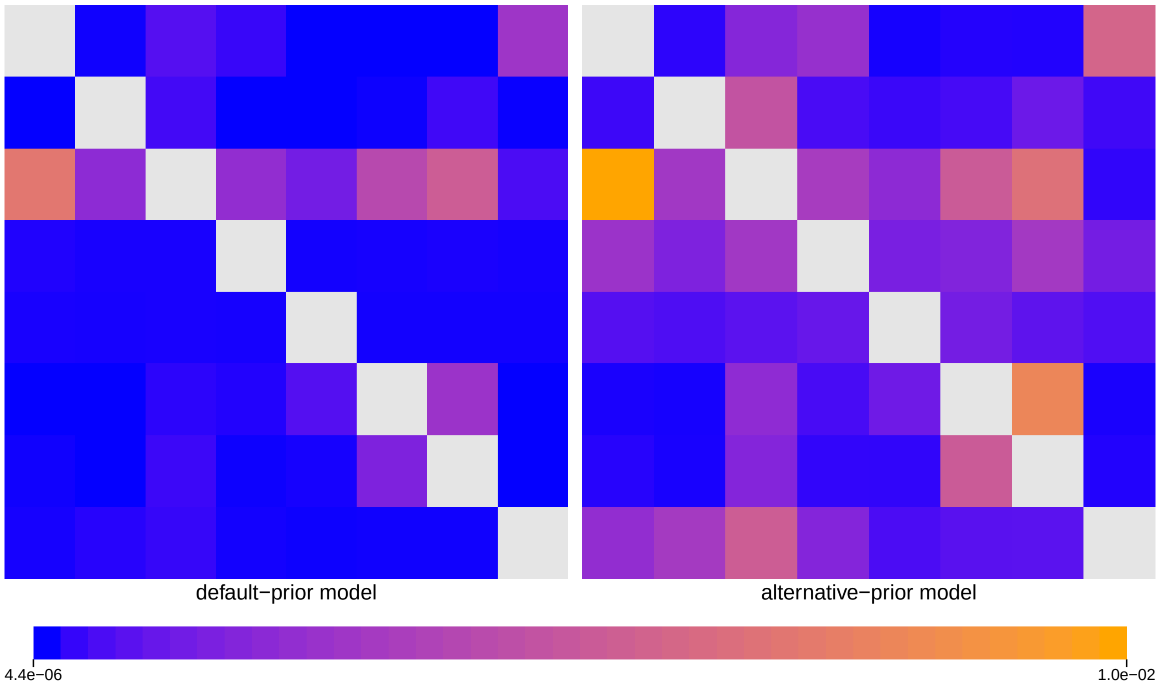


**Figure S91: The impact of prior choice on pairwise dispersal rates for HIV dataset C.** Heatmaps summarize posterior-mean estimates of the instantaneous rate of dispersal between each pair of geographic areas, $q_{ij}$, under the default (left) and alternative (right) prior models.

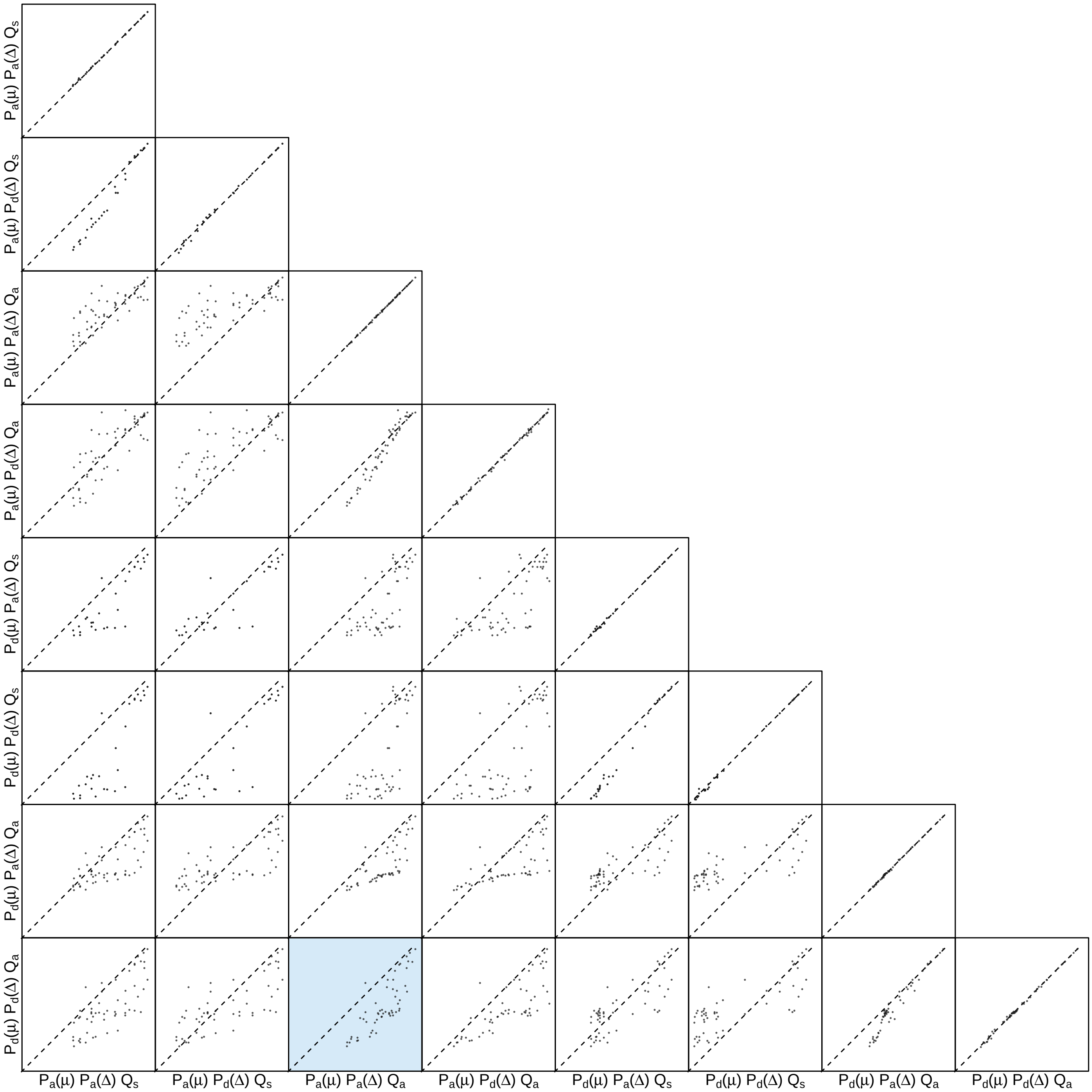


**Figure S92: The impact of prior choice on pairwise dispersal rates for HIV dataset C.** Each cell of the plot compares posterior-mean estimate of the rate of dispersal between each pair of geographic areas, $q_{ij}$, between each pair of prior models. $x$- and $y$-axis of each cell are both on log scale. Diagonal cells are comparisons between half and the other half of the replicates under the same prior model, assessing the convergence of MCMC simulations; off-diagonal cells are comparisons between different prior models, demonstrating the impact of the prior model on the estimates of pairwise dispersal rates. The shaded cell corresponds to the comparison between the preferred default- and the preferred alternative-prior models (*c.f.*, Figure S91). Notation for the prior models is as described in Table S12.

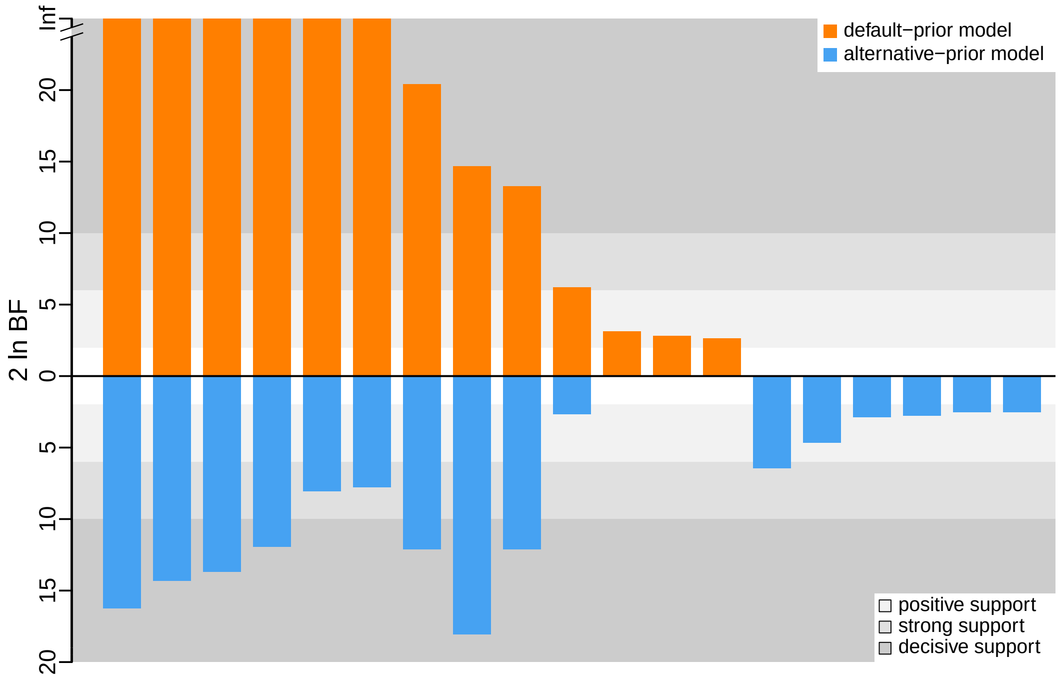


**Figure S93: The impact of prior choice on the inferred support for dispersal routes for HIV dataset C.** We compare the evidential support for each dispersal route for HIV dataset C under the default (orange) and alternative (blue) prior models. Each bar indicates the $2 \ln \mathrm{BF}$ for the corresponding dispersal route between two areas; only supported dispersal routes (*i.e.*, $2 \ln \mathrm{BF} > 2$) are plotted.

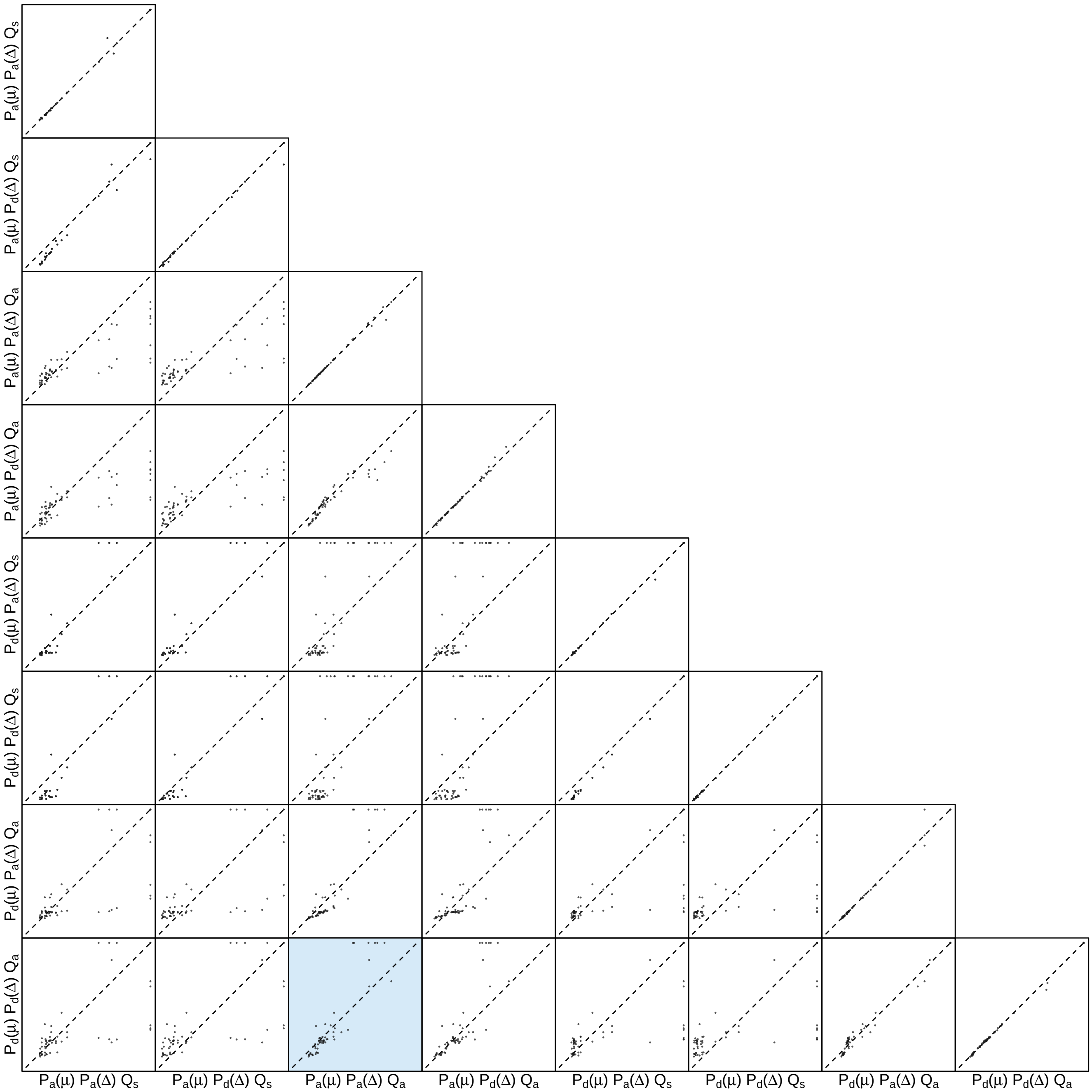


**Figure S94: The impact of prior choice on the inferred support for dispersal routes for HIV dataset C.** Each cell of the plot compares the inferred support (2 ln BF) for pairwise dispersal routes between each pair of prior models. Diagonal cells are comparisons between half and the other half of the replicates under the same prior model, assessing the convergence of MCMC simulations; off-diagonal cells are comparisons between different prior models, demonstrating the impact of the prior model on the inferred support for pairwise dispersal routes. The shaded cell corresponds to the comparison between the preferred default- and the preferred alternative-prior models (*c.f.*, Figure S93). Notation for the prior models is as described in Table S12.

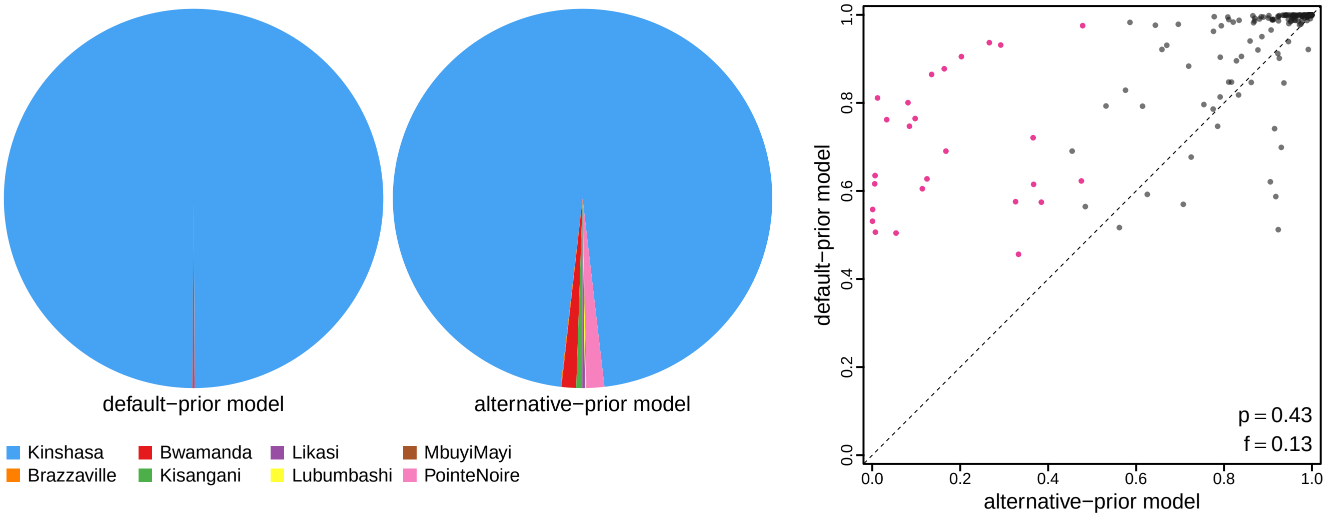


**Figure S95: The impact of prior choice on ancestral-area estimates for HIV dataset C.** The left panel compares the posterior probability of each ancestral area at the root node under the default- and alternative-prior models for HIV dataset C. The right panel plots the posterior probability of the most probable ancestral area under the default-prior model for each node in the MCC tree (y-axis) against the corresponding posterior probability of that area under the alternative-prior model (x-axis). Pink dots represent the internal nodes where the MAP ancestral area inferred under the default-prior model differs from that inferred under the alternative-prior models. The statistic $p$ denotes the fraction of internal nodes that are shared under the default- and alternative-prior models; $f$, is the fraction of shared nodes where the MAP ancestral area differs under the default- and alternative-prior models. Note that the posterior probabilities of the MAP ancestral area under the default-prior model are generally higher than those under the alternative-prior model (*i.e.*, the default-prior model tends to mask uncertainty in the ancestral-area estimates).

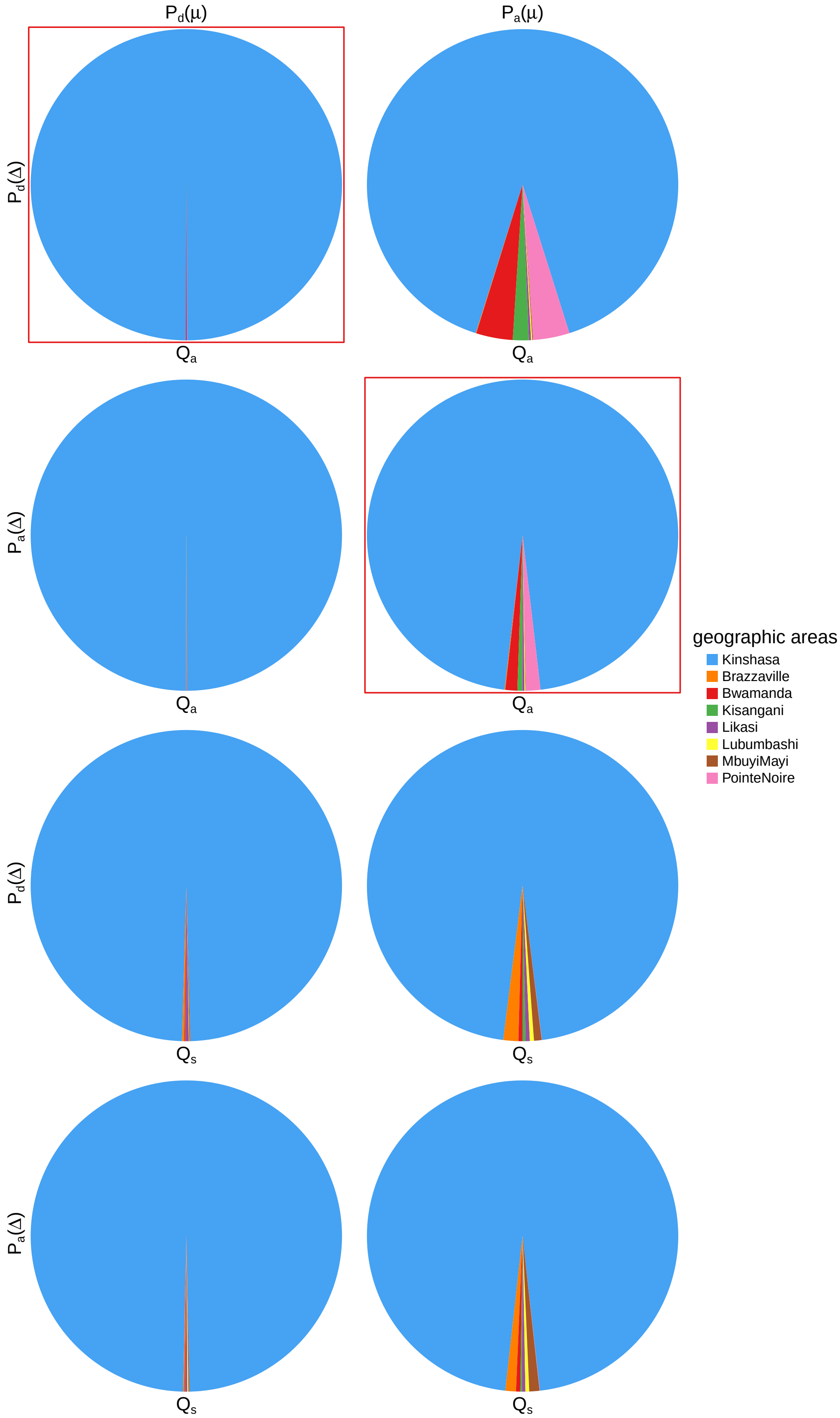


**Figure S96: The impact of prior choice on ancestral-area estimates at the root node for HIV dataset C.** The posterior probability of each ancestral area at the root node under each prior model for HIV dataset C. The pie chart enclosed in red corresponds to the comparison between the preferred default- and the preferred alternative-prior models (*c.f.*, Figure S95, left panel). Model notation follows the description in Table S12.

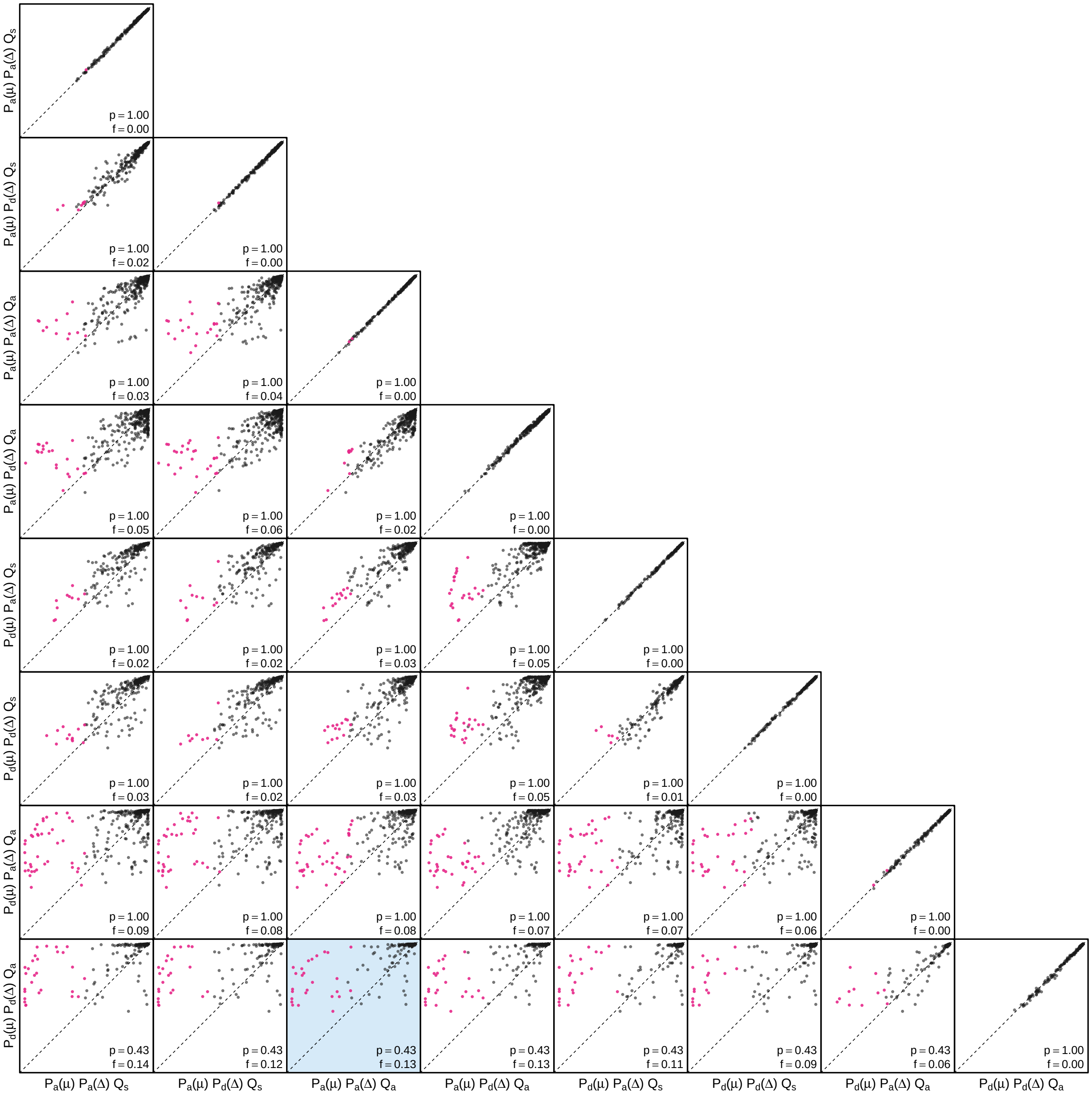


**Figure S97: The impact of prior choice on the MAP ancestral-area estimates at internal nodes for HIV dataset C.** Pairwise scatter plots compare the posterior probability of the MAP ancestral area inferred under each combination of prior models. Diagonal cells are comparisons between two replicates under the same prior model (assessing the convergence of MCMC simulations); off-diagonal cells are comparisons between different prior models, demonstrating the impact of the prior model on both the posterior probability and the identity of the MAP ancestral-area estimates at internal nodes. Pink dots represent the internal nodes where the MAP ancestral area inferred under the default-prior model differs from that inferred under the alternative-prior models. The statistic $p$ denotes the fraction of internal nodes that are shared under the default- and alternative-prior models; $f$ is the fraction of shared nodes where the MAP ancestral area differs under the default- and alternative-prior models. The shaded cell corresponds to the comparison between the preferred default- and the preferred alternative-prior models (*c.f.*, Figure S95, left panel). Model notation follows the description in Table S12.

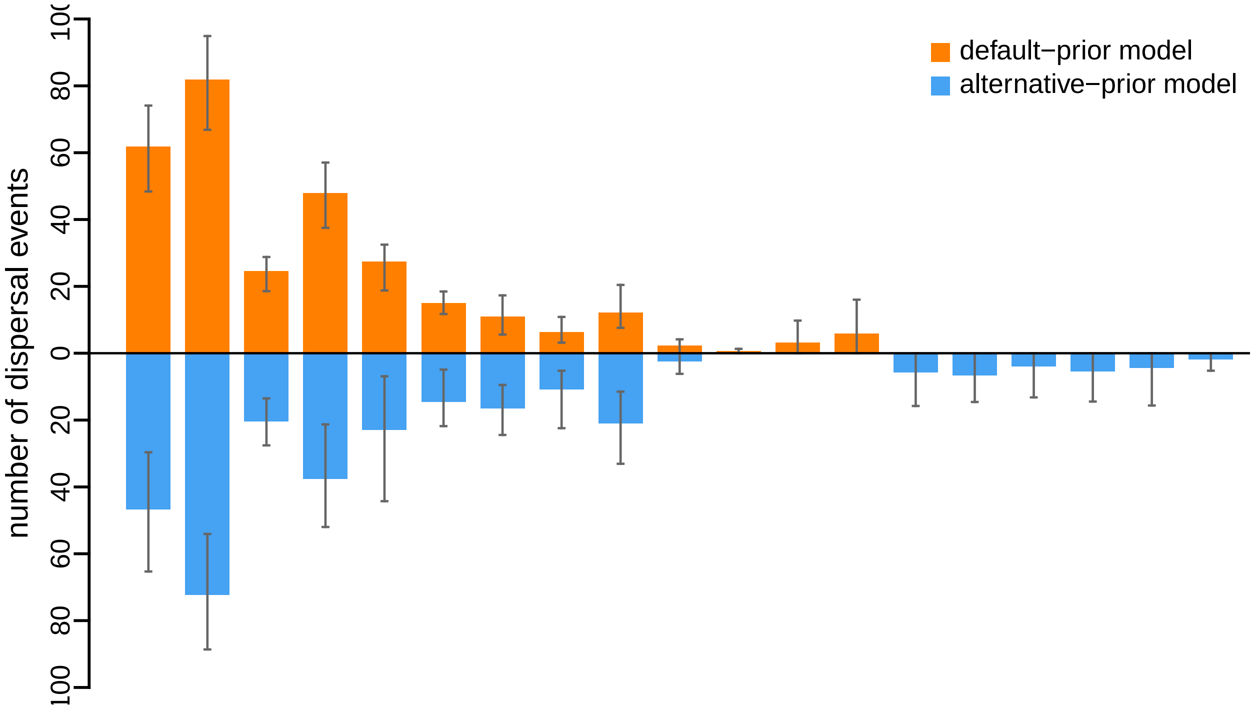


**Figure S98: The impact of prior choice on the inferred number of dispersal events between each pair of areas for HIV dataset C.** The reflected bar plot depicts the number of dispersal events inferred under the default (orange) and alternative (blue) prior models for HIV dataset C. Each bar indicates the posterior-mean number of dispersal events between a pair of areas; whiskers indicate the 95% credible interval. Note that only the number of dispersal events over the "significant" dispersal routes (*i.e.*, $2 \ln \mathrm{BF} > 2$) are figured.

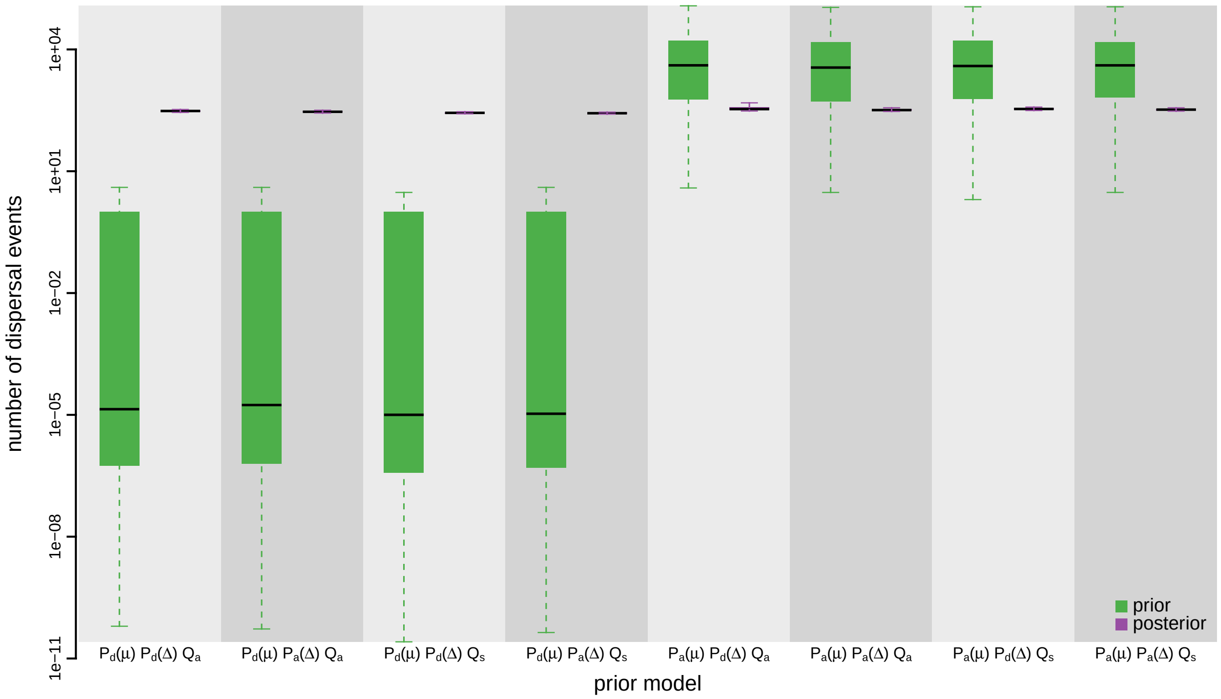


**Figure S99: The impact of prior choice on the inferred total number of dispersal events between all areas for HIV dataset C.** Each pair of boxplots depicts the specified prior (green) and corresponding posterior estimate (purple) of the number of dispersal events between all areas under each prior model: the center of each box indicates the posterior-median number of dispersal events; the box and whiskers indicate the corresponding 50% and 95% credible intervals, respectively. Model notation follows the description in Table S12.

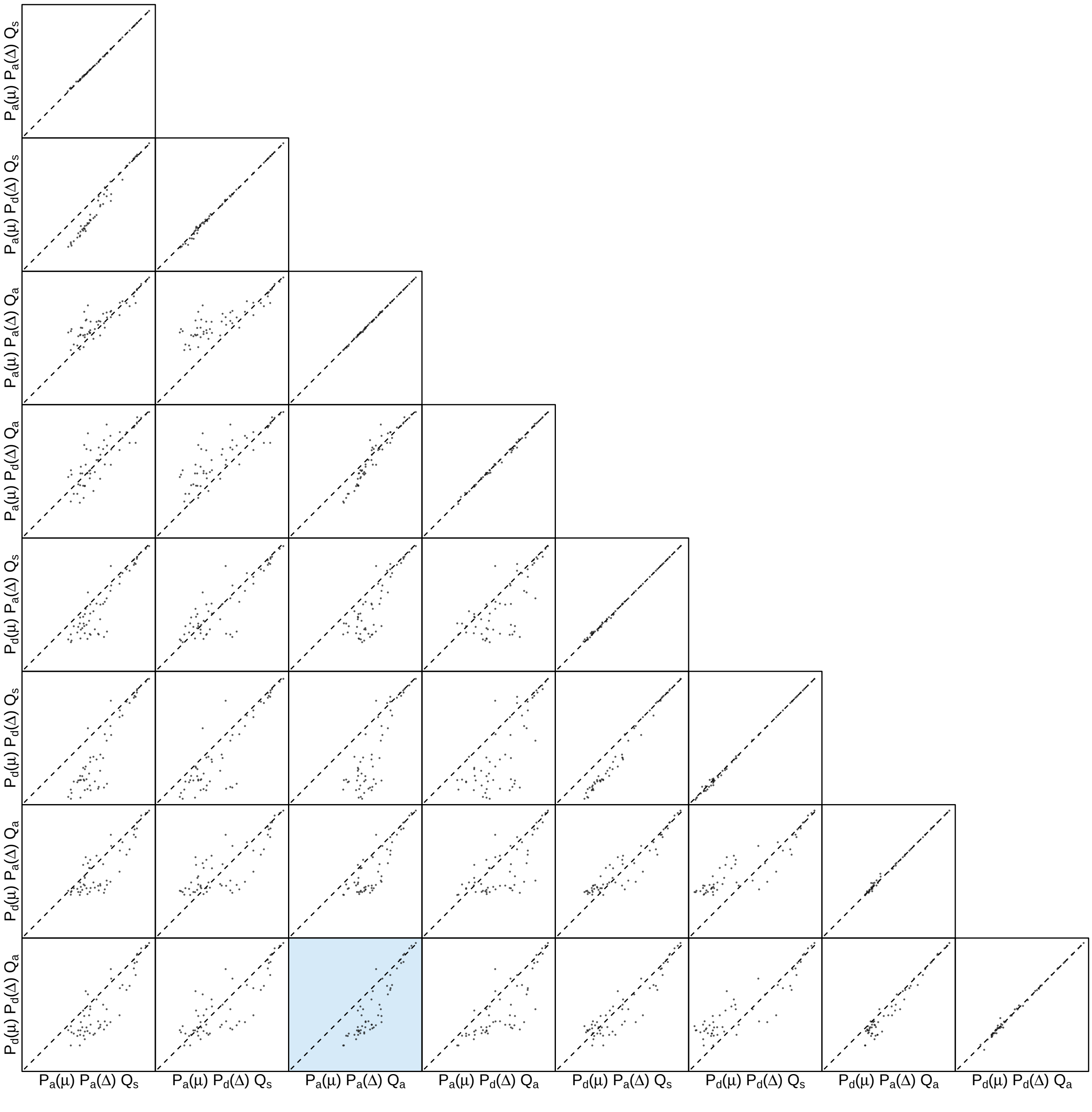


**Figure S100: The impact of prior choice on the inferred number of dispersal events between each pair of areas for HIV dataset C.** Each cell of the plot compares the inferred number of pairwise dispersal events between each pair of prior models. *x*- and *y*-axis of each cell are both on log scale. Diagonal cells are comparisons between half and the other half of the replicates under the same prior model, assessing the convergence of MCMC simulations; off-diagonal cells are comparisons between different prior models, demonstrating the impact of the prior model on the inferred number of pairwise dispersal events. The shaded cell corresponds to the comparison between the preferred default- and the preferred alternative-prior models (*c.f.*, Figure S98). Model notation follows the description in Table S12.

###### S3.3.4 Influenza Virus

Bedford et al. (2015) inferred the geographic dynamics of the four most prevalent, globally circulating human seasonal influenza viruses (A/H3N2, A/H1N1, B/Victoria, and B/Yamagata). The authors collected complete sequences of the HA1 domain of the hemagglutinin (HA) gene for these influenza viruses, sampled across most of the major geographic areas between 2000–2012. They applied down-sampling schemes to the complete dataset to reduce the impact of possible surveillance biases, resulting into three data(sub)sets that were used to assess the robustness of the main conclusions to sampling effects. These includes: the "large" datasets (containing between 1999 to 4006 sequences for each virus); the "small" datasets (containing between 1240 to 1391 sequences for each virus), and the "alternative" datasets (containing between 1223 to 1967 sequences for each virus) (details see Bedford et al. 2015).

We explored the impact of prior choice on phylodynamic inferences for two of the datasets from their study; the "small" versions of the A/H3N2 and B/Yamagata datasets. Our choice of datasets was motivated by computational considerations (the comprehensive series of analyses in our study entails a very large computational burden even for the "small" datasets), coupled with our desire to include one virus from each of the two major types of human seasonal influenza virus (*i.e.*, A and B).

We acquired the marginal posterior probability distribution of phylogenies (inferred from the sequence data and used to perform sequential analyses in Bedford et al. 2015) directly from the Github repository of the original study. Each posterior distribution included 101 trees, which was then treated as the prior distribution of phylogenies in the second step of the sequential phylodynamic inference. The `trees` files containing these distributions are available in our GitHub and Dryad repositories. We acquired the sampling geographic location data from the XML scripts provided by the original study; this sampling-area data are available in our GitHub and Dryad repositories.

The MCMC simulations used in the second step of our sequential analyses and of the analyses used to estimate marginal likelihoods under each prior model are described above in Section S3.1. Details of these analyses are available in the XML scripts included in our GitHub and Dryad repositories.

###### S3.3.4.1 A/H3N2 Dataset

*The Impact of Prior Choice on Biogeographic Model Fit*

**Table S13: Marginal-likelihood estimates of each of the eight prior models for Influenza A/H3N2 dataset.** Columns 2–5 list marginal likelihoods inferred from four replicate analyses. Columns 6–7 list the mean and standard deviation of these marginal-likelihood estimates. The last column lists the marginal-likelihood estimates computed by combining the samples from the replicate power-posterior MCMC simulations. Candidate models are listed in rows and include all possible combinations of: (1) instantaneous-rate matrices (symmetric, $Q_s$ or asymmetric, $Q_a$); (2) priors on the average dispersal rate [default, $P_d(\mu)$ or alternative, $P_a(\mu)$], and; (3) priors on the number of dispersal routes [default, $P_d(\Delta)$ or alternative, $P_a(\Delta)$]. The preferred default- and alternative-prior models are indicated in bold text.

| Model | replicate1 | replicate2 | replicate3 | replicate4 | mean | sd | combined |
|---|---|---|---|---|---|---|---|
| $\boldsymbol{P_d(\mu)Q_aP_d(\Delta)}$ | **-2874.26** | **-2874.16** | **-2873.86** | **-2873.33** | **-2873.90** | **0.42** | **-2873.68** |
| $P_d(\mu)Q_aP_a(\Delta)$ | -2825.17 | -2825.57 | -2825.15 | -2825.22 | -2825.28 | 0.20 | -2825.27 |
| $P_d(\mu)Q_sP_d(\Delta)$ | -2886.53 | -2886.96 | -2887.53 | -2887.54 | -2887.14 | 0.49 | -2887.06 |
| $P_d(\mu)Q_sP_a(\Delta)$ | -2838.53 | -2837.93 | -2838.49 | -2838.33 | -2838.32 | 0.27 | -2838.12 |
| $P_a(\mu)Q_aP_d(\Delta)$ | -2300.60 | -2300.77 | -2300.68 | -2300.60 | -2300.66 | 0.08 | -2300.65 |
| $\boldsymbol{P_a(\mu)Q_aP_a(\Delta)}$ | **-2275.52** | **-2275.49** | **-2275.48** | **-2275.42** | **-2275.48** | **0.04** | **-2275.47** |
| $P_a(\mu)Q_sP_d(\Delta)$ | -2310.78 | -2309.85 | -2312.15 | -2311.25 | -2311.01 | 0.96 | -2310.93 |
| $P_a(\mu)Q_sP_a(\Delta)$ | -2280.06 | -2280.62 | -2280.40 | -2280.54 | -2280.40 | 0.25 | -2280.41 |

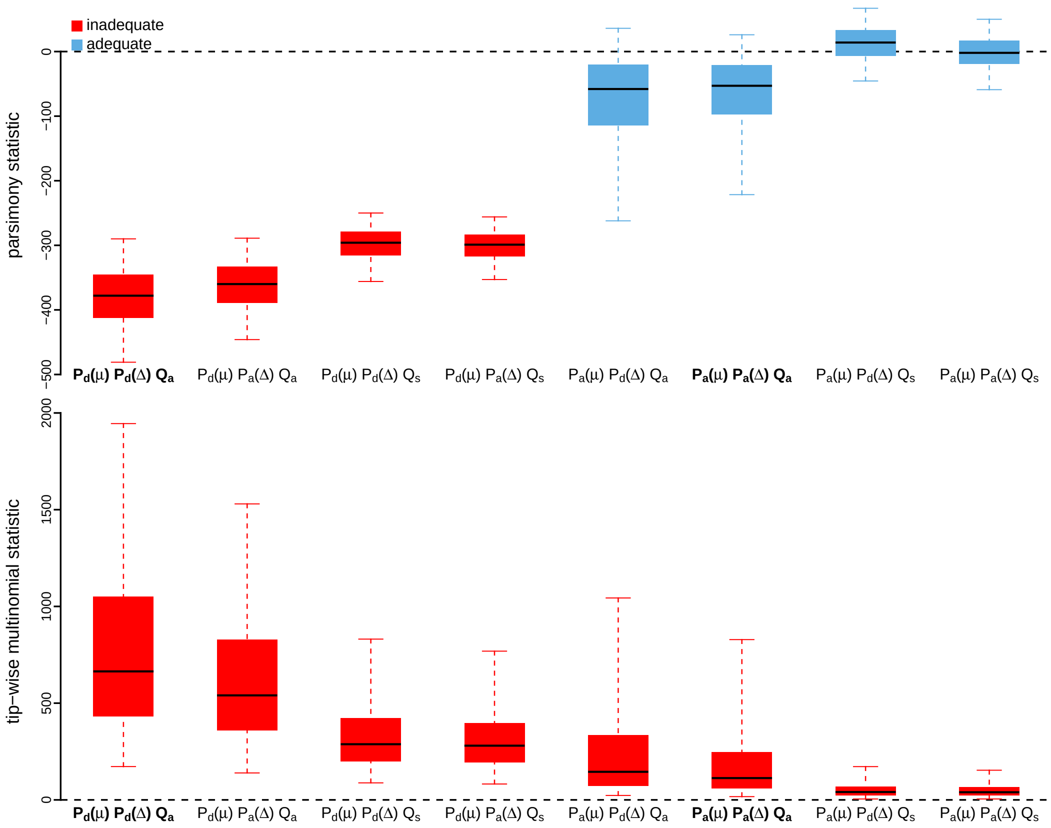


**Figure S101: Posterior-predictive distributions of the parsimony statistic (top panel) and the tip-wise multinomial statistic (bottom panel) under each of the eight prior models for Influenza A/H3N2 dataset.** Boxplots depict the posterior-predictive distributions of the statistic under each of the eight candidate prior models: the center of each box is the median predictive value of the summary statistic; the box and whiskers indicate the corresponding 50% and 95% posterior-predictive intervals, respectively. The horizontal dashed line indicates when the simulated and observed datasets produce identical value for the summary statistic. A model is judged to be inadequate (*i.e.*, incapable of generating geographic datasets that are similar to the observed data) if its 95% posterior-predictive interval does not overlap with the dashed line. Here the preferred default prior model and preferred alternative prior model are both inadequate. Notation for the candidate models is described in Table S13; the preferred default- and alternative-prior models are indicated in bold text.

*The Impact of Prior Choice on Average Dispersal Rate*

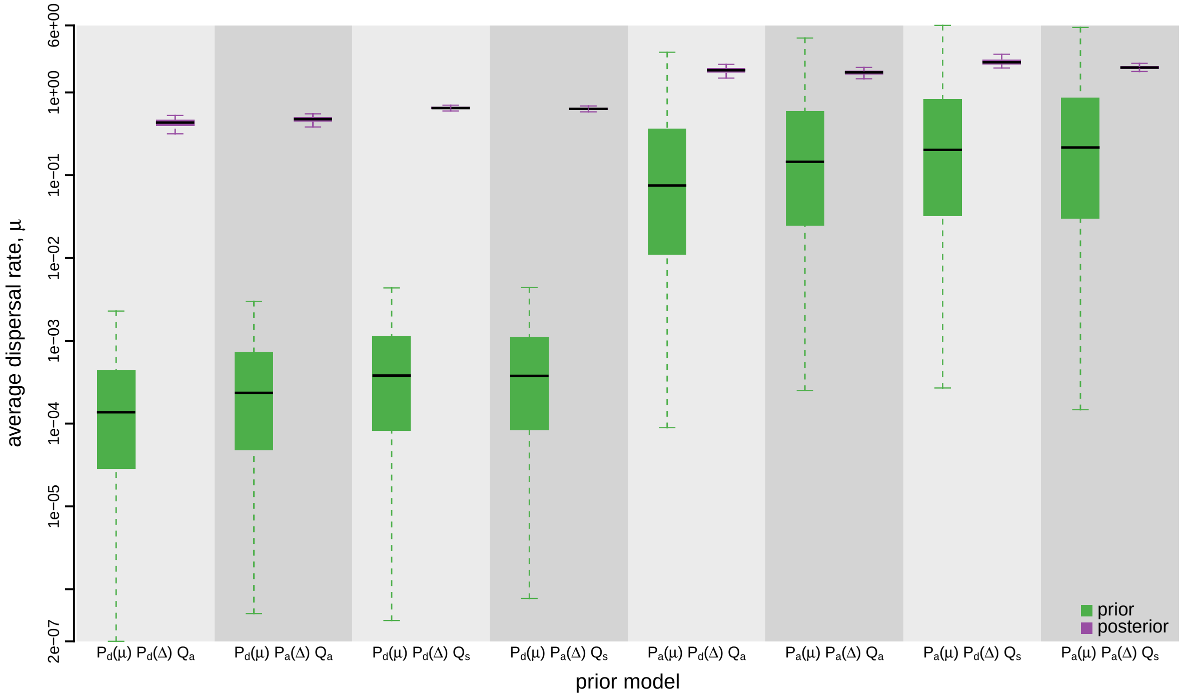


**Figure S102: The impact of prior choice on the average dispersal rate for Influenza A/H3N2 dataset.** Each pair of boxplots depicts the specified prior (green) and corresponding posterior estimate (purple) of the average dispersal rate, $\mu$, under each prior model: the center of each box indicates the median rate; the box and whiskers indicate the corresponding 50% and 95% credible intervals, respectively. Notation for the prior models is as described in Table S13.

*The Impact of Prior Choice on Pairwise Dispersal Rates*

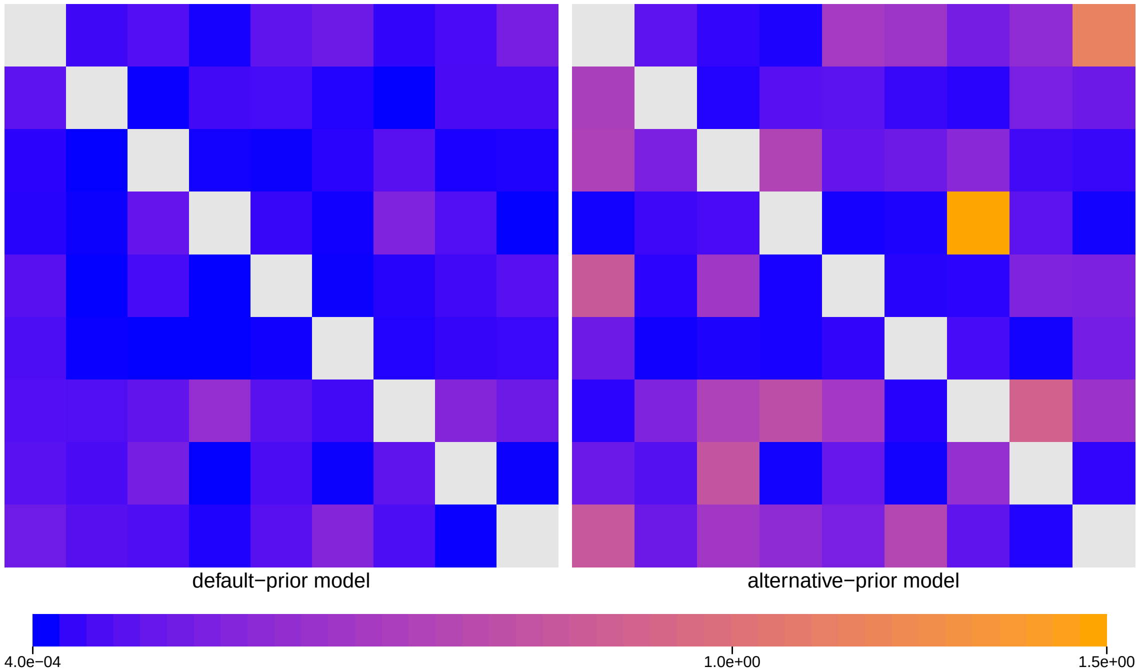


**Figure S103: The impact of prior choice on pairwise dispersal rates for Influenza A/H3N2 dataset.** Heatmaps summarize posterior-mean estimates of the instantaneous rate of dispersal between each pair of geographic areas, $q_{ij}$, under the default (left) and alternative (right) prior models.

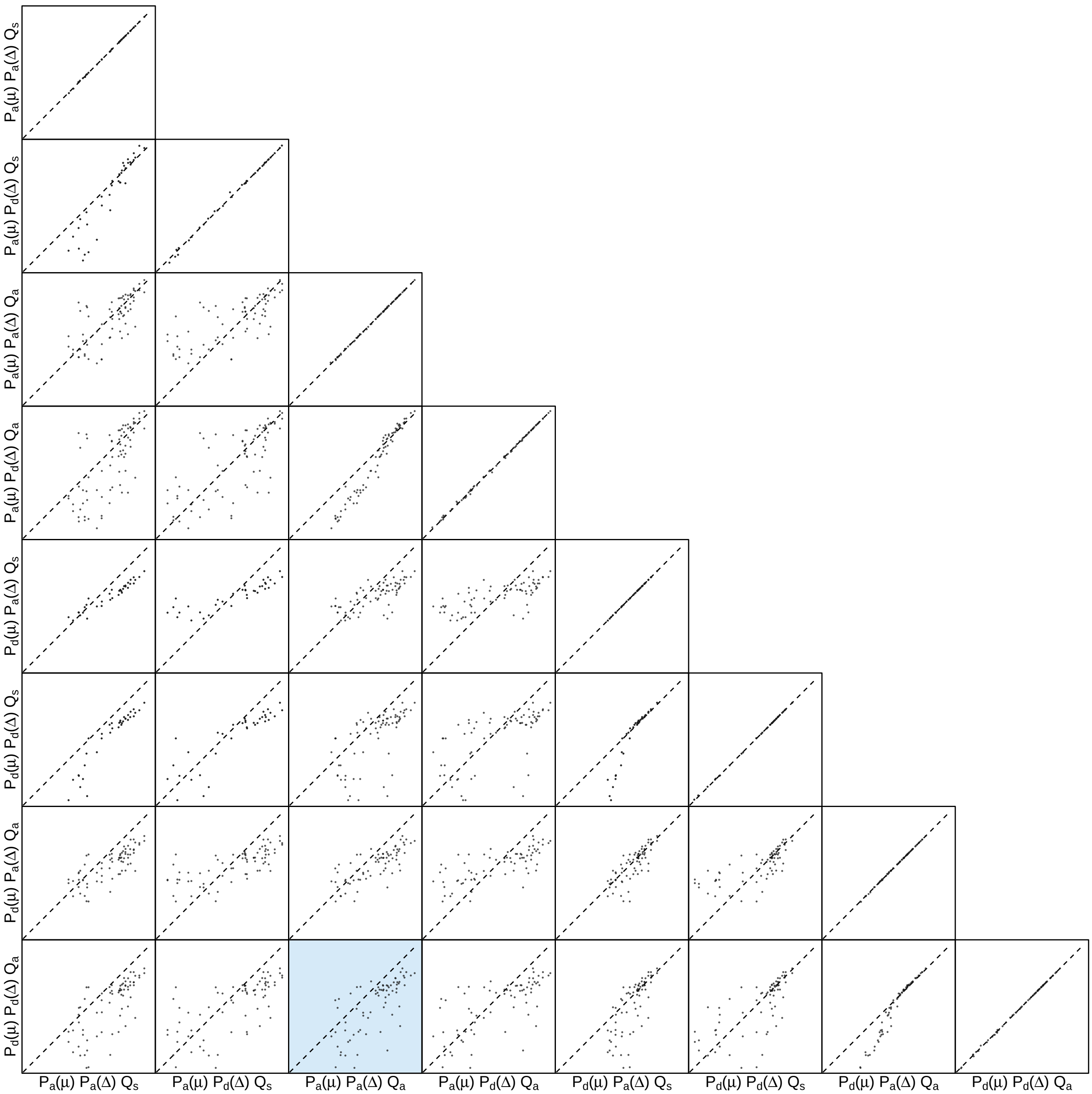


**Figure S104: The impact of prior choice on pairwise dispersal rates for Influenza A/H3N2 dataset.** Each cell of the plot compares posterior-mean estimate of the rate of dispersal between each pair of geographic areas, $q_{ij}$, between each pair of prior models. $x$- and $y$-axis of each cell are both on log scale. Diagonal cells are comparisons between half and the other half of the replicates under the same prior model, assessing the convergence of MCMC simulations; off-diagonal cells are comparisons between different prior models, demonstrating the impact of the prior model on the estimates of pairwise dispersal rates. The shaded cell corresponds to the comparison between the preferred default- and the preferred alternative-prior models (*c.f.*, Figure S103). Notation for the prior models is as described in Table S13.

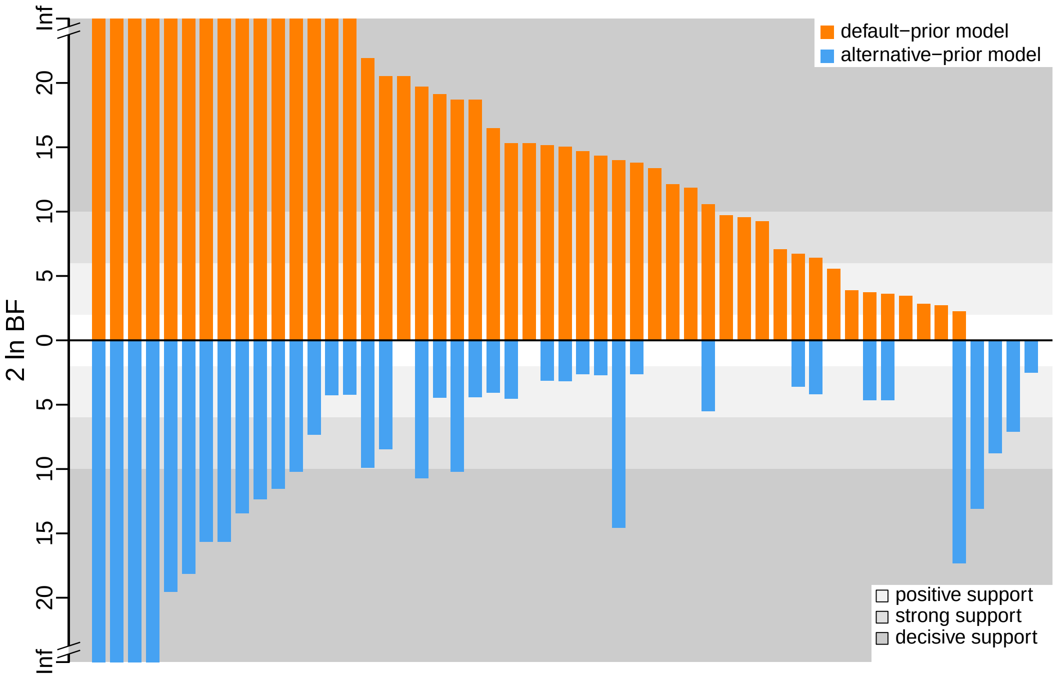


**Figure S105: The impact of prior choice on the inferred support for dispersal routes for Influenza A/H3N2 dataset.** We compare the evidential support for each dispersal route for Influenza A/H3N2 dataset under the default (orange) and alternative (blue) prior models. Each bar indicates the $2 \ln \mathrm{BF}$ for the corresponding dispersal route between two areas; only supported dispersal routes (*i.e.*, $2 \ln \mathrm{BF} > 2$) are plotted.

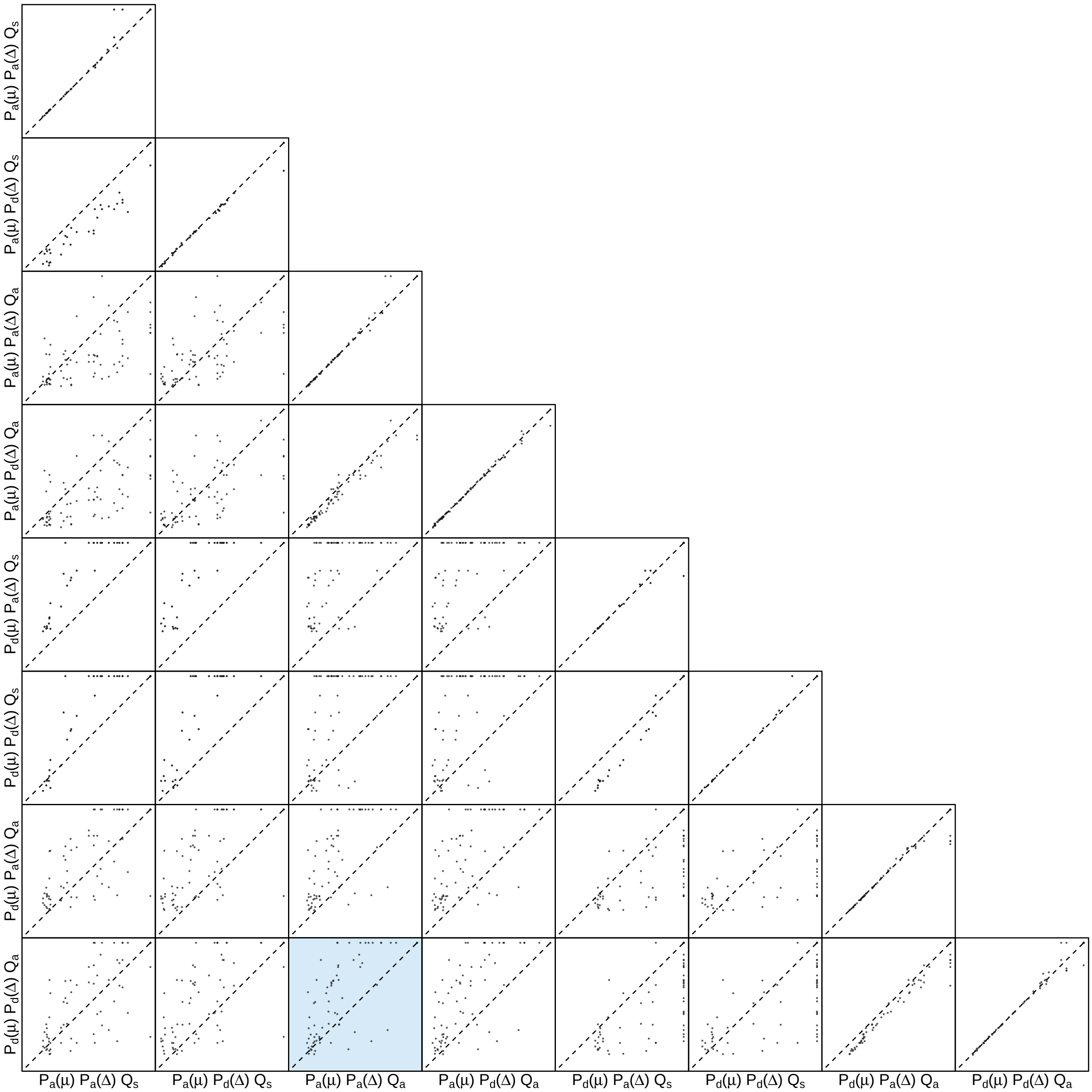


**Figure S106: The impact of prior choice on the inferred support for dispersal routes for Influenza A/H3N2 dataset.** Each cell of the plot compares the inferred support (2 ln BF) for pairwise dispersal routes between each pair of prior models. Diagonal cells are comparisons between half and the other half of the replicates under the same prior model, assessing the convergence of MCMC simulations; off-diagonal cells are comparisons between different prior models, demonstrating the impact of the prior model on the inferred support for pairwise dispersal routes. The shaded cell corresponds to the comparison between the preferred default- and the preferred alternative-prior models (*c.f.*, Figure S105). Notation for the prior models is as described in Table S13.

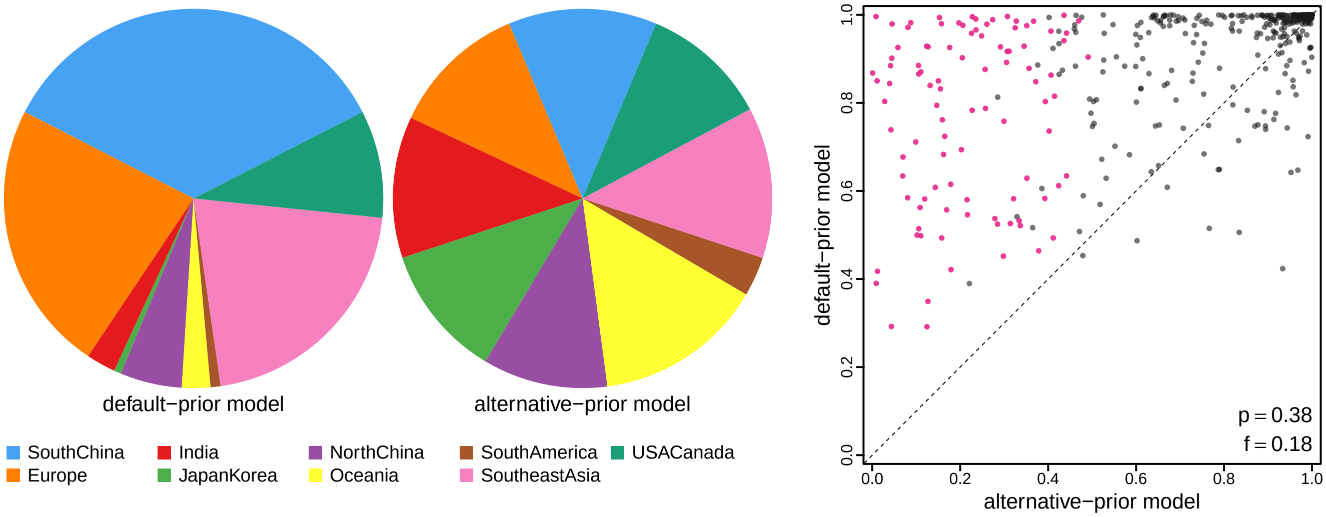


**Figure S107: The impact of prior choice on ancestral-area estimates for Influenza A/H3N2 dataset.** The left panel compares the posterior probability of each ancestral area at the root node under the default- and alternative-prior models for Influenza A/H3N2 dataset. The right panel plots the posterior probability of the most probable ancestral area under the default-prior model for each node in the MCC tree (y-axis) against the corresponding posterior probability of that area under the alternative-prior model (x-axis). Pink dots represent the internal nodes where the MAP ancestral area inferred under the default-prior model differs from that inferred under the alternative-prior models. The statistic $p$ denotes the fraction of internal nodes that are shared under the default- and alternative-prior models; $f$, is the fraction of shared nodes where the MAP ancestral area differs under the default- and alternative-prior models. Note that the posterior probabilities of the MAP ancestral area under the default-prior model are generally higher than those under the alternative-prior model (*i.e.*, the default-prior model tends to mask uncertainty in the ancestral-area estimates).

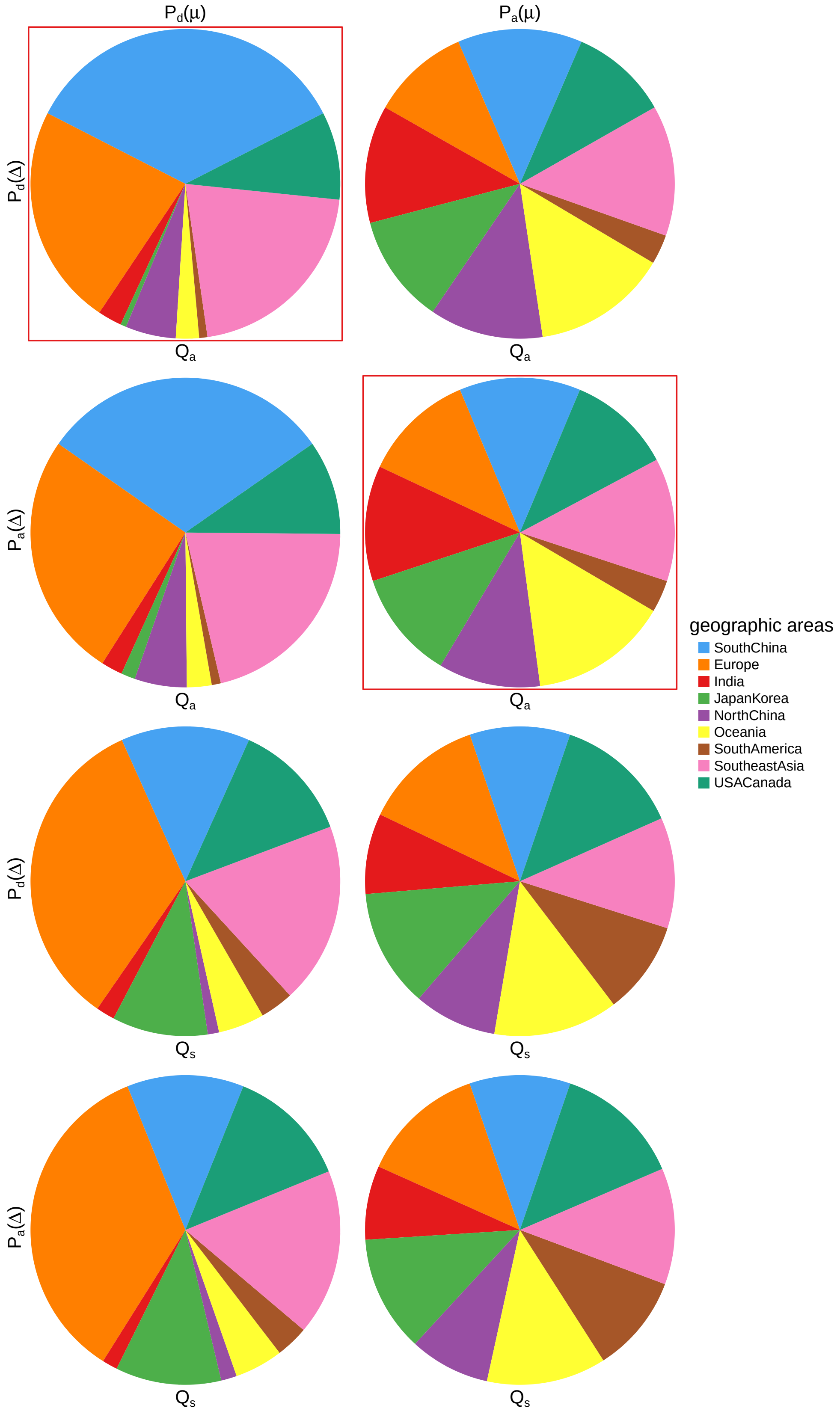


**Figure S108: The impact of prior choice on ancestral-area estimates at the root node for Influenza A/H3N2 dataset.** The posterior probability of each ancestral area at the root node under each prior model for Influenza A/H3N2 dataset. The pie chart enclosed in red corresponds to the comparison between the preferred default- and the preferred alternative-prior models (*c.f.*, Figure S107, left panel). Model notation follows the description in Table S13.

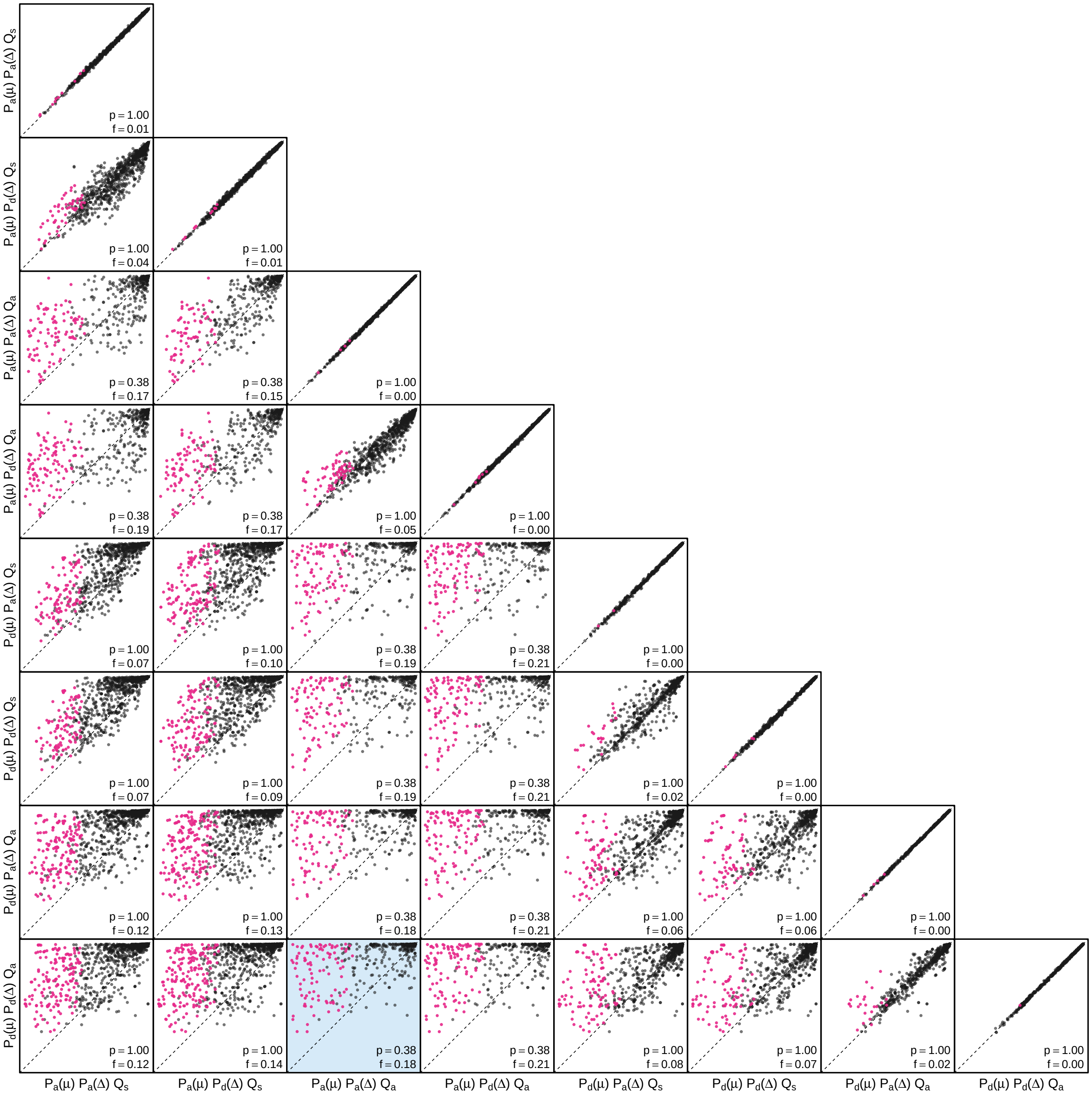


**Figure S109: The impact of prior choice on the MAP ancestral-area estimates at internal nodes for Influenza A/H3N2 dataset.** Pairwise scatter plots compare the posterior probability of the MAP ancestral area inferred under each combination of prior models. Diagonal cells are comparisons between two replicates under the same prior model (assessing the convergence of MCMC simulations); off-diagonal cells are comparisons between different prior models, demonstrating the impact of the prior model on both the posterior probability and the identity of the MAP ancestral-area estimates at internal nodes. Pink dots represent the internal nodes where the MAP ancestral area inferred under the default-prior model differs from that inferred under the alternative-prior models. The statistic $p$ denotes the fraction of internal nodes that are shared under the default- and alternative-prior models; $f$ is the fraction of shared nodes where the MAP ancestral area differs under the default- and alternative-prior models. The shaded cell corresponds to the comparison between the preferred default- and the preferred alternative-prior models (*c.f.*, Figure S107, left panel). Model notation follows the description in Table S13.

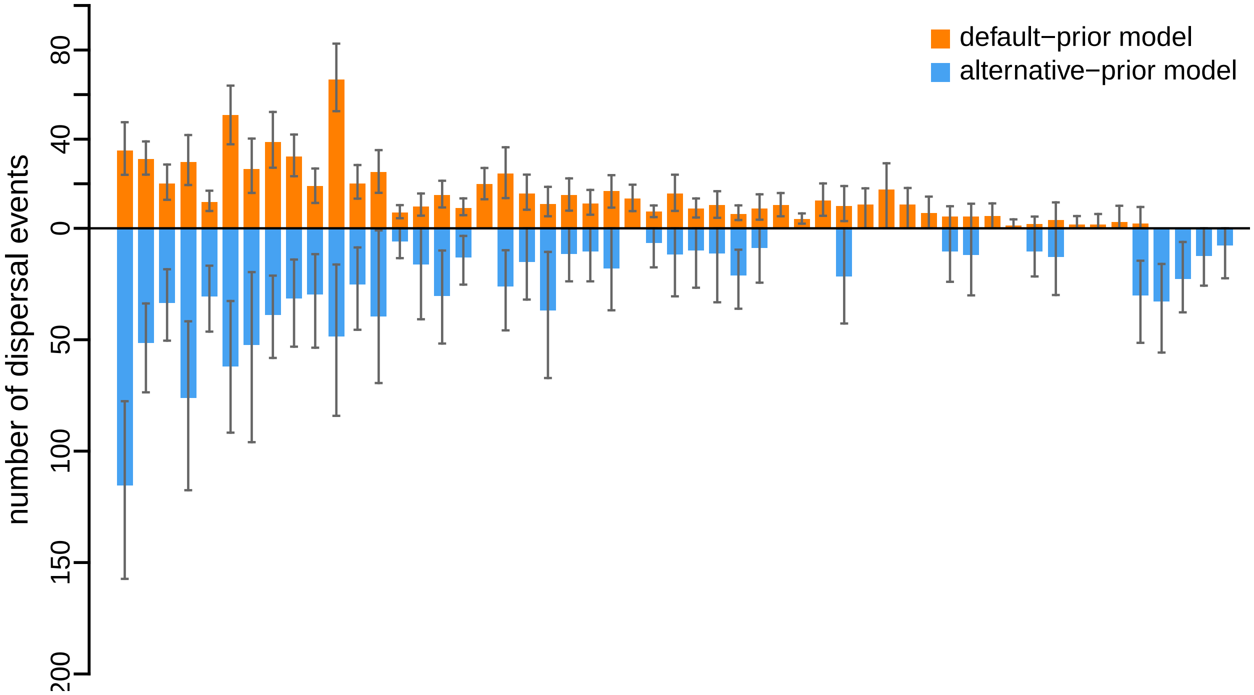


**Figure S110: The impact of prior choice on the inferred number of dispersal events between each pair of areas for Influenza A/H3N2 dataset.** The reflected bar plot depicts the number of dispersal events inferred under the default (orange) and alternative (blue) prior models for Influenza A/H3N2 dataset. Each bar indicates the posterior-mean number of dispersal events between a pair of areas; whiskers indicate the 95% credible interval. Note that only the number of dispersal events over the "significant" dispersal routes (*i.e.*, $2 \ln \mathrm{BF} > 2$) are figured.

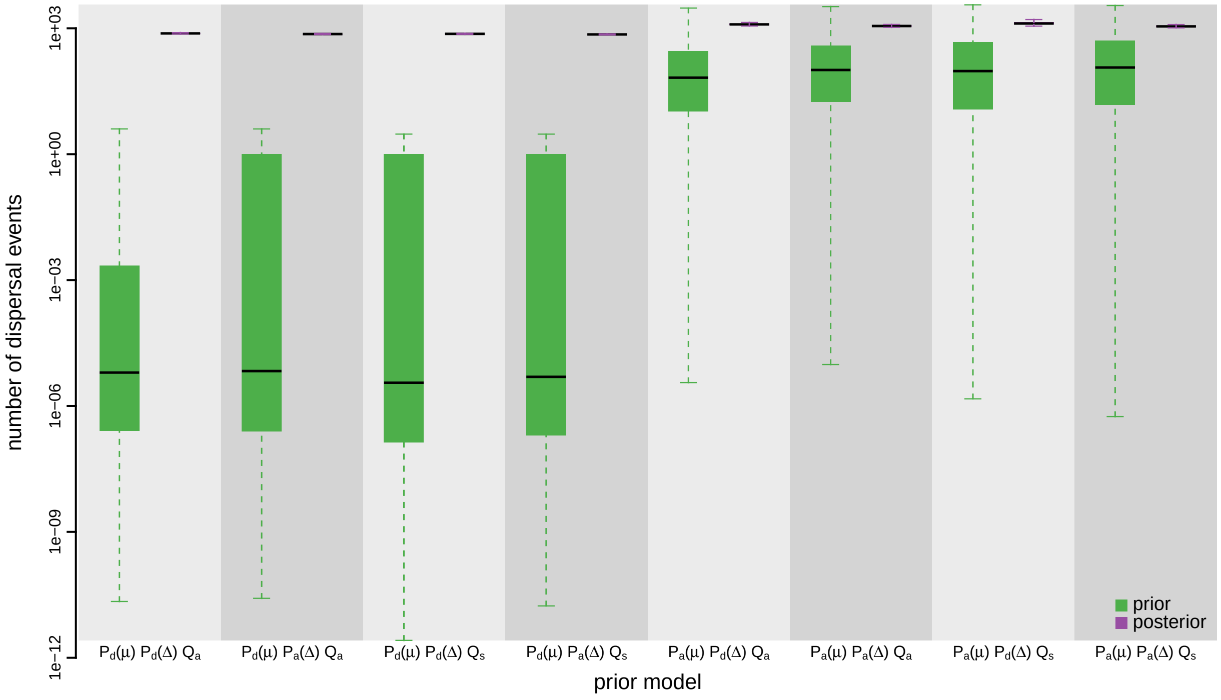


**Figure S111: The impact of prior choice on the inferred total number of dispersal events between all areas for Influenza A/H3N2 dataset.** Each pair of boxplots depicts the specified prior (green) and corresponding posterior estimate (purple) of the number of dispersal events between all areas under each prior model: the center of each box indicates the posterior-median number of dispersal events; the box and whiskers indicate the corresponding 50% and 95% credible intervals, respectively. Model notation follows the description in Table S13.

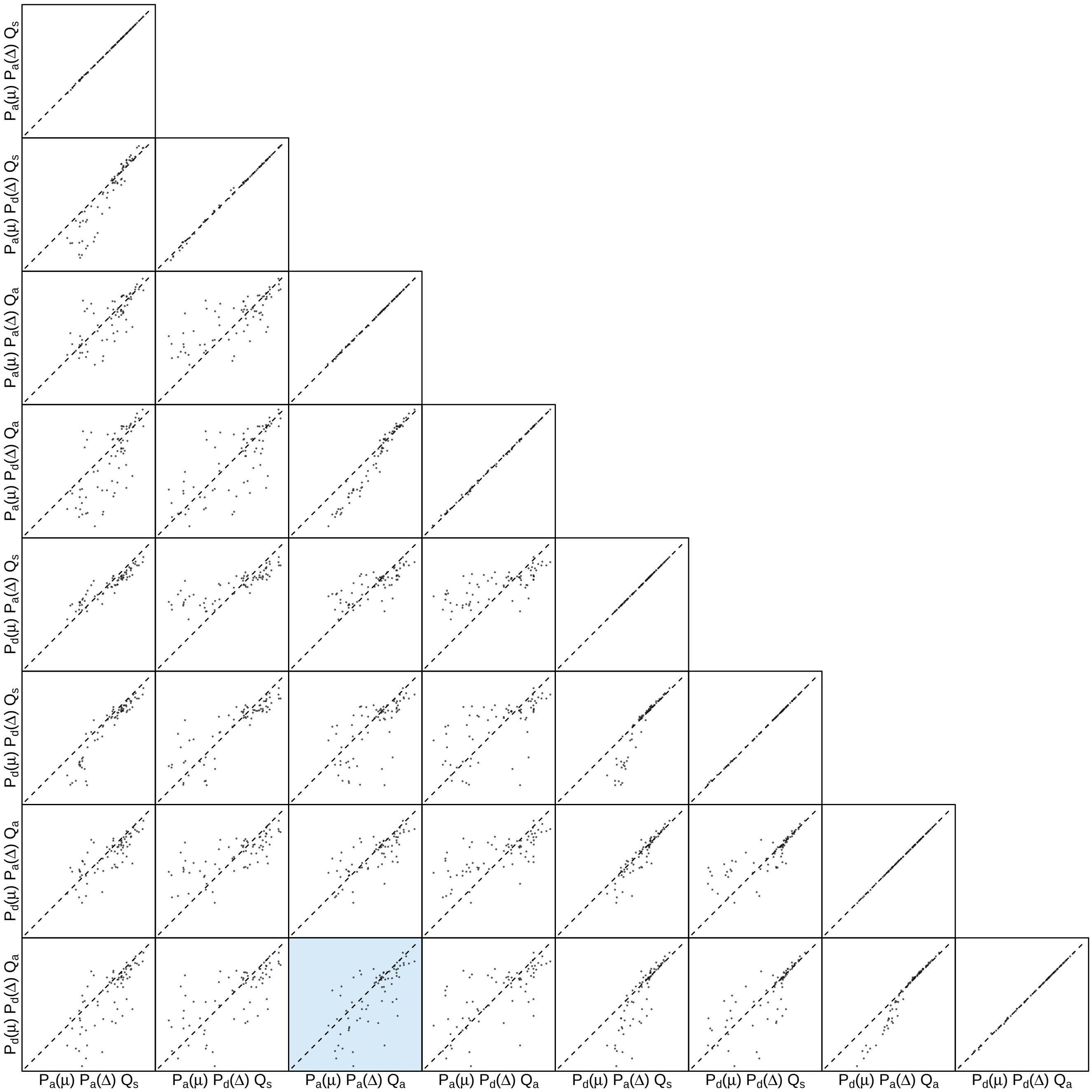


**Figure S112: The impact of prior choice on the inferred number of dispersal events between each pair of areas for Influenza A/H3N2 dataset.** Each cell of the plot compares the inferred number of pairwise dispersal events between each pair of prior models. *x*- and *y*-axis of each cell are both on log scale. Diagonal cells are comparisons between half and the other half of the replicates under the same prior model, assessing the convergence of MCMC simulations; off-diagonal cells are comparisons between different prior models, demonstrating the impact of the prior model on the inferred number of pairwise dispersal events. The shaded cell corresponds to the comparison between the preferred default- and the preferred alternative-prior models (*c.f.*, Figure S110). Model notation follows the description in Table S13.

###### S3.3.4.2 B/Yamagata Dataset

*The Impact of Prior Choice on Biogeographic Model Fit*

**Table S14: Marginal-likelihood estimates of each of the eight prior models for Influenza B/Yamagata dataset.** Columns 2–5 list marginal likelihoods inferred from four replicate analyses. Columns 6–7 list the mean and standard deviation of these marginal-likelihood estimates. The last column lists the marginal-likelihood estimates computed by combining the samples from the replicate power-posterior MCMC simulations. Candidate models are listed in rows and include all possible combinations of: (1) instantaneous-rate matrices (symmetric, $Q_s$ or asymmetric, $Q_a$); (2) priors on the average dispersal rate [default, $P_d(\mu)$ or alternative, $P_a(\mu)$], and; (3) priors on the number of dispersal routes [default, $P_d(\Delta)$ or alternative, $P_a(\Delta)$]. The preferred default- and alternative-prior models are indicated in bold text.

| Model | replicate1 | replicate2 | replicate3 | replicate4 | mean | sd | combined |
|---|---|---|---|---|---|---|---|
| $\mathbf{P_d(\mu) Q_a P_d(\Delta)}$ | **-2333.12** | **-2334.61** | **-2333.34** | **-2334.12** | **-2333.80** | **0.69** | **-2333.48** |
| $P_d(\mu) Q_a P_a(\Delta)$ | -2293.25 | -2293.21 | -2293.48 | -2293.10 | -2293.26 | 0.16 | -2293.25 |
| $P_d(\mu) Q_s P_d(\Delta)$ | -2351.22 | -2351.73 | -2351.95 | -2351.83 | -2351.68 | 0.32 | -2351.56 |
| $P_d(\mu) Q_s P_a(\Delta)$ | -2308.93 | -2308.12 | -2308.29 | -2307.89 | -2308.31 | 0.45 | -2308.11 |
| $P_a(\mu) Q_a P_d(\Delta)$ | -1897.70 | -1897.89 | -1897.71 | -1897.45 | -1897.69 | 0.18 | -1897.68 |
| $\mathbf{P_a(\mu) Q_a P_a(\Delta)}$ | **-1872.89** | **-1872.90** | **-1872.85** | **-1872.88** | **-1872.88** | **0.02** | **-1872.87** |
| $P_a(\mu) Q_s P_d(\Delta)$ | -1929.77 | -1930.08 | -1930.90 | -1930.34 | -1930.27 | 0.48 | -1930.24 |
| $P_a(\mu) Q_s P_a(\Delta)$ | -1898.25 | -1898.70 | -1898.84 | -1898.62 | -1898.60 | 0.25 | -1898.59 |

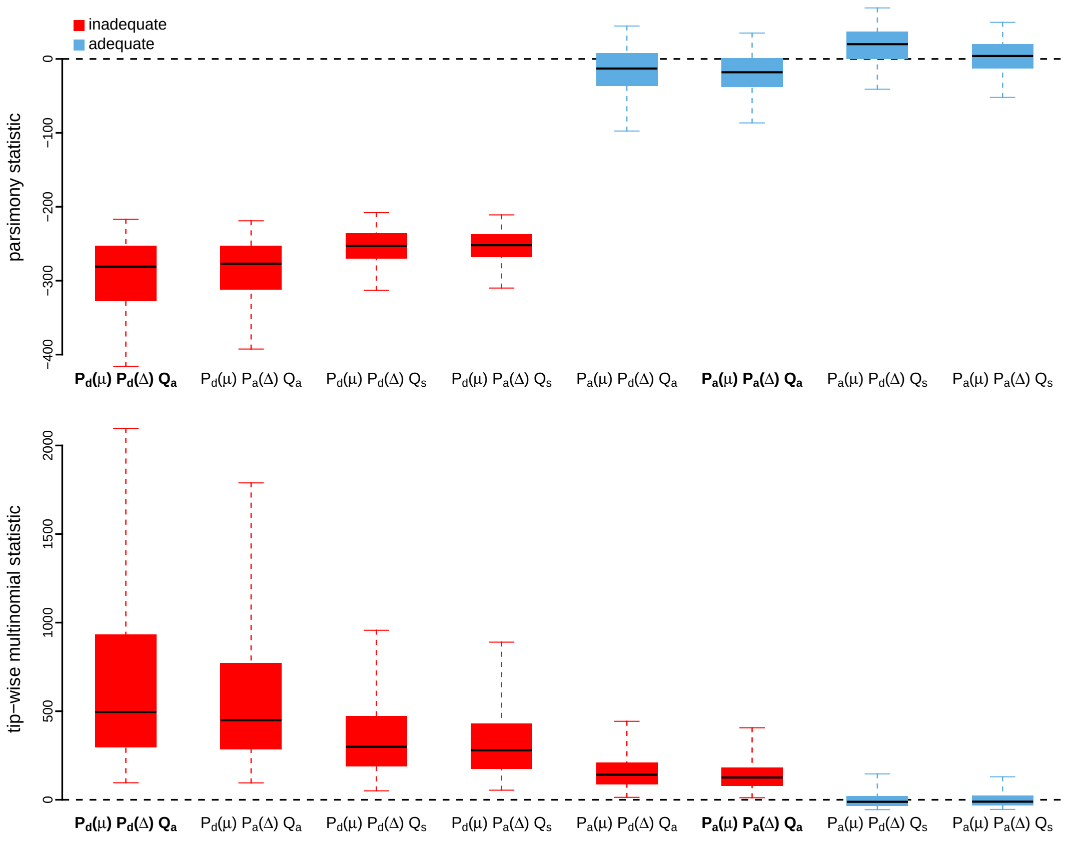


**Figure S113: Posterior-predictive distributions of the parsimony statistic (top panel) and the tip-wise multinomial statistic (bottom panel) under each of the eight prior models for Influenza B/Yamagata dataset.** Boxplots depict the posterior-predictive distributions of the statistic under each of the eight candidate prior models: the center of each box is the median predictive value of the summary statistic; the box and whiskers indicate the corresponding 50% and 95% posterior-predictive intervals, respectively. The horizontal dashed line indicates when the simulated and observed datasets produce identical value for the summary statistic. A model is judged to be inadequate (*i.e.*, incapable of generating geographic datasets that are similar to the observed data) if its 95% posterior-predictive interval does not overlap with the dashed line. Here the preferred default prior model and preferred alternative prior model are both inadequate. Notation for the candidate models is described in Table S14; the preferred default- and alternative-prior models are indicated in bold text.

*The Impact of Prior Choice on Average Dispersal Rate*

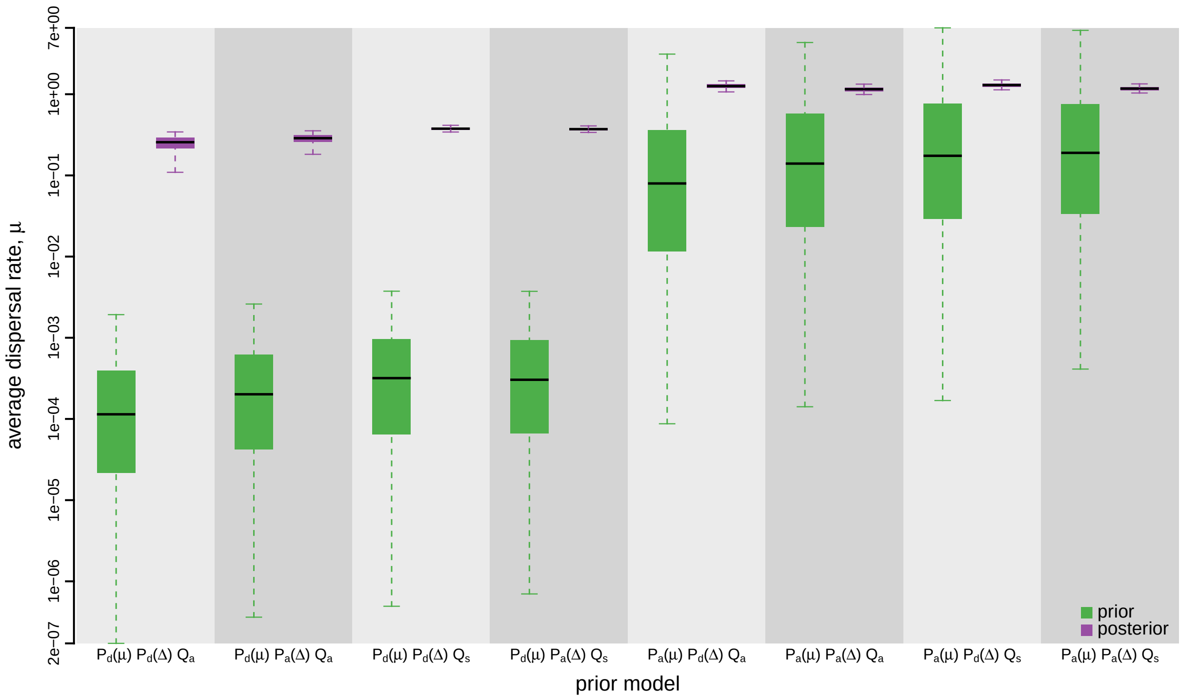


**Figure S114: The impact of prior choice on the average dispersal rate for Influenza B/Yamagata dataset.** Each pair of boxplots depicts the specified prior (green) and corresponding posterior estimate (purple) of the average dispersal rate, $\mu$, under each prior model: the center of each box indicates the median rate; the box and whiskers indicate the corresponding 50% and 95% credible intervals, respectively. Notation for the prior models is as described in Table S14.

*The Impact of Prior Choice on Pairwise Dispersal Rates*

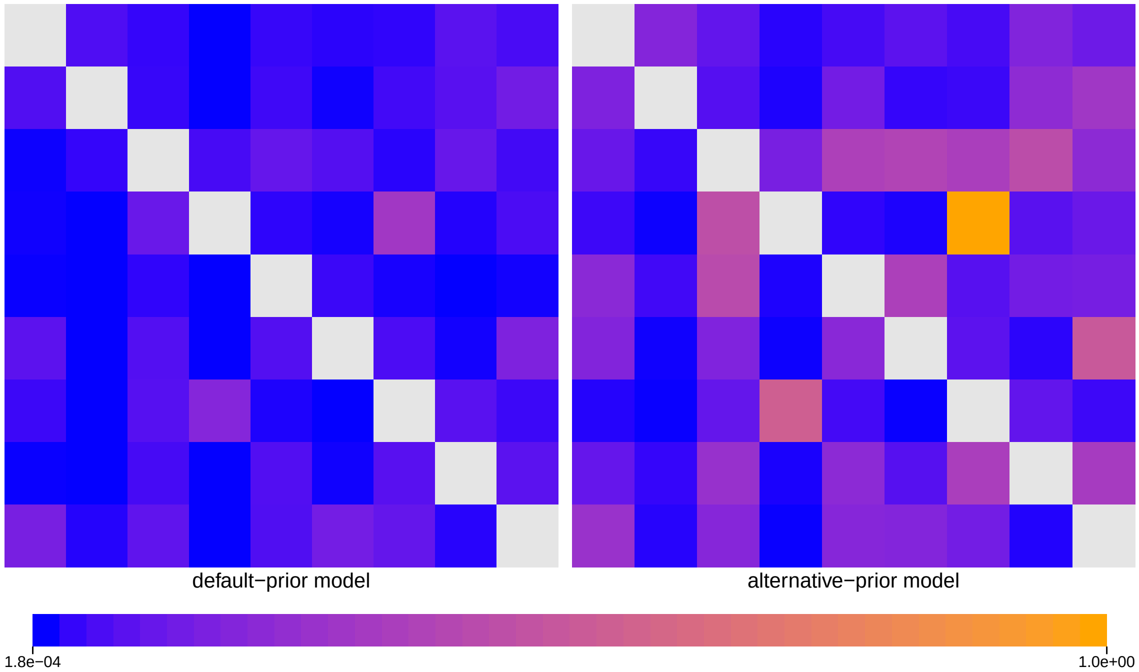


**Figure S115: The impact of prior choice on pairwise dispersal rates for Influenza B/Yamagata dataset.** Heatmaps summarize posterior-mean estimates of the instantaneous rate of dispersal between each pair of geographic areas, $q_{ij}$, under the default (left) and alternative (right) prior models.

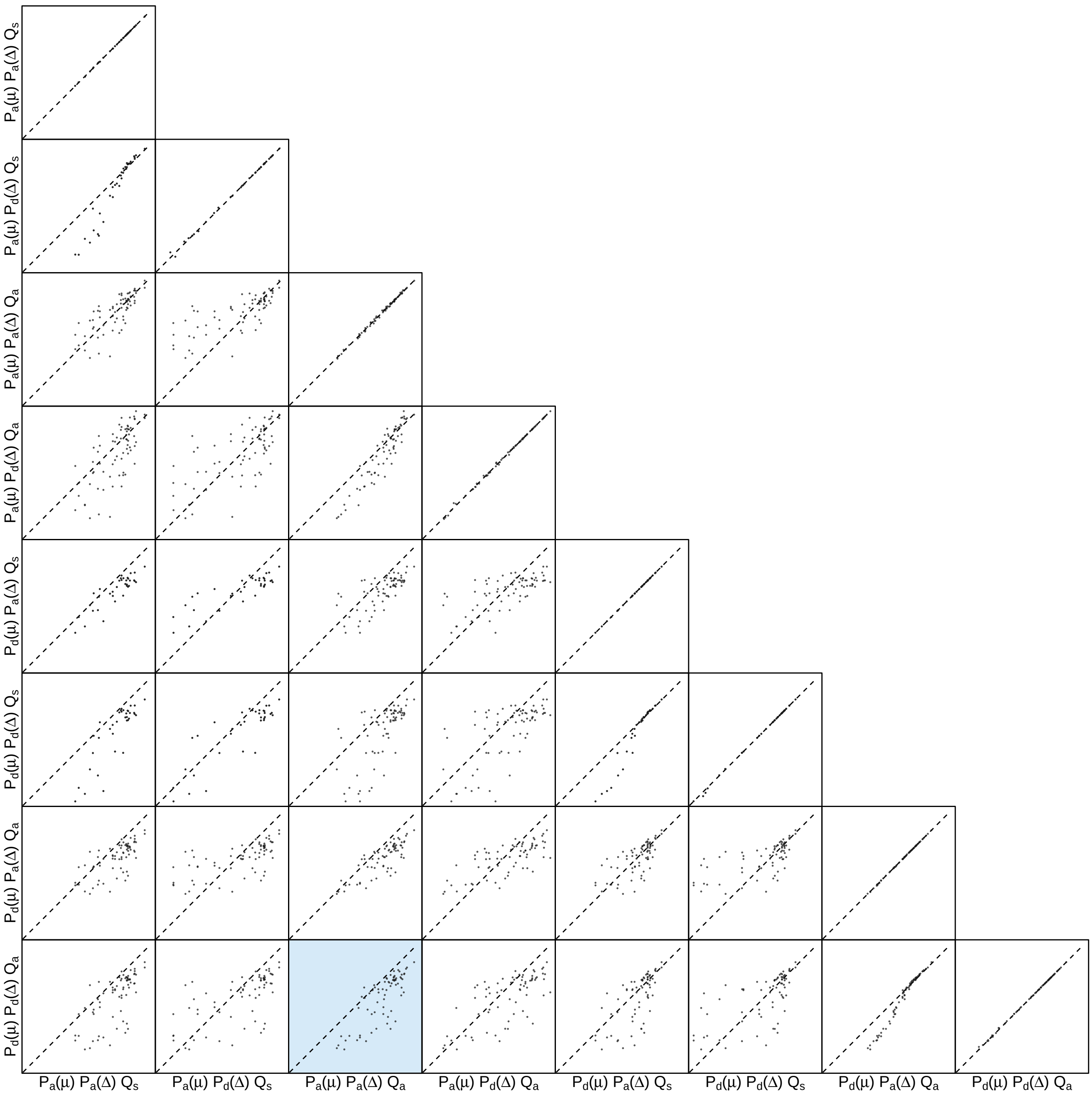


**Figure S116: The impact of prior choice on pairwise dispersal rates for Influenza B/Yamagata dataset.** Each cell of the plot compares posterior-mean estimate of the rate of dispersal between each pair of geographic areas, $q_{ij}$, between each pair of prior models. $x$- and $y$-axis of each cell are both on log scale. Diagonal cells are comparisons between half and the other half of the replicates under the same prior model, assessing the convergence of MCMC simulations; off-diagonal cells are comparisons between different prior models, demonstrating the impact of the prior model on the estimates of pairwise dispersal rates. The shaded cell corresponds to the comparison between the preferred default- and the preferred alternative-prior models (*c.f.*, Figure S115). Notation for the prior models is as described in Table S14.

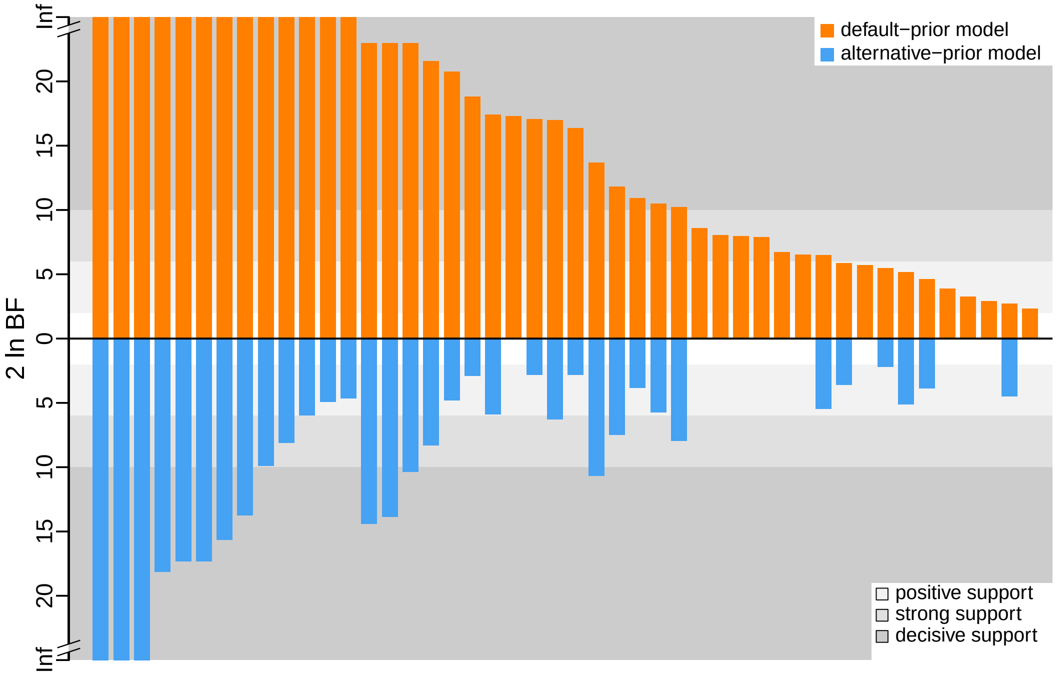


**Figure S117: The impact of prior choice on the inferred support for dispersal routes for Influenza B/Yamagata dataset.** We compare the evidential support for each dispersal route for Influenza B/Yamagata dataset under the default (orange) and alternative (blue) prior models. Each bar indicates the $2 \ln \mathrm{BF}$ for the corresponding dispersal route between two areas; only supported dispersal routes (*i.e.*, $2 \ln \mathrm{BF} > 2$) are plotted.

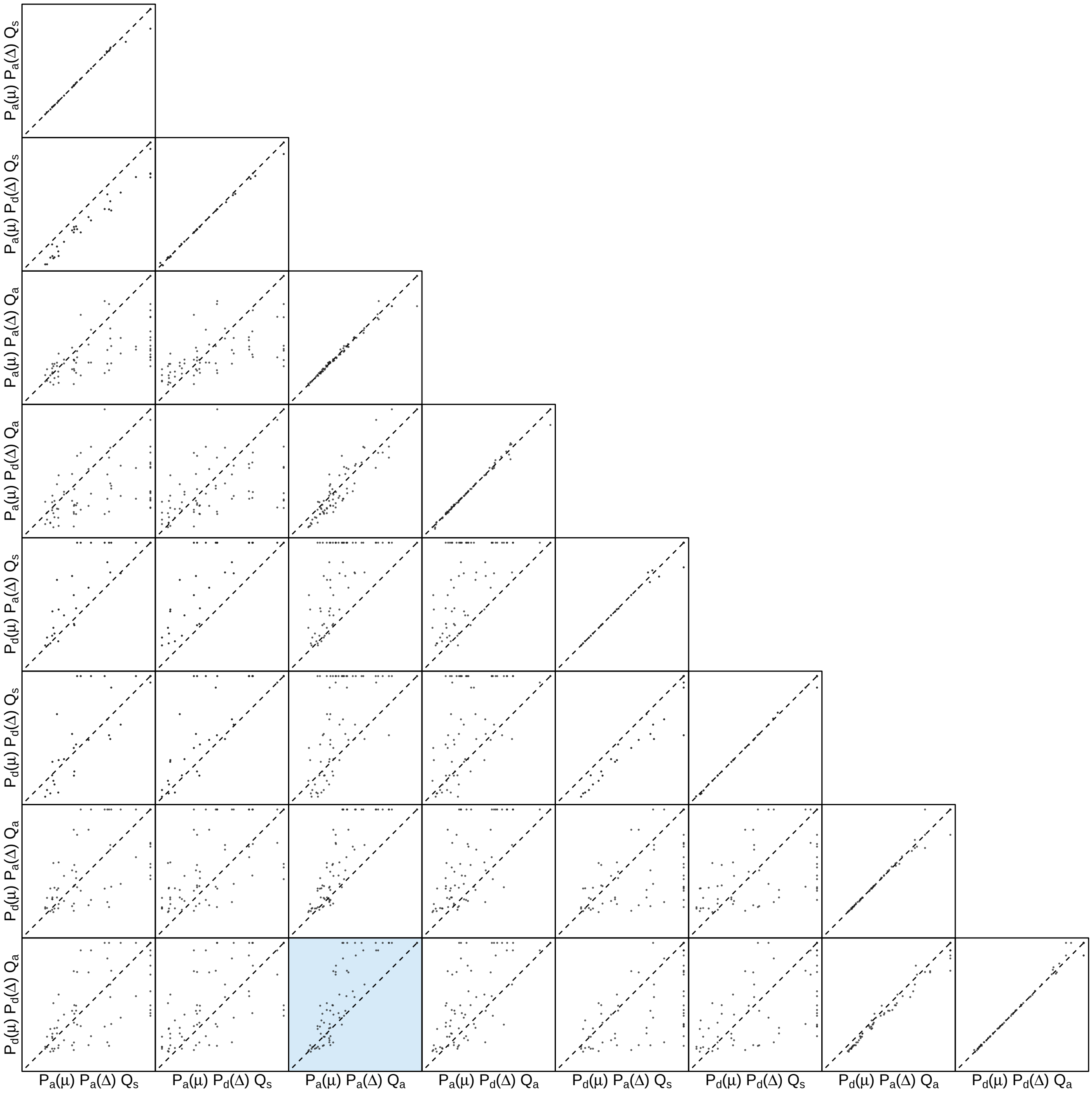


**Figure S118: The impact of prior choice on the inferred support for dispersal routes for Influenza B/Yamagata dataset.** Each cell of the plot compares the inferred support ($2 \ln \mathrm{BF}$) for pairwise dispersal routes between each pair of prior models. Diagonal cells are comparisons between half and the other half of the replicates under the same prior model, assessing the convergence of MCMC simulations; off-diagonal cells are comparisons between different prior models, demonstrating the impact of the prior model on the inferred support for pairwise dispersal routes. The shaded cell corresponds to the comparison between the preferred default- and the preferred alternative-prior models (*c.f.*, Figure S117). Notation for the prior models is as described in Table S14.

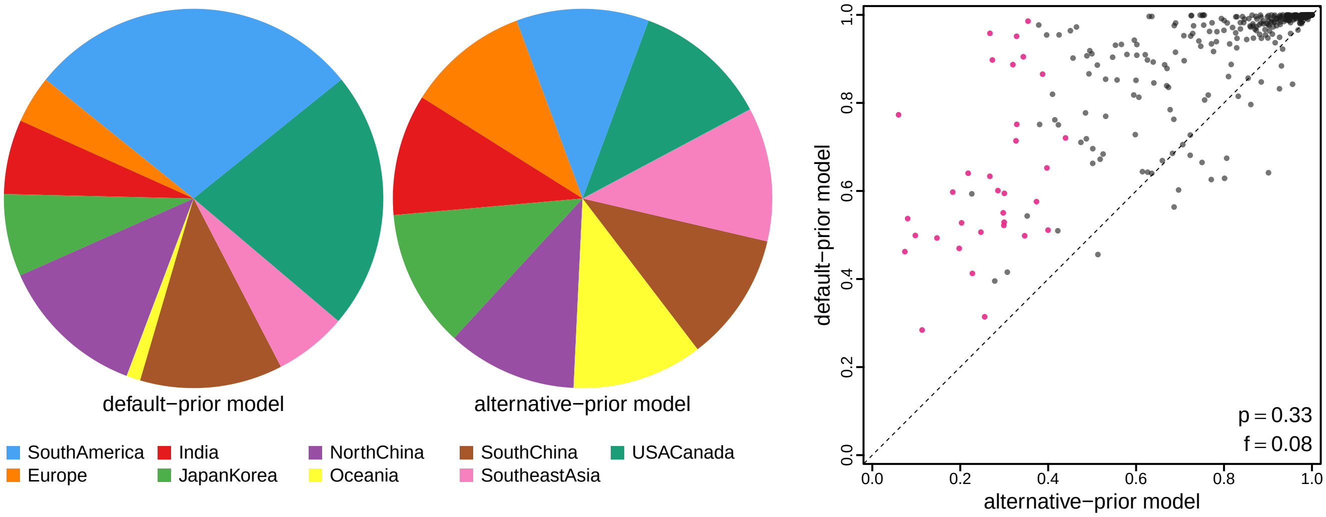


**Figure S119: The impact of prior choice on ancestral-area estimates for Influenza B/Yamagata dataset.** The left panel compares the posterior probability of each ancestral area at the root node under the default- and alternative-prior models for Influenza B/Yamagata dataset. The right panel plots the posterior probability of the most probable ancestral area under the default-prior model for each node in the MCC tree (y-axis) against the corresponding posterior probability of that area under the alternative-prior model (x-axis). Pink dots represent the internal nodes where the MAP ancestral area inferred under the default-prior model differs from that inferred under the alternative-prior models. The statistic $p$ denotes the fraction of internal nodes that are shared under the default- and alternative-prior models; $f$, is the fraction of shared nodes where the MAP ancestral area differs under the default- and alternative-prior models. Note that the posterior probabilities of the MAP ancestral area under the default-prior model are generally higher than those under the alternative-prior model (*i.e.*, the default-prior model tends to mask uncertainty in the ancestral-area estimates).

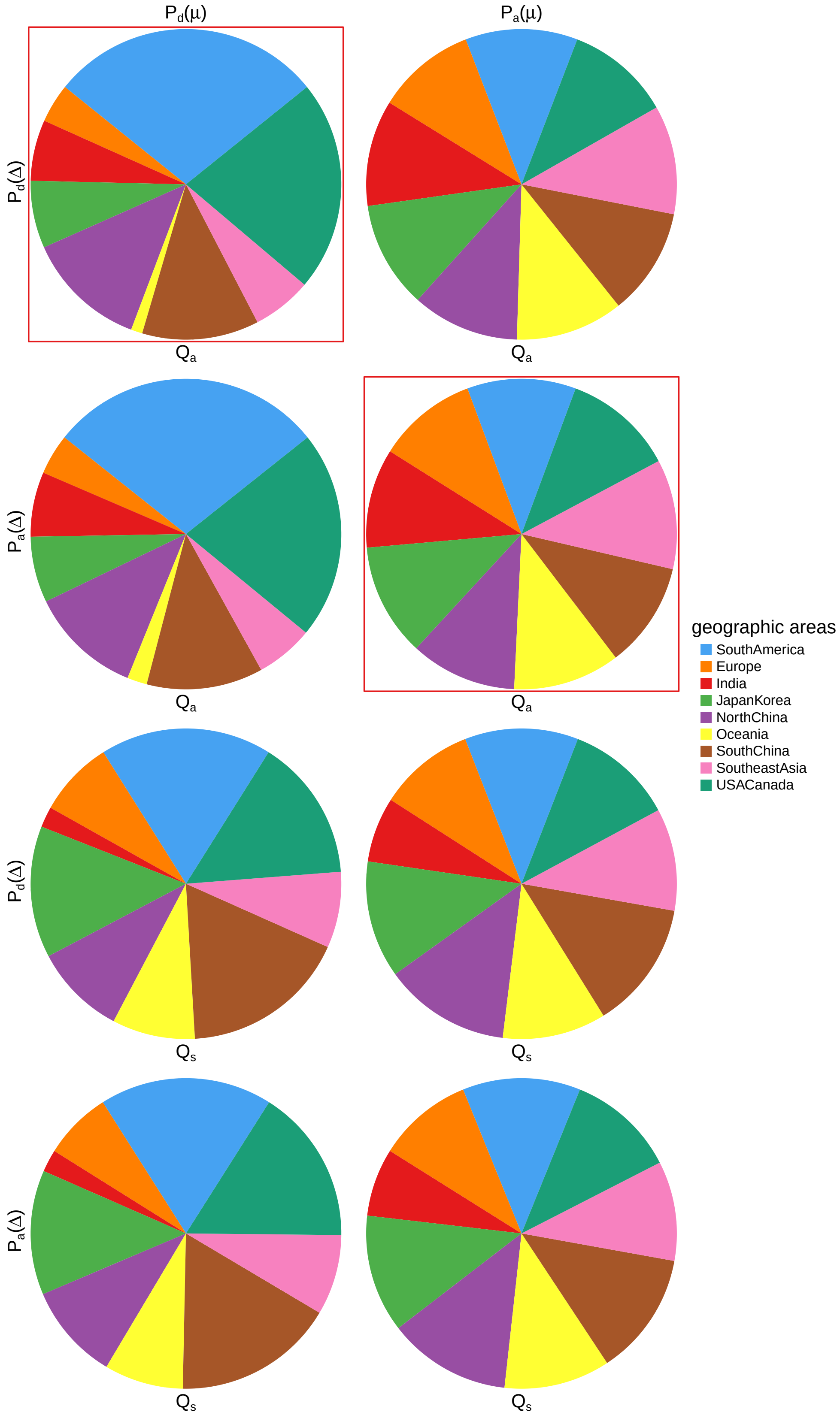


**Figure S120: The impact of prior choice on ancestral-area estimates at the root node for Influenza B/Yamagata dataset.** The posterior probability of each ancestral area at the root node under each prior model for Influenza B/Yamagata dataset. The pie chart enclosed in red corresponds to the comparison between the preferred default- and the preferred alternative-prior models (*c.f.*, Figure S119, left panel). Model notation follows the description in Table S14.

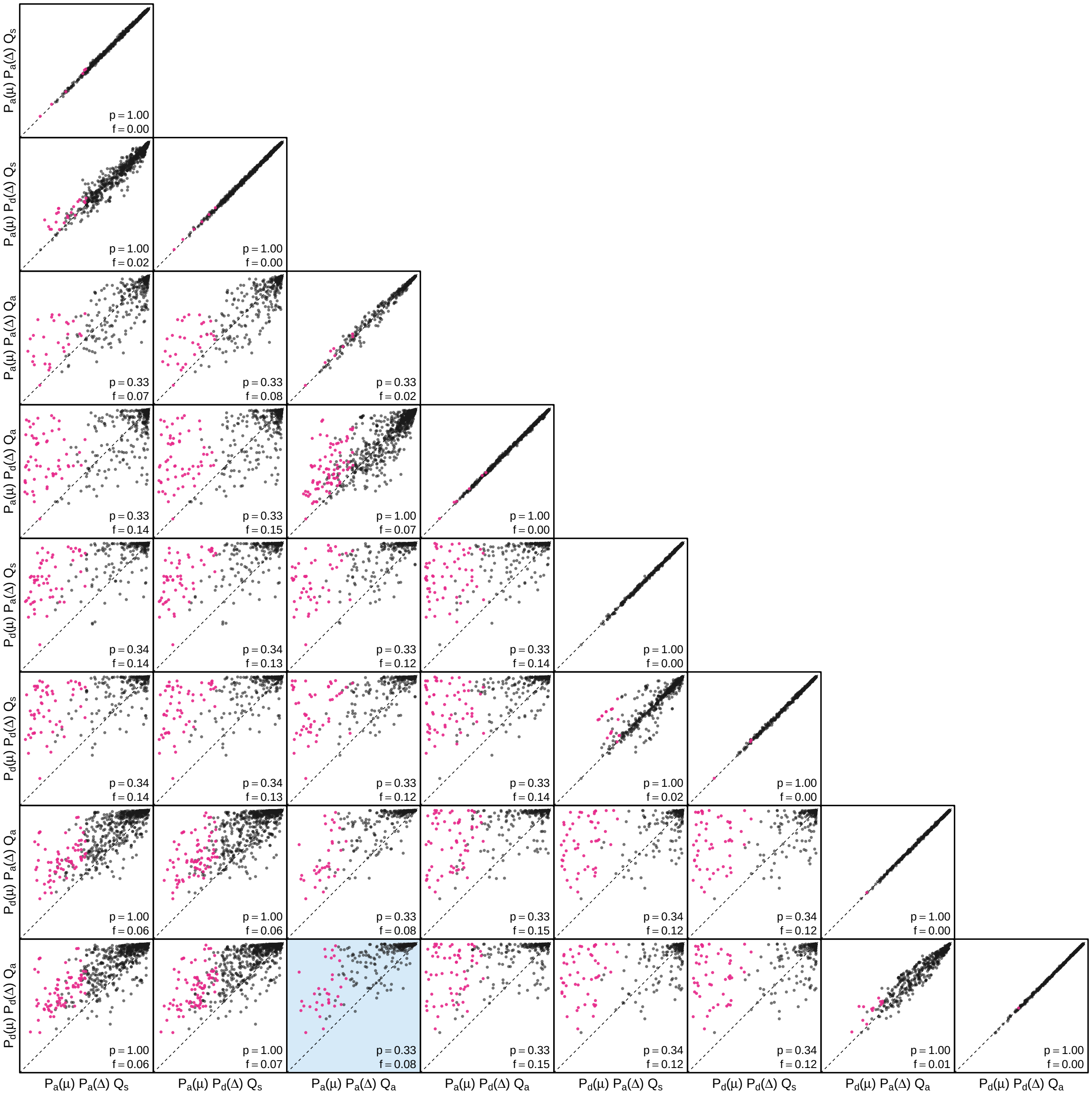


**Figure S121: The impact of prior choice on the MAP ancestral-area estimates at internal nodes for Influenza B/Yamagata dataset.** Pairwise scatter plots compare the posterior probability of the MAP ancestral area inferred under each combination of prior models. Diagonal cells are comparisons between two replicates under the same prior model (assessing the convergence of MCMC simulations); off-diagonal cells are comparisons between different prior models, demonstrating the impact of the prior model on both the posterior probability and the identity of the MAP ancestral-area estimates at internal nodes. Pink dots represent the internal nodes where the MAP ancestral area inferred under the default-prior model differs from that inferred under the alternative-prior models. The statistic $p$ denotes the fraction of internal nodes that are shared under the default- and alternative-prior models; $f$ is the fraction of shared nodes where the MAP ancestral area differs under the default- and alternative-prior models. The shaded cell corresponds to the comparison between the preferred default- and the preferred alternative-prior models (*c.f.*, Figure S119, left panel). Model notation follows the description in Table S14.

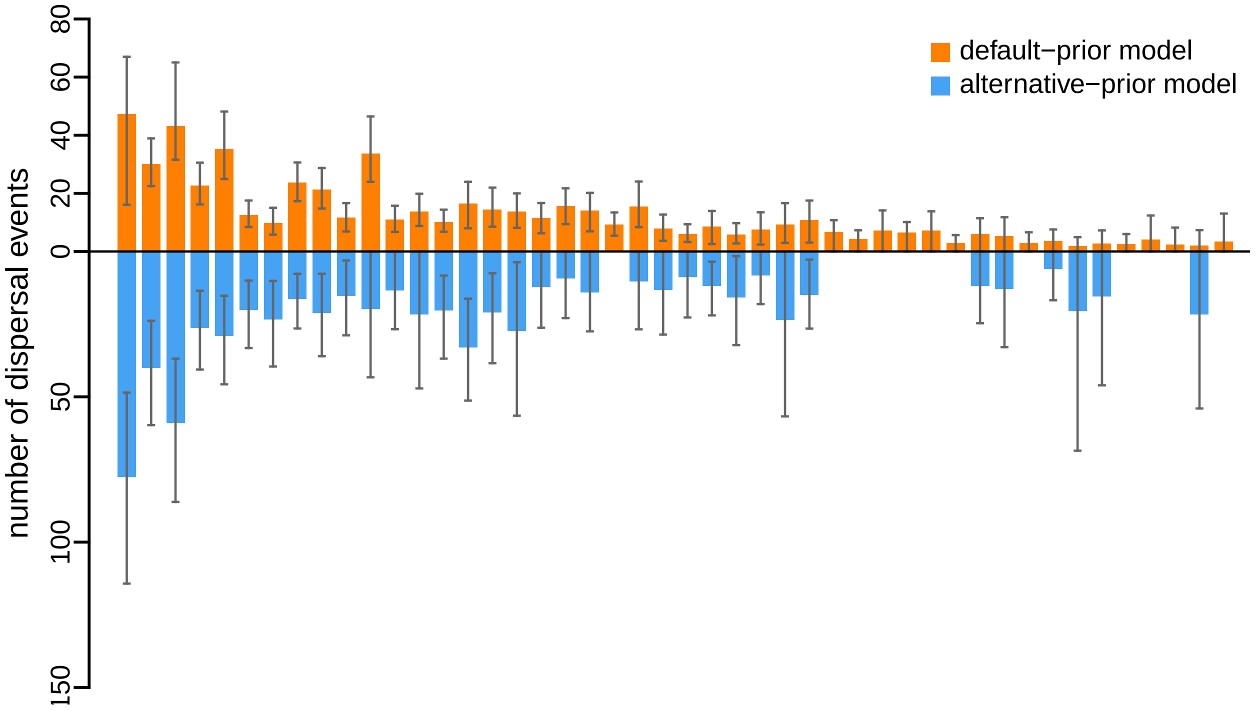


**Figure S122: The impact of prior choice on the inferred number of dispersal events between each pair of areas for Influenza B/Yamagata dataset.** The reflected bar plot depicts the number of dispersal events inferred under the default (orange) and alternative (blue) prior models for Influenza B/Yamagata dataset. Each bar indicates the posterior-mean number of dispersal events between a pair of areas; whiskers indicate the 95% credible interval. Note that only the number of dispersal events over the "significant" dispersal routes (*i.e.*, $2 \ln \text{BF} > 2$) are figured.

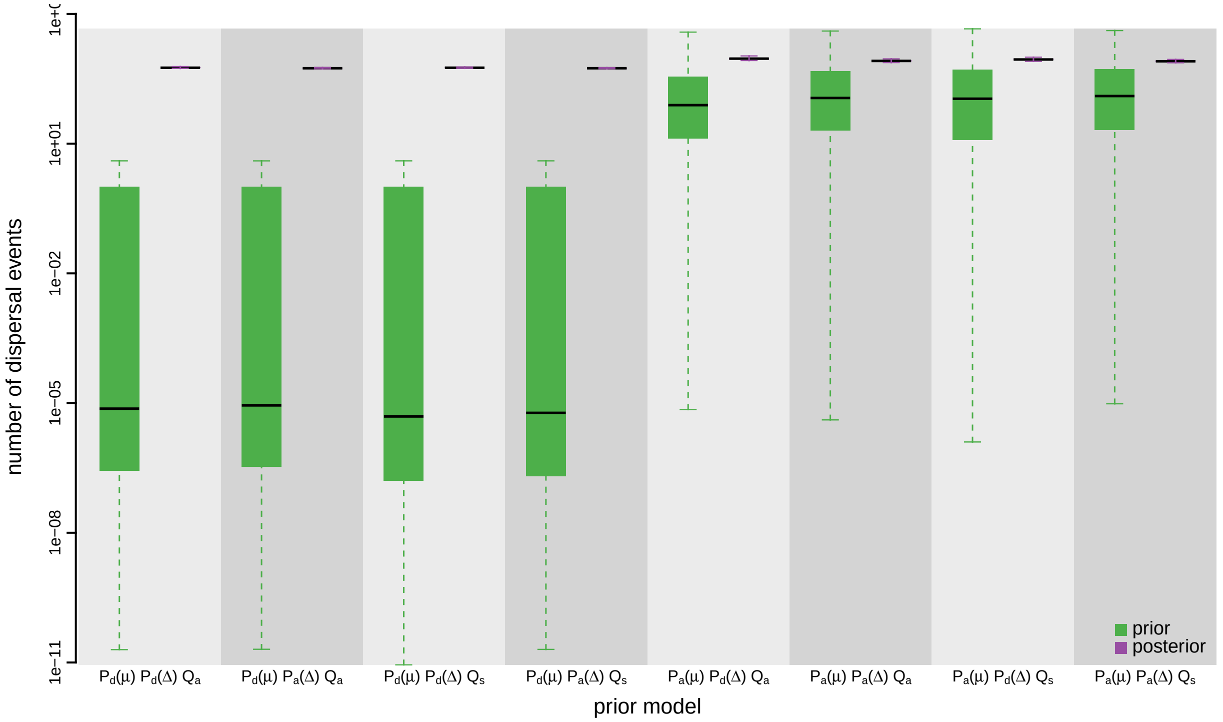


**Figure S123: The impact of prior choice on the inferred total number of dispersal events between all areas for Influenza B/Yamagata dataset.** Each pair of boxplots depicts the specified prior (green) and corresponding posterior estimate (purple) of the number of dispersal events between all areas under each prior model: the center of each box indicates the posterior-median number of dispersal events; the box and whiskers indicate the corresponding 50% and 95% credible intervals, respectively. Model notation follows the description in Table S14.

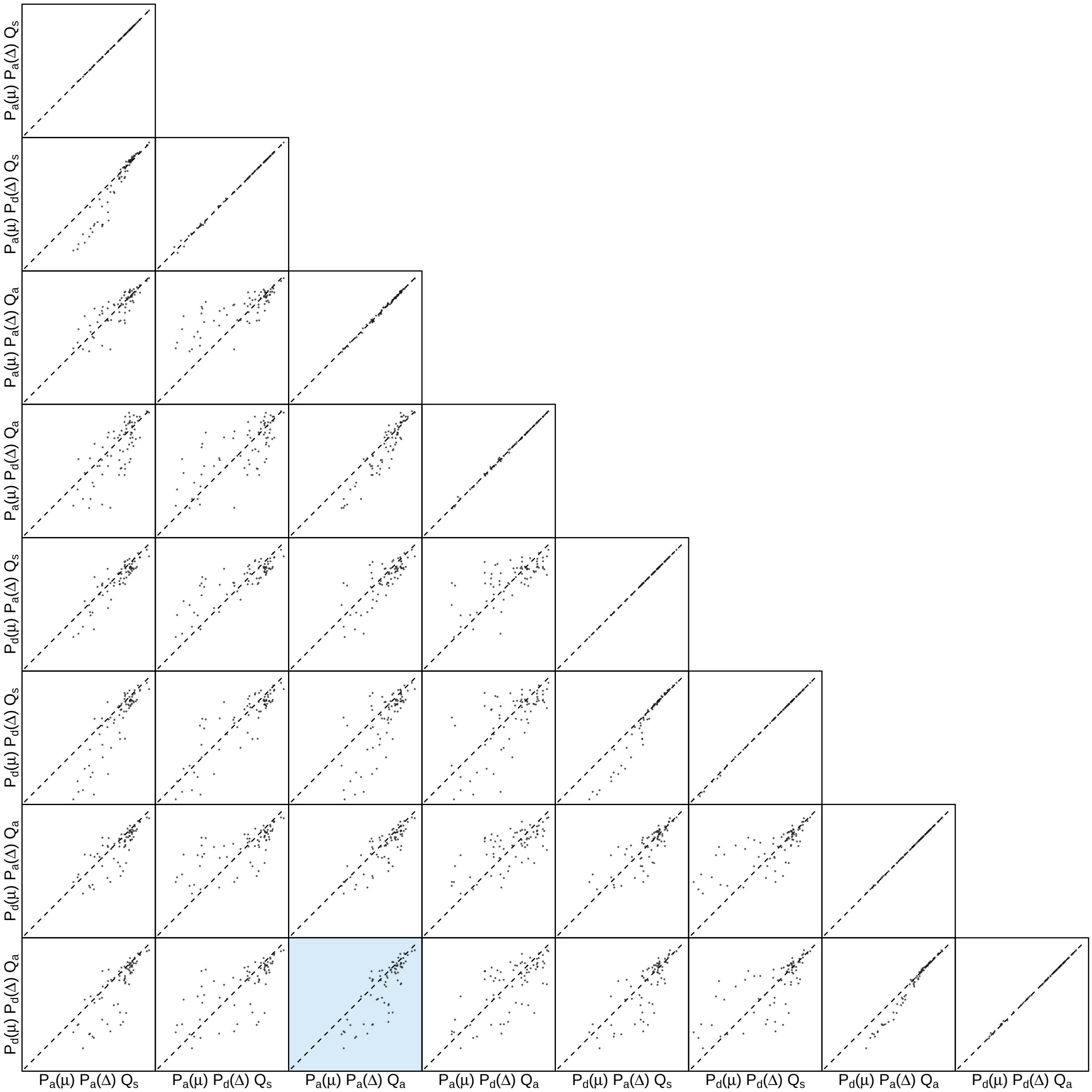


**Figure S124: The impact of prior choice on the inferred number of dispersal events between each pair of areas for Influenza B/Yamagata dataset.** Each cell of the plot compares the inferred number of pairwise dispersal events between each pair of prior models. *x*- and *y*-axis of each cell are both on log scale. Diagonal cells are comparisons between half and the other half of the replicates under the same prior model, assessing the convergence of MCMC simulations; off-diagonal cells are comparisons between different prior models, demonstrating the impact of the prior model on the inferred number of pairwise dispersal events. The shaded cell corresponds to the comparison between the preferred default- and the preferred alternative-prior models (*c.f.*, Figure S122). Model notation follows the description in Table S14.

###### S3.3.5 Rabies Virus

Yao et al. (2015) explored the geographic dynamics of the rabies virus in China based on sequences sampled across 19 provinces between 1986–2012. The authors performed separate phylodynamic analyses on two main lineages, Clade I and Clade II. Our reanalyses are based on the dataset defined by Clade I.

We acquired the sampling time and location data, as well as the GenBank accession numbers from Table S1 in Yao et al. (2015), and then obtained the nucleotide sequences from GenBank. This dataset has 141 sequences distributed among 18 geographic areas. We aligned the nucleotide sequences using `MUSCLE` version 3.8 (Edgar 2004). The files containing the GenBank accession numbers, the sequence alignment, and the sampling time and location data are available in our GitHub and Dryad repositories.

To infer the marginal posterior distribution of phylogenies given the sequence alignment, we specified a phylogenetic model with the following components: (1) the GTR+I+$\Gamma_4$ substitution model (Tavaré 1986; Yang 1994; Gu et al. 1995); (2) the uncorrelated lognormal (UCLN) branch-rate prior model (Drummond et al. 2006; Rannala and Yang 2007), and; (3) the Gaussian Markov Random Field (GMRF) Bayesian Skyride coalescent node-age model (Minin et al. 2008). Details of these analyses are available in the XML scripts included in our GitHub and Dryad repositories.

We ran four independent MCMC simulations in `BEAST` version 1.8.2 for 200 million generations each, sampling every 15000 generations. We first assessed the performance of each MCMC simulation using `Tracer` version 1.7.1 (Rambaut et al. 2018), removed the first 10% of samples from each chain as the burn-in, and then combined the remaining posterior samples of trees from the replicate simulations using `LogCombiner` version 1.8.2. This resulted in a posterior sample of 1200 trees (available in our GitHub and Dryad repositories), which we then used as the prior distribution of phylogenies for the second step of our sequential analyses.

The MCMC simulations used in the second step of our sequential analyses and of the analyses used to estimate marginal likelihoods under each prior model are described above in Section S3.1. Details of these analyses are available in the XML scripts included in our GitHub and Dryad repositories.

**Table S15: Marginal-likelihood estimates of the eight prior models for the Rabies virus dataset.** Columns 2–5 list marginal likelihoods inferred from four replicate analyses. Columns 6–7 list the mean and standard deviation of these marginal-likelihood estimates. The last column lists the marginal-likelihood estimates computed by combining the samples from the replicate power-posterior MCMC simulations. Candidate models are listed in rows and include all possible combinations of: (1) instantaneous-rate matrices (symmetric, $Q_s$ or asymmetric, $Q_a$); (2) priors on the average dispersal rate [default, $P_d(\mu)$ or alternative, $P_a(\mu)$], and; (3) priors on the number of dispersal routes [default, $P_d(\Delta)$ or alternative, $P_a(\Delta)$]. The preferred default- and alternative-prior models are indicated in bold text.

| Model | replicate1 | replicate2 | replicate3 | replicate4 | mean | sd | combined |
|---|---|---|---|---|---|---|---|
| $\boldsymbol{P_d(\mu) Q_a P_d(\Delta)}$ | **-305.71** | **-305.90** | **-306.00** | **-305.91** | **-305.88** | **0.12** | **-305.87** |
| $P_d(\mu) Q_a P_a(\Delta)$ | -296.80 | -296.81 | -296.74 | -296.67 | -296.75 | 0.06 | -296.75 |
| $P_d(\mu) Q_s P_d(\Delta)$ | -319.33 | -319.90 | -319.12 | -319.13 | -319.37 | 0.37 | -319.20 |
| $P_d(\mu) Q_s P_a(\Delta)$ | -299.37 | -299.85 | -299.47 | -299.63 | -299.58 | 0.21 | -299.52 |
| $P_a(\mu) Q_a P_d(\Delta)$ | -262.24 | -262.24 | -262.07 | -262.54 | -262.27 | 0.19 | -262.26 |
| $\boldsymbol{P_a(\mu) Q_a P_a(\Delta)}$ | **-258.28** | **-258.18** | **-258.16** | **-257.92** | **-258.13** | **0.15** | **-258.13** |
| $P_a(\mu) Q_s P_d(\Delta)$ | -269.21 | -269.26 | -269.49 | -269.30 | -269.32 | 0.12 | -269.24 |
| $P_a(\mu) Q_s P_a(\Delta)$ | -258.67 | -258.73 | -258.40 | -258.49 | -258.57 | 0.15 | -258.48 |

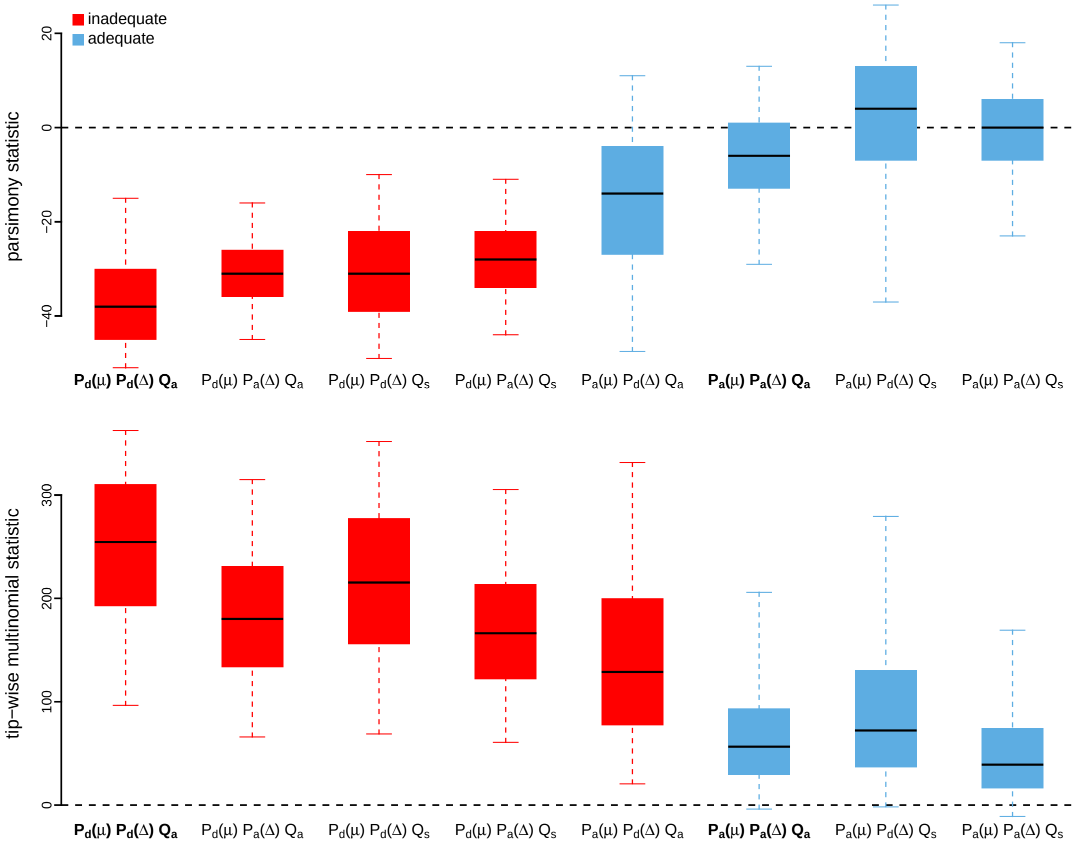


**Figure S125: Posterior-predictive distributions of the parsimony statistic (top panel) and the tip-wise multinomial statistic (bottom panel) under each of the eight prior models for the Rabies virus dataset.** Boxplots depict the posterior-predictive distributions of the statistic under each of the eight candidate prior models: the center of each box is the median predictive value of the summary statistic; the box and whiskers indicate the corresponding 50% and 95% posterior-predictive intervals, respectively. The horizontal dashed line indicates when the simulated and observed datasets produce identical value for the summary statistic. A model is judged to be inadequate (*i.e.*, incapable of generating geographic datasets that are similar to the observed data) if its 95% posterior-predictive interval does not overlap with the dashed line. Here the preferred default prior model is inadequate, whereas the preferred alternative prior model is adequate. Notation for the candidate models is described in Table S15; the preferred default- and alternative-prior models are indicated in bold text.

*The Impact of Prior Choice on Average Dispersal Rate*

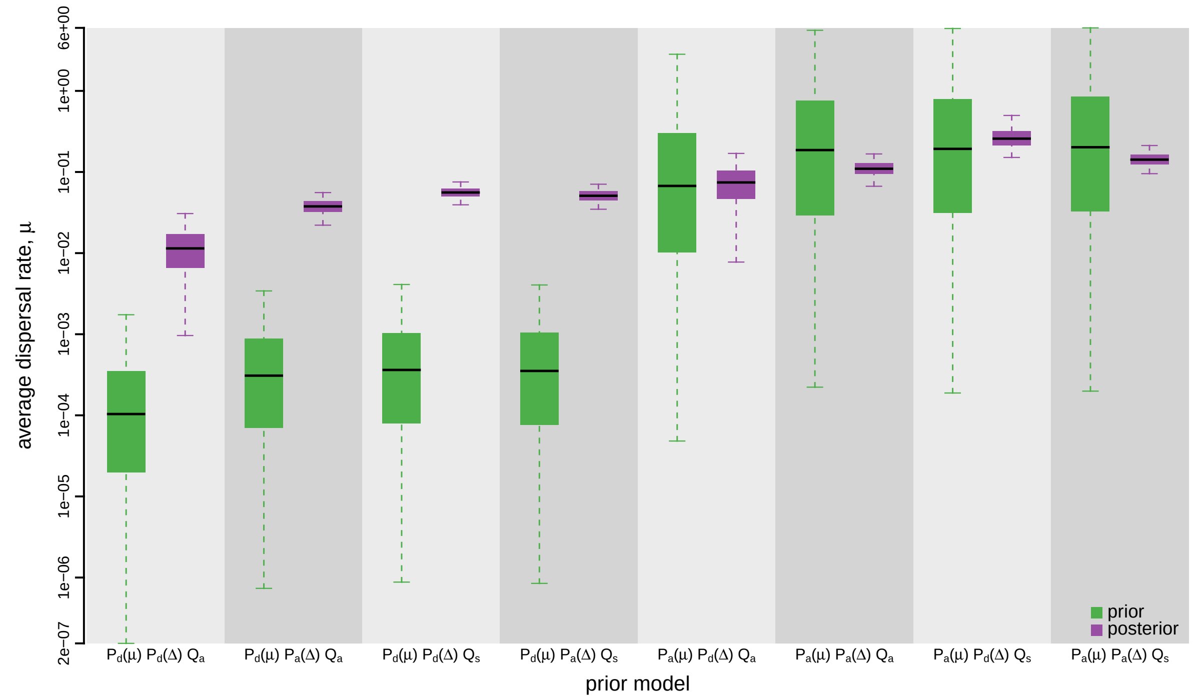


**Figure S126: The impact of prior choice on the average rate of dispersal for the Rabies virus dataset.** Each pair of boxplots depicts the specified prior (green) and corresponding posterior estimate (purple) of the average dispersal rate, $\mu$, under each prior model: the center of each box indicates the median rate; the box and whiskers indicate the corresponding 50% and 95% credible intervals, respectively. Notation for the prior models is as described in Table S15.

*The Impact of Prior Choice on Pairwise Dispersal Rates*

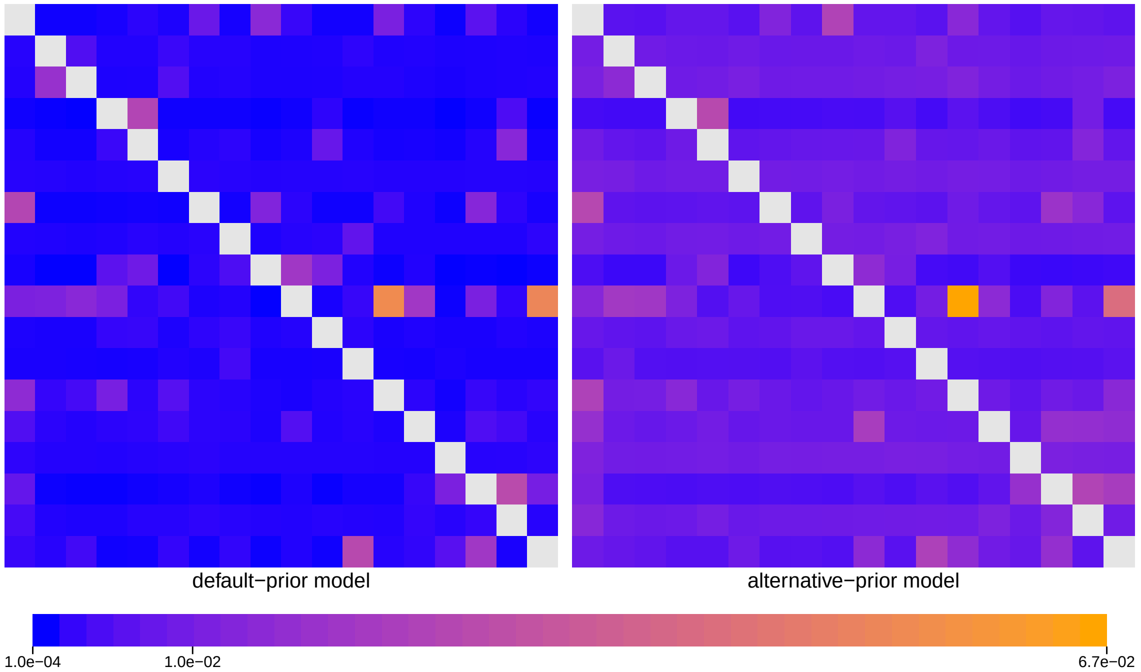


**Figure S127: The impact of prior choice on pairwise dispersal rates for the Rabies virus dataset.** Heatmaps summarize posterior-mean estimates of the instantaneous rate of dispersal between each pair of geographic areas, $q_{ij}$, under the default (left) and alternative (right) prior models.

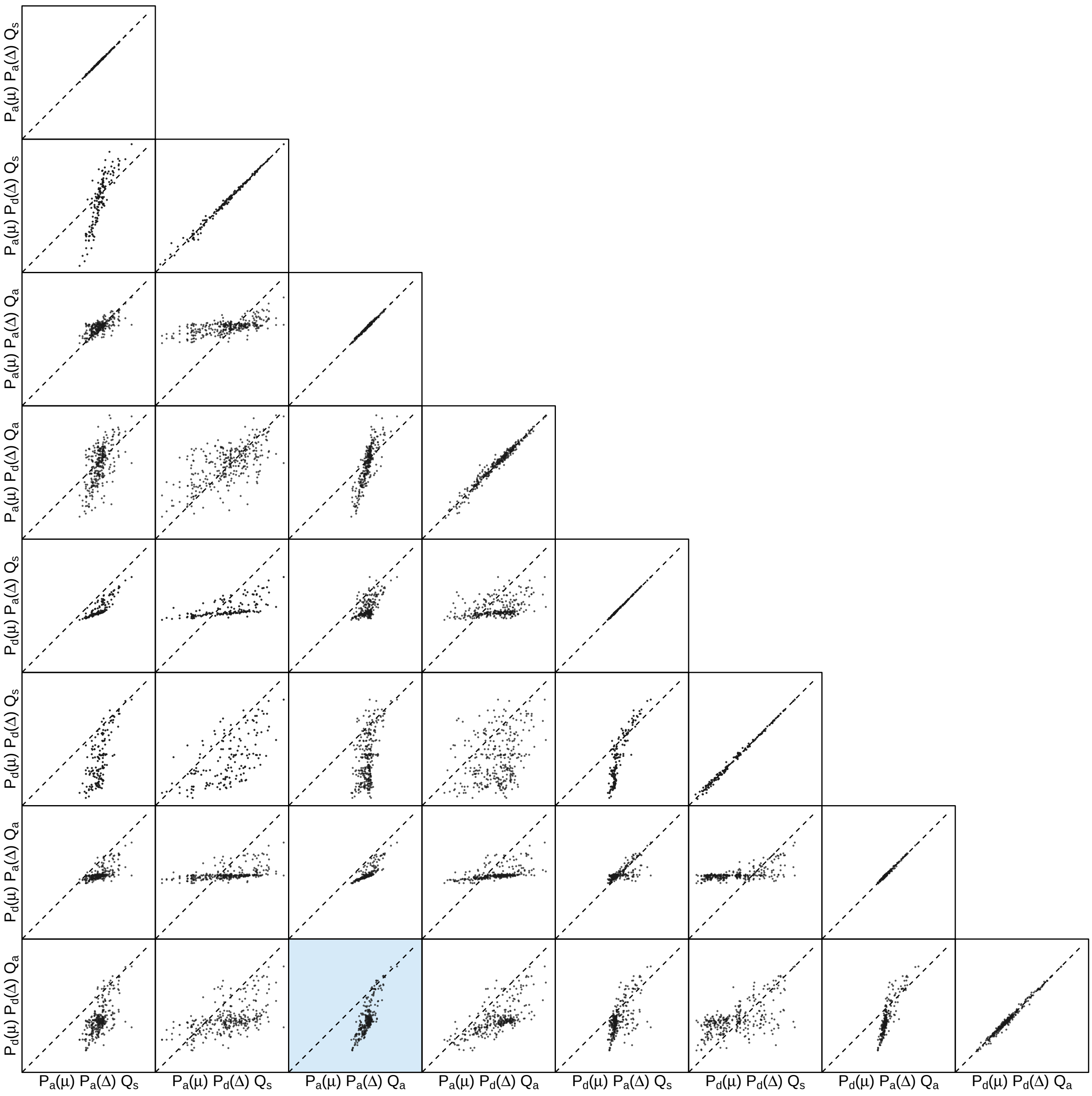


**Figure S128: The impact of prior choice on pairwise dispersal rates for the Rabies virus dataset.** Each cell of the plot compares posterior-mean estimate of the rate of dispersal between each pair of geographic areas, $q_{ij}$, between each pair of prior models. $x$- and $y$-axis of each cell are both on log scale. Diagonal cells are comparisons between half and the other half of the replicates under the same prior model, assessing the convergence of MCMC simulations; off-diagonal cells are comparisons between different prior models, demonstrating the impact of the prior model on the estimates of pairwise dispersal rates. The shaded cell corresponds to the comparison between the preferred default- and the preferred alternative-prior models (*c.f.*, Figure S127). Notation for the prior models is as described in Table S15.

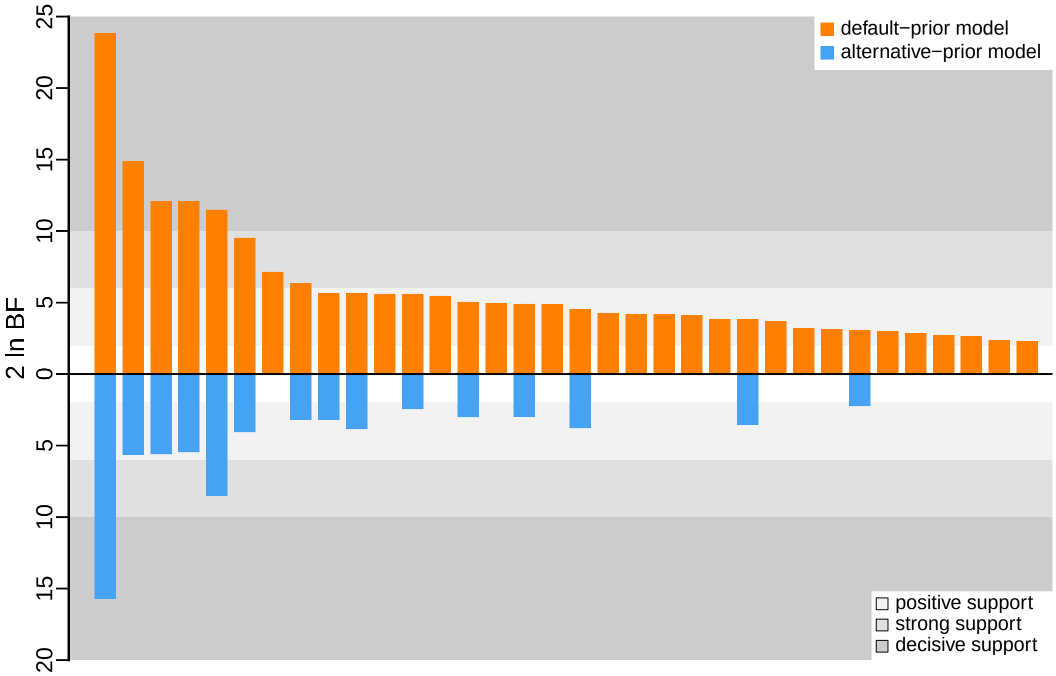


**Figure S129: The impact of prior choice on the inferred support for dispersal routes for the Rabies virus dataset.** We compare the evidential support for each dispersal route for the Rabies virus dataset under the default (orange) and alternative (blue) prior models. Each bar indicates the $2 \ln \text{BF}$ for the corresponding dispersal route between two areas; only supported dispersal routes (*i.e.*, $2 \ln \text{BF} > 2$) are plotted.

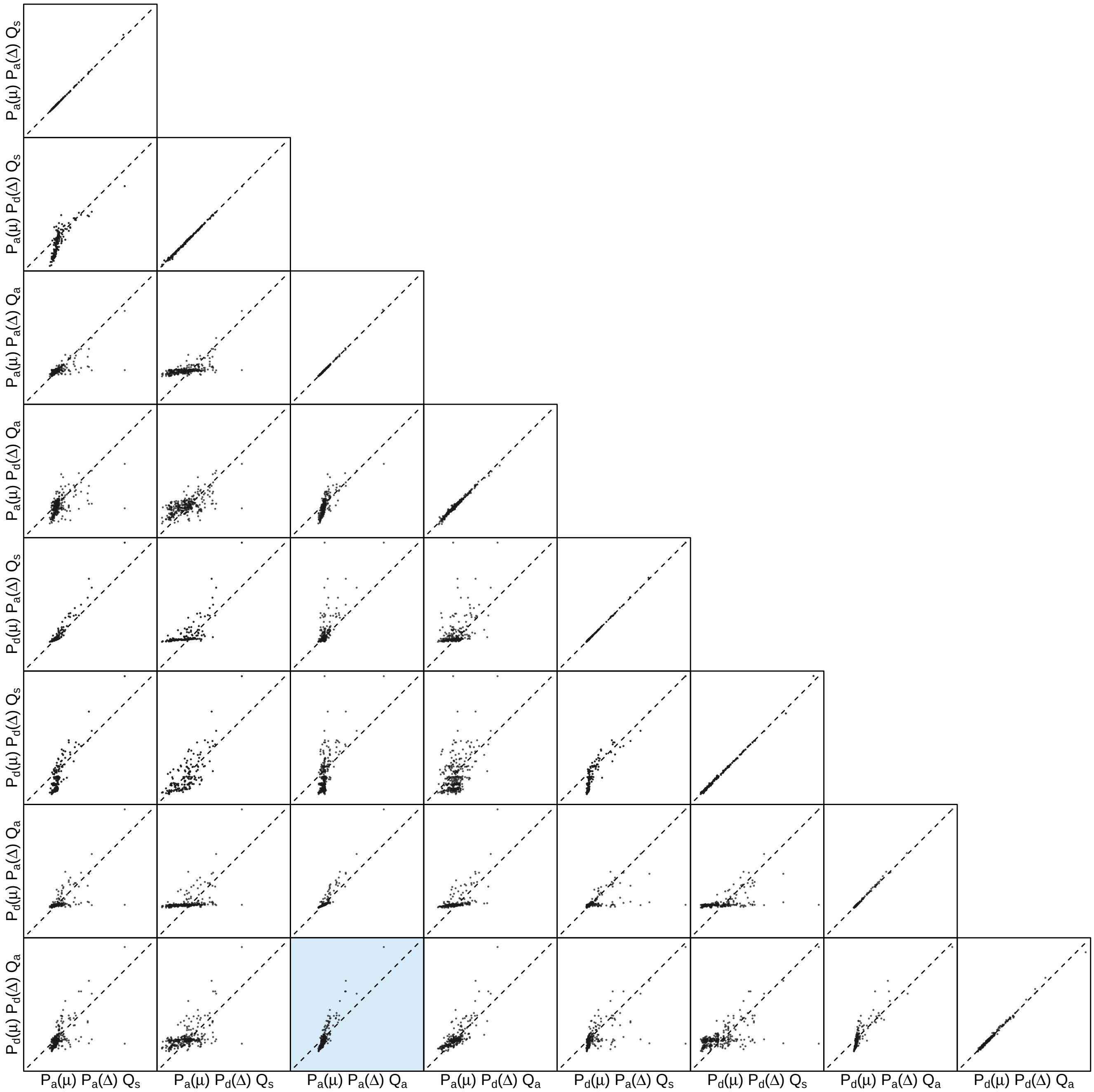


**Figure S130: The impact of prior choice on the inferred support for dispersal routes for the Rabies virus dataset.** Each cell of the plot compares the inferred support ($2 \ln \mathrm{BF}$) for pairwise dispersal routes between each pair of prior models. Diagonal cells are comparisons between half and the other half of the replicates under the same prior model, assessing the convergence of MCMC simulations; off-diagonal cells are comparisons between different prior models, demonstrating the impact of the prior model on the inferred support for pairwise dispersal routes. The shaded cell corresponds to the comparison between the preferred default- and the preferred alternative-prior models (*c.f.*, Figure S129). Notation for the prior models is as described in Table S15.

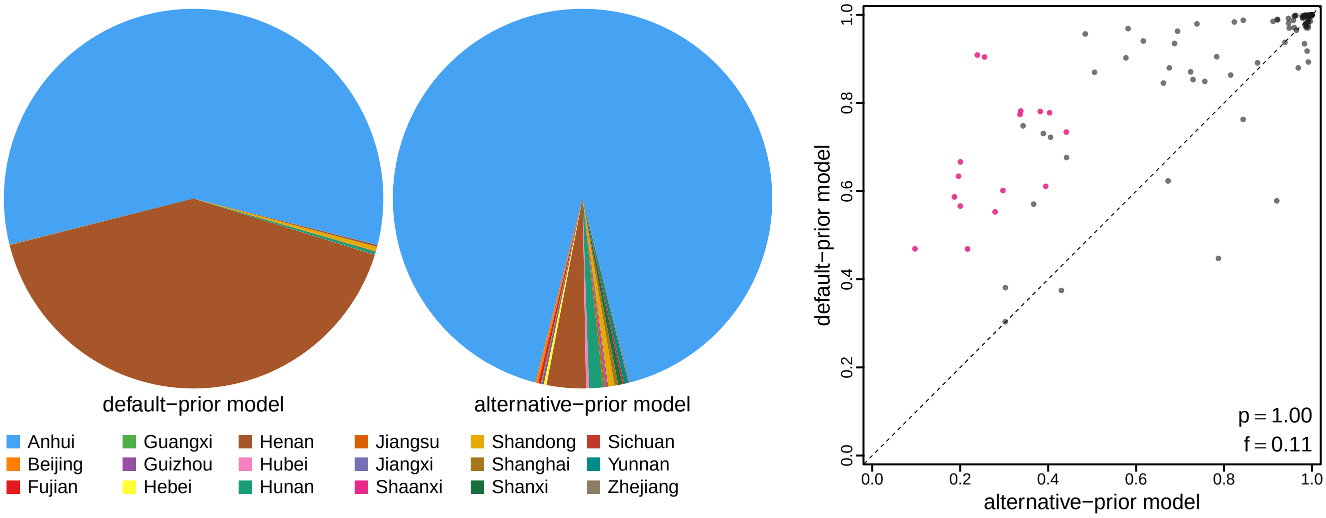


**Figure S131: The impact of prior choice on ancestral-area estimates for the Rabies virus dataset.** The left panel compares the posterior probability of each ancestral area at the root node under the default- and alternative-prior models for the Rabies virus dataset. The right panel plots the posterior probability of the most probable ancestral area under the default-prior model for each node in the MCC tree (y-axis) against the corresponding posterior probability of that area under the alternative-prior model (x-axis). Pink dots represent the internal nodes where the MAP ancestral area inferred under the default-prior model differs from that inferred under the alternative-prior models. The statistic $p$ denotes the fraction of internal nodes that are shared under the default- and alternative-prior models; $f$, is the fraction of shared nodes where the MAP ancestral area differs under the default- and alternative-prior models. Note that the posterior probabilities of the MAP ancestral area under the default-prior model are generally higher than those under the alternative-prior model (*i.e.*, the default-prior model tends to mask uncertainty in the ancestral-area estimates).

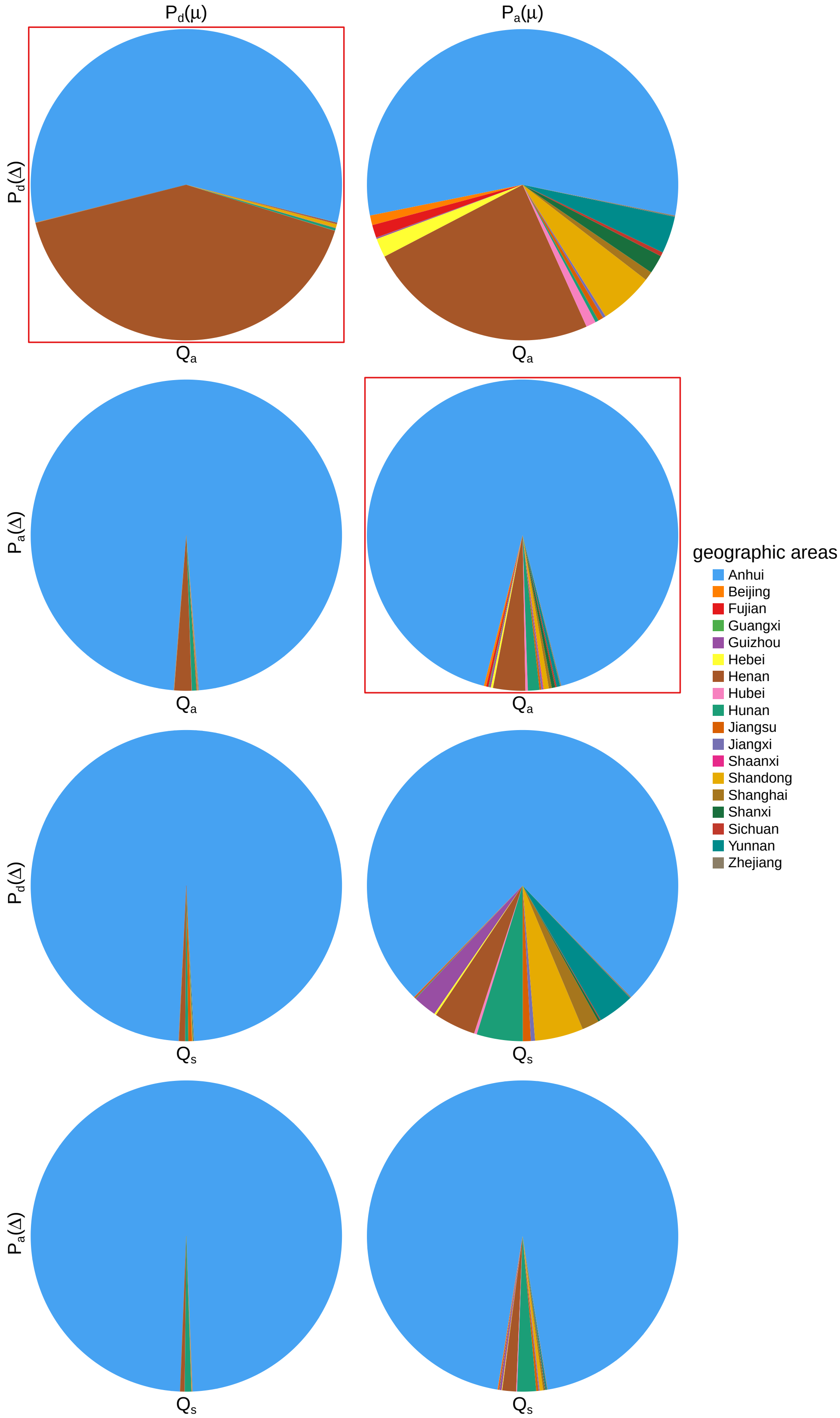


**Figure S132: The impact of prior choice on ancestral-area estimates at the root node for the Rabies virus dataset.** The posterior probability of each ancestral area at the root node under each prior model for the Rabies virus dataset. The pie chart enclosed in red corresponds to the comparison between the preferred default- and the preferred alternative-prior models (*c.f.*, Figure S131, left panel). Model notation follows the description in Table S15.

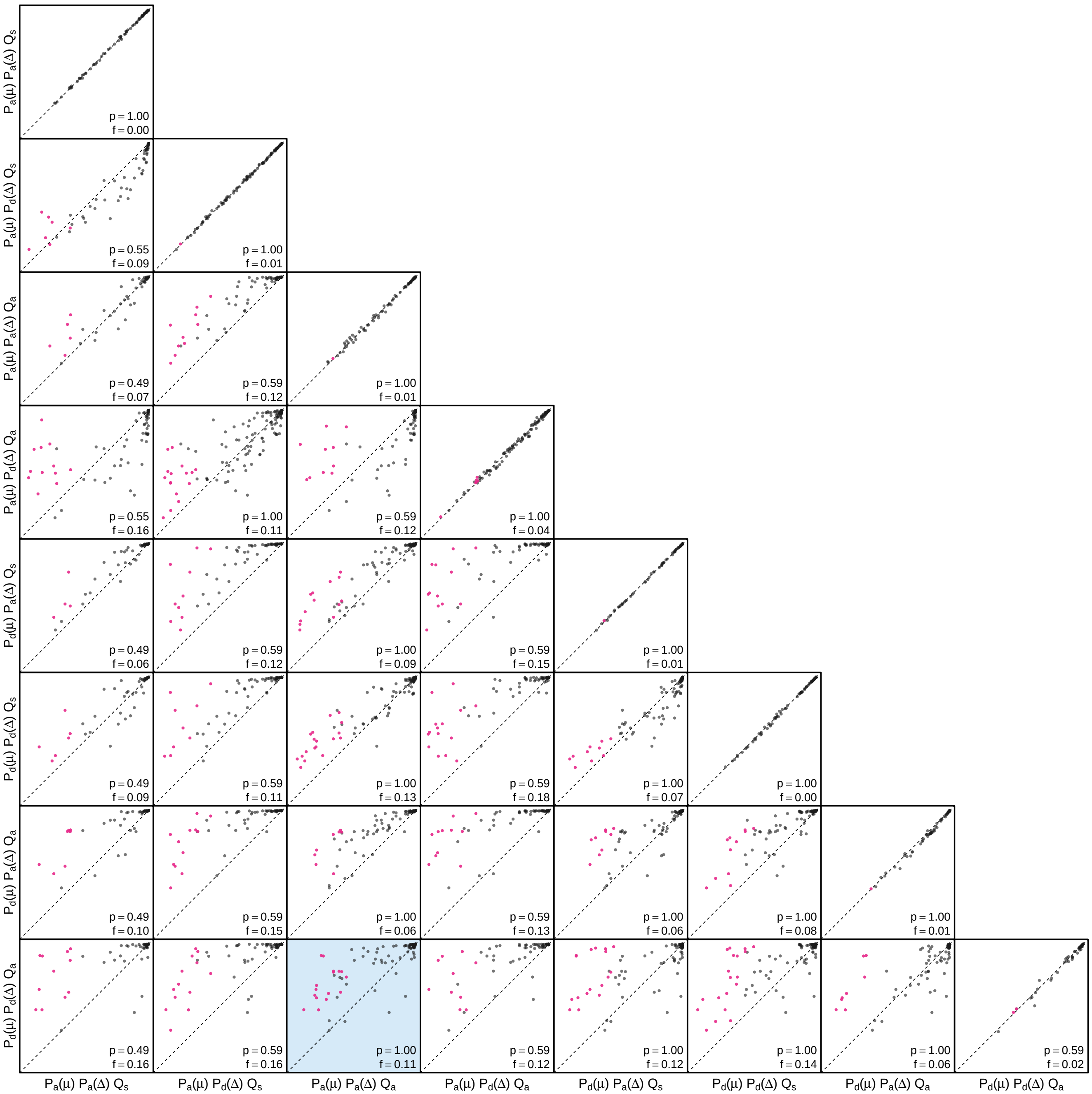


**Figure S133: The impact of prior choice on the MAP ancestral-area estimates at internal nodes for the Rabies virus dataset.** Pairwise scatter plots compare the posterior probability of the MAP ancestral area inferred under each combination of prior models. Diagonal cells are comparisons between two replicates under the same prior model (assessing the convergence of MCMC simulations); off-diagonal cells are comparisons between different prior models, demonstrating the impact of the prior model on both the posterior probability and the identity of the MAP ancestral-area estimates at internal nodes. Pink dots represent the internal nodes where the MAP ancestral area inferred under the default-prior model differs from that inferred under the alternative-prior models. The statistic $p$ denotes the fraction of internal nodes that are shared under the default- and alternative-prior models; $f$ is the fraction of shared nodes where the MAP ancestral area differs under the default- and alternative-prior models. The shaded cell corresponds to the comparison between the preferred default- and the preferred alternative-prior models (*c.f.*, Figure S131, right panel). Model notation follows the description in Table S15.

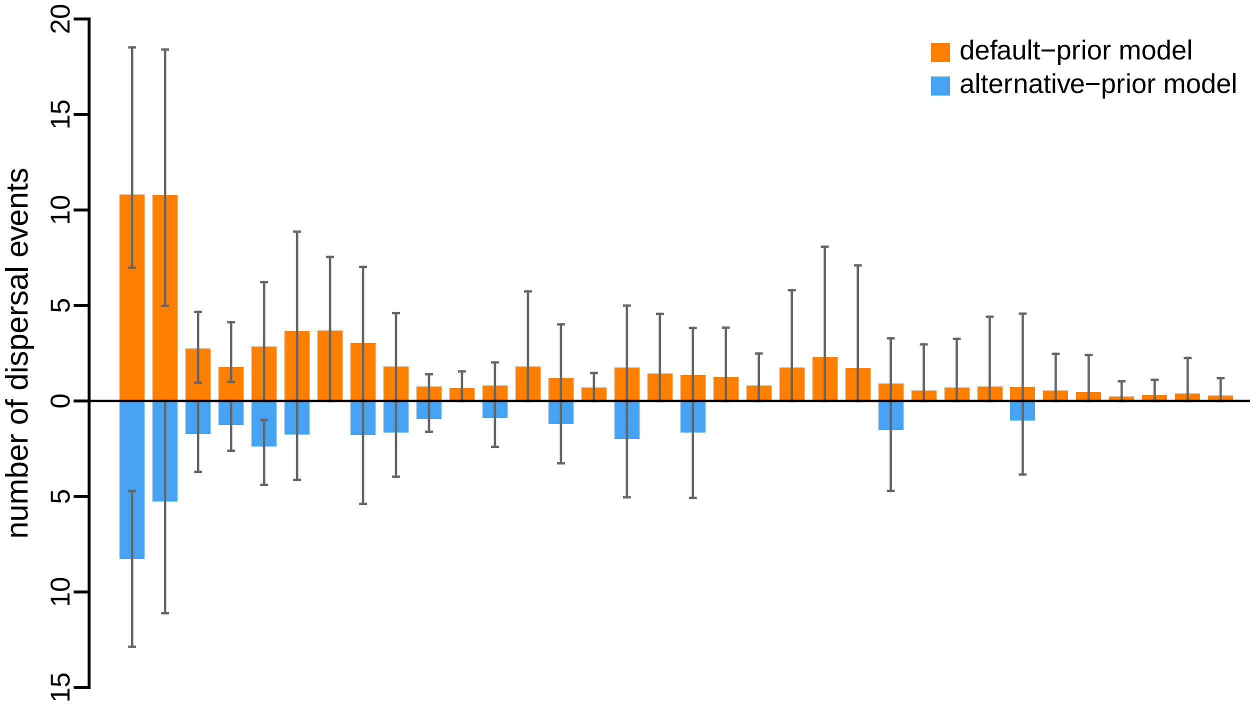


**Figure S134: The impact of prior choice on the inferred number of dispersal events between each pair of areas for the Rabies virus dataset.** The reflected bar plot depicts the number of dispersal events inferred under the default (orange) and alternative (blue) prior models for the Rabies virus dataset. Each bar indicates the posterior-mean number of dispersal events between a pair of areas; whiskers indicate the 95% credible interval. Note that only the number of dispersal events over the "significant" dispersal routes (*i.e.*, $2 \ln \mathrm{BF} > 2$) are figured.

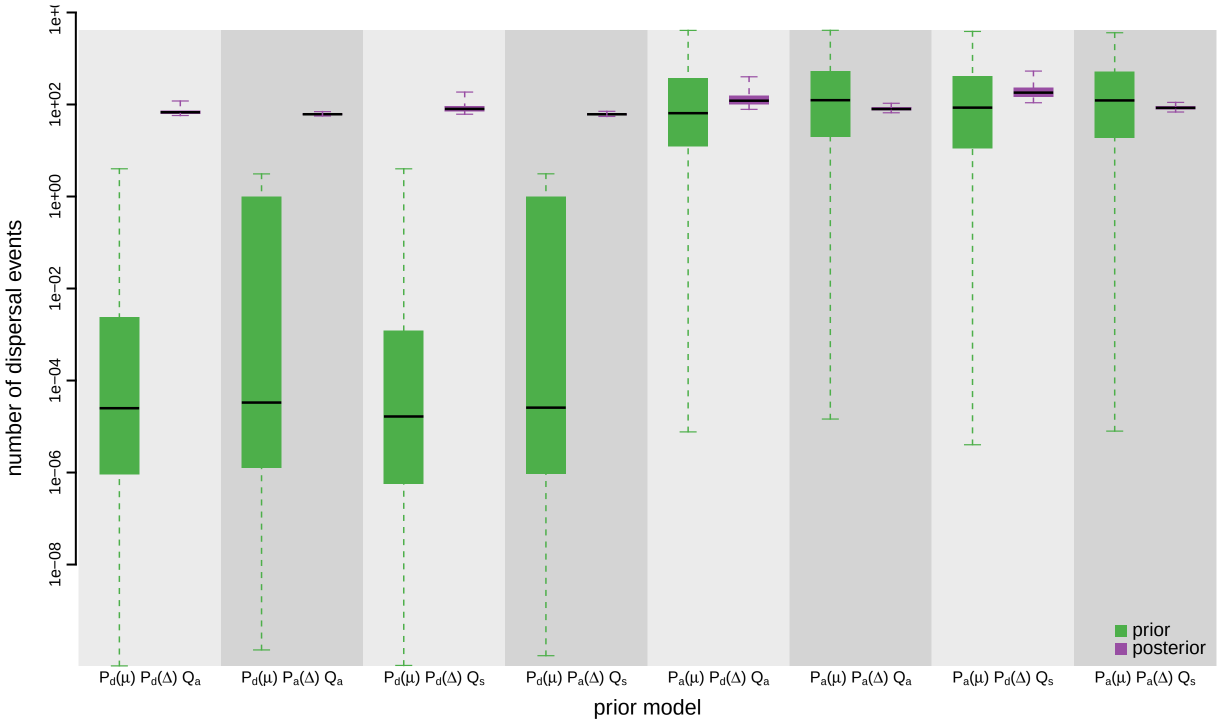


**Figure S135: The impact of prior choice on the inferred total number of dispersal events between all areas for the Rabies virus dataset.** Each pair of boxplots depicts the specified prior (green) and corresponding posterior estimate (purple) of the number of dispersal events between all areas under each prior model: the center of each box indicates the posterior-median number of dispersal events; the box and whiskers indicate the corresponding 50% and 95% credible intervals, respectively. Model notation follows the description in Table S15.

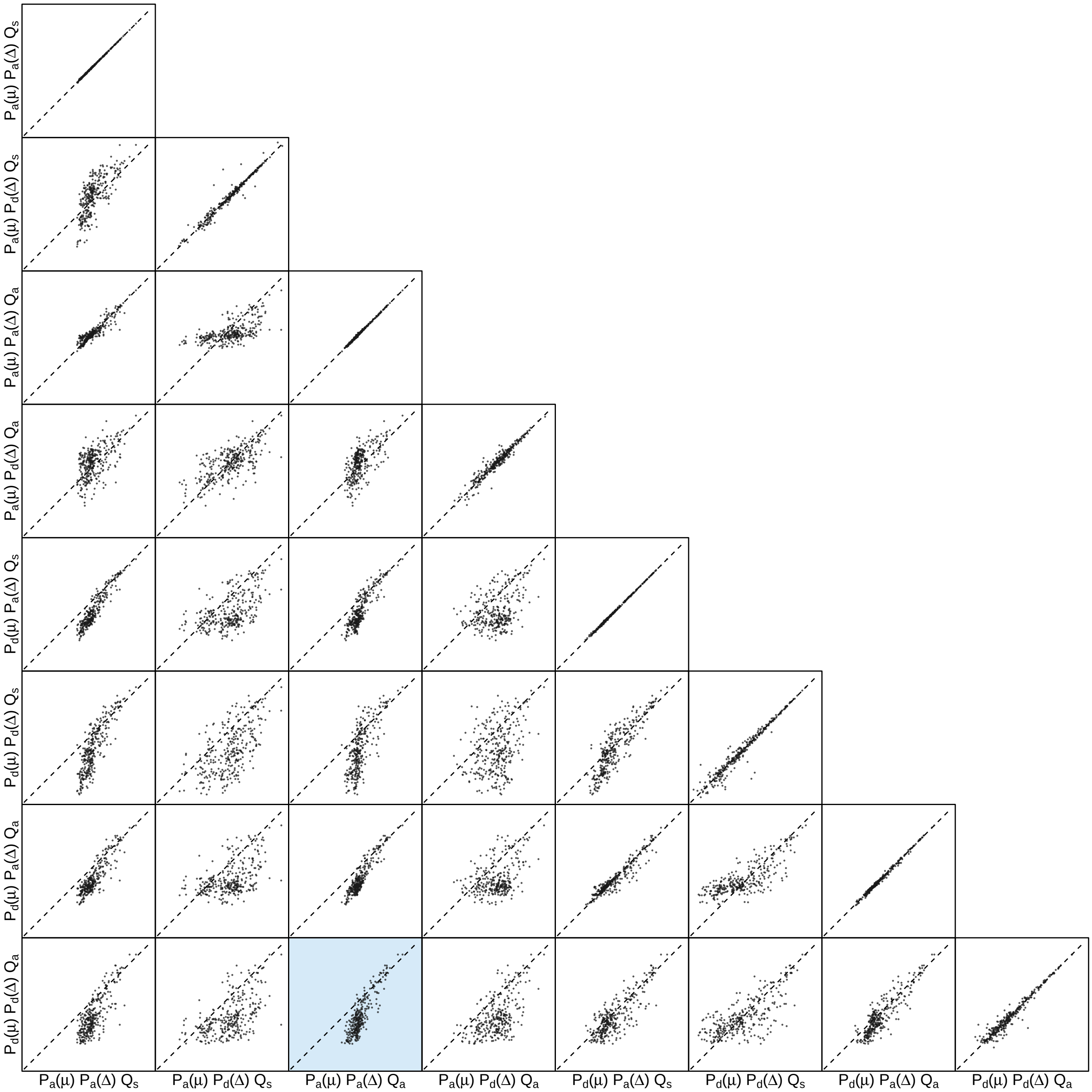


**Figure S136: The impact of prior choice on the inferred number of dispersal events between each pair of areas for the Rabies virus dataset.** Each cell of the plot compares the inferred number of pairwise dispersal events between each pair of prior models. *x*- and *y*-axis of each cell are both on log scale. Diagonal cells are comparisons between half and the other half of the replicates under the same prior model, assessing the convergence of MCMC simulations; off-diagonal cells are comparisons between different prior models, demonstrating the impact of the prior model on the inferred number of pairwise dispersal events. The shaded cell corresponds to the comparison between the preferred default- and the preferred alternative-prior models (*c.f.*, Figure S134). Model notation follows the description in Table S15.

###### S3.3.6 SARS-CoV-2 Global

Gao et al. (2022) explored the early spread of the COVID-19 pandemic and the efficacy of mitigation measures on limiting the spread using publicly available SARS-CoV-2 genomic sequences collected during the early phase of the pandemic. Here we used one of the datasets produced in that study to assess the impact of prior misspecification on phylodynamic inference of biogeographic history. This dataset contains 1271 SARS-CoV-2 genomic sequences that were originally obtained from the Global Initiative on Sharing All Influenza Data (GISAID, Shu and McCauley 2017). Details about the data curation process can be found in Gao et al. (2022). An alignment were then inferred using `MUSCLE` version 3.8 (Edgar 2004) with the curated dataset. The sampling time and geographic location associated with each sequence of the alignment were also acquired from GISAID.

Different from the other datasets, where we performed the second step of the sequential phylodynamic inferences by marginalizing over the posterior distribution of trees inferred using sequence data and the associated sampling times (but not sampling locations), here we conditioned on the maximum clade credibility (MCC) tree summarized from the posterior distribution to ensure numerical stability of the analyses. This MCC tree was obtained directly from Gao et al. (2022); details (including model and prior specification, as well as the `BEAST` analyses settings) about the analyses that estimated the tree can be found in that study. Following Gao et al. (2022), we discretized the globe into 23 geographic areas to; see Gao et al. (2022) for detailed description of the geographic-area delineation. The MCC tree and the file containing the associated sampling time and location of each sequence are available in our GitHub and Dryad repositories.

The MCMC simulations used in the second step of our sequential analyses and of the analyses used to estimate marginal likelihoods under each prior model are described above in Section S3.1. Details of these analyses are available in the XML scripts included in our GitHub and Dryad repositories.

**Table S16: Marginal-likelihood estimates of the eight prior models for the SARS-CoV-2 Global dataset.** Columns 2–5 list marginal likelihoods inferred from four replicate analyses. Columns 6–7 list the mean and standard deviation of these marginal-likelihood estimates. The last column lists the marginal-likelihood estimates computed by combining the samples from the replicate power-posterior MCMC simulations. Candidate models are listed in rows and include all possible combinations of: (1) instantaneous-rate matrices (symmetric, $Q_s$ or asymmetric, $Q_a$); (2) priors on the average dispersal rate [default, $P_d(\mu)$ or alternative, $P_a(\mu)$], and; (3) priors on the number of dispersal routes [default, $P_d(\Delta)$ or alternative, $P_a(\Delta)$]. The preferred default- and alternative-prior models are indicated in bold text.

| Model | replicate1 | replicate2 | replicate3 | replicate4 | mean | sd | combined |
|---|---|---|---|---|---|---|---|
| $\boldsymbol{P_d(\mu)Q_aP_d(\Delta)}$ | **-2519.38** | **-2519.41** | **-2519.39** | **-2519.15** | **-2519.33** | **0.12** | **-2519.27** |
| $P_d(\mu)Q_aP_a(\Delta)$ | -2503.66 | -2503.87 | -2503.32 | -2504.18 | -2503.76 | 0.36 | -2503.71 |
| $P_d(\mu)Q_sP_d(\Delta)$ | -2560.17 | -2559.85 | -2559.58 | -2559.94 | -2559.88 | 0.24 | -2559.85 |
| $P_d(\mu)Q_sP_a(\Delta)$ | -2502.78 | -2503.01 | -2502.83 | -2502.98 | -2502.90 | 0.11 | -2502.89 |
| $P_a(\mu)Q_aP_d(\Delta)$ | -2158.24 | -2159.08 | -2159.01 | -2159.29 | -2158.9 | 0.46 | -2158.83 |
| $\boldsymbol{P_a(\mu)Q_aP_a(\Delta)}$ | **-2159.81** | **-2160.07** | **-2160.09** | **-2159.42** | **-2159.85** | **0.31** | **-2159.82** |
| $P_a(\mu)Q_sP_d(\Delta)$ | -2259.81 | -2260.13 | -2259.88 | -2260.11 | -2259.98 | 0.16 | -2259.81 |
| $P_a(\mu)Q_sP_a(\Delta)$ | -2200.55 | -2200.29 | -2200.44 | -2200.43 | -2200.43 | 0.11 | -2200.42 |

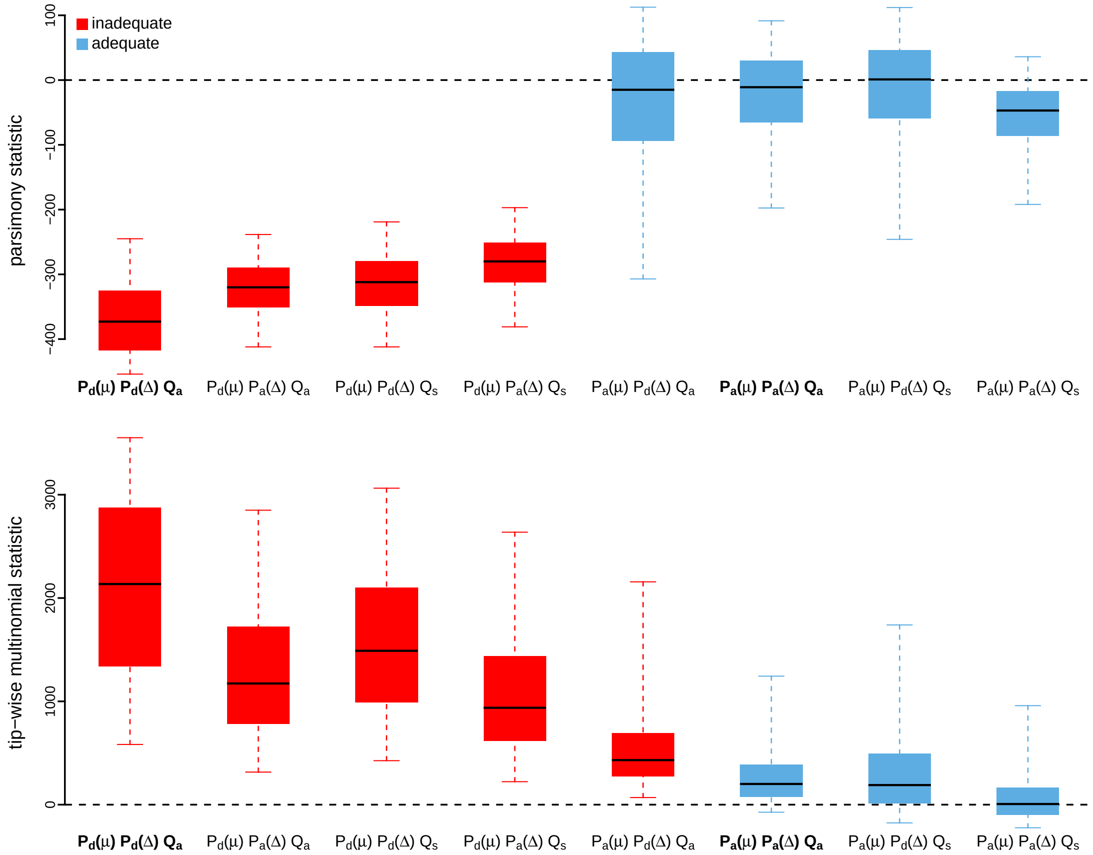


**Figure S137: Posterior-predictive distributions of the parsimony statistic (top panel) and the tip-wise multinomial statistic (bottom panel) under each of the eight prior models for the SARS-CoV-2 Global dataset.** Boxplots depict the posterior-predictive distributions of the statistic under each of the eight candidate prior models: the center of each box is the median predictive value of the summary statistic; the box and whiskers indicate the corresponding 50% and 95% posterior-predictive intervals, respectively. The horizontal dashed line indicates when the simulated and observed datasets produce identical value for the summary statistic. A model is judged to be inadequate (*i.e.*, incapable of generating geographic datasets that are similar to the observed data) if its 95% posterior-predictive interval does not overlap with the dashed line. Here the preferred default prior model is inadequate, whereas the preferred alternative prior model is adequate. Notation for the candidate models is described in Table S16; the preferred default- and alternative-prior models are indicated in bold text.

*The Impact of Prior Choice on Average Dispersal Rate*

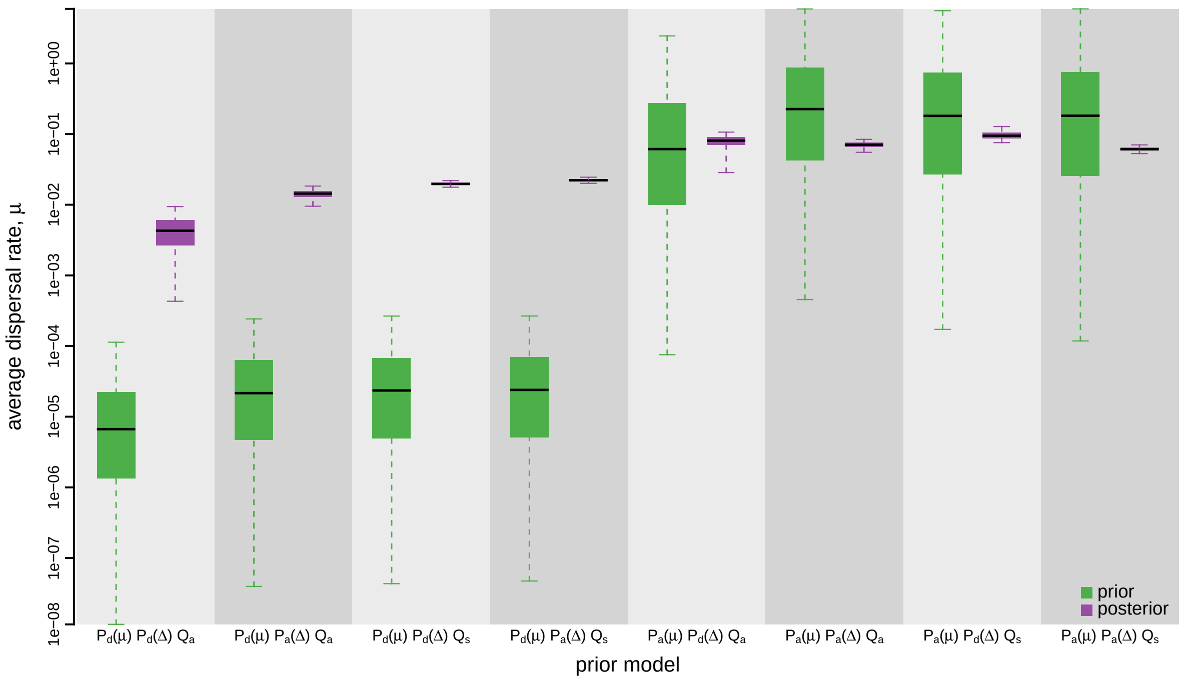


**Figure S138: The impact of prior choice on the average rate of dispersal for the SARS-CoV-2 Global dataset.** Each pair of boxplots depicts the specified prior (green) and corresponding posterior estimate (purple) of the average dispersal rate, $\mu$, under each prior model: the center of each box indicates the median rate; the box and whiskers indicate the corresponding 50% and 95% credible intervals, respectively. Notation for the prior models is as described in Table S16.

*The Impact of Prior Choice on Pairwise Dispersal Rates*

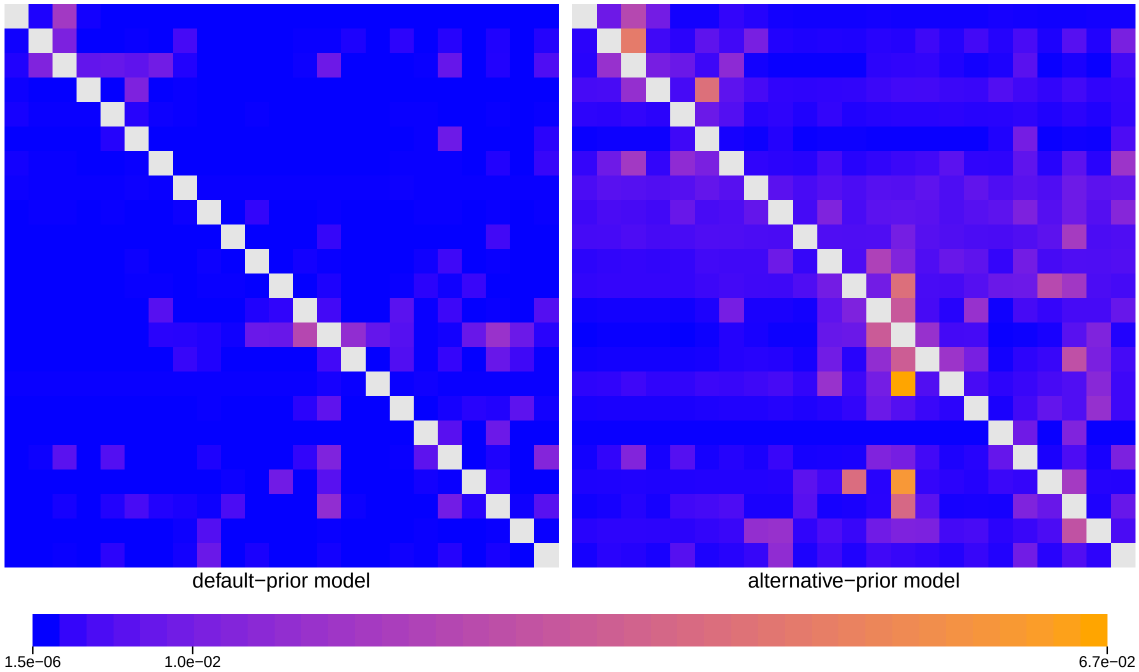


**Figure S139: The impact of prior choice on pairwise dispersal rates for the SARS-CoV-2 Global dataset.** Heatmaps summarize posterior-mean estimates of the instantaneous rate of dispersal between each pair of geographic areas, $q_{ij}$, under the default (left) and alternative (right) prior models.

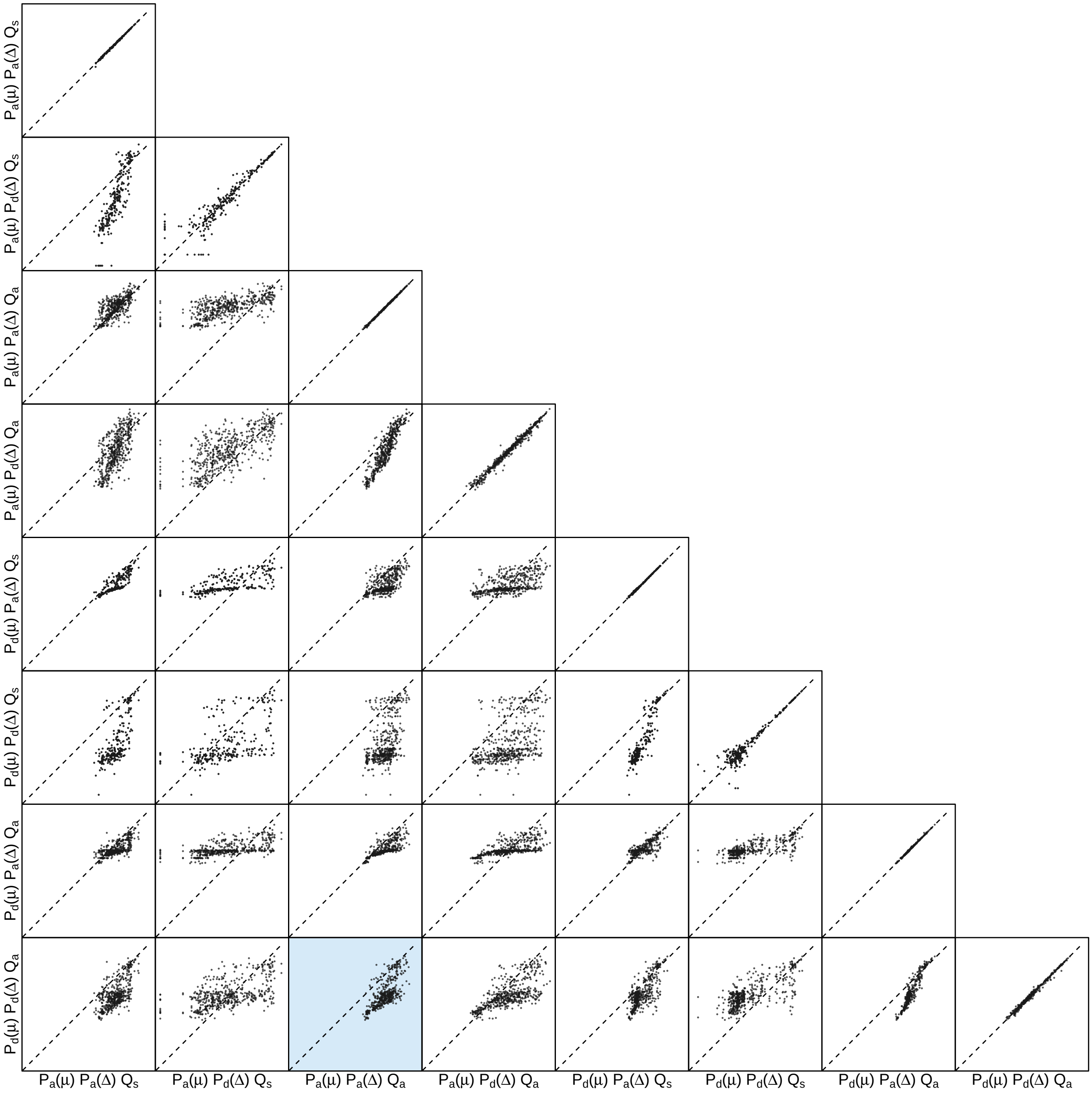


**Figure S140: The impact of prior choice on pairwise dispersal rates for the SARS-CoV-2 Global dataset.** Each cell of the plot compares posterior-mean estimate of the rate of dispersal between each pair of geographic areas, $q_{ij}$, between each pair of prior models. $x$- and $y$-axis of each cell are both on log scale. Diagonal cells are comparisons between half and the other half of the replicates under the same prior model, assessing the convergence of MCMC simulations; off-diagonal cells are comparisons between different prior models, demonstrating the impact of the prior model on the estimates of pairwise dispersal rates. The shaded cell corresponds to the comparison between the preferred default- and the preferred alternative-prior models (*c.f.*, Figure S139). Notation for the prior models is as described in Table S16.

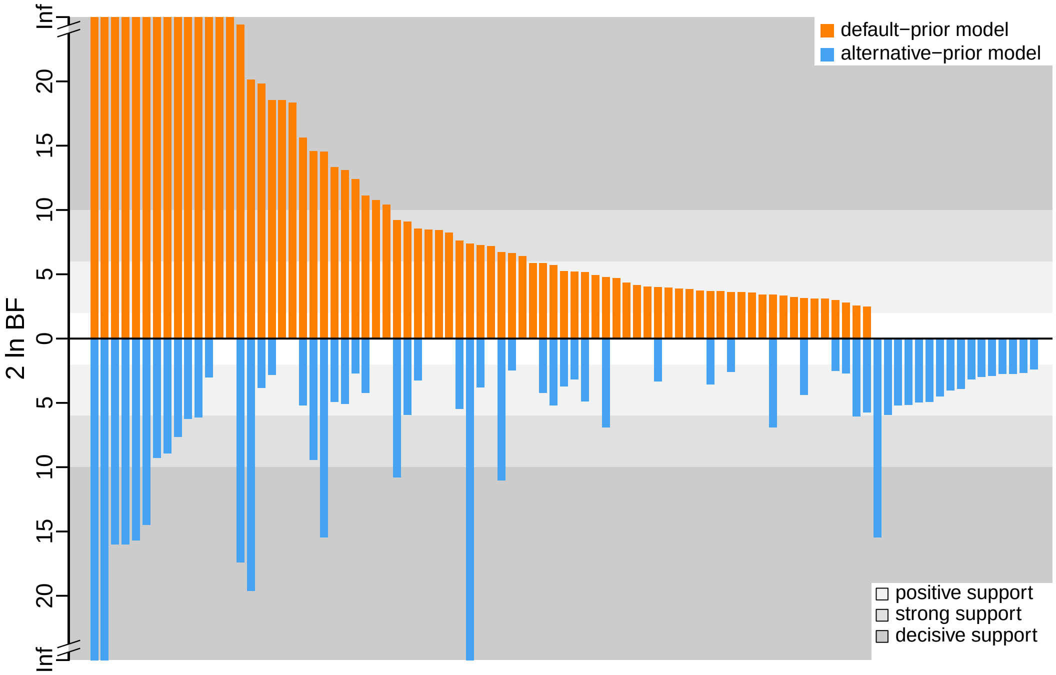


**Figure S141: The impact of prior choice on the inferred support for dispersal routes for the SARS-CoV-2 Global dataset.** We compare the evidential support for each dispersal route for the SARS-CoV-2 Global dataset under the default (orange) and alternative (blue) prior models. Each bar indicates the $2 \ln \mathrm{BF}$ for the corresponding dispersal route between two areas; only supported dispersal routes (*i.e.*, $2 \ln \mathrm{BF} > 2$) are plotted.

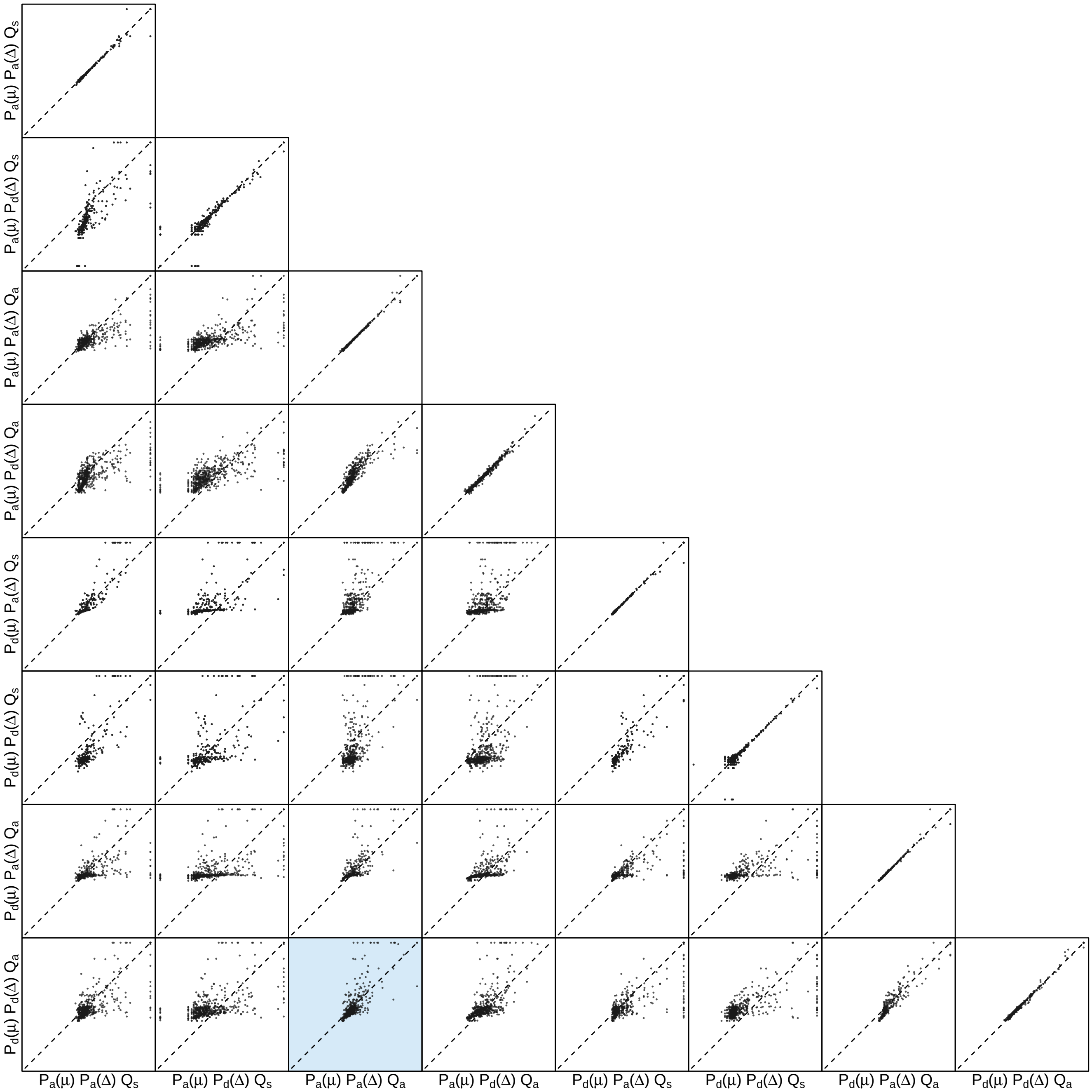


**Figure S142: The impact of prior choice on the inferred support for dispersal routes for the SARS-CoV-2 Global dataset.** Each cell of the plot compares the inferred support ($2 \ln \mathrm{BF}$) for pairwise dispersal routes between each pair of prior models. Diagonal cells are comparisons between half and the other half of the replicates under the same prior model, assessing the convergence of MCMC simulations; off-diagonal cells are comparisons between different prior models, demonstrating the impact of the prior model on the inferred support for pairwise dispersal routes. The shaded cell corresponds to the comparison between the preferred default- and the preferred alternative-prior models (*c.f.*, Figure S141). Notation for the prior models is as described in Table S16.

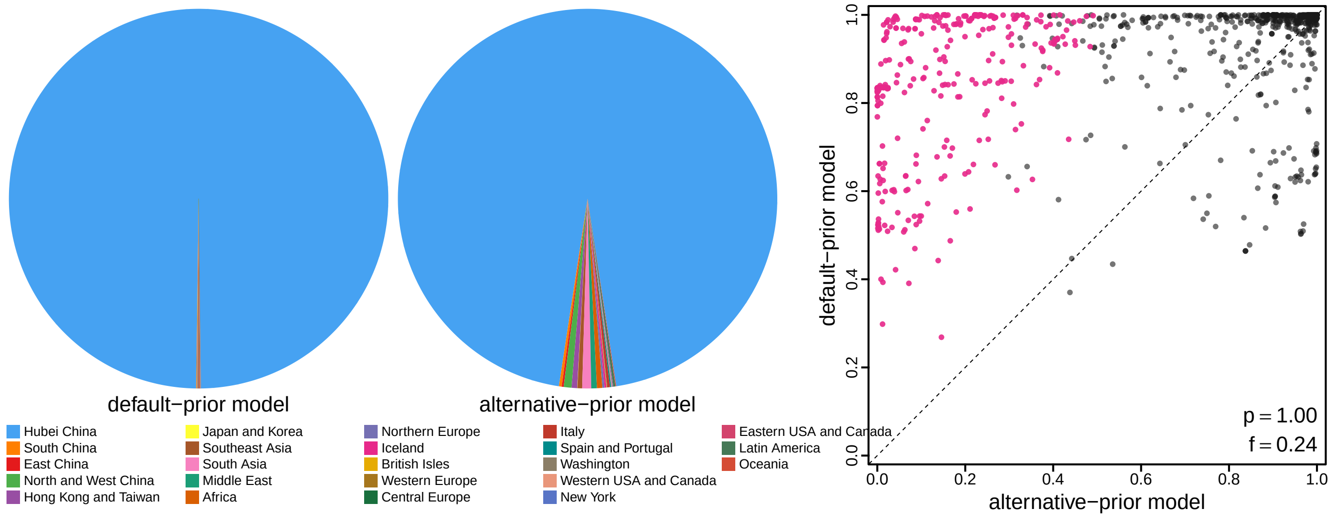


**Figure S143: The impact of prior choice on ancestral-area estimates for the SARS-CoV-2 Global dataset.** The left panel compares the posterior probability of each ancestral area at the root node under the default- and alternative-prior models for the SARS-CoV-2 Global dataset. The right panel plots the posterior probability of the most probable ancestral area under the default-prior model for each node in the MCC tree (y-axis) against the corresponding posterior probability of that area under the alternative-prior model (x-axis). Pink dots represent the internal nodes where the MAP ancestral area inferred under the default-prior model differs from that inferred under the alternative-prior models. The statistic $p$ denotes the fraction of internal nodes that are shared under the default- and alternative-prior models; $f$, is the fraction of shared nodes where the MAP ancestral area differs under the default- and alternative-prior models. Note that the posterior probabilities of the MAP ancestral area under the default-prior model are generally higher than those under the alternative-prior model (*i.e.*, the default-prior model tends to mask uncertainty in the ancestral-area estimates).

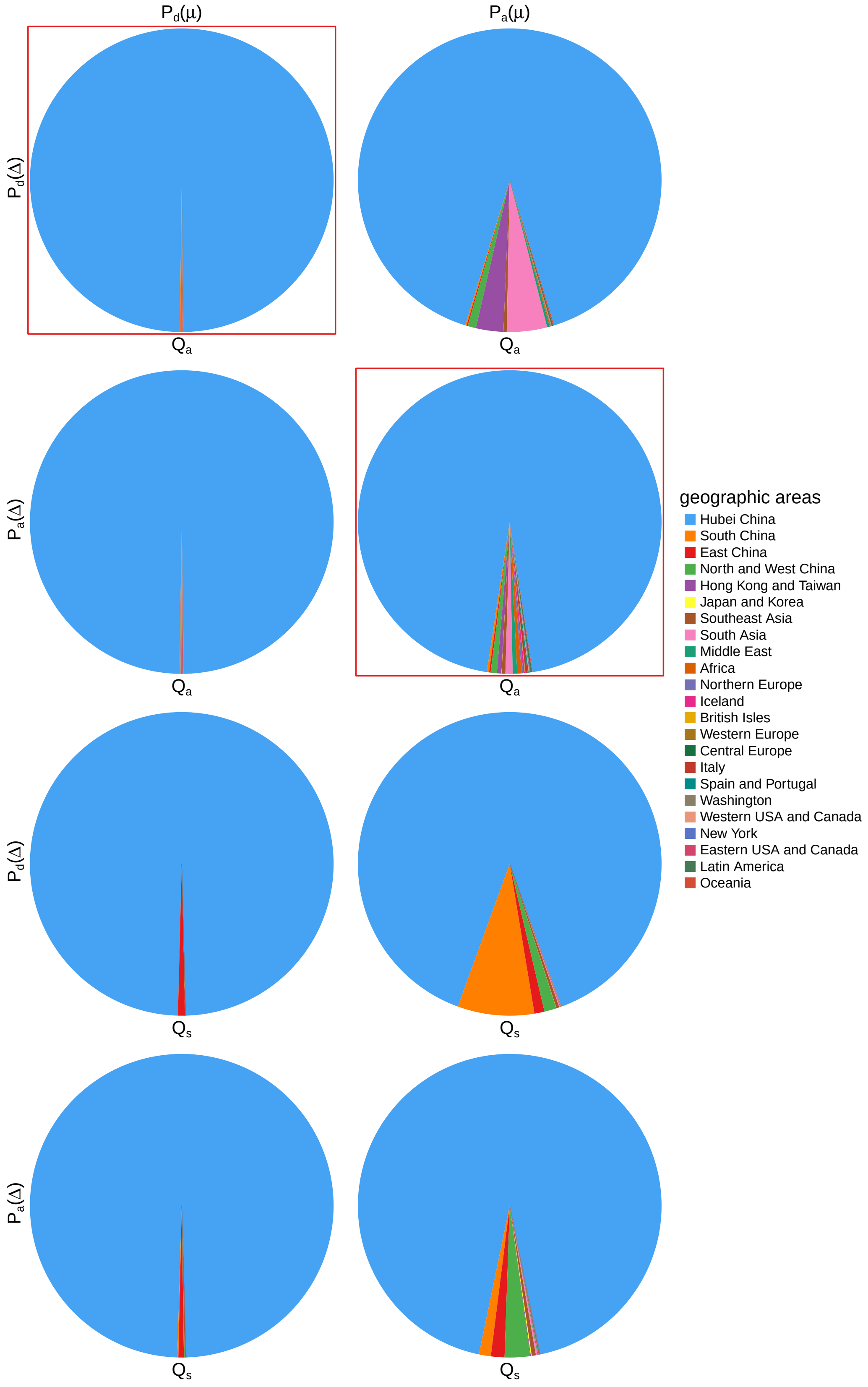


**Figure S144: The impact of prior choice on ancestral-area estimates at the root node for the SARS-CoV-2 Global dataset.** The posterior probability of each ancestral area at the root node under each prior model for the SARS-CoV-2 Global dataset. The pie chart enclosed in red corresponds to the comparison between the preferred default- and the preferred alternative-prior models (*c.f.*, Figure S143, left panel). Model notation follows the description in Table S16.

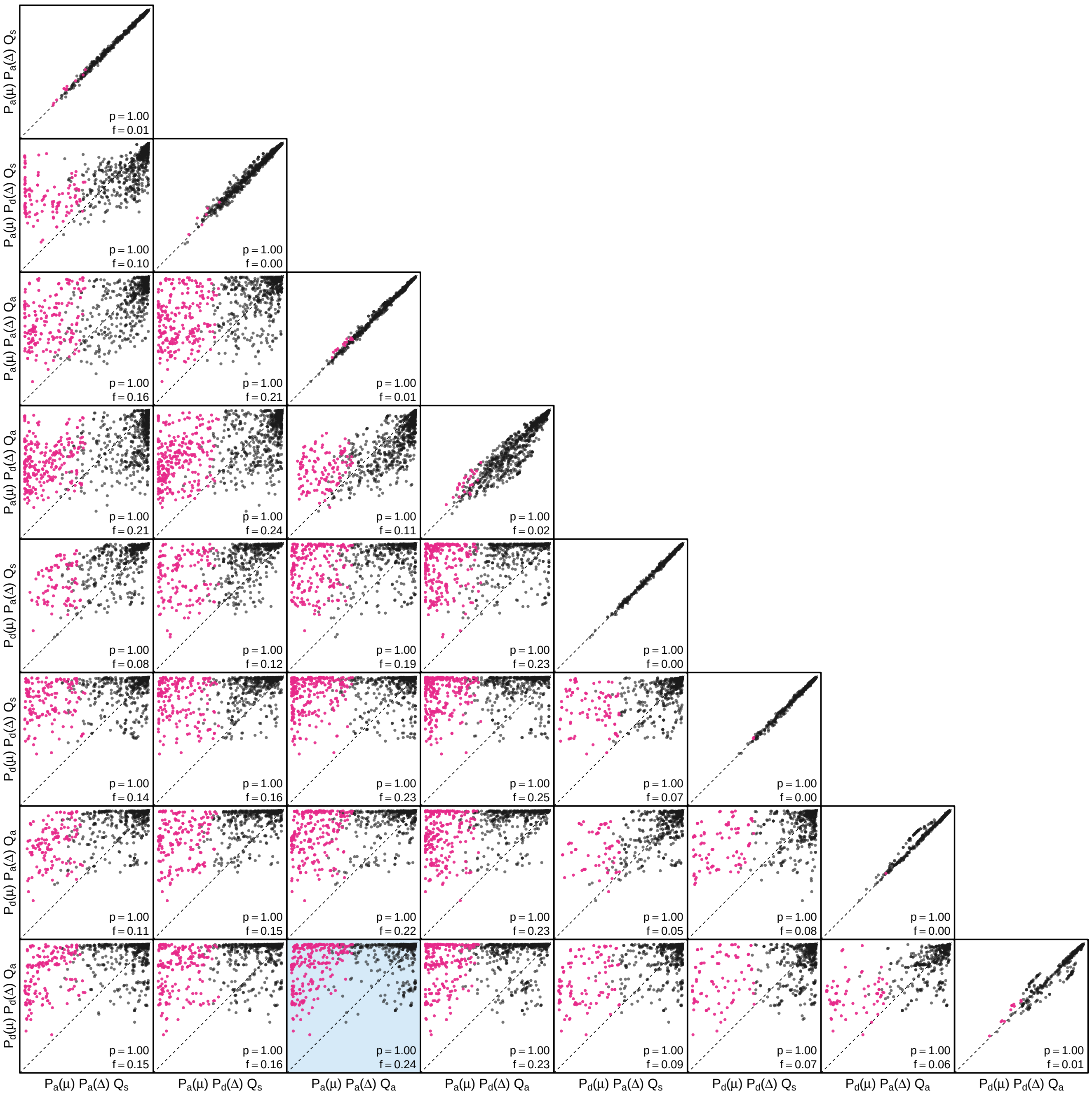


**Figure S145: The impact of prior choice on the MAP ancestral-area estimates at internal nodes for the SARS-CoV-2 Global dataset.** Pairwise scatter plots compare the posterior probability of the MAP ancestral area inferred under each combination of prior models. Diagonal cells are comparisons between two replicates under the same prior model (assessing the convergence of MCMC simulations); off-diagonal cells are comparisons between different prior models, demonstrating the impact of the prior model on both the posterior probability and the identity of the MAP ancestral-area estimates at internal nodes. Pink dots represent the internal nodes where the MAP ancestral area inferred under the default-prior model differs from that inferred under the alternative-prior models. The statistic $p$ denotes the fraction of internal nodes that are shared under the default- and alternative-prior models; $f$ is the fraction of shared nodes where the MAP ancestral area differs under the default- and alternative-prior models. The shaded cell corresponds to the comparison between the preferred default- and the preferred alternative-prior models (*c.f.*, Figure S143, right panel). Model notation follows the description in Table S16.

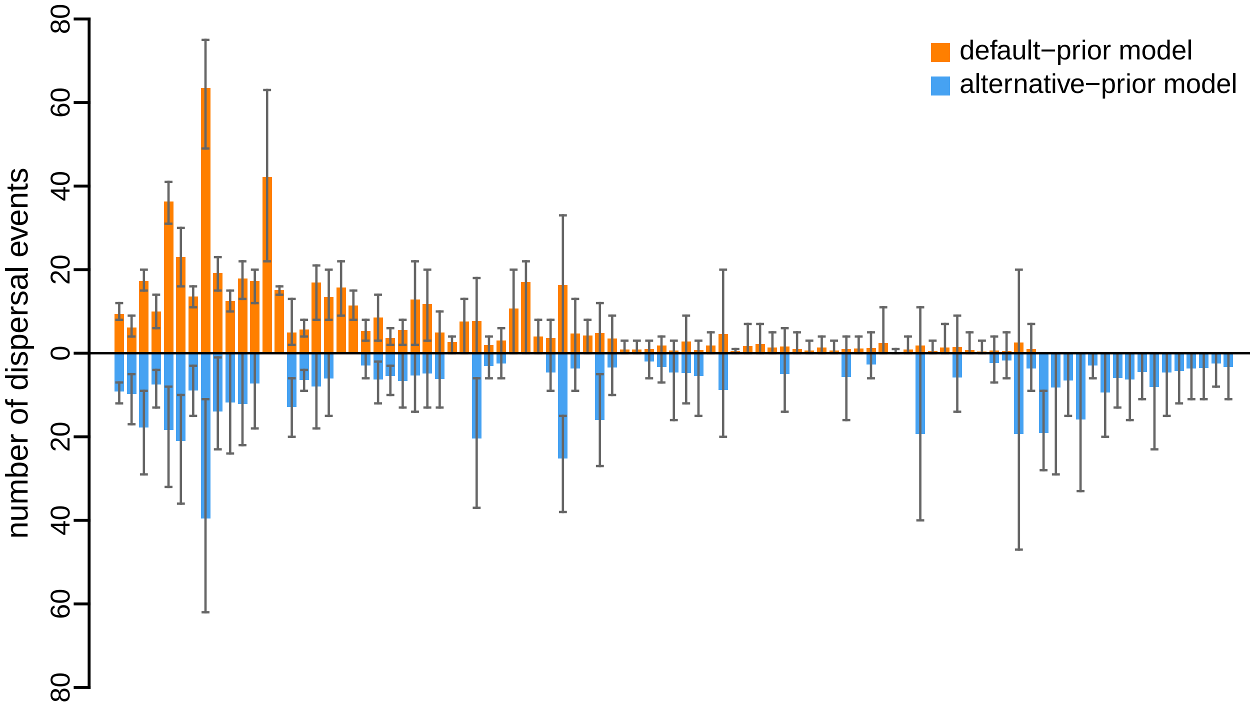


**Figure S146: The impact of prior choice on the inferred number of dispersal events between each pair of areas for the SARS-CoV-2 Global dataset.** The reflected bar plot depicts the number of dispersal events inferred under the default (orange) and alternative (blue) prior models for the SARS-CoV-2 Global dataset. Each bar indicates the posterior-mean number of dispersal events between a pair of areas; whiskers indicate the 95% credible interval. Note that only the number of dispersal events over the "significant" dispersal routes (*i.e.*, $2 \ln \mathrm{BF} > 2$) are figured.

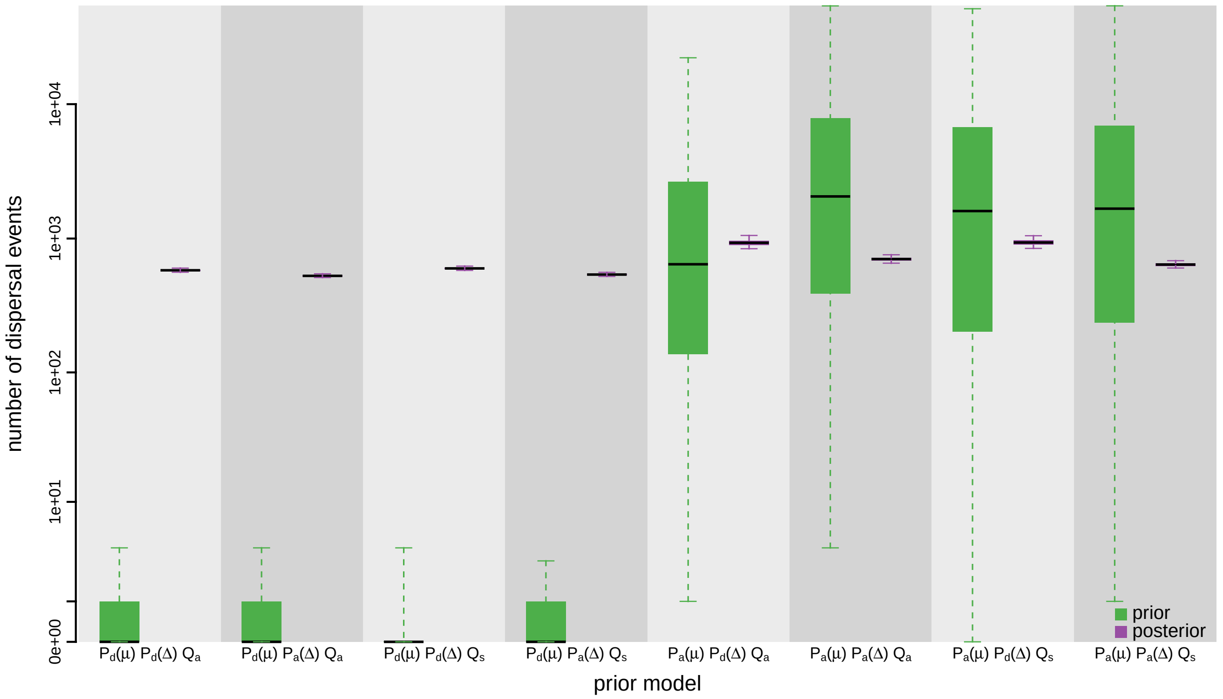


**Figure S147: The impact of prior choice on the inferred total number of dispersal events between all areas for the SARS-CoV-2 Global dataset.** Each pair of boxplots depicts the specified prior (green) and corresponding posterior estimate (purple) of the number of dispersal events between all areas under each prior model: the center of each box indicates the posterior-median number of dispersal events; the box and whiskers indicate the corresponding 50% and 95% credible intervals, respectively. Model notation follows the description in Table S16.

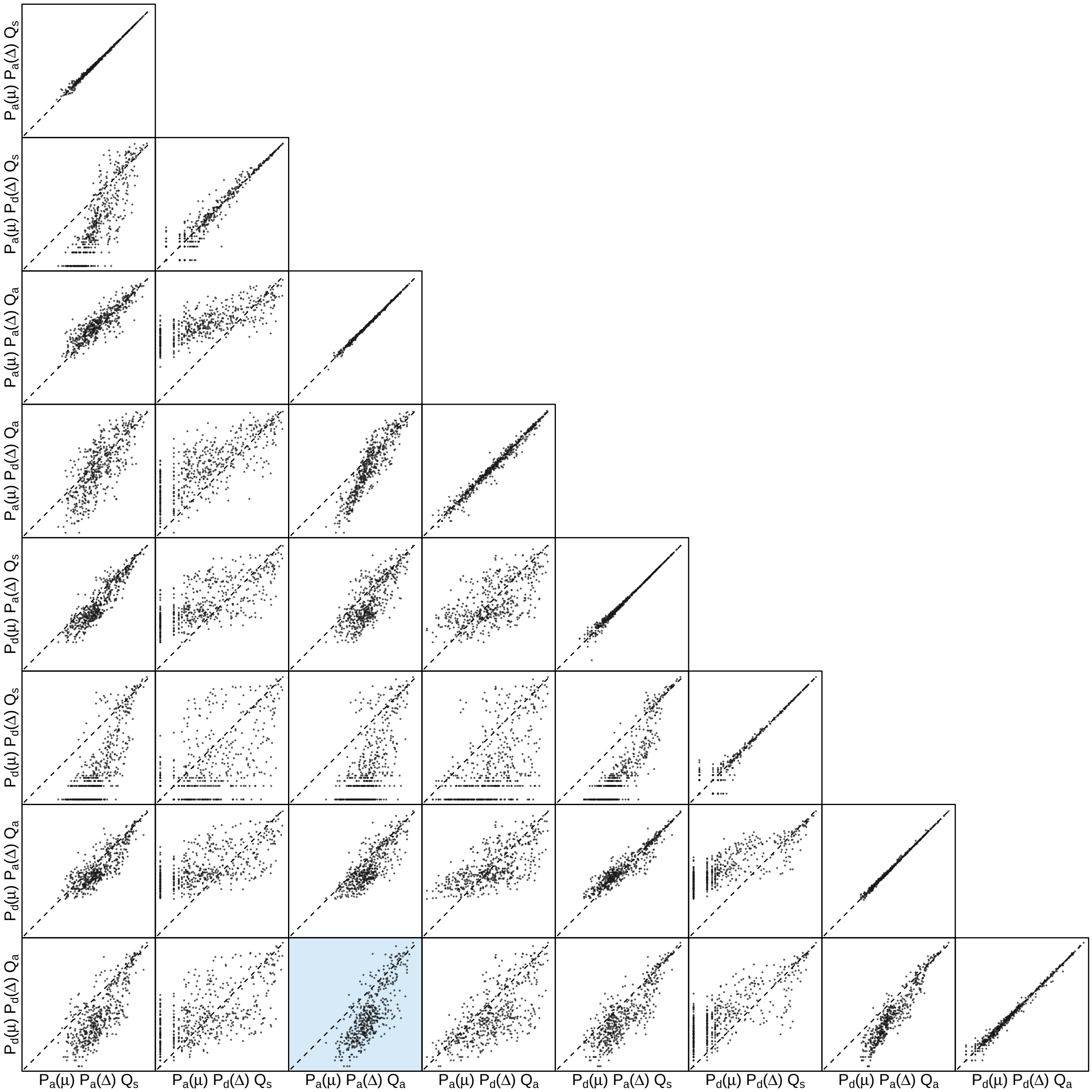


**Figure S148: The impact of prior choice on the inferred number of dispersal events between each pair of areas for the SARS-CoV-2 Global dataset.** Each cell of the plot compares the inferred number of pairwise dispersal events between each pair of prior models. *x*- and *y*-axis of each cell are both on log scale. Diagonal cells are comparisons between half and the other half of the replicates under the same prior model, assessing the convergence of MCMC simulations; off-diagonal cells are comparisons between different prior models, demonstrating the impact of the prior model on the inferred number of pairwise dispersal events. The shaded cell corresponds to the comparison between the preferred default- and the preferred alternative-prior models (*c.f.*, Figure S146). Model notation follows the description in Table S16.

###### S3.3.7 SARS-CoV-2 B.1.1.7 US Dataset

Alpert et al. (2021) explored the introduction, establishment, and geographic dispersal dynamics of the B.1.1.7 variant of SARS-CoV-2 in the United States. The authors produced a dataset containing 1908 SARS-CoV-2 genomic sequences, subsampled from all the B.1.1.7 variant genomes available on GISAID (Shu and McCauley 2017) as of February 26, 2021, focussing on the samples from the US. They discretized the geographic space by states (for the US samples) or by Europe or not Europe (for the international samples), resulting in 22 (20 US and 2 international) geographic areas. Details about the data and the curation procedures can be found in Alpert et al. (2021).

Here we explored the impact of prior choice on phylodynamic inference of biogeographic history using this dataset. Following the original study, we conditioned on the summary tree (obtained directly from the Github repository of Alpert et al. 2021) inferred without the geographic data (*i.e.*, using the sequence data and the sampling time for each sequence) to perform the biogeographic inference. This summary tree and the file containing the associated sampling time and location of each sequence are available in our GitHub and Dryad repositories.

The MCMC simulations used in the second step of our sequential analyses and of the analyses used to estimate marginal likelihoods under each prior model are described above in Section S3.1. Details of these analyses are available in the XML scripts included in our GitHub and Dryad repositories.

**Table S17: Marginal-likelihood estimates of the eight prior models for the SARS-CoV-2 B.1.1.7 US dataset.** Columns 2–5 list marginal likelihoods inferred from four replicate analyses. Columns 6–7 list the mean and standard deviation of these marginal-likelihood estimates. The last column lists the marginal-likelihood estimates computed by combining the samples from the replicate power-posterior MCMC simulations. Candidate models are listed in rows and include all possible combinations of: (1) instantaneous-rate matrices (symmetric, $Q_s$ or asymmetric, $Q_a$); (2) priors on the average dispersal rate [default, $P_d(\mu)$ or alternative, $P_a(\mu)$], and; (3) priors on the number of dispersal routes [default, $P_d(\Delta)$ or alternative, $P_a(\Delta)$]. The preferred default- and alternative-prior models are indicated in bold text.

| Model | replicate1 | replicate2 | replicate3 | replicate4 | mean | sd | combined |
|---|---|---|---|---|---|---|---|
| $\mathbf{P_d(\mu) Q_a P_d(\Delta)}$ | **-1983.46** | **-1983.59** | **-1984.04** | **-1983.21** | **-1983.57** | **0.35** | **-1983.46** |
| $P_d(\mu) Q_a P_a(\Delta)$ | -1966.52 | -1966.96 | -1967.14 | -1967.98 | -1967.15 | 0.61 | -1966.99 |
| $P_d(\mu) Q_s P_d(\Delta)$ | -2050.76 | -2050.91 | -2050.71 | -2050.22 | -2050.65 | 0.30 | -2050.61 |
| $P_d(\mu) Q_s P_a(\Delta)$ | -1978.79 | -1978.72 | -1978.59 | -1978.97 | -1978.77 | 0.16 | -1978.75 |
| $P_a(\mu) Q_a P_d(\Delta)$ | -1730.19 | -1729.43 | -1728.80 | -1729.41 | -1729.46 | 0.57 | -1729.21 |
| $\mathbf{P_a(\mu) Q_a P_a(\Delta)}$ | **-1720.99** | **-1721.28** | **-1721.09** | **-1721.16** | **-1721.13** | **0.12** | **-1721.10** |
| $P_a(\mu) Q_s P_d(\Delta)$ | -1844.13 | -1845.14 | -1845.60 | -1846.47 | -1845.33 | 0.97 | -1845.14 |
| $P_a(\mu) Q_s P_a(\Delta)$ | -1791.29 | -1791.15 | -1790.99 | -1791.46 | -1791.22 | 0.20 | -1791.21 |

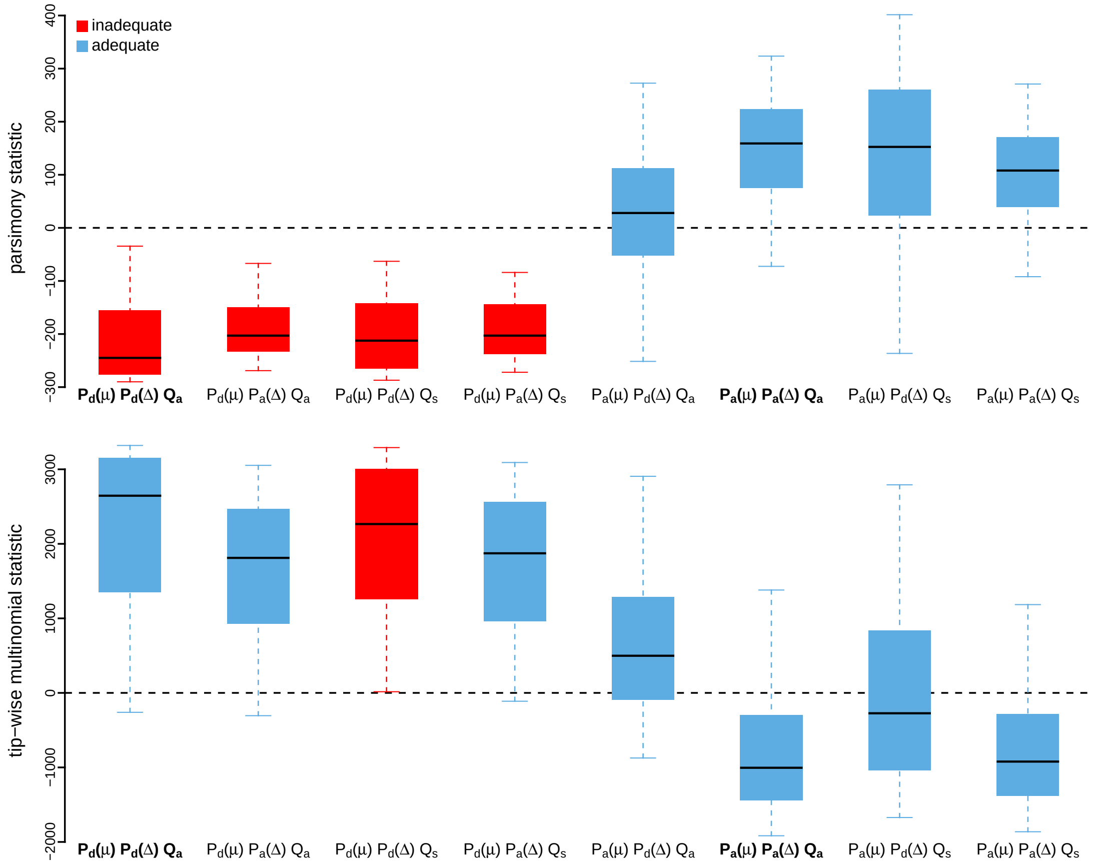


**Figure S149: Posterior-predictive distributions of the parsimony statistic (top panel) and the tip-wise multinomial statistic (bottom panel) under each of the eight prior models for the SARS-CoV-2 B.1.1.7 US dataset.** Boxplots depict the posterior-predictive distributions of the statistic under each of the eight candidate prior models: the center of each box is the median predictive value of the summary statistic; the box and whiskers indicate the corresponding 50% and 95% posterior-predictive intervals, respectively. The horizontal dashed line indicates when the simulated and observed datasets produce identical value for the summary statistic. A model is judged to be inadequate (*i.e.*, incapable of generating geographic datasets that are similar to the observed data) if its 95% posterior-predictive interval does not overlap with the dashed line. Here the preferred default prior model is inadequate, whereas the preferred alternative prior model is adequate. Notation for the candidate models is described in Table S17; the preferred default- and alternative-prior models are indicated in bold text.

*The Impact of Prior Choice on Average Dispersal Rate*

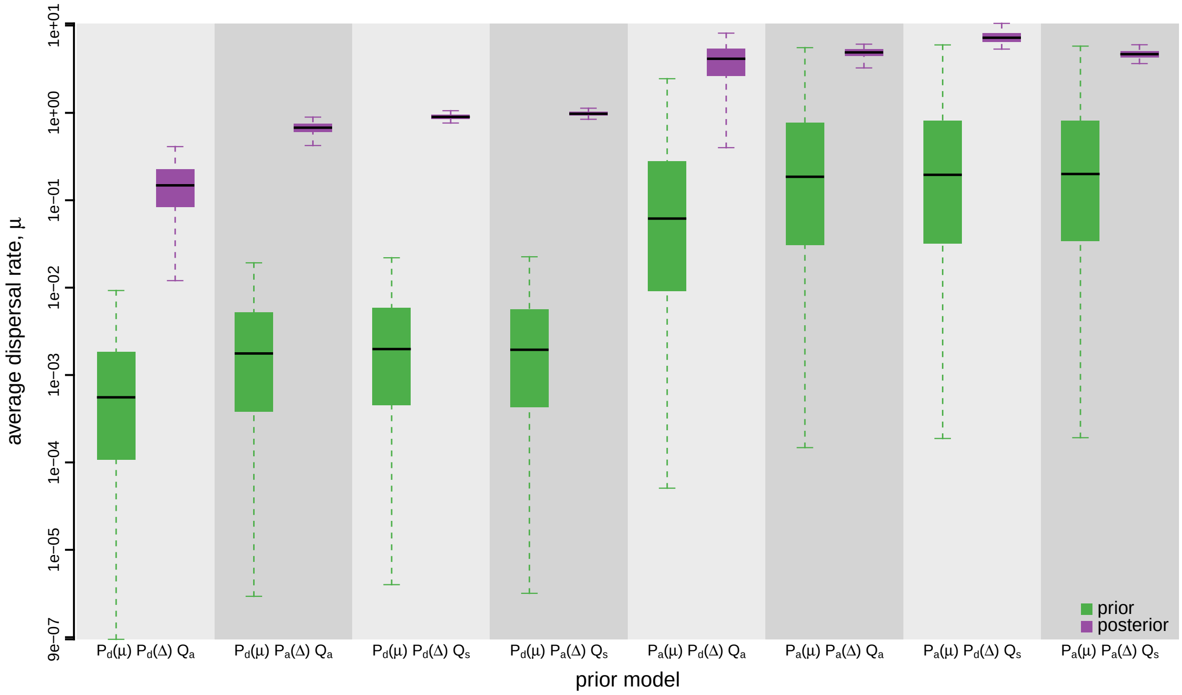


**Figure S150: The impact of prior choice on the average rate of dispersal for the SARS-CoV-2 B.1.1.7 US dataset.** Each pair of boxplots depicts the specified prior (green) and corresponding posterior estimate (purple) of the average dispersal rate, $\mu$, under each prior model: the center of each box indicates the median rate; the box and whiskers indicate the corresponding 50% and 95% credible intervals, respectively. Notation for the prior models is as described in Table S17.

*The Impact of Prior Choice on Pairwise Dispersal Rates*

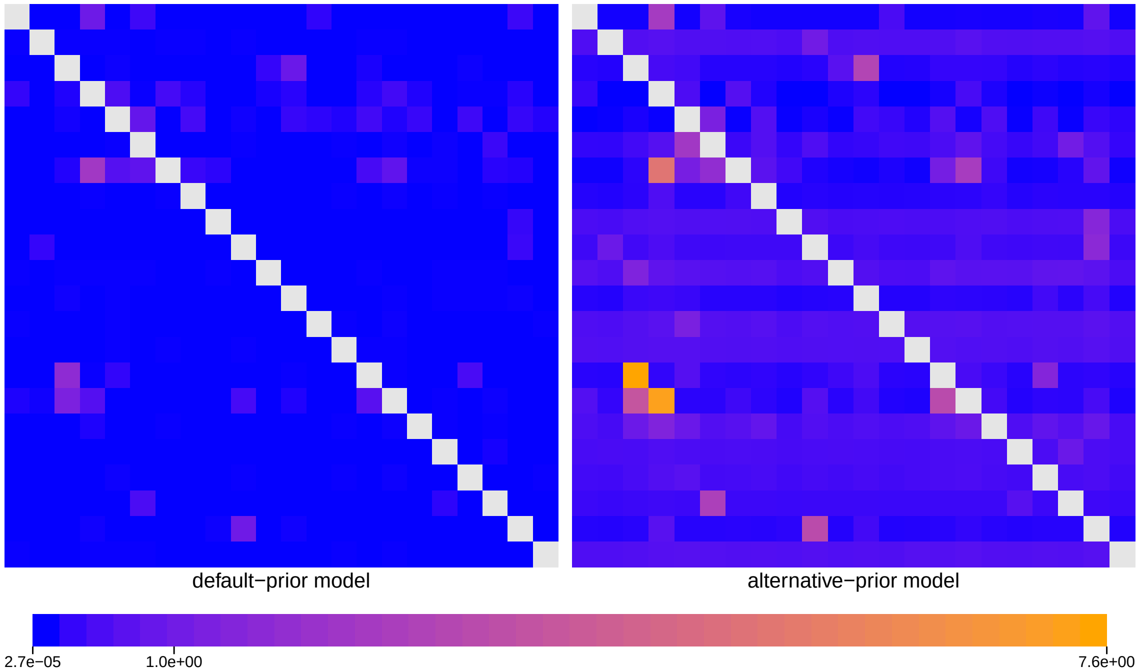


**Figure S151: The impact of prior choice on pairwise dispersal rates for the SARS-CoV-2 B.1.1.7 US dataset.** Heatmaps summarize posterior-mean estimates of the instantaneous rate of dispersal between each pair of geographic areas, $q_{ij}$, under the default (left) and alternative (right) prior models.

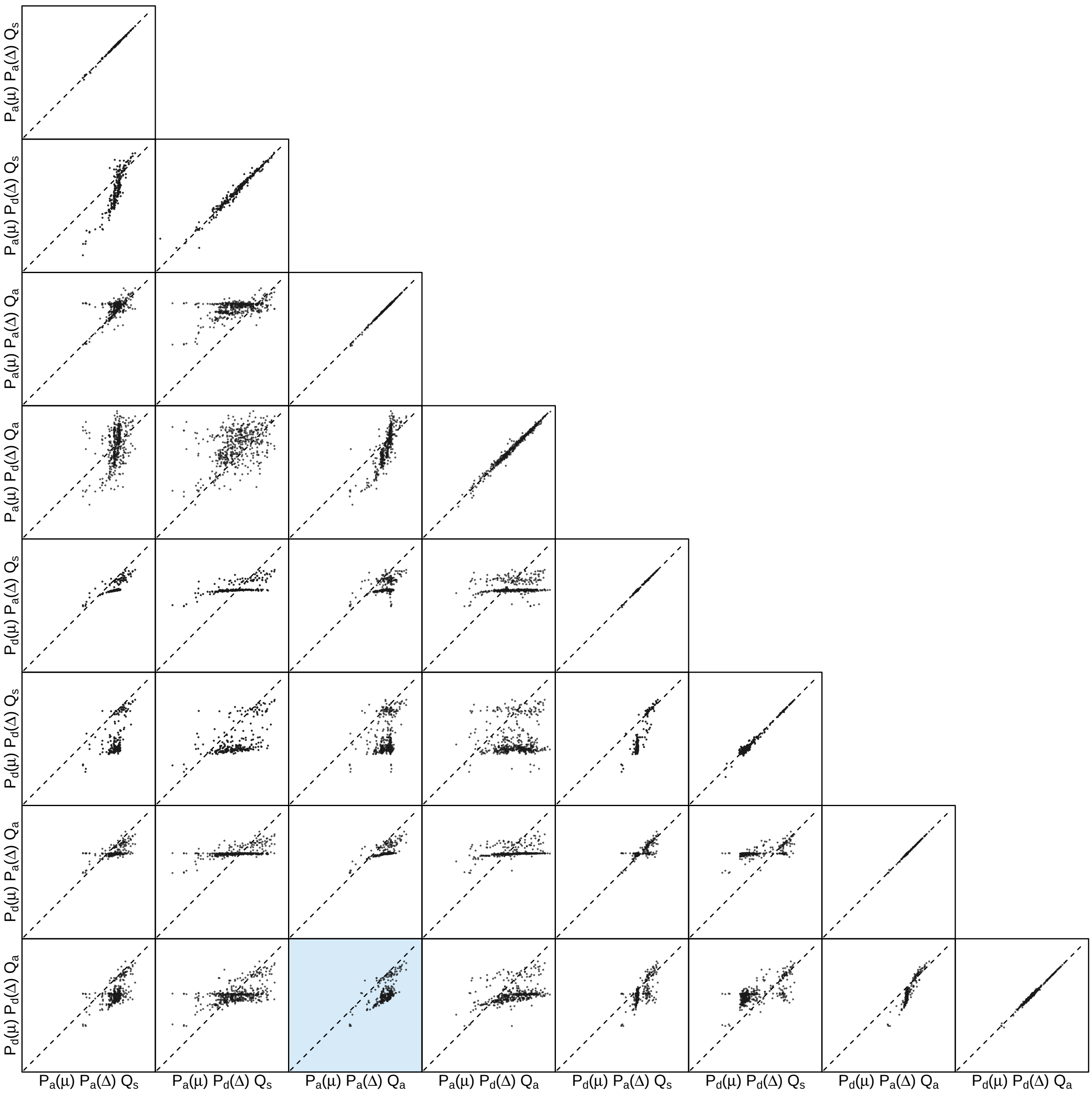


**Figure S152: The impact of prior choice on pairwise dispersal rates for the SARS-CoV-2 B.1.1.7 US dataset.** Each cell of the plot compares posterior-mean estimate of the rate of dispersal between each pair of geographic areas, $q_{ij}$, between each pair of prior models. $x$- and $y$-axis of each cell are both on log scale. Diagonal cells are comparisons between half and the other half of the replicates under the same prior model, assessing the convergence of MCMC simulations; off-diagonal cells are comparisons between different prior models, demonstrating the impact of the prior model on the estimates of pairwise dispersal rates. The shaded cell corresponds to the comparison between the preferred default- and the preferred alternative-prior models (*c.f.*, Figure S151). Notation for the prior models is as described in Table S17.

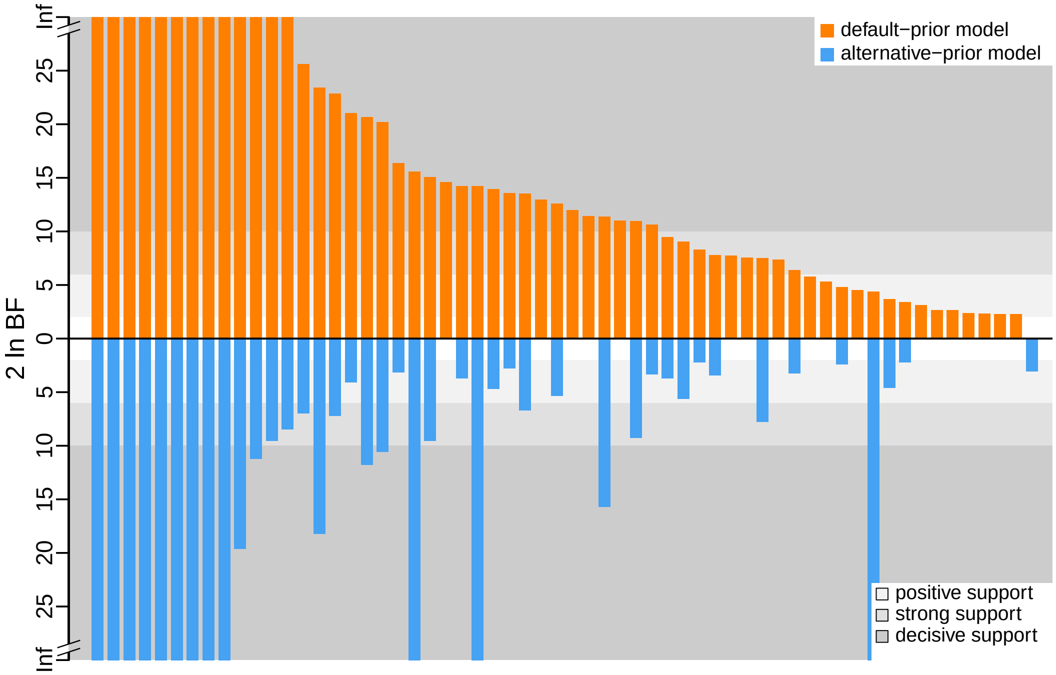


**Figure S153: The impact of prior choice on the inferred support for dispersal routes for the SARS-CoV-2 B.1.1.7 US dataset.** We compare the evidential support for each dispersal route for the SARS-CoV-2 B.1.1.7 US dataset under the default (orange) and alternative (blue) prior models. Each bar indicates the $2 \ln \mathrm{BF}$ for the corresponding dispersal route between two areas; only supported dispersal routes (*i.e.*, $2 \ln \mathrm{BF} > 2$) are plotted.

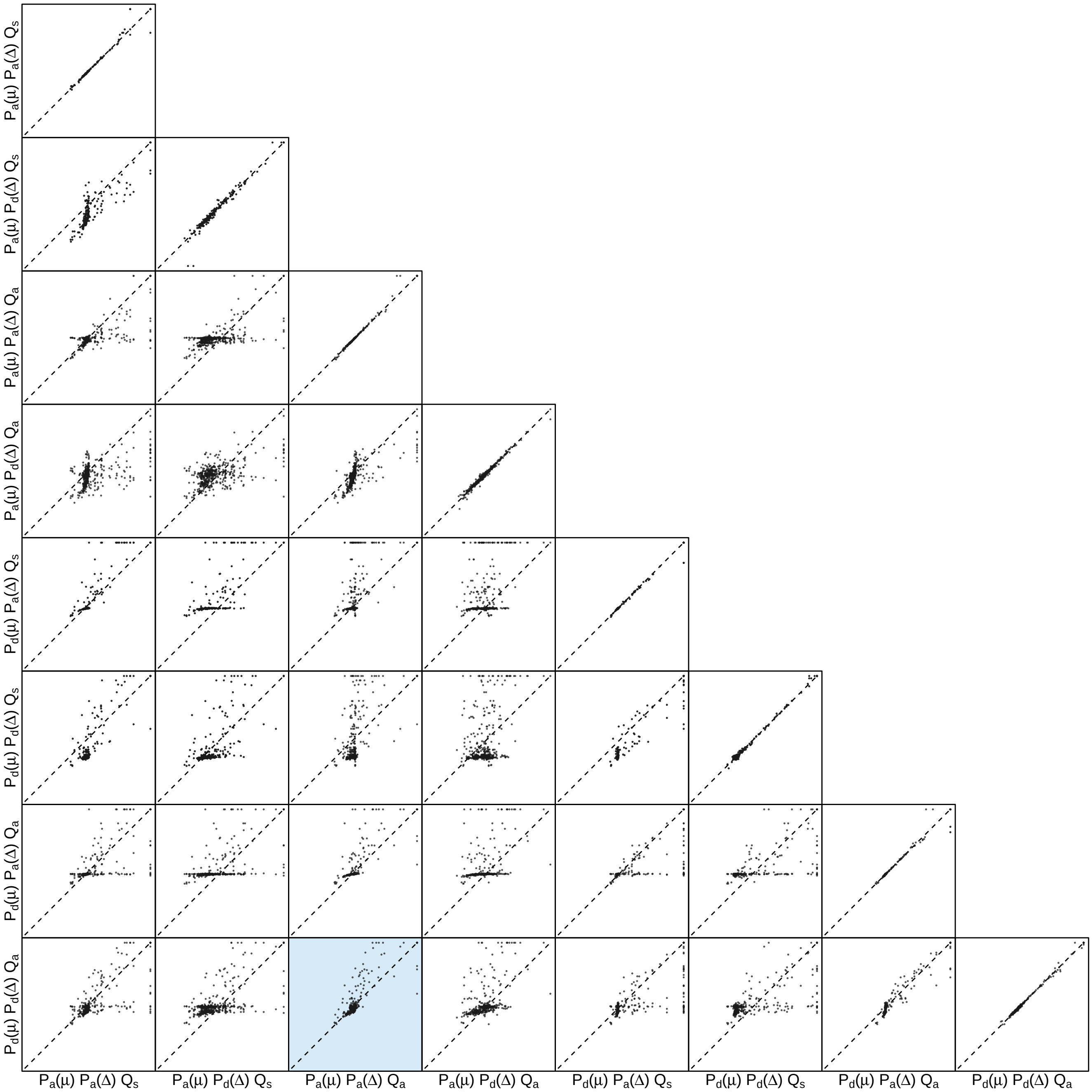


**Figure S154: The impact of prior choice on the inferred support for dispersal routes for the SARS-CoV-2 B.1.1.7 US dataset.** Each cell of the plot compares the inferred support ($2 \ln \mathrm{BF}$) for pairwise dispersal routes between each pair of prior models. Diagonal cells are comparisons between half and the other half of the replicates under the same prior model, assessing the convergence of MCMC simulations; off-diagonal cells are comparisons between different prior models, demonstrating the impact of the prior model on the inferred support for pairwise dispersal routes. The shaded cell corresponds to the comparison between the preferred default- and the preferred alternative-prior models (*c.f.*, Figure S153). Notation for the prior models is as described in Table S17.

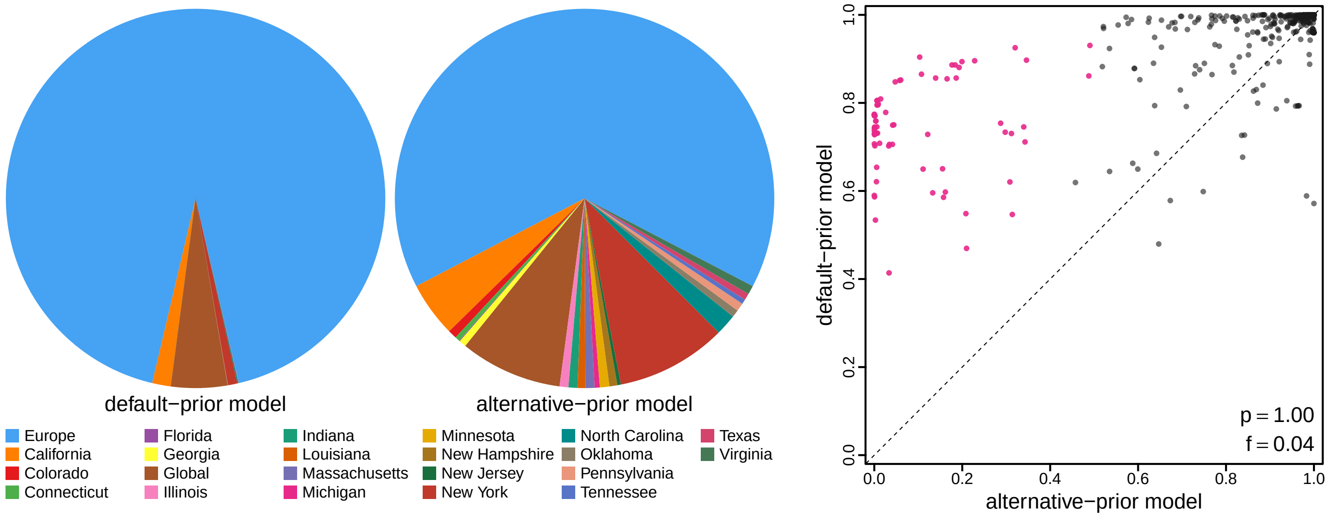


**Figure S155: The impact of prior choice on ancestral-area estimates for the SARS-CoV-2 B.1.1.7 US dataset.** The left panel compares the posterior probability of each ancestral area at the root node under the default- and alternative-prior models for the SARS-CoV-2 B.1.1.7 US dataset. The right panel plots the posterior probability of the most probable ancestral area under the default-prior model for each node in the MCC tree (y-axis) against the corresponding posterior probability of that area under the alternative-prior model (x-axis). Pink dots represent the internal nodes where the MAP ancestral area inferred under the default-prior model differs from that inferred under the alternative-prior models. The statistic $p$ denotes the fraction of internal nodes that are shared under the default- and alternative-prior models; $f$, is the fraction of shared nodes where the MAP ancestral area differs under the default- and alternative-prior models. Note that the posterior probabilities of the MAP ancestral area under the default-prior model are generally higher than those under the alternative-prior model (*i.e.*, the default-prior model tends to mask uncertainty in the ancestral-area estimates).

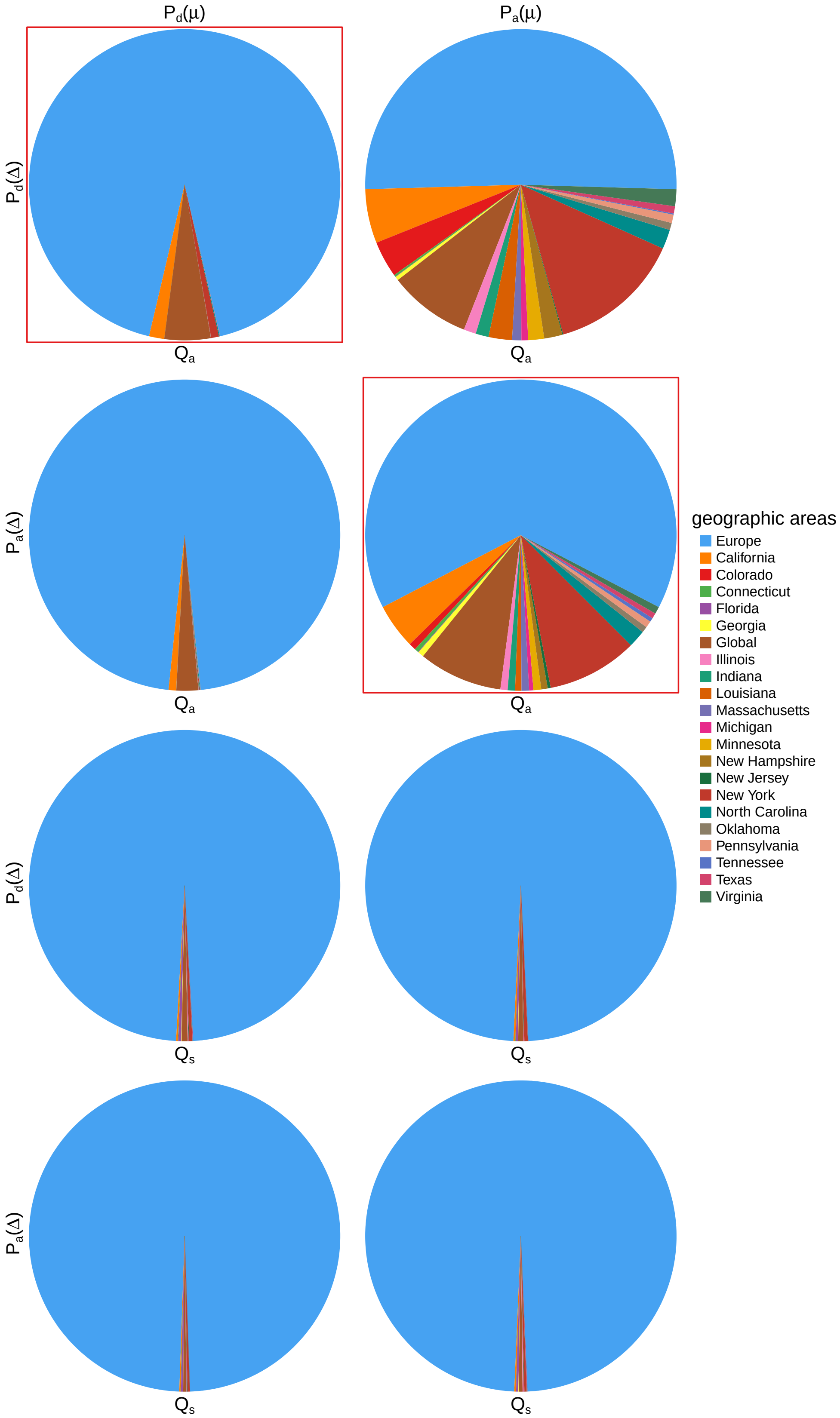


**Figure S156: The impact of prior choice on ancestral-area estimates at the root node for the SARS-CoV-2 B.1.1.7 US dataset.** The posterior probability of each ancestral area at the root node under each prior model for the SARS-CoV-2 B.1.1.7 US dataset. The pie chart enclosed in red corresponds to the comparison between the preferred default- and the preferred alternative-prior models (*c.f.*, Figure S155, left panel). Model notation follows the description in Table S17.

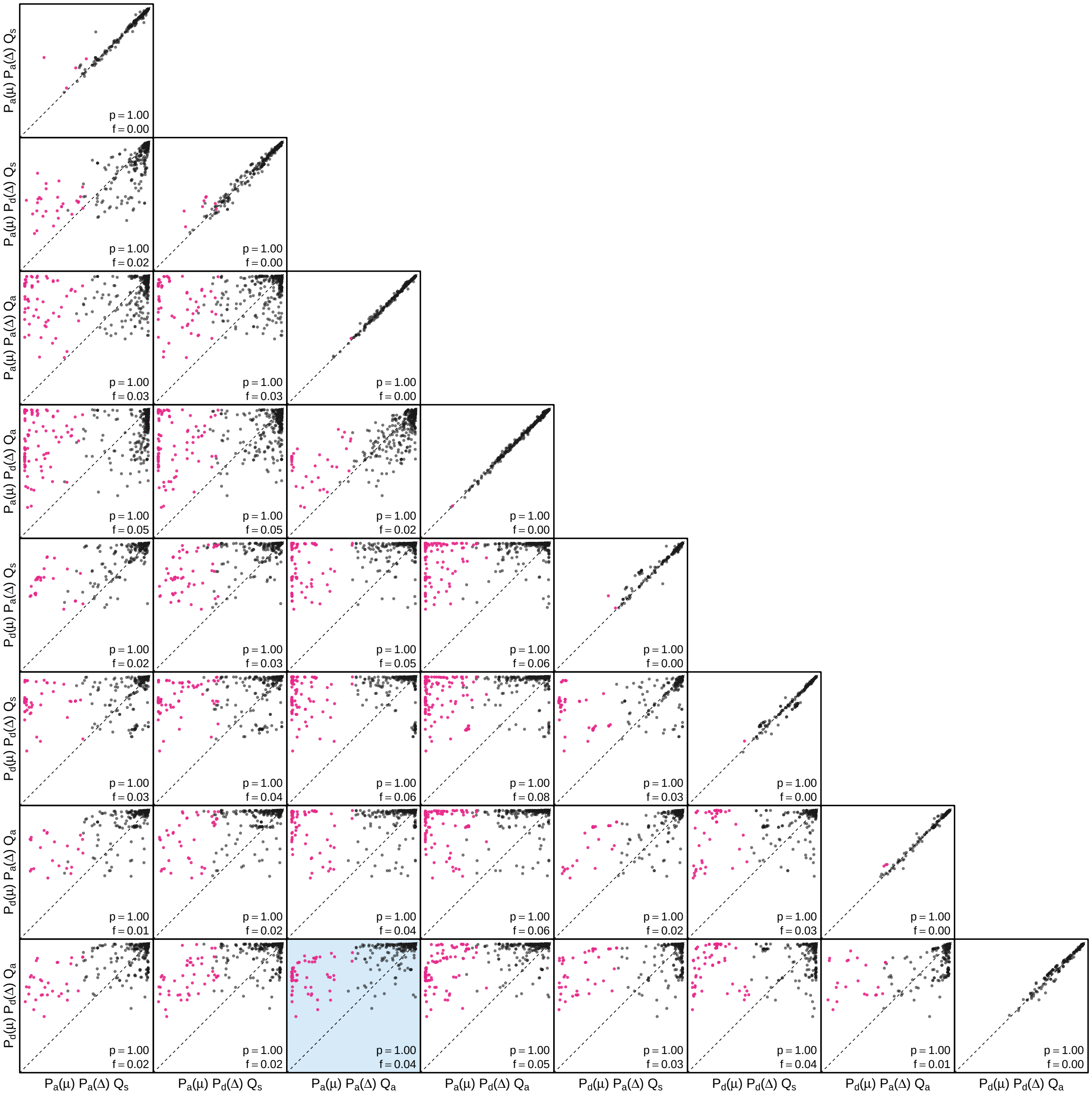


**Figure S157: The impact of prior choice on the MAP ancestral-area estimates at internal nodes for the SARS-CoV-2 B.1.1.7 US dataset.** Pairwise scatter plots compare the posterior probability of the MAP ancestral area inferred under each combination of prior models. Diagonal cells are comparisons between two replicates under the same prior model (assessing the convergence of MCMC simulations); off-diagonal cells are comparisons between different prior models, demonstrating the impact of the prior model on both the posterior probability and the identity of the MAP ancestral-area estimates at internal nodes. Pink dots represent the internal nodes where the MAP ancestral area inferred under the default-prior model differs from that inferred under the alternative-prior models. The statistic $p$ denotes the fraction of internal nodes that are shared under the default- and alternative-prior models; $f$ is the fraction of shared nodes where the MAP ancestral area differs under the default- and alternative-prior models. The shaded cell corresponds to the comparison between the preferred default- and the preferred alternative-prior models (*c.f.*, Figure S155, right panel). Model notation follows the description in Table S17.

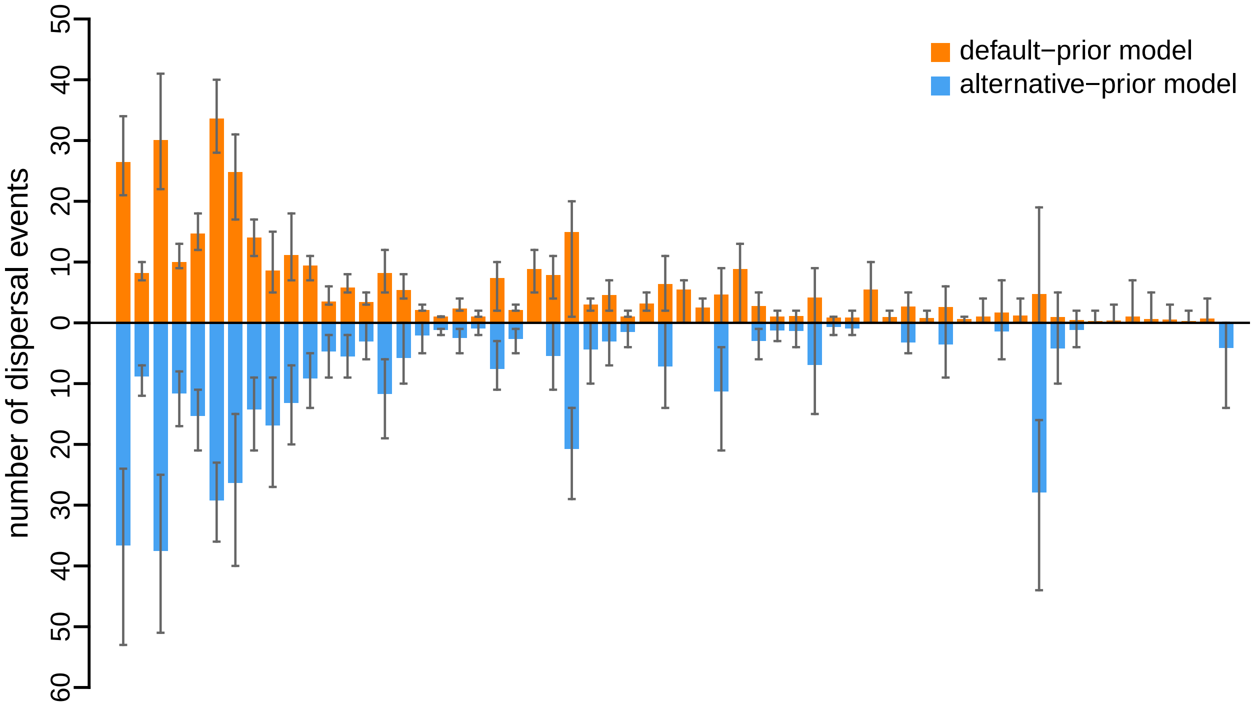


**Figure S158: The impact of prior choice on the inferred number of dispersal events between each pair of areas for the SARS-CoV-2 B.1.1.7 US dataset.** The reflected bar plot depicts the number of dispersal events inferred under the default (orange) and alternative (blue) prior models for the SARS-CoV-2 B.1.1.7 US dataset. Each bar indicates the posterior-mean number of dispersal events between a pair of areas; whiskers indicate the 95% credible interval. Note that only the number of dispersal events over the "significant" dispersal routes (*i.e.*, $2 \ln \mathrm{BF} > 2$) are figured.

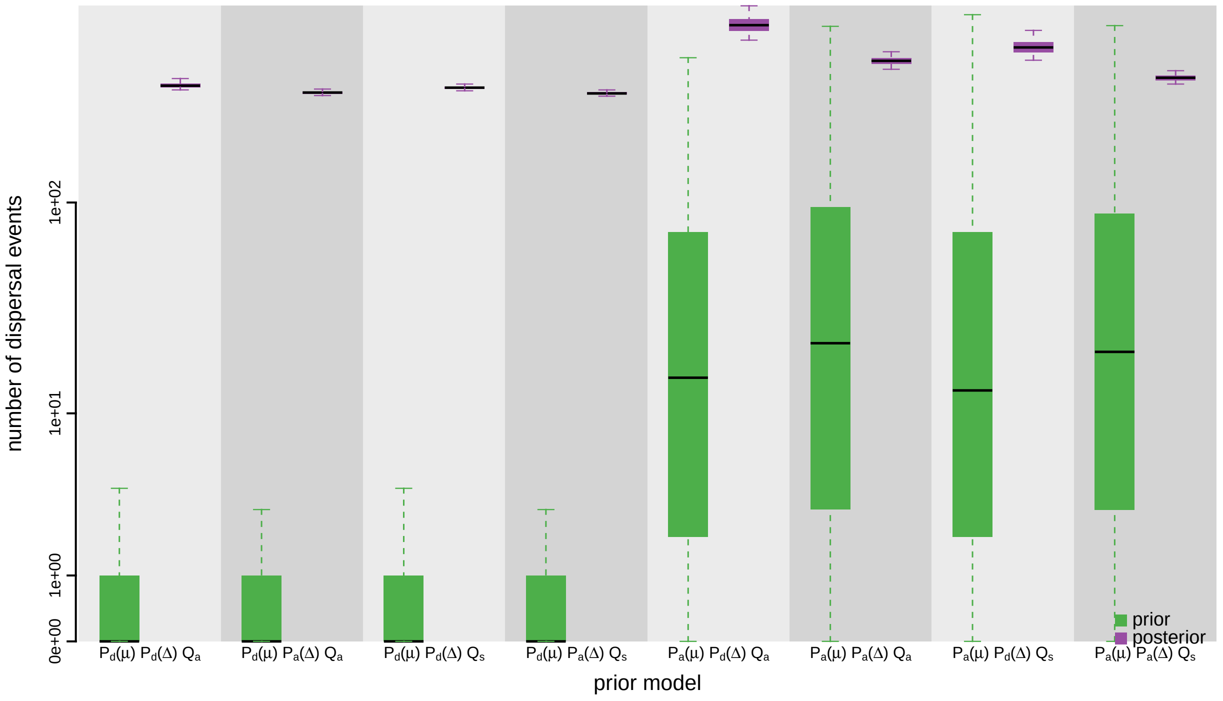


**Figure S159: The impact of prior choice on the inferred total number of dispersal events between all areas for the SARS-CoV-2 B.1.1.7 US dataset.** Each pair of boxplots depicts the specified prior (green) and corresponding posterior estimate (purple) of the number of dispersal events between all areas under each prior model: the center of each box indicates the posterior-median number of dispersal events; the box and whiskers indicate the corresponding 50% and 95% credible intervals, respectively. Model notation follows the description in Table S17.

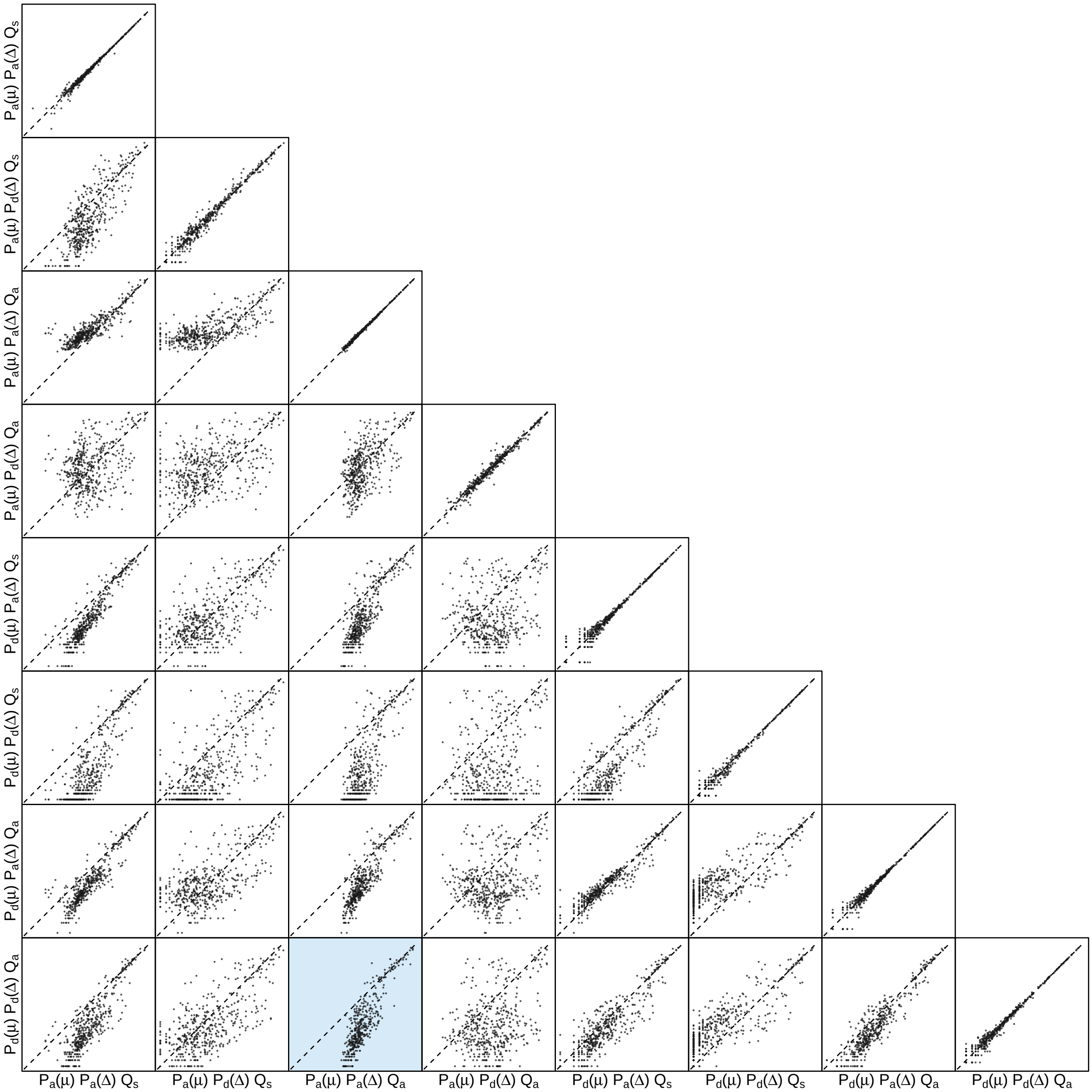


**Figure S160: The impact of prior choice on the inferred number of dispersal events between each pair of areas for the SARS-CoV-2 B.1.1.7 US dataset.** Each cell of the plot compares the inferred number of pairwise dispersal events between each pair of prior models. *x*- and *y*-axis of each cell are both on log scale. Diagonal cells are comparisons between half and the other half of the replicates under the same prior model, assessing the convergence of MCMC simulations; off-diagonal cells are comparisons between different prior models, demonstrating the impact of the prior model on the inferred number of pairwise dispersal events. The shaded cell corresponds to the comparison between the preferred default- and the preferred alternative-prior models (*c.f.*, Figure S158). Model notation follows the description in Table S17.

###### S3.3.8 SARS-CoV-2 Brazil

Candido et al. (2020) investigated the early spread of the SARS-CoV-2 epidemic in Brazil and the efficacy of mitigation measures on limiting that spread. The authors combined newly sequenced SARS-CoV-2 genomes sampled from Brazil with the genomes available on GISAID (Shu and McCauley 2017) as April 24, 2020 to produce a SARS-CoV-2 sequence dataset focussing on the epidemic in Brazil. This dataset contains 1182 SARS-CoV-2 genomes, including 490 sampled from Brazil and 692 subsampled from the sequences collected outside of Brazil. The authors then discretized the geographic space with three different ways, generating three geographic datasets, including: (1) Brazil or not Brazil (totaling two areas, scheme A); (2) five Brazilian regions ("Southeast", "Northeast", "North", "Centre-West", and "South") and five international regions (North America, Europe, Asia, Oceania, and Africa) (totaling 10 areas, scheme B), and; (3) 21 Brazil states and one other area representing the sampling location for all the international sequences (totaling 22 areas, scheme C). Details about the data and the curation procedures can be found in Candido et al. (2020).

Here we explored the impact of prior choice on phylodynamic inference of biogeographic history using the second (SchemeB) and third (SchemeC) geographic datasets. We acquired the marginal posterior probability distribution of phylogenies (inferred from the sequence data and used to perform sequential analyses in Candido et al. 2020) directly from the Dryad repository of the original study. This posterior distribution included 1000 trees, which was then treated as the prior distribution of phylogenies in the second step of the sequential phylodynamic inference. The `trees` file containing this distribution is available in our GitHub and Dryad repositories. We acquired the sampling geographic location data from the XML scripts provided by the original study; this sampling-area data are available in our GitHub and Dryad repositories.

The MCMC simulations used in the second step of our sequential analyses and of the analyses used to estimate marginal likelihoods under each prior model are described above in Section S3.1. Details of these analyses are available in the XML scripts included in our GitHub and Dryad repositories.

##### S3.3.8.1 SchemeB Dataset

*The Impact of Prior Choice on Biogeographic Model Fit*

**Table S18: Marginal-likelihood estimates of each of the eight prior models for SARS-CoV-2 Brazil SchemeB dataset.** Columns 2–5 list marginal likelihoods inferred from four replicate analyses. Columns 6–7 list the mean and standard deviation of these marginal-likelihood estimates. The last column lists the marginal-likelihood estimates computed by combining the samples from the replicate power-posterior MCMC simulations. Candidate models are listed in rows and include all possible combinations of: (1) instantaneous-rate matrices (symmetric, $Q_s$ or asymmetric, $Q_a$); (2) priors on the average dispersal rate [default, $P_d(\mu)$ or alternative, $P_a(\mu)$], and; (3) priors on the number of dispersal routes [default, $P_d(\Delta)$ or alternative, $P_a(\Delta)$]. The preferred default- and alternative-prior models are indicated in bold text.

| Model | replicate1 | replicate2 | replicate3 | replicate4 | mean | sd | combined |
|---|---|---|---|---|---|---|---|
| $\mathbf{P_d(\mu)Q_aP_d(\Delta)}$ | **-1754.37** | **-1754.32** | **-1754.21** | **-1754.52** | **-1754.35** | **0.13** | **-1754.35** |
| $P_d(\mu)Q_aP_a(\Delta)$ | -1745.85 | -1745.79 | -1745.86 | -1745.85 | -1745.84 | 0.03 | -1745.83 |
| $P_d(\mu)Q_sP_d(\Delta)$ | -1783.18 | -1782.99 | -1783.00 | -1783.05 | -1783.06 | 0.09 | -1783.05 |
| $P_d(\mu)Q_sP_a(\Delta)$ | -1768.00 | -1768.00 | -1767.99 | -1767.95 | -1767.99 | 0.02 | -1767.98 |
| $P_a(\mu)Q_aP_d(\Delta)$ | -1550.79 | -1550.83 | -1550.75 | -1550.71 | -1550.77 | 0.05 | -1550.77 |
| $\mathbf{P_a(\mu)Q_aP_a(\Delta)}$ | **-1536.90** | **-1536.98** | **-1536.88** | **-1536.91** | **-1536.92** | **0.04** | **-1536.91** |
| $P_a(\mu)Q_sP_d(\Delta)$ | -1601.28 | -1601.34 | -1601.19 | -1601.27 | -1601.27 | 0.06 | -1601.27 |
| $P_a(\mu)Q_sP_a(\Delta)$ | -1577.29 | -1577.52 | -1577.53 | -1577.46 | -1577.45 | 0.11 | -1577.45 |

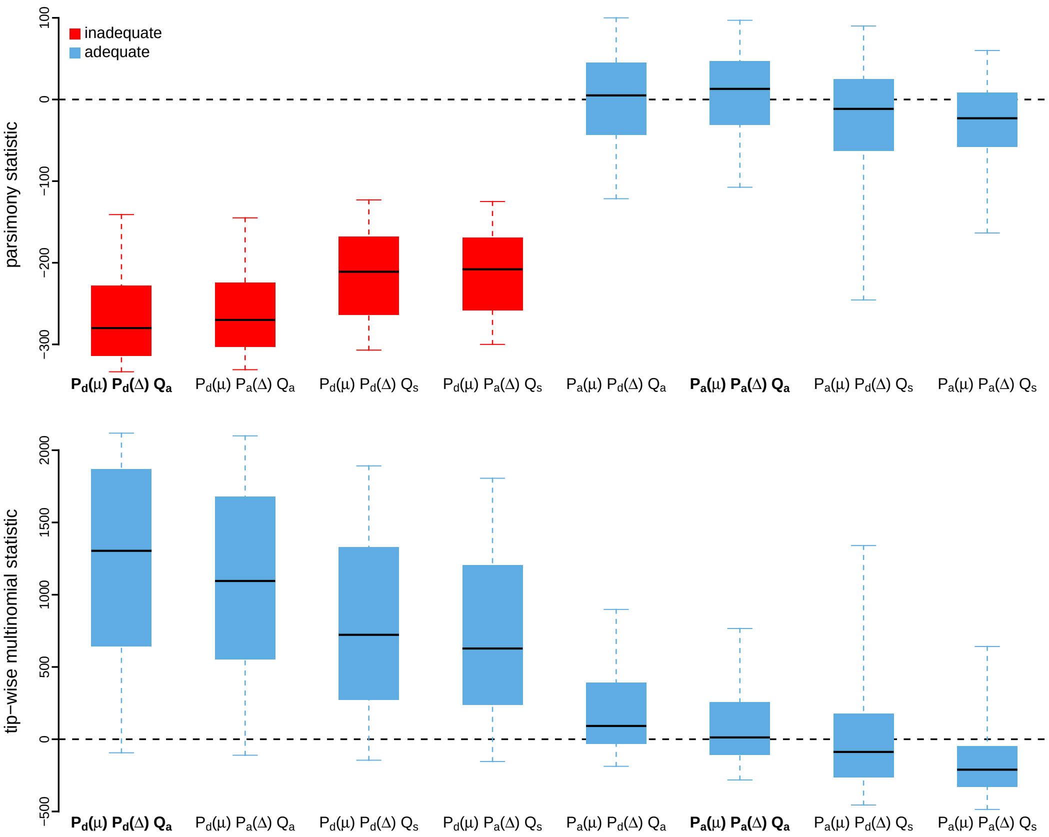


**Figure S161: Posterior-predictive distributions of the parsimony statistic (top panel) and the tip-wise multinomial statistic (bottom panel) under each of the eight prior models for SARS-CoV-2 Brazil SchemeB dataset.** Boxplots depict the posterior-predictive distributions of the statistic under each of the eight candidate prior models: the center of each box is the median predictive value of the summary statistic; the box and whiskers indicate the corresponding 50% and 95% posterior-predictive intervals, respectively. The horizontal dashed line indicates when the simulated and observed datasets produce identical value for the summary statistic. A model is judged to be inadequate (*i.e.*, incapable of generating geographic datasets that are similar to the observed data) if its 95% posterior-predictive interval does not overlap with the dashed line. Here the preferred default prior model is inadequate, whereas the preferred alternative prior model is adequate. Notation for the candidate models is described in Table S18; the preferred default- and alternative-prior models are indicated in bold text.

*The Impact of Prior Choice on Average Dispersal Rate*

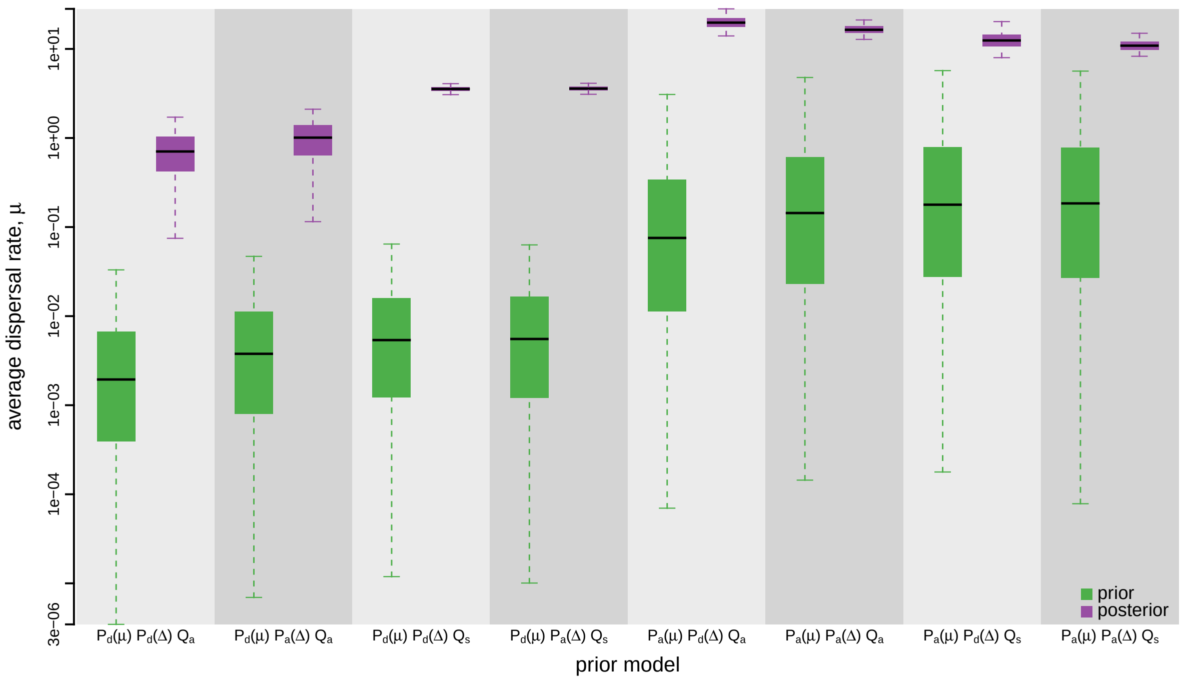


**Figure S162: The impact of prior choice on the average dispersal rate for SARS-CoV-2 Brazil SchemeB dataset.** Each pair of boxplots depicts the specified prior (green) and corresponding posterior estimate (purple) of the average dispersal rate, $\mu$, under each prior model: the center of each box indicates the median rate; the box and whiskers indicate the corresponding 50% and 95% credible intervals, respectively. Notation for the prior models is as described in Table S18.

*The Impact of Prior Choice on Pairwise Dispersal Rates*

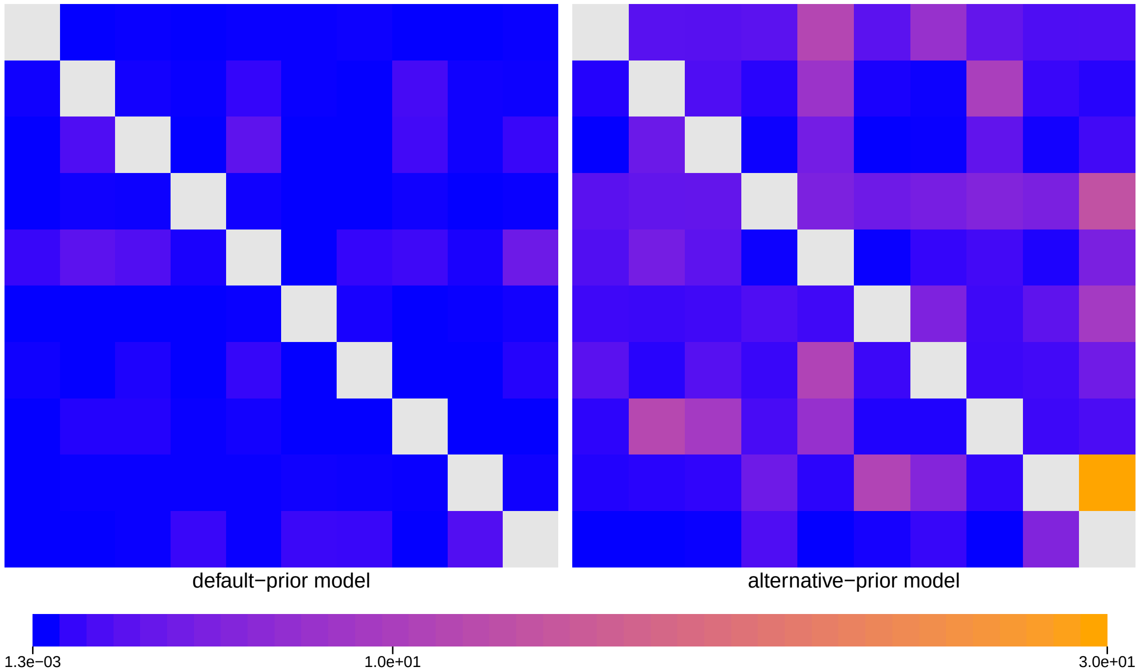


**Figure S163: The impact of prior choice on pairwise dispersal rates for SARS-CoV-2 Brazil SchemeB dataset.** Heatmaps summarize posterior-mean estimates of the instantaneous rate of dispersal between each pair of geographic areas, $q_{ij}$, under the default (left) and alternative (right) prior models.

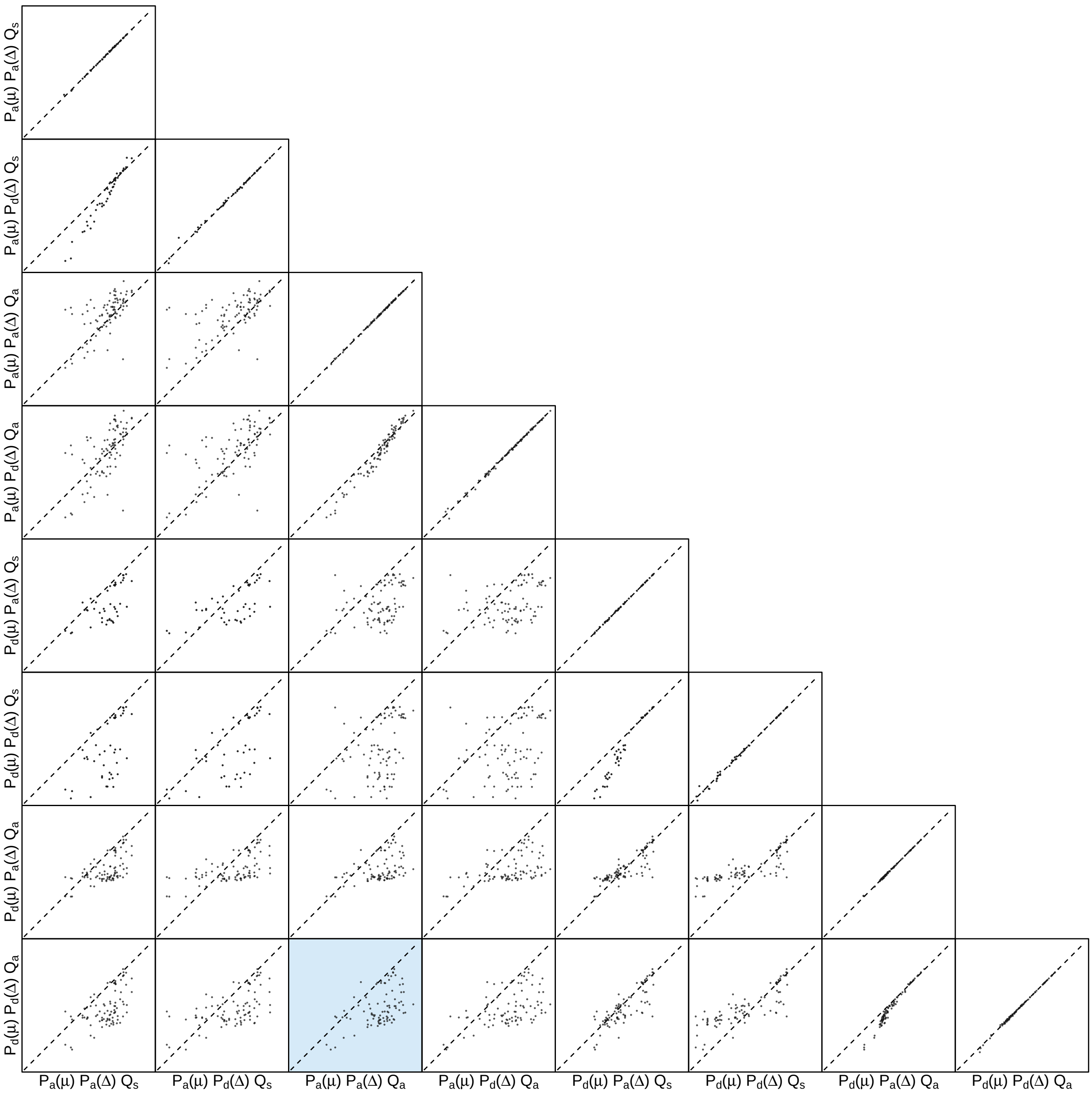


**Figure S164: The impact of prior choice on pairwise dispersal rates for SARS-CoV-2 Brazil SchemeB dataset.** Each cell of the plot compares posterior-mean estimate of the rate of dispersal between each pair of geographic areas, $q_{ij}$, between each pair of prior models. $x$- and $y$-axis of each cell are both on log scale. Diagonal cells are comparisons between half and the other half of the replicates under the same prior model, assessing the convergence of MCMC simulations; off-diagonal cells are comparisons between different prior models, demonstrating the impact of the prior model on the estimates of pairwise dispersal rates. The shaded cell corresponds to the comparison between the preferred default- and the preferred alternative-prior models (*c.f.*, Figure S163). Notation for the prior models is as described in Table S18.

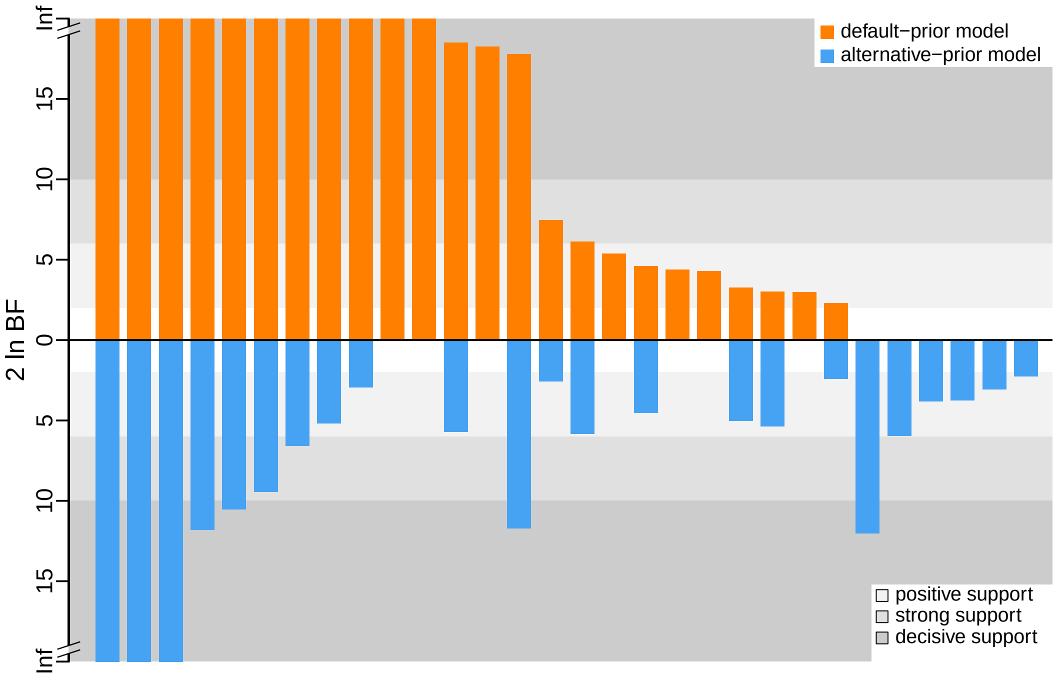


**Figure S165: The impact of prior choice on the inferred support for dispersal routes for SARS-CoV-2 Brazil SchemeB dataset.** We compare the evidential support for each dispersal route for SARS-CoV-2 Brazil SchemeB dataset under the default (orange) and alternative (blue) prior models. Each bar indicates the $2 \ln \mathrm{BF}$ for the corresponding dispersal route between two areas; only supported dispersal routes (*i.e.*, $2 \ln \mathrm{BF} > 2$) are plotted.

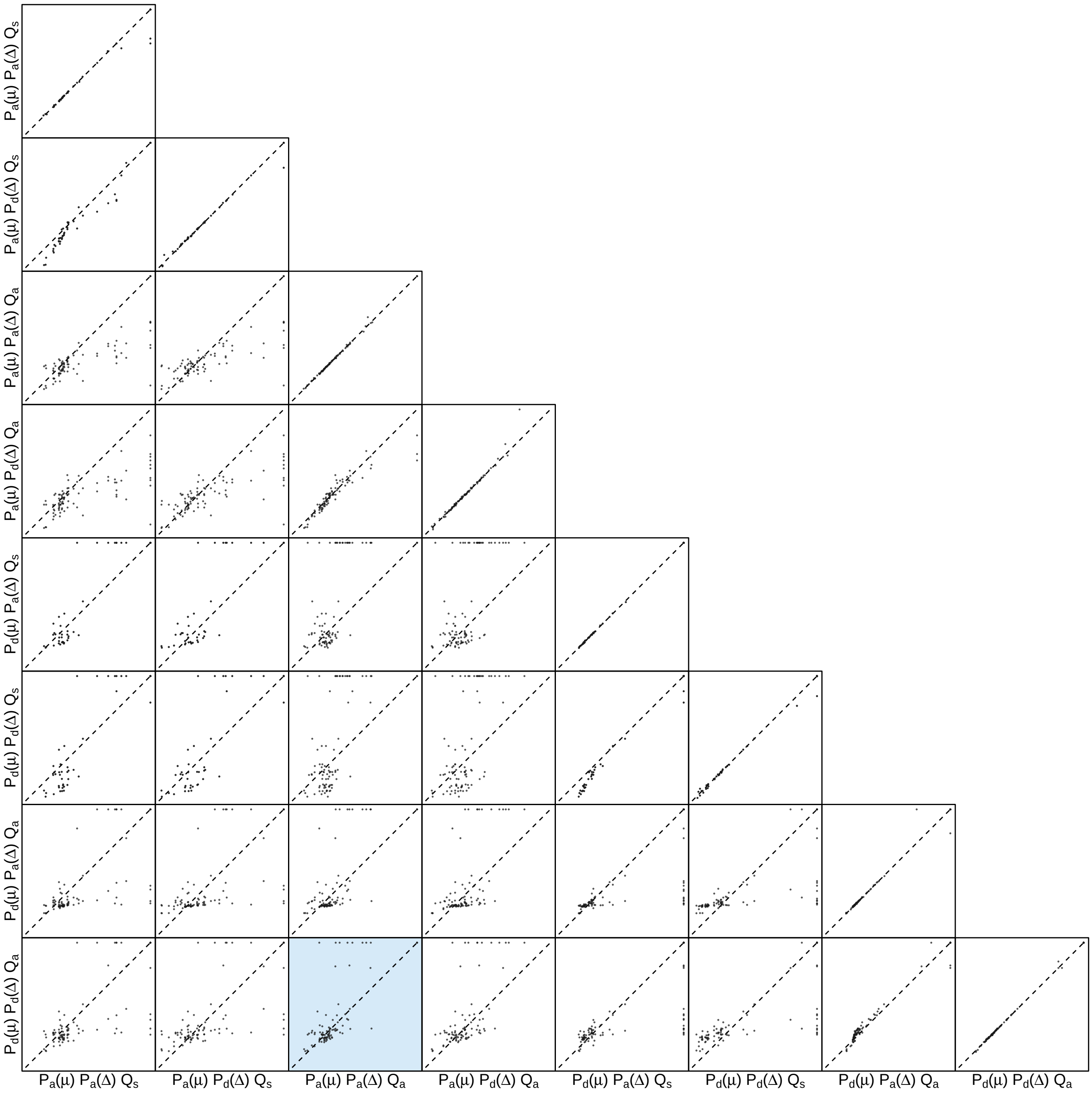


**Figure S166: The impact of prior choice on the inferred support for dispersal routes for SARS-CoV-2 Brazil SchemeB dataset.** Each cell of the plot compares the inferred support ($2 \ln \mathrm{BF}$) for pairwise dispersal routes between each pair of prior models. Diagonal cells are comparisons between half and the other half of the replicates under the same prior model, assessing the convergence of MCMC simulations; off-diagonal cells are comparisons between different prior models, demonstrating the impact of the prior model on the inferred support for pairwise dispersal routes. The shaded cell corresponds to the comparison between the preferred default- and the preferred alternative-prior models (*c.f.*, Figure S165). Notation for the prior models is as described in Table S18.

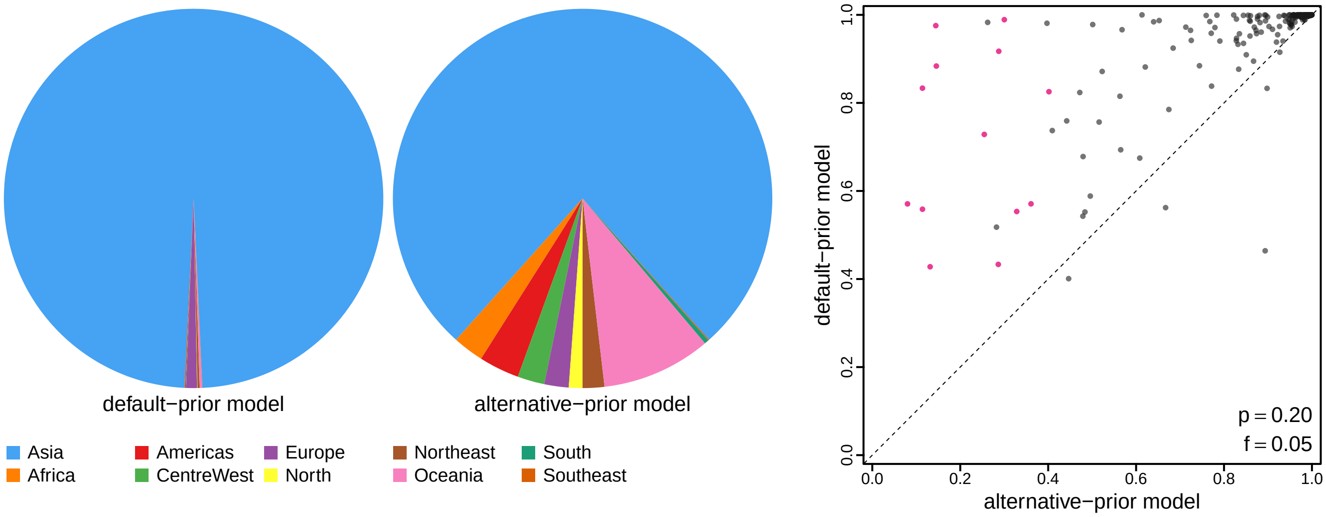


**Figure S167: The impact of prior choice on ancestral-area estimates for SARS-CoV-2 Brazil SchemeB dataset.** The left panel compares the posterior probability of each ancestral area at the root node under the default- and alternative-prior models for SARS-CoV-2 Brazil SchemeB dataset. The right panel plots the posterior probability of the most probable ancestral area under the default-prior model for each node in the MCC tree (y-axis) against the corresponding posterior probability of that area under the alternative-prior model (x-axis). Pink dots represent the internal nodes where the MAP ancestral area inferred under the default-prior model differs from that inferred under the alternative-prior models. The statistic $p$ denotes the fraction of internal nodes that are shared under the default- and alternative-prior models; $f$, is the fraction of shared nodes where the MAP ancestral area differs under the default- and alternative-prior models. Note that the posterior probabilities of the MAP ancestral area under the default-prior model are generally higher than those under the alternative-prior model (*i.e.*, the default-prior model tends to mask uncertainty in the ancestral-area estimates).

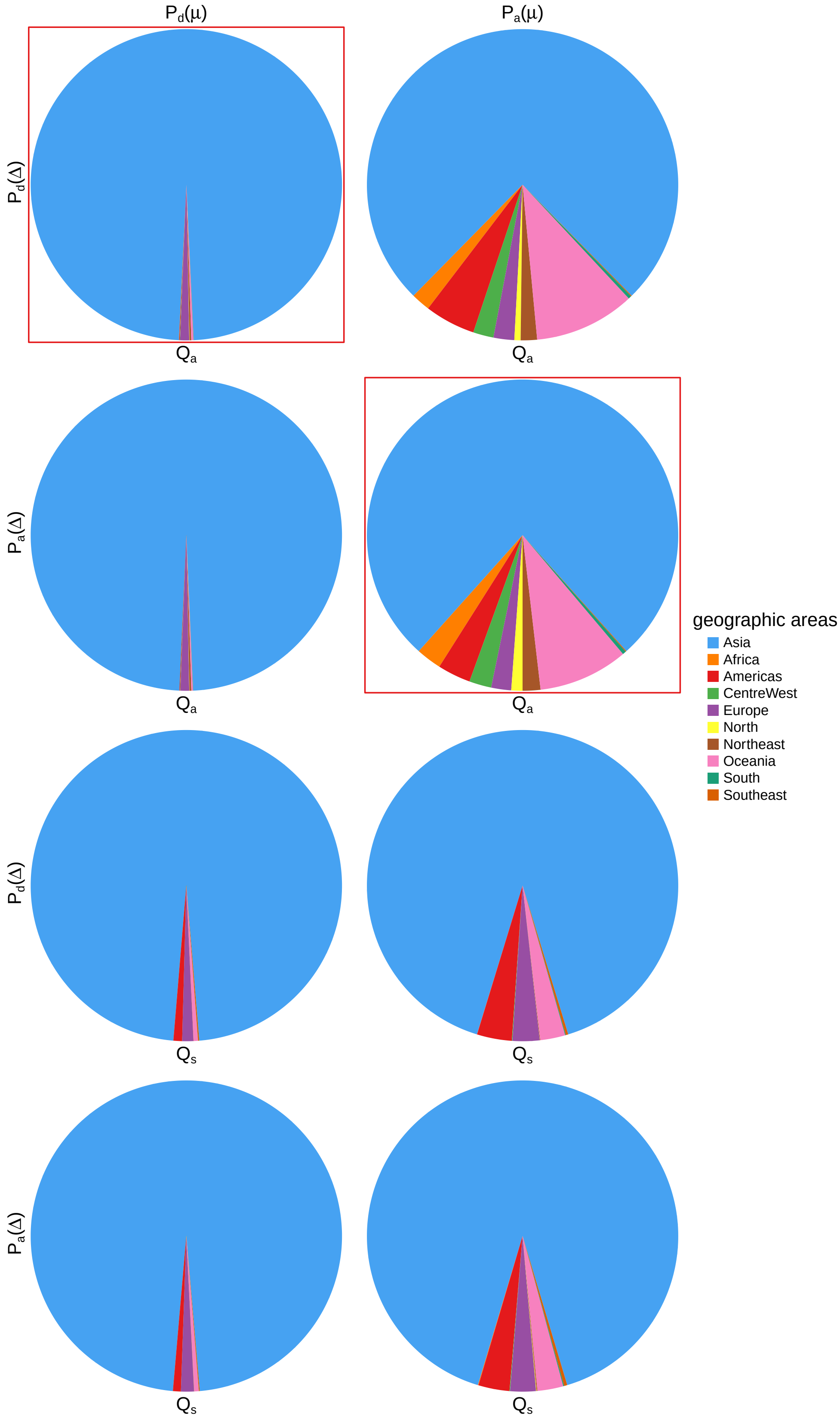


**Figure S168: The impact of prior choice on ancestral-area estimates at the root node for SARS-CoV-2 Brazil SchemeB dataset.** The posterior probability of each ancestral area at the root node under each prior model for SARS-CoV-2 Brazil SchemeB dataset. The pie chart enclosed in red corresponds to the comparison between the preferred default- and the preferred alternative-prior models (*c.f.*, Figure S167, left panel). Model notation follows the description in Table S18.

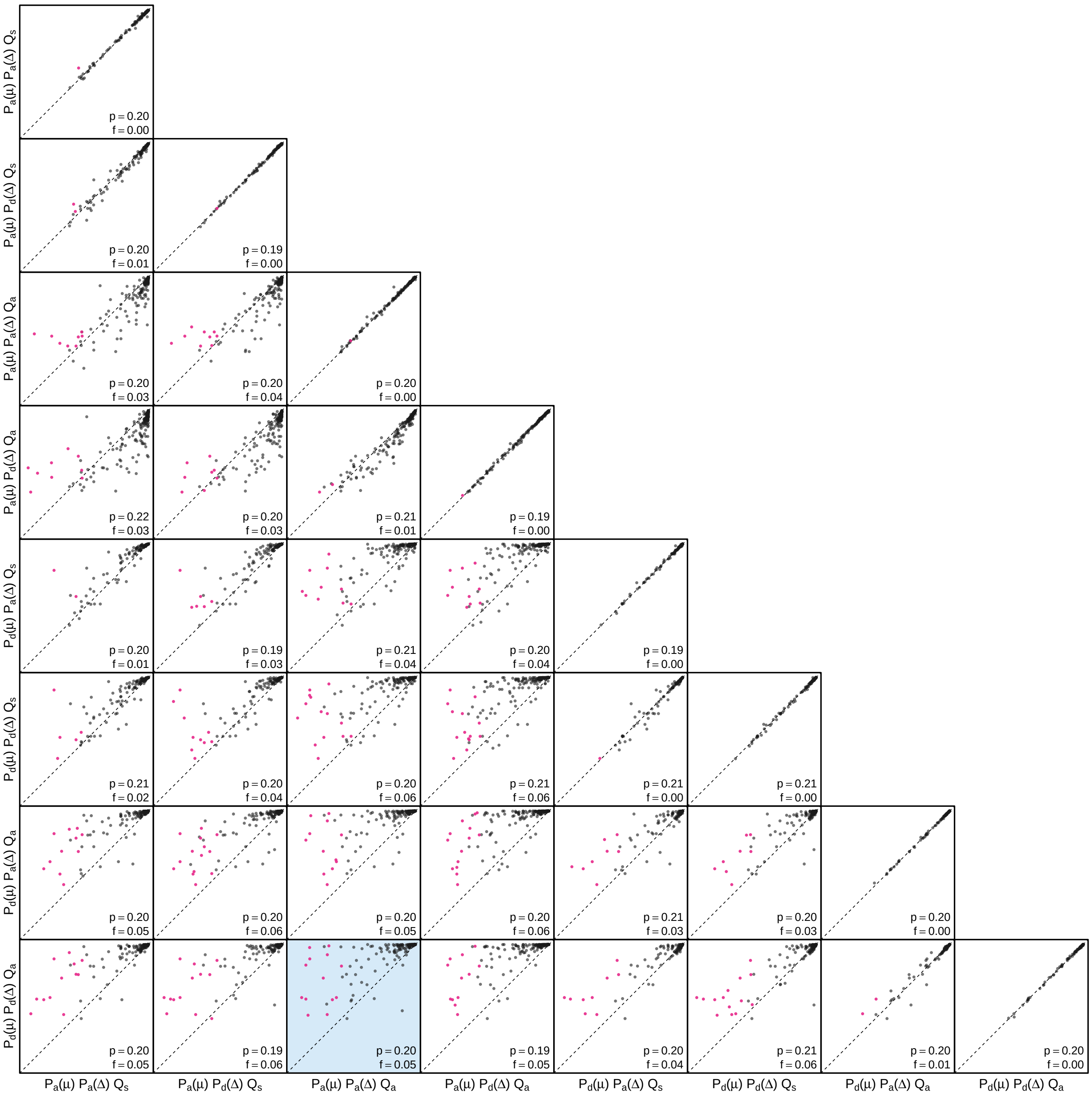


**Figure S169: The impact of prior choice on the MAP ancestral-area estimates at internal nodes for SARS-CoV-2 Brazil SchemeB dataset.** Pairwise scatter plots compare the posterior probability of the MAP ancestral area inferred under each combination of prior models. Diagonal cells are comparisons between two replicates under the same prior model (assessing the convergence of MCMC simulations); off-diagonal cells are comparisons between different prior models, demonstrating the impact of the prior model on both the posterior probability and the identity of the MAP ancestral-area estimates at internal nodes. Pink dots represent the internal nodes where the MAP ancestral area inferred under the default-prior model differs from that inferred under the alternative-prior models. The statistic $p$ denotes the fraction of internal nodes that are shared under the default- and alternative-prior models; $f$ is the fraction of shared nodes where the MAP ancestral area differs under the default- and alternative-prior models. The shaded cell corresponds to the comparison between the preferred default- and the preferred alternative-prior models (*c.f.*, Figure S167, left panel). Model notation follows the description in Table S18.

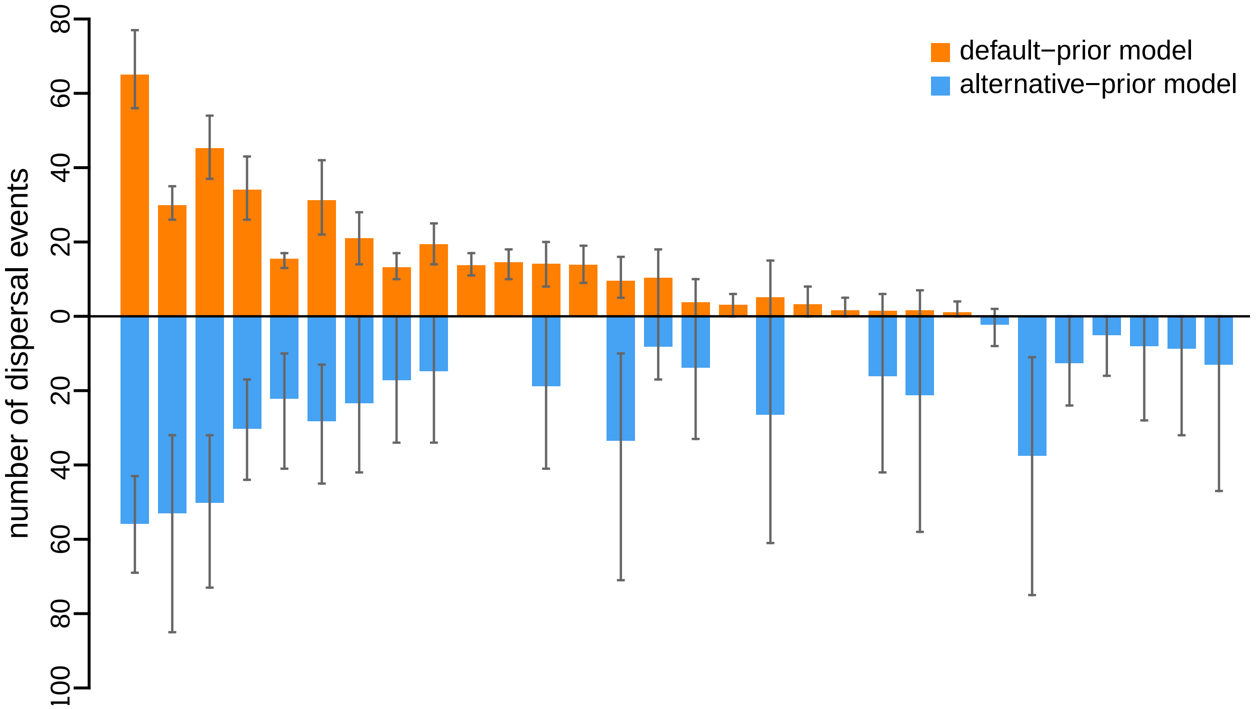


**Figure S170: The impact of prior choice on the inferred number of dispersal events between each pair of areas for SARS-CoV-2 Brazil SchemeB dataset.** The reflected bar plot depicts the number of dispersal events inferred under the default (orange) and alternative (blue) prior models for SARS-CoV-2 Brazil SchemeB dataset. Each bar indicates the posterior-mean number of dispersal events between a pair of areas; whiskers indicate the 95% credible interval. Note that only the number of dispersal events over the "significant" dispersal routes (*i.e.*, $2 \ln \mathrm{BF} > 2$) are figured.

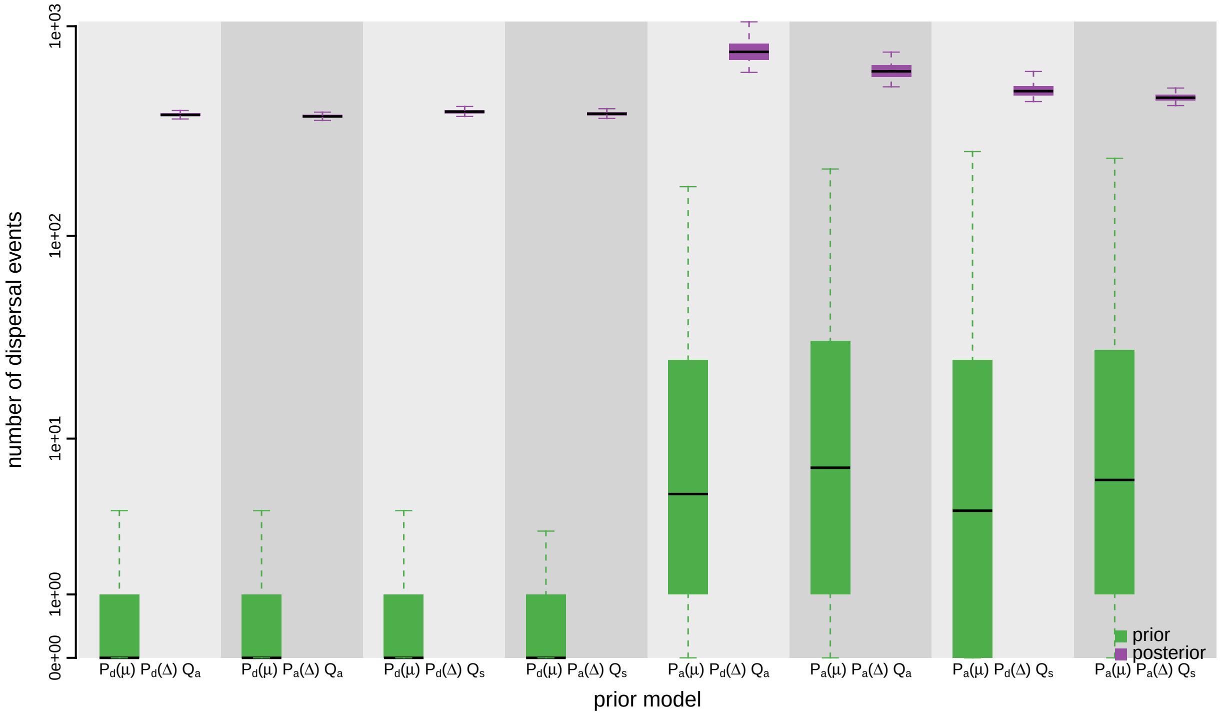


**Figure S171: The impact of prior choice on the inferred total number of dispersal events between all areas for SARS-CoV-2 Brazil SchemeB dataset.** Each pair of boxplots depicts the specified prior (green) and corresponding posterior estimate (purple) of the number of dispersal events between all areas under each prior model: the center of each box indicates the posterior-median number of dispersal events; the box and whiskers indicate the corresponding 50% and 95% credible intervals, respectively. Model notation follows the description in Table S18.

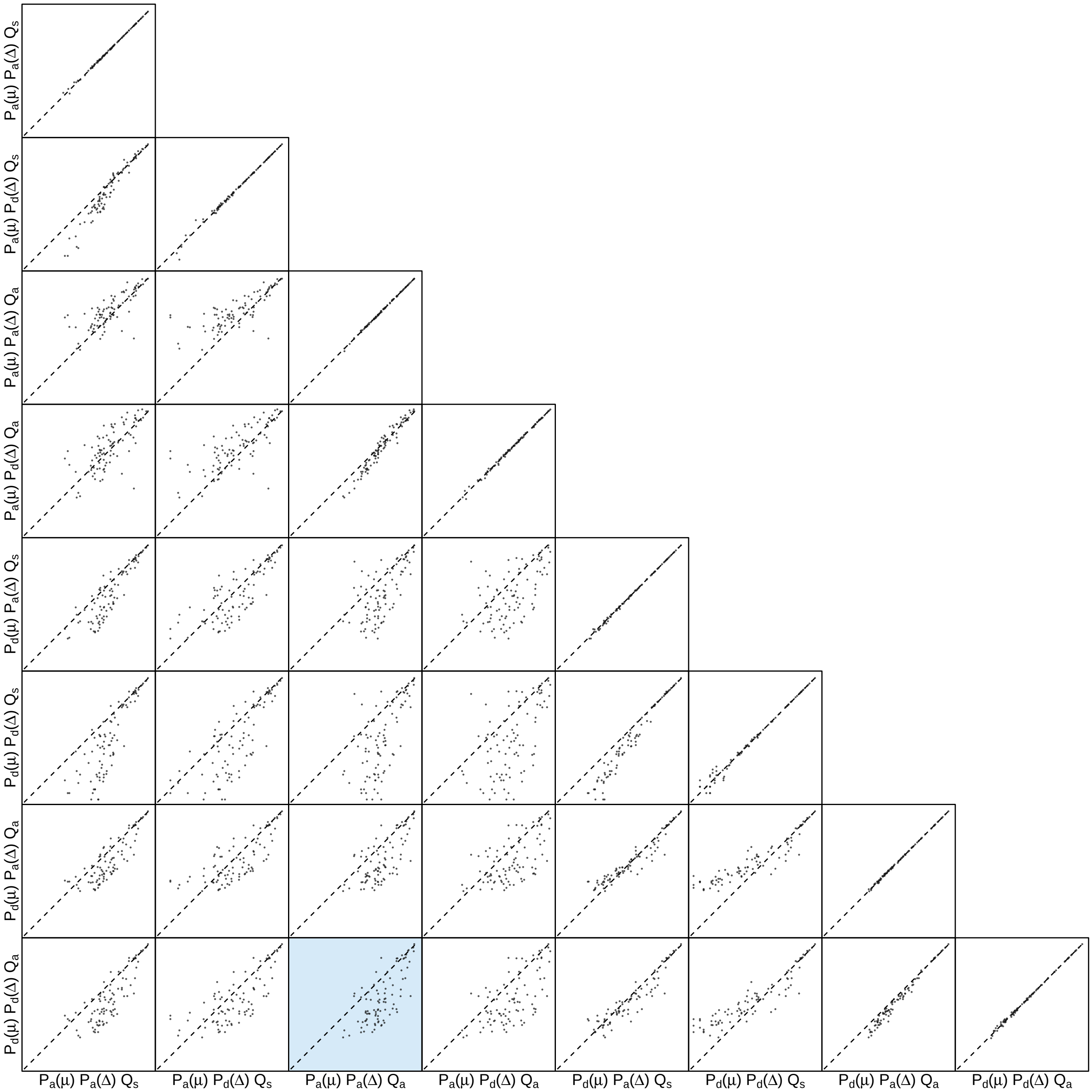


**Figure S172: The impact of prior choice on the inferred number of dispersal events between each pair of areas for SARS-CoV-2 Brazil SchemeB dataset.** Each cell of the plot compares the inferred number of pairwise dispersal events between each pair of prior models. *x*- and *y*-axis of each cell are both on log scale. Diagonal cells are comparisons between half and the other half of the replicates under the same prior model, assessing the convergence of MCMC simulations; off-diagonal cells are comparisons between different prior models, demonstrating the impact of the prior model on the inferred number of pairwise dispersal events. The shaded cell corresponds to the comparison between the preferred default- and the preferred alternative-prior models (*c.f.*, Figure S170). Model notation follows the description in Table S18.

##### S3.3.8.2 SchemeC Dataset

*The Impact of Prior Choice on Biogeographic Model Fit*

**Table S19: Marginal-likelihood estimates of each of the eight prior models for SARS-CoV-2 Brazil SchemeC dataset.** Columns 2–5 list marginal likelihoods inferred from four replicate analyses. Columns 6–7 list the mean and standard deviation of these marginal-likelihood estimates. The last column lists the marginal-likelihood estimates computed by combining the samples from the replicate power-posterior MCMC simulations. Candidate models are listed in rows and include all possible combinations of: (1) instantaneous-rate matrices (symmetric, $Q_s$ or asymmetric, $Q_a$); (2) priors on the average dispersal rate [default, $P_d(\mu)$ or alternative, $P_a(\mu)$], and; (3) priors on the number of dispersal routes [default, $P_d(\Delta)$ or alternative, $P_a(\Delta)$]. The preferred default- and alternative-prior models are indicated in bold text.

| Model | replicate1 | replicate2 | replicate3 | replicate4 | mean | sd | combined |
|---|---|---|---|---|---|---|---|
| $\boldsymbol{P_d(\mu)Q_aP_d(\Delta)}$ | **-1372.20** | **-1372.01** | **-1372.04** | **-1372.08** | **-1372.08** | **0.09** | **-1372.07** |
| $P_d(\mu)Q_aP_a(\Delta)$ | -1360.90 | -1360.65 | -1360.92 | -1360.99 | -1360.87 | 0.15 | -1360.88 |
| $P_d(\mu)Q_sP_d(\Delta)$ | -1382.69 | -1382.94 | -1382.91 | -1382.78 | -1382.83 | 0.12 | -1382.82 |
| $P_d(\mu)Q_sP_a(\Delta)$ | -1362.59 | -1362.43 | -1362.42 | -1362.15 | -1362.40 | 0.18 | -1362.39 |
| $P_a(\mu)Q_aP_d(\Delta)$ | -1224.12 | -1224.26 | -1224.87 | -1225.90 | -1224.79 | 0.81 | -1224.60 |
| $\boldsymbol{P_a(\mu)Q_aP_a(\Delta)}$ | **-1225.53** | **-1225.72** | **-1225.46** | **-1225.14** | **-1225.46** | **0.24** | **-1225.45** |
| $P_a(\mu)Q_sP_d(\Delta)$ | -1296.75 | -1297.79 | -1297.17 | -1297.53 | -1297.31 | 0.45 | -1297.26 |
| $P_a(\mu)Q_sP_a(\Delta)$ | -1249.73 | -1249.95 | -1249.99 | -1249.78 | -1249.86 | 0.13 | -1249.86 |

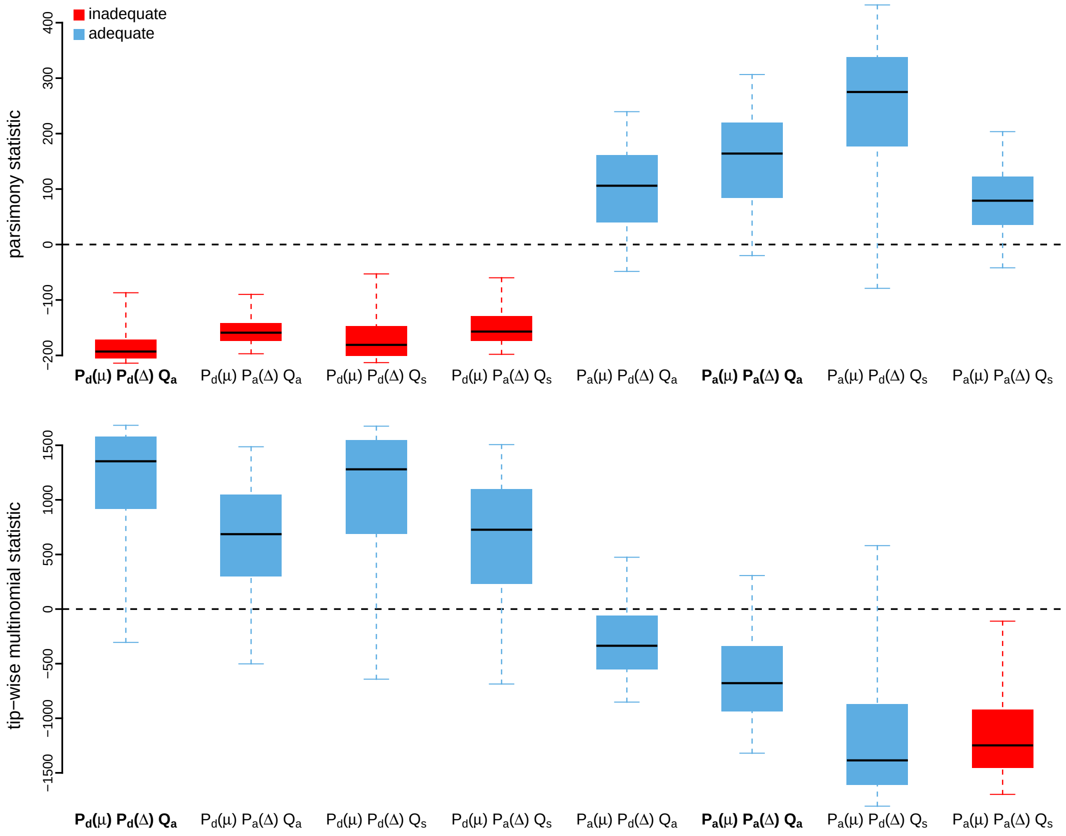


**Figure S173: Posterior-predictive distributions of the parsimony statistic (top panel) and the tip-wise multinomial statistic (bottom panel) under each of the eight prior models for SARS-CoV-2 Brazil SchemeC dataset.** Boxplots depict the posterior-predictive distributions of the statistic under each of the eight candidate prior models: the center of each box is the median predictive value of the summary statistic; the box and whiskers indicate the corresponding 50% and 95% posterior-predictive intervals, respectively. The horizontal dashed line indicates when the simulated and observed datasets produce identical value for the summary statistic. A model is judged to be inadequate (*i.e.*, incapable of generating geographic datasets that are similar to the observed data) if its 95% posterior-predictive interval does not overlap with the dashed line. Here the preferred default prior model is inadequate, whereas the preferred alternative prior model is adequate. Notation for the candidate models is described in Table S19; the preferred default- and alternative-prior models are indicated in bold text.

*The Impact of Prior Choice on Average Dispersal Rate*

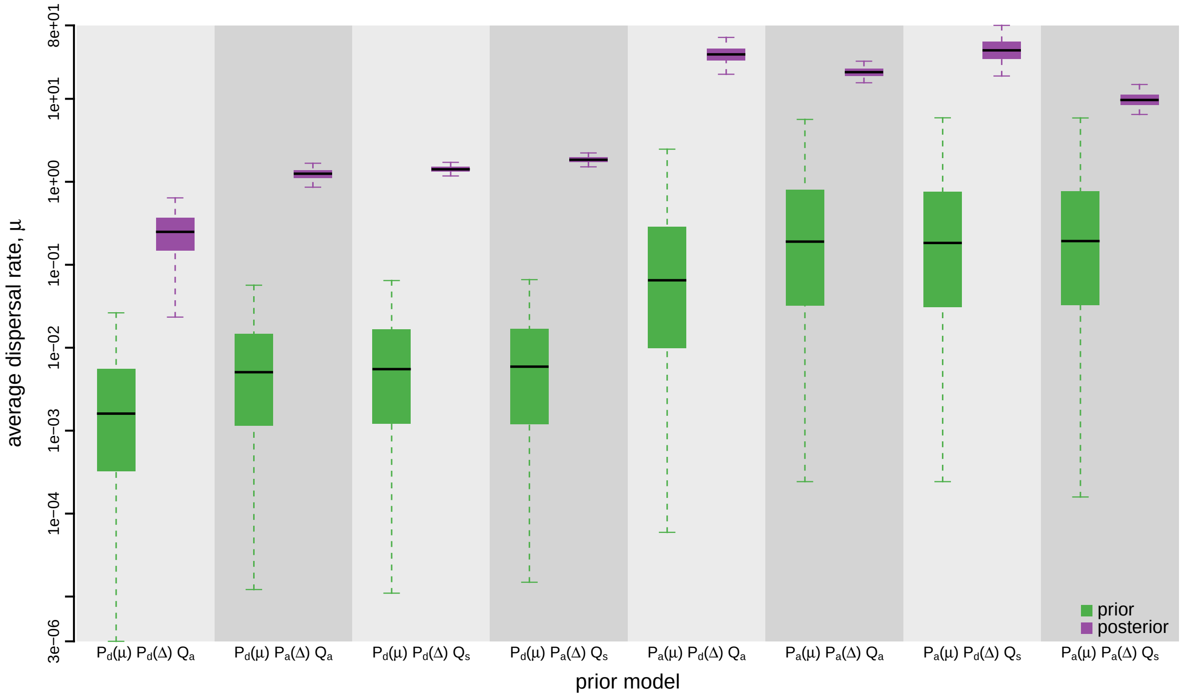


**Figure S174: The impact of prior choice on the average dispersal rate for SARS-CoV-2 Brazil SchemeC dataset.** Each pair of boxplots depicts the specified prior (green) and corresponding posterior estimate (purple) of the average dispersal rate, $\mu$, under each prior model: the center of each box indicates the median rate; the box and whiskers indicate the corresponding 50% and 95% credible intervals, respectively. Notation for the prior models is as described in Table S19.

*The Impact of Prior Choice on Pairwise Dispersal Rates*

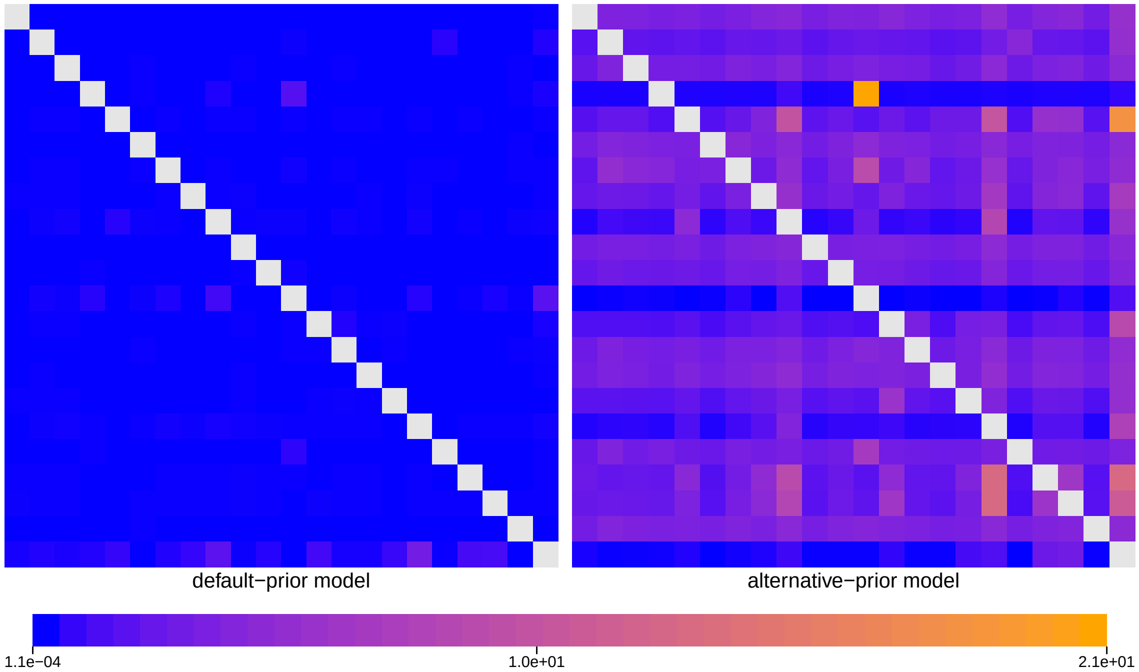


**Figure S175: The impact of prior choice on pairwise dispersal rates for SARS-CoV-2 Brazil SchemeC dataset.** Heatmaps summarize posterior-mean estimates of the instantaneous rate of dispersal between each pair of geographic areas, $q_{ij}$, under the default (left) and alternative (right) prior models.

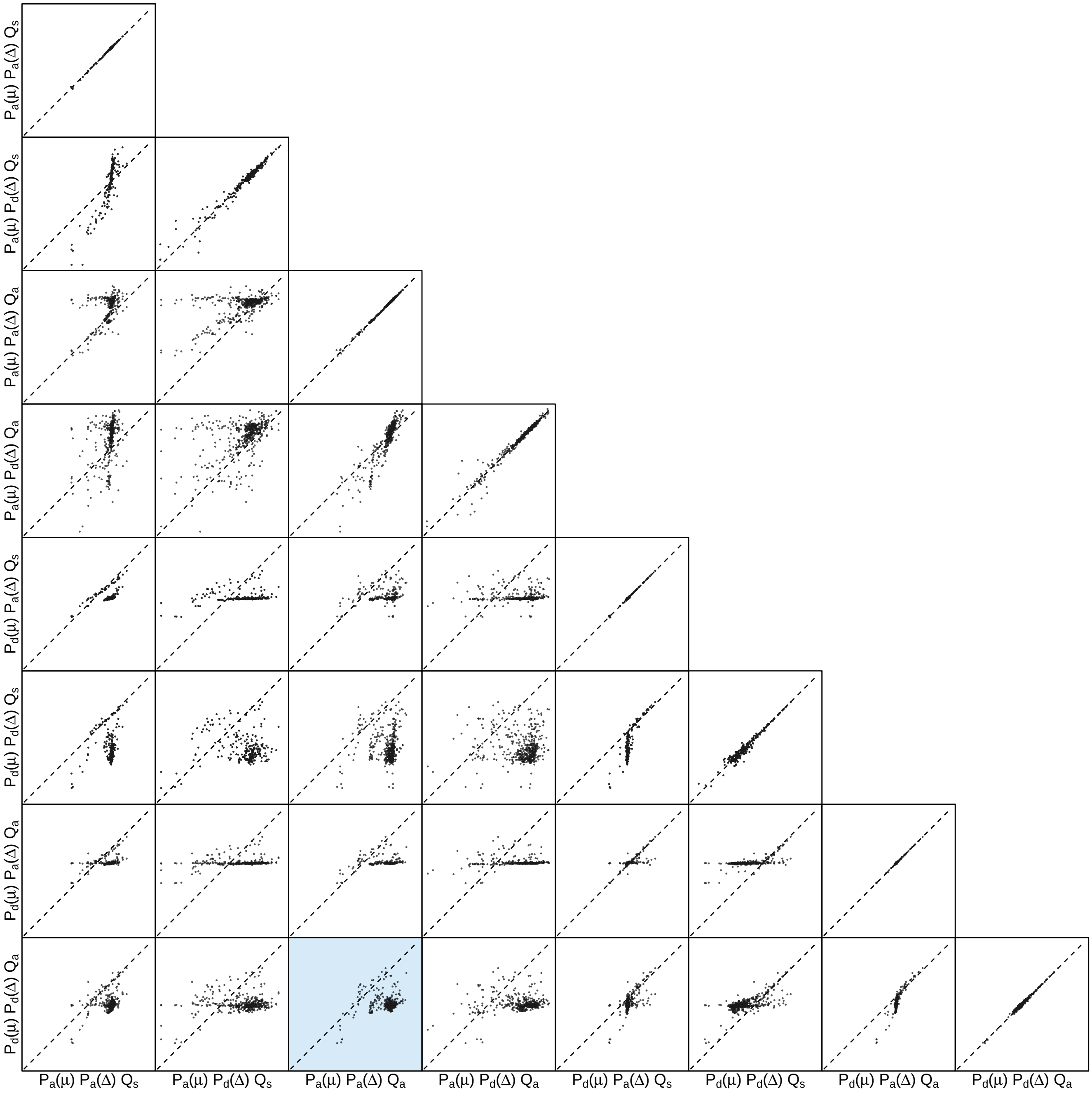


**Figure S176: The impact of prior choice on pairwise dispersal rates for SARS-CoV-2 Brazil SchemeC dataset.** Each cell of the plot compares posterior-mean estimate of the rate of dispersal between each pair of geographic areas, $q_{ij}$, between each pair of prior models. $x$- and $y$-axis of each cell are both on log scale. Diagonal cells are comparisons between half and the other half of the replicates under the same prior model, assessing the convergence of MCMC simulations; off-diagonal cells are comparisons between different prior models, demonstrating the impact of the prior model on the estimates of pairwise dispersal rates. The shaded cell corresponds to the comparison between the preferred default- and the preferred alternative-prior models (*c.f.*, Figure S175). Notation for the prior models is as described in Table S19.

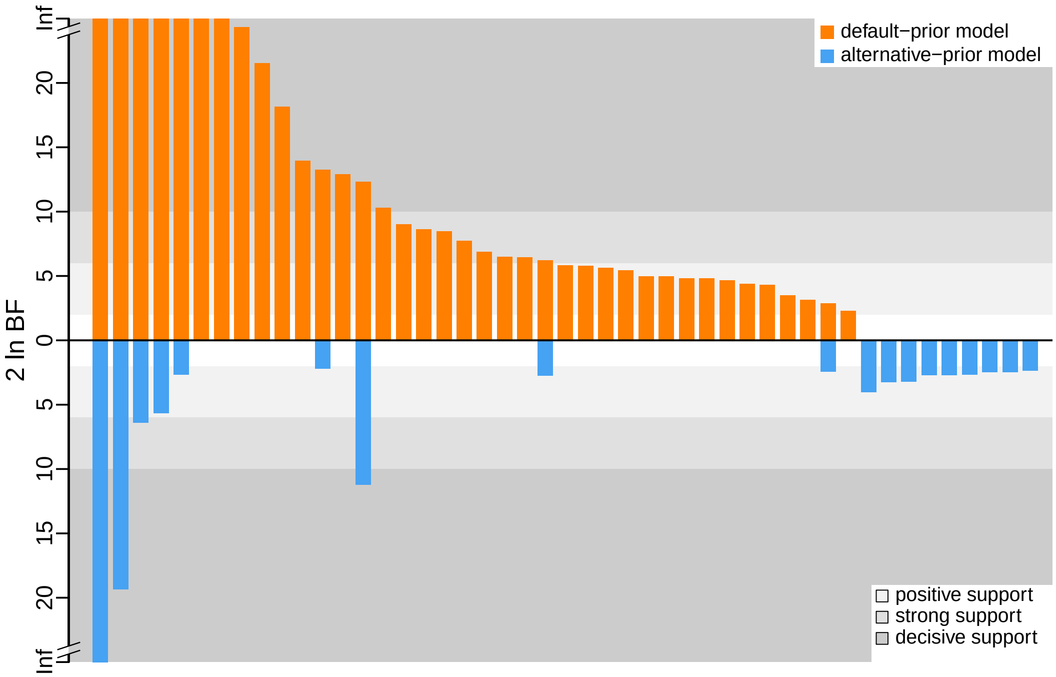


**Figure S177: The impact of prior choice on the inferred support for dispersal routes for SARS-CoV-2 Brazil SchemeC dataset.** We compare the evidential support for each dispersal route for SARS-CoV-2 Brazil SchemeC dataset under the default (orange) and alternative (blue) prior models. Each bar indicates the $2 \ln \mathrm{BF}$ for the corresponding dispersal route between two areas; only supported dispersal routes (*i.e.*, $2 \ln \mathrm{BF} > 2$) are plotted.

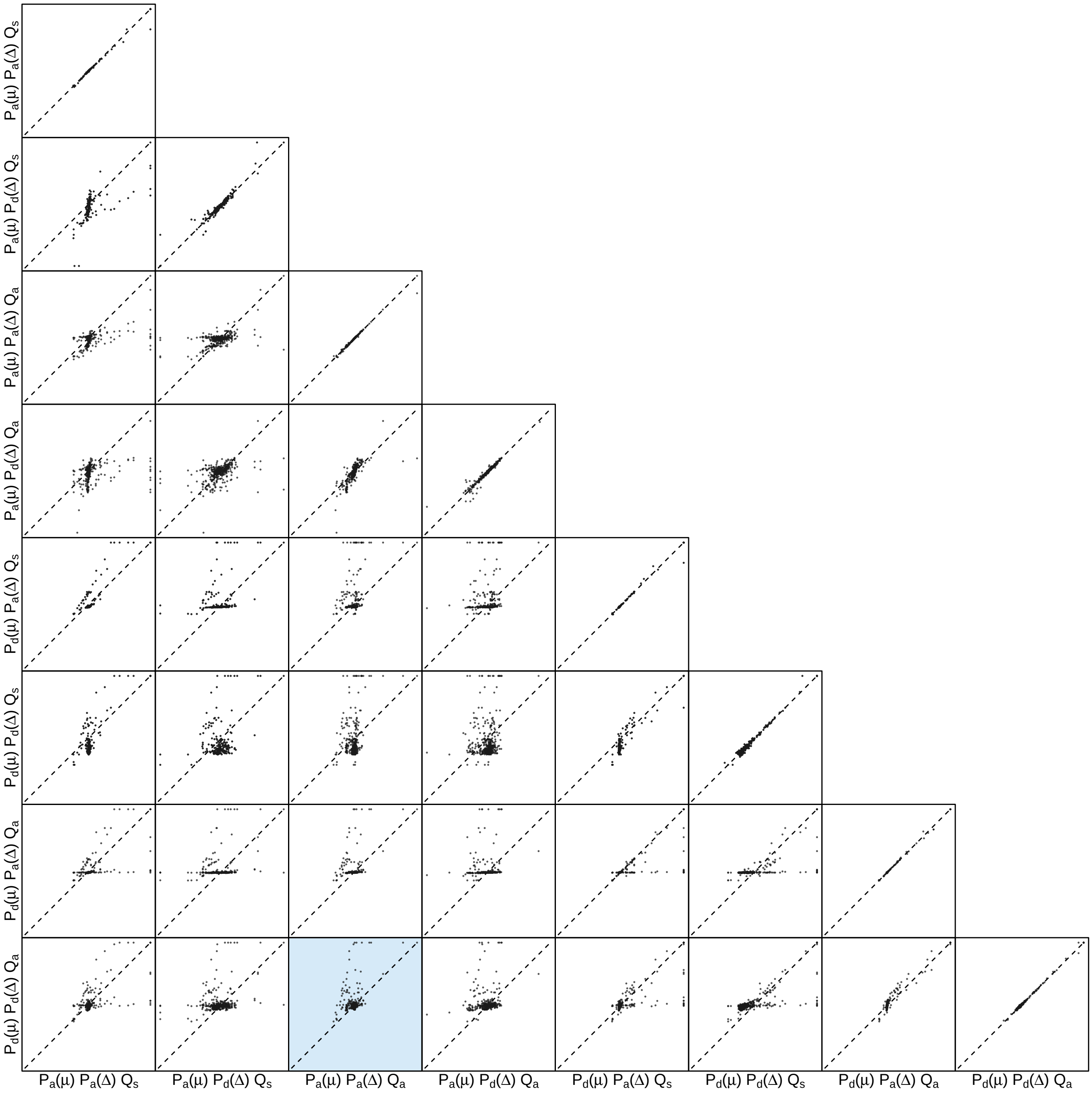


**Figure S178: The impact of prior choice on the inferred support for dispersal routes for SARS-CoV-2 Brazil SchemeC dataset.** Each cell of the plot compares the inferred support ($2 \ln \mathrm{BF}$) for pairwise dispersal routes between each pair of prior models. Diagonal cells are comparisons between half and the other half of the replicates under the same prior model, assessing the convergence of MCMC simulations; off-diagonal cells are comparisons between different prior models, demonstrating the impact of the prior model on the inferred support for pairwise dispersal routes. The shaded cell corresponds to the comparison between the preferred default- and the preferred alternative-prior models (*c.f.*, Figure S177). Notation for the prior models is as described in Table S19.

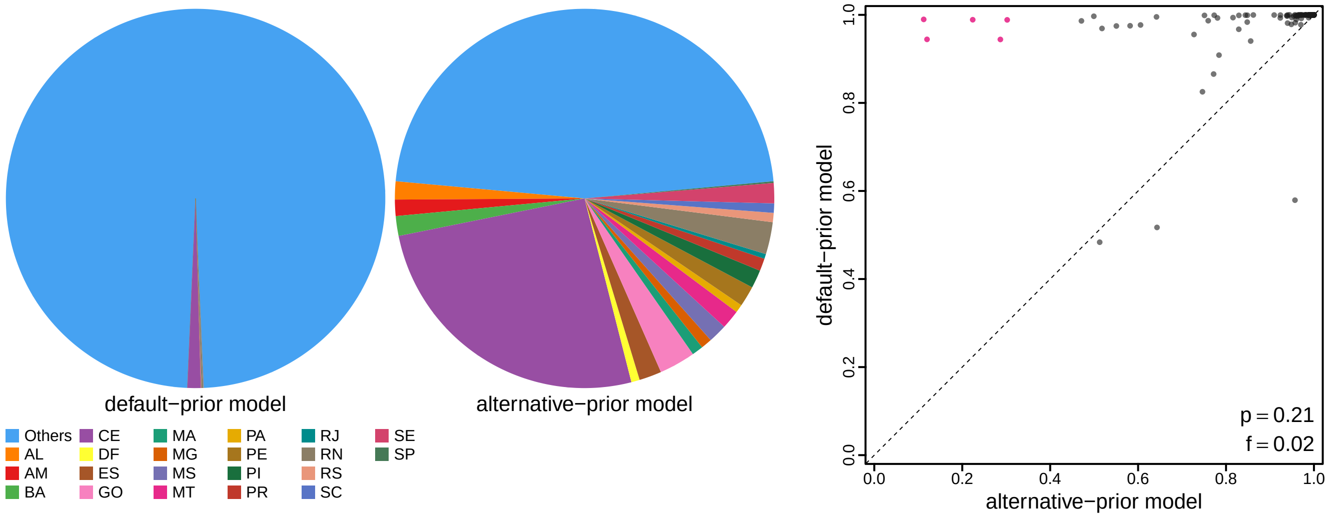


**Figure S179: The impact of prior choice on ancestral-area estimates for SARS-CoV-2 Brazil SchemeC dataset.** The left panel compares the posterior probability of each ancestral area at the root node under the default- and alternative-prior models for SARS-CoV-2 Brazil SchemeC dataset. The right panel plots the posterior probability of the most probable ancestral area under the default-prior model for each node in the MCC tree (y-axis) against the corresponding posterior probability of that area under the alternative-prior model (x-axis). Pink dots represent the internal nodes where the MAP ancestral area inferred under the default-prior model differs from that inferred under the alternative-prior models. The statistic $p$ denotes the fraction of internal nodes that are shared under the default- and alternative-prior models; $f$, is the fraction of shared nodes where the MAP ancestral area differs under the default- and alternative-prior models. Note that the posterior probabilities of the MAP ancestral area under the default-prior model are generally higher than those under the alternative-prior model (*i.e.*, the default-prior model tends to mask uncertainty in the ancestral-area estimates).

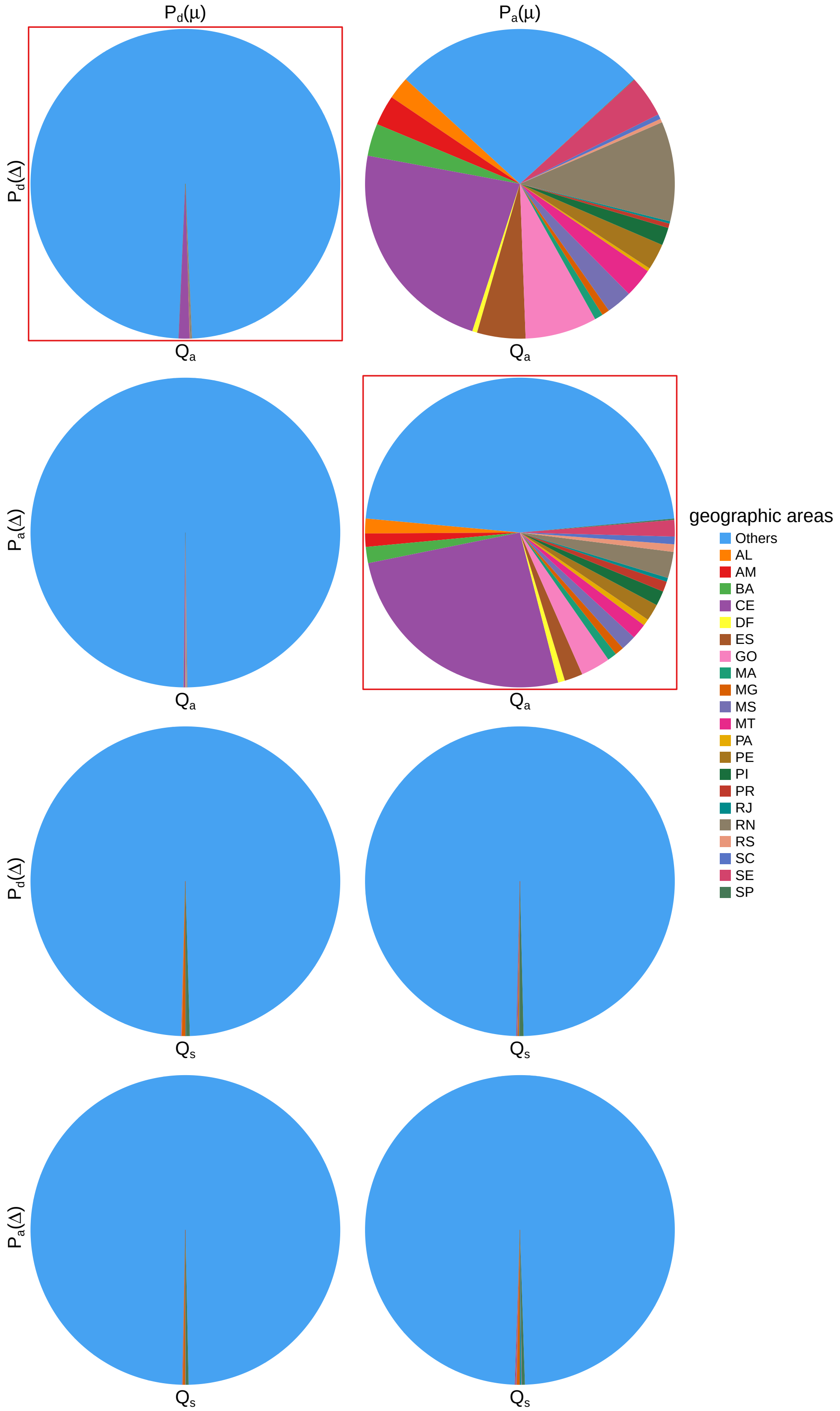


**Figure S180: The impact of prior choice on ancestral-area estimates at the root node for SARS-CoV-2 Brazil SchemeC dataset.** The posterior probability of each ancestral area at the root node under each prior model for SARS-CoV-2 Brazil SchemeC dataset. The pie chart enclosed in red corresponds to the comparison between the preferred default- and the preferred alternative-prior models (*c.f.*, Figure S179, left panel). Model notation follows the description in Table S19.

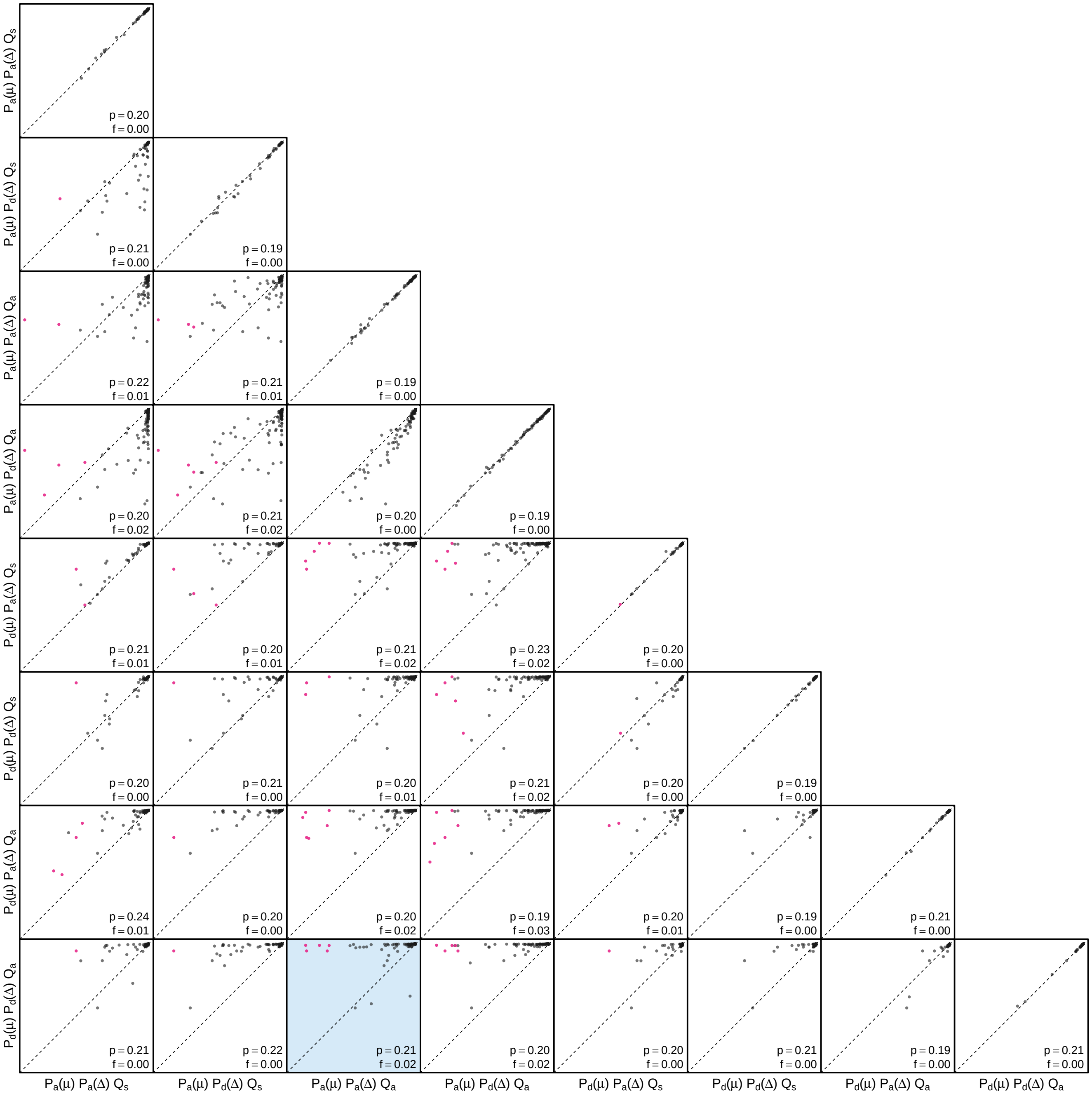


**Figure S181: The impact of prior choice on the MAP ancestral-area estimates at internal nodes for SARS-CoV-2 Brazil SchemeC dataset.** Pairwise scatter plots compare the posterior probability of the MAP ancestral area inferred under each combination of prior models. Diagonal cells are comparisons between two replicates under the same prior model (assessing the convergence of MCMC simulations); off-diagonal cells are comparisons between different prior models, demonstrating the impact of the prior model on both the posterior probability and the identity of the MAP ancestral-area estimates at internal nodes. Pink dots represent the internal nodes where the MAP ancestral area inferred under the default-prior model differs from that inferred under the alternative-prior models. The statistic $p$ denotes the fraction of internal nodes that are shared under the default- and alternative-prior models; $f$ is the fraction of shared nodes where the MAP ancestral area differs under the default- and alternative-prior models. The shaded cell corresponds to the comparison between the preferred default- and the preferred alternative-prior models (*c.f.*, Figure S179, left panel). Model notation follows the description in Table S19.

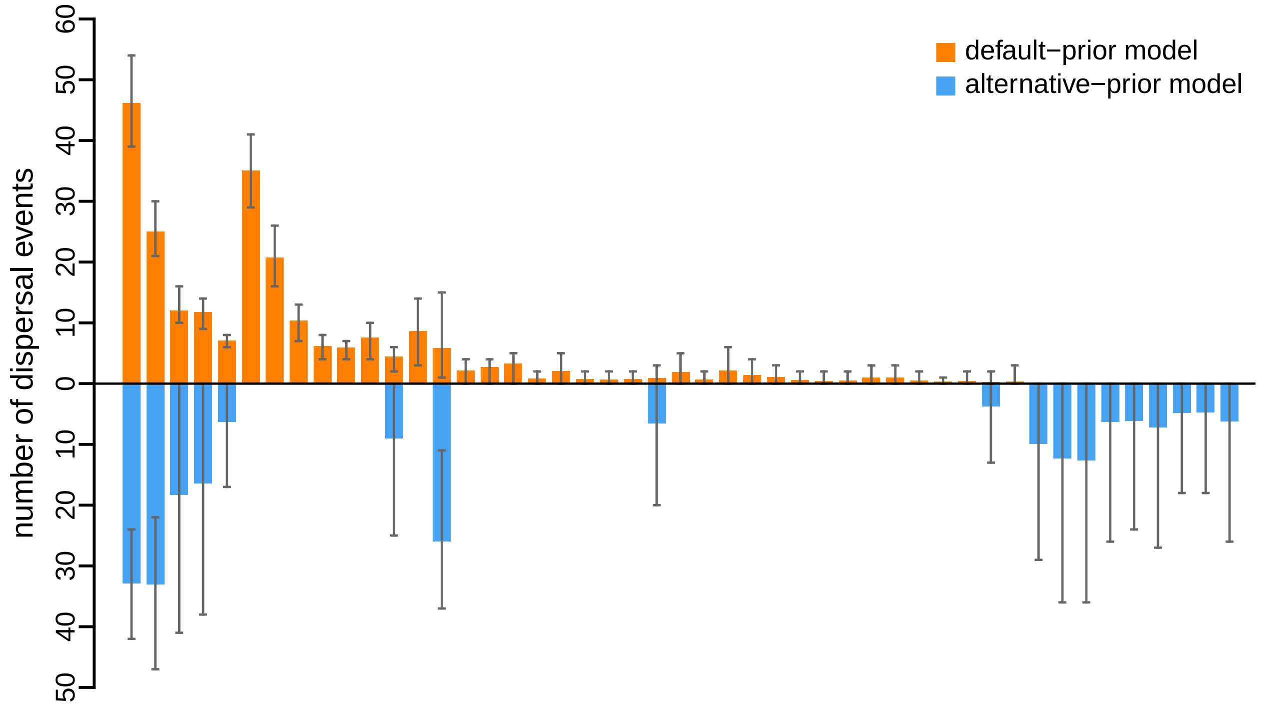


**Figure S182: The impact of prior choice on the inferred number of dispersal events between each pair of areas for SARS-CoV-2 Brazil SchemeC dataset.** The reflected bar plot depicts the number of dispersal events inferred under the default (orange) and alternative (blue) prior models for SARS-CoV-2 Brazil SchemeC dataset. Each bar indicates the posterior-mean number of dispersal events between a pair of areas; whiskers indicate the 95% credible interval. Note that only the number of dispersal events over the "significant" dispersal routes (*i.e.*, $2 \ln \mathrm{BF} > 2$) are figured.

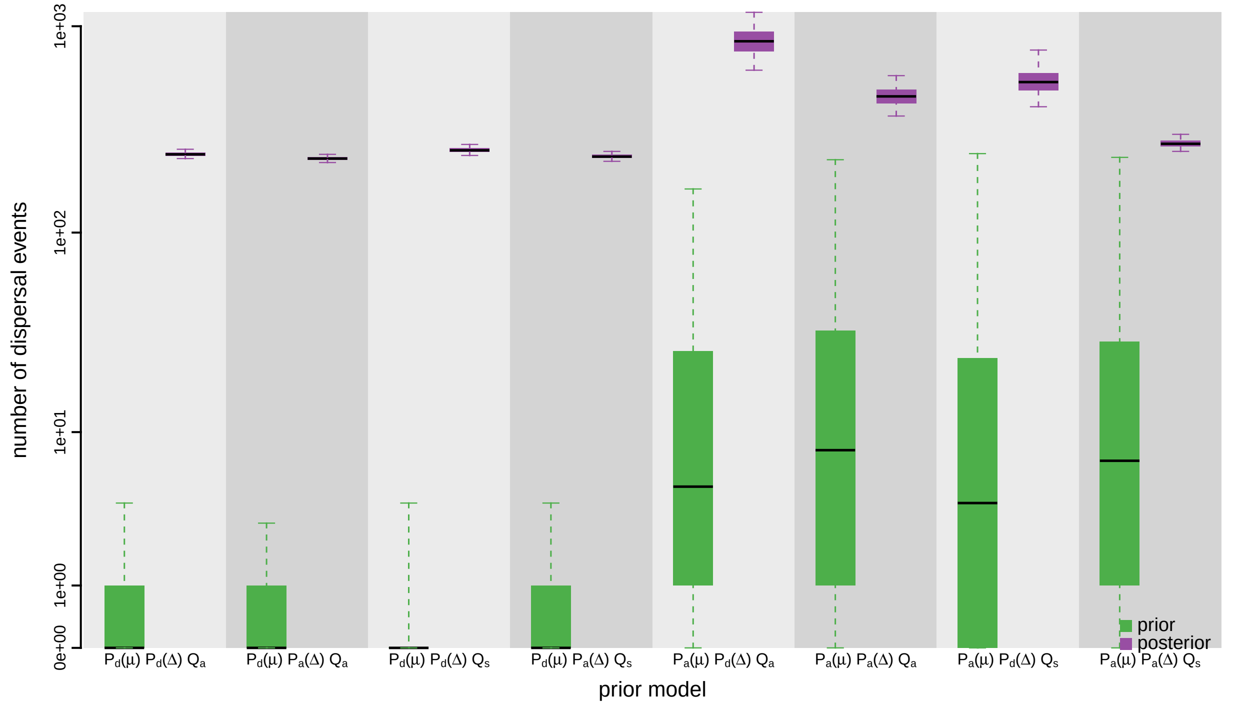


**Figure S183: The impact of prior choice on the inferred total number of dispersal events between all areas for SARS-CoV-2 Brazil SchemeC dataset.** Each pair of boxplots depicts the specified prior (green) and corresponding posterior estimate (purple) of the number of dispersal events between all areas under each prior model: the center of each box indicates the posterior-median number of dispersal events; the box and whiskers indicate the corresponding 50% and 95% credible intervals, respectively. Model notation follows the description in Table S19.

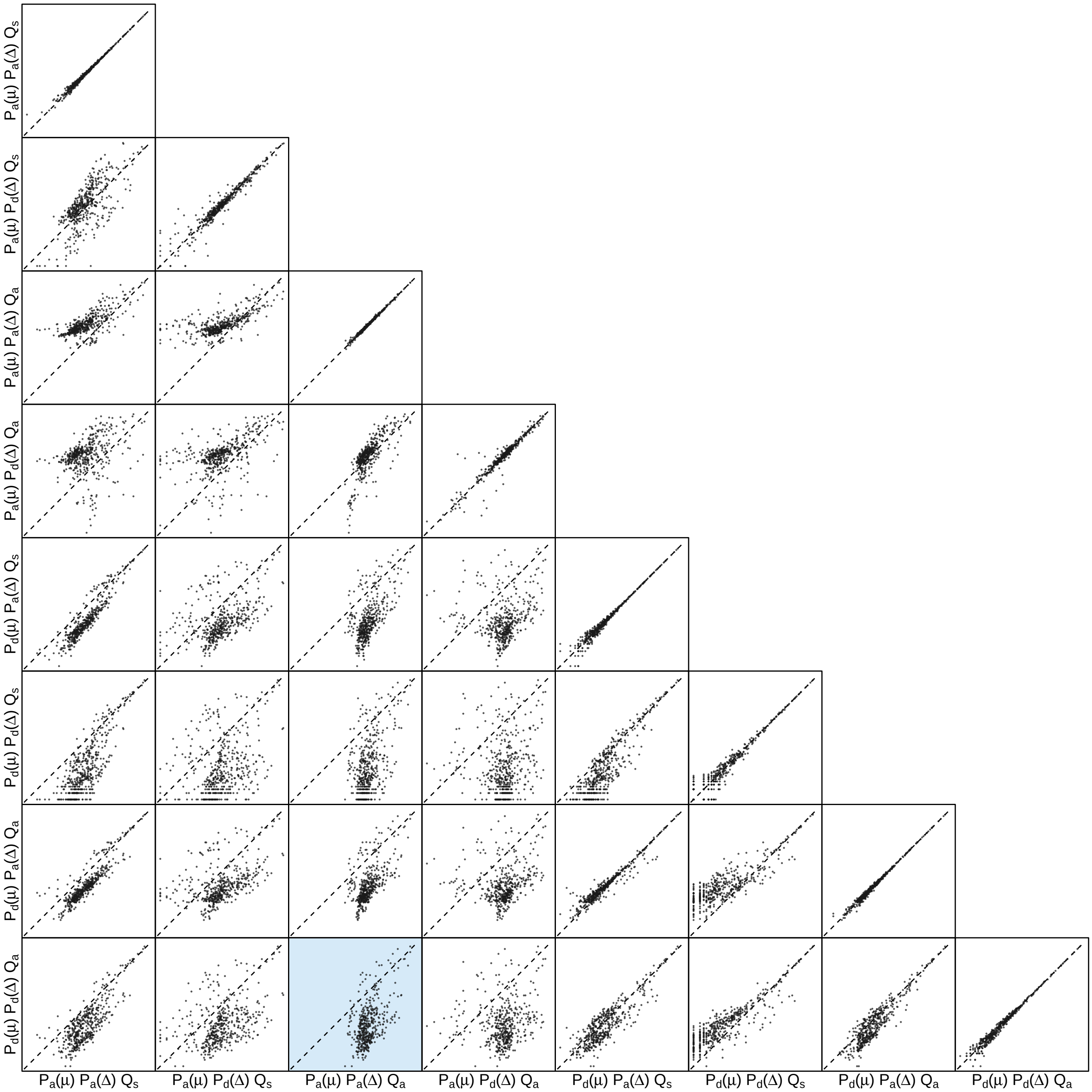


**Figure S184: The impact of prior choice on the inferred number of dispersal events between each pair of areas for SARS-CoV-2 Brazil SchemeC dataset.** Each cell of the plot compares the inferred number of pairwise dispersal events between each pair of prior models. *x*- and *y*-axis of each cell are both on log scale. Diagonal cells are comparisons between half and the other half of the replicates under the same prior model, assessing the convergence of MCMC simulations; off-diagonal cells are comparisons between different prior models, demonstrating the impact of the prior model on the inferred number of pairwise dispersal events. The shaded cell corresponds to the comparison between the preferred default- and the preferred alternative-prior models (*c.f.*, Figure S182). Model notation follows the description in Table S19.

#### S4 References


Aho, A. V., Hopcroft, J. E., and Ullman, J. D. (1983). *Data structures and algorithms*. Addison-Wesley.

Alpert, T., Brito, A. F., Lasek-Nesselquist, E., Rothman, J., Valesano, A. L., MacKay, M. J., Petrone, M. E., Breban, M. I., Watkins, A. E., Vogels, C. B., et al. (2021). Early introductions and transmission of SARS-CoV-2 variant B. 1.1. 7 in the United States. *Cell*, 184(10):2595–2604.

Archer, K., Gessel, I. M., Graves, C., and Liang, X. (2020). Counting acyclic and strong digraphs by descents. *Discrete Mathematics*, 343(11):112041.

Ayres, D. L., Cummings, M. P., Baele, G., Darling, A. E., Lewis, P. O., Swofford, D. L., Huelsenbeck, J. P., Lemey, P., Rambaut, A., and Suchard, M. A. (2019). BEAGLE 3: Improved performance, scaling, and usability for a high-performance computing library for statistical phylogenetics. *Systematic Biology*, 68(6):1052–1061.

Ayres, D. L., Darling, A., Zwickl, D. J., Beerli, P., Holder, M. T., Lewis, P. O., Huelsenbeck, J. P., Ronquist, F., Swofford, D. L., Cummings, M. P., et al. (2012). BEAGLE: an application programming interface and high-performance computing library for statistical phylogenetics. *Systematic Biology*, 61(1):170–173.

Baele, G., Lemey, P., Bedford, T., Rambaut, A., Suchard, M. A., and Alekseyenko, A. V. (2012). Improving the accuracy of demographic and molecular clock model comparison while accommodating phylogenetic uncertainty. *Molecular Biology and Evolution*, 29(9):2157–2167.

Bedford, T., Riley, S., Barr, I. G., Broor, S., Chadha, M., Cox, N. J., Daniels, R. S., Gunasekaran, C. P., Hurt, A. C., Kelso, A., et al. (2015). Global circulation patterns of seasonal influenza viruses vary with antigenic drift. *Nature*, 523(7559):217.

Bielejec, F., Baele, G., Vrancken, B., Suchard, M. A., Rambaut, A., and Lemey, P. (2016). Spread3: interactive visualization of spatiotemporal history and trait evolutionary processes. *Molecular Biology and Evolution*, 33(8):2167–2169.

Bielejec, F., Rambaut, A., Suchard, M. A., and Lemey, P. (2011). Spread: spatial phylogenetic reconstruction of evolutionary dynamics. *Bioinformatics*, 27(20):2910–2912.

Blanquart, S. and Lartillot, N. (2006). A Bayesian compound stochastic process for modeling nonstationary and nonhomogeneous sequence evolution. *Molecular Biology and Evolution*, 23(11):2058–2071.

Bollback, J. P. (2002). Bayesian model adequacy and choice in phylogenetics. *Molecular Biology and Evolution*, 19(7):1171–1180.

Candido, D. S., Claro, I. M., De Jesus, J. G., Souza, W. M., Moreira, F. R., Dellicour, S., Mellan, T. A., Du Plessis, L., Pereira, R. H., Sales, F. C., et al. (2020). Evolution and epidemic spread of SARS-CoV-2 in Brazil. *Science*, 369(6508):1255–1260.

Dash, P. K., Sharma, S., Soni, M., Agarwal, A., Sahni, A. K., and Parida, M. (2015). Complete genome sequencing and evolutionary phylogeography analysis of Indian isolates of Dengue virus type 1. *Virus Research*, 195:124–134.

Drummond, A. J., Ho, S. Y., Phillips, M. J., and Rambaut, A. (2006). Relaxed phylogenetics and dating with confidence. *PLoS Biology*, 4(5):e88.

Drummond, A. J., Suchard, M. A., Xie, D., and Rambaut, A. (2012). Bayesian phylogenetics with BEAUti and the BEAST 1.7. *Molecular Biology and Evolution*, 29(8):1969–1973.

Edgar, R. C. (2004). MUSCLE: multiple sequence alignment with high accuracy and high throughput. *Nucleic Acids Research*, 32(5):1792–1797.

Edwards, C. J., Suchard, M. A., Lemey, P., Welch, J. J., Barnes, I., Fulton, T. L., Barnett, R., O'Connell, T. C., Coxon, P., Monaghan, N., et al. (2011). Ancient hybridization and an Irish origin for the modern polar bear matriline. *Current Biology*, 21(15):1251–1258.

Faria, N. R., Rambaut, A., Suchard, M. A., Baele, G., Bedford, T., Ward, M. J., Tatem, A. J., Sousa, J. D., Arinaminpathy, N., Pépin, J., et al. (2014). The early spread and epidemic ignition of HIV-1 in human populations. *Science*, 346(6205):56–61.

Felsenstein, J. (1973). Maximum-likelihood estimation of evolutionary trees from continuous characters. *American Journal of Human Genetics*, 25(5):471.

Felsenstein, J. (1981). Evolutionary trees from DNA sequences: a maximum likelihood approach. *Journal of Molecular Evolution*, 17(6):368–376.

Galtier, N. and Gouy, M. (1998). Inferring pattern and process: maximum-likelihood implementation of a nonhomogeneous model of DNA sequence evolution for phylogenetic analysis. *Molecular Biology and Evolution*, 15(7):871–879.

Gao, J., May, M. R., Rannala, B., and Moore, B. R. (2022). New phylogenetic models incorporating interval-specific dispersal dynamics improve inference of disease spread. *Molecular Biology and Evolution*, 39(8):msac159.

Gelman, A., Carlin, J. B., Stern, H. S., Dunson, D. B., Vehtari, A., and Rubin, D. B. (2013). *Bayesian data analysis*. CRC press.

Gelman, A., Meng, X.-L., and Stern, H. (1996). Posterior predictive assessment of model fitness via realized discrepancies. *Statistica Sinica*, pages 733–760.

Gelman, A. and Rubin, D. B. (1992). Inferences from iterative simulation using multiple sequences. *Statistical Science*, 7:457–511.

Goldman, N. (1993). Statistical tests of models of DNA substitution. *Journal of Molecular Evolution*, 36(2):182–198.

Gu, X., Fu, Y.-X., and Li, W.-H. (1995). Maximum likelihood estimation of the heterogeneity of substitution rate among nucleotide sites. *Molecular Biology and Evolution*, 12(4):546–557.

Harary, F. and Palmer, E. M. (2014). *Graphical enumeration*. Elsevier.

Hasegawa, M., Kishino, H., and Yano, T.-a. (1985). Dating of the human-ape splitting by a molecular clock of mitochondrial dna. *Journal of Molecular Evolution*, 22(2):160–174.

Hasegawa, M., Yano, T.-a., and Kishino, H. (1984). A new molecular clock of mitochondrial DNA and the evolution of Hominoids. *Proceedings of the Japan Academy, series B*, 60(4):95–98.

Hobolth, A. and Stone, E. A. (2009). Simulation from endpoint-conditioned, continuous-time Markov chains on a finite state space, with applications to molecular evolution. *The Annals of Applied Statistics*, 3(3):1204.

Lartillot, N. and Philippe, H. (2006). Computing Bayes factors using theromodynamic integration. *Systematic Biology*, 55:195–207.

Lele, S. R., Dennis, B., and Lutscher, F. (2007). Data cloning: easy maximum likelihood estimation for complex ecological models using Bayesian Markov chain Monte Carlo methods. *Ecology Letters*, 10(7):551–563.

Lemey, P., Rambaut, A., Drummond, A. J., and Suchard, M. A. (2009). Bayesian phylogeography finds its roots. *PLoS Computational Biology*, 5(9):e1000520.

Minin, V. N., Bloomquist, E. W., and Suchard, M. A. (2008). Smooth skyride through a rough skyline: Bayesian coalescent-based inference of population dynamics. *Molecular Biology and Evolution*, 25(7):1459–1471.

Minin, V. N. and Suchard, M. A. (2008a). Counting labeled transitions in continuous-time Markov models of evolution. *Journal of Mathematical Biology*, 56(3):391–412.

Minin, V. N. and Suchard, M. A. (2008b). Fast, accurate and simulation-free stochastic mapping. *Philosophical Transactions of the Royal Society B: Biological Sciences*, 363(1512):3985–3995.

Nielsen, R. (2002). Mapping mutations on phylogenies. *Systematic Biology*, 51(5):729–739.

O'Brien, J. D., Minin, V. N., and Suchard, M. A. (2009). Learning to count: robust estimates for labeled distances between molecular sequences. *Molecular Biology and Evolution*, 26(4):801–814.

OEIS Foundation Inc. (2022). The On-Line Encyclopedia of Integer Sequences. Published electronically at http://oeis.org.

Plummer, M., Best, N., Cowles, K., and Vines, K. (2006). CODA: convergence diagnosis and output analysis for mcmc. *R news*, 6(1):7–11.

Ponciano, J. M., Burleigh, J. G., Braun, E. L., and Taper, M. L. (2012). Assessing parameter identifiability in phylogenetic models using data cloning. *Systematic Biology*, 61(6):955–972.

Ponciano, J. M., Taper, M. L., Dennis, B., and Lele, S. R. (2009). Hierarchical models in ecology: confidence intervals, hypothesis testing, and model selection using data cloning. *Ecology*, 90(2):356–362.

R Core Team (2020). *R: A Language and Environment for Statistical Computing*. R Foundation for Statistical Computing, Vienna, Austria.

Rambaut, A., Drummond, A. J., Xie, D., Baele, G., and Suchard, M. A. (2018). Posterior summarization in Bayesian phylogenetics using Tracer 1.7. *Systematic Biology*, 67(5):901.

Rannala, B. and Yang, Z. (2007). Inferring speciation times under an episodic molecular clock. *Systematic Biology*, 56(3):453–466.

Revell, L. J. (2012). phytools: an R package for phylogenetic comparative biology (and other things). *Methods in Ecology and Evolution*, 3(2):217–223.

Robert, C. P. (1993). Prior feedback: A Bayesian approach to maximum likelihood estimation. *Computational Statistics*, 8:279–294.

Rodrigue, N., Philippe, H., and Lartillot, N. (2007). Uniformization for sampling realizations of markov processes: applications to bayesian implementations of codon substitution models. *Bioinformatics*, 24(1):56–62.

Schliep, K. P. (2010). phangorn: phylogenetic analysis in R. *Bioinformatics*, 27(4):592–593.

Schneider, T. D., Stormo, G. D., Gold, L., and Ehrenfeucht, A. (1986). Information content of binding sites on nucleotide sequences. *Journal of molecular biology*, 188(3):415–431.

Shannon, C. E. (1948). A mathematical theory of communication. *The Bell system technical journal*, 27(3):379–423.

Shu, Y. and McCauley, J. (2017). GISAID: Global initiative on sharing all influenza data–from vision to reality. *Eurosurveillance*, 22(13):30494.

Suchard, M. A., Lemey, P., Baele, G., Ayres, D. L., Drummond, A. J., and Rambaut, A. (2018). Bayesian phylogenetic and phylodynamic data integration using BEAST 1.10. *Virus Evolution*, 4(1):vey016.

Tamura, K. and Nei, M. (1993). Estimation of the number of nucleotide substitutions in the control region of mitochondrial DNA in humans and chimpanzees. *Molecular Biology and Evolution*, 10(3):512–526.

Tavaré, S. (1986). Some probabilistic and statistical problems in the analysis of DNA sequences. *Lectures on Mathematics in the Life Sciences*, 17(2):57–86.

The PARI Group (2022). *PARI/GP version* `2.15.1`. Univ. Bordeaux. available from http://pari.math.u-bordeaux.fr/.

Wilfert, L., Long, G., Leggett, H., Schmid-Hempel, P., Butlin, R., Martin, S., and Boots, M. (2016). Deformed wing virus is a recent global epidemic in honeybees driven by *Varroa* mites. *Science*, 351(6273):594–597.

Xie, W., Lewis, P. O., Fan, Y., Kuo, L., and Chen, M.-H. (2011). Improving marginal likelihood estimation for Bayesian phylogenetic model selection. *Systematic Biology*, 60:150–160.

Yang, Z. (1994). Maximum likelihood phylogenetic estimation from DNA sequences with variable rates over sites: approximate methods. *Journal of Molecular Evolution*, 39(3):306–314.

Yang, Z. (2014). *Molecular Evolution: a Statistical Approach*. Oxford University Press.

Yang, Z. and Roberts, D. (1995). On the use of nucleic acid sequences to infer early branchings in the tree of life. *Molecular Biology and Evolution*, 12(3):451–458.

Yao, H.-W., Yang, Y., Liu, K., Li, X.-L., Zuo, S.-Q., Sun, R.-X., Fang, L.-Q., and Cao, W.-C. (2015). The spatiotemporal expansion of human Rabies and its probable explanation in mainland China, 2004-2013. *PLoS Neglected Tropical Diseases*, 9(2):e0003502.
